# Autism spectrum disorder trios from consanguineous populations are enriched for rare biallelic variants, identifying 32 new candidate genes

**DOI:** 10.1101/2021.12.24.21268340

**Authors:** Ricardo Harripaul, Ansa Rabia, Nasim Vasli, Anna Mikhailov, Ashlyn Rodrigues, Stephen F. Pastore, Tahir Muhammad, Thulasi Thiruvallur Madanagopal, Aisha Nasir Hashmi, Clinton Tran, Cassandra Stan, Katherine Aw, Clement Zai, Maleeha Azam, Saqib Mahmood, Abolfazl Heidari, Raheel Qamar, Leon French, Shreejoy Tripathy, Zehra Agha, Muhammad Iqbal, Majid Ghadami, Susan L. Santangelo, Bita Bozorgmehr, Laila Al Ayadhi, Roksana Sasanfar, Shazia Maqbool, James A. Knowles, Muhammad Ayub, John B Vincent

## Abstract

**Background:** Autism spectrum disorder (ASD) is a neurodevelopmental disorder that affects about 1 in 36 children in the United States, imposing enormous economic and socioemotional burden on families and communities. Genetic studies of ASD have identified *de novo* copy number variants (CNVs) and point mutations that contribute significantly to the genetic architecture, but the majority of these studies were conducted in populations unsuited for detecting autosomal recessive (AR) inheritance. However, several ASD studies in consanguineous populations point towards AR as an under-appreciated source of ASD variants.

**Methods:** We used whole exome sequencing to look for rare variants for ASD in 115 proband-mother-father trios from populations with high rates of consanguinity, namely Pakistan, Iran, and Saudi Arabia. Consanguinity was assessed through microarray genotyping.

**Results:** We report 84 candidate disease-predisposing single nucleotide variants and indels, with 58% biallelic, 25% autosomal dominant/*de novo*, and the rest X-linked, in 39 trios. 52% of the variants were loss of function (LoF) or putative LoF (pLoF), and 47% nonsynonymous. We found an enrichment of biallelic variants, both in sixteen genes previously reported for AR ASD and/or intellectual disability (ID) and 32 previously unreported AR candidate genes (including *DAGLA*, *ENPP6*, *FAXDC2*, *ILDR2*, *KSR2*, *PKD1L1*, *SCN10A*, *SHH*, and *SLC36A1*). We also identified eight candidate biallelic exonic loss CNVs.

**Conclusions:** The significant enrichment for biallelic variants among individuals with high F_roh_ coefficients, compared with low F_roh_, either in known or candidate AR genes, confirms that genetic architecture for ASD among consanguineous populations is different to non-consanguineous populations. Assessment of consanguinity may assist in the genetic diagnostic process for ASD.

## Introduction

Autism spectrum disorder (ASD) is characterized by deficits in social communication, repetitive/restricted behaviours and interests. Apart from a small percentage of individuals who recover (1), ASD is a life-long condition and only about 20% have good outcomes, as reported in a recent meta-analysis of longitudinal studies (2). Phenotypically, individuals with ASD are heterogeneous in their level of intellectual functioning, and co-morbid psychiatric and behavioural problems. These factors play an important role in the long term end points of the disease (3). In a systematic review the prevalence of ASD was estimated to be 0.76% internationally (4). In another systematic review of international data, the median estimate was 62/10,000 (5). In this review, the ASD prevalence showed little variation by geographic region, ethnic, cultural and socioeconomic factors (5).

The reported prevalence of ASD has increased with time. In the 1960s it was estimated to affect as few as 1 in 10,000 individuals (5), prevalence studies from the 1980s suggested that as many as 72 in 10,000 individuals had ASD, rising to 1% in the 2000s (5,6). More recent studies report prevalence rates of more than 2%. (7–9). In the United States the prevalence across eleven sites in 2016 was 18.5 per 1,000 (one in 54) children (10) and increasing to one in 36 in the most recent report (11). This increase is partly due to changes in diagnostic criteria, reporting practices, and increased awareness (12–15).

A systematic review of literature from South Asian countries reported a prevalence of ASD in the range of 0.09-1.07 % (16). To date, there has been no community study of ASD prevalence in Pakistan. Studies from child psychiatry clinics reported rates of 2.4% and 3.2%, 4.5%, 5% (17–20). Clinical services and special schools for ASD are available in Pakistan, but are typically limited to major cities (21). The literature on clinical presentation is limited. One study with a small sample size shows symptoms consistent with studies elsewhere in the world (22).

Consanguineous marriages lead to a marked increase in the frequency of rare recessive disorders (23,24). Many genetic variants will only be pathogenic in recessive form; this includes variants for ID, and almost certainly for ASD too. Populations with a high proportion of consanguinity have been important for describing autosomal recessive genes in ID in Pakistan (25,26), Iran (27,28), Syria (29), and Saudi Arabia (30), yet, to date, relatively few reports of AR genes underlying ASD in consanguineous populations have been published (31–33). Previous work has highlighted recessive inheritance as an important component of the genetic architecture of ASD. For instance, a large study of consanguineous versus non-consanguineous families in India concluded that consanguinity increases the risk for ASD with an odds ratio of 3.22 (34). Morrow and colleagues used SNP microarrays to map homozygous loci in 104 small ASD families from the Middle East, Turkey and Pakistan, finding homozygous deletions implicating *SLC9A9*, *PCDH10*, *CNTN3* and others (31).

Research in outbred populations also supports AR inheritance as an important piece of the genetic puzzle for ASD. For example, it has been estimated that loss-of-function (LoF) recessive mutations contribute 3% of ASD genetics in two US-based case-control cohorts (35). These findings are not limited to population isolates or ethnic subgroups (36,37). Identification of recessive genes in outbred populations is problematic, as the analysis pipelines for WES/WGS data are non-optimal for discovering compound heterozygous mutations.

Despite the higher prevalence of recessive inheritance in consanguineous populations, data from studies of developmental disorder (DD) suggest that *de novo* variants are also prominent, albeit in a lower proportion e.g. in a UK study of 6040 families from the Deciphering Developmental Disorders (DDD) study, individuals of Pakistani ancestry had 29.8% *de novo* compared with 49.9% in the European ancestry UK population (38). In an Iranian ID study, the *de novo* rate was 27.86% (39).

Describing the genetic architecture of ASD is an important step towards understanding the pathogenesis of the disorder. There has been significant progress to date, and studies of ASD have shown that *de novo* copy number variants (CNVs) and point mutations play a prominent role, however, the majority of these studies have been conducted in outbred populations (40). Here, we report on a study of 84 genetic and 11 genomic variants (eight homozygous; 3 heterozygous) from a cohort of 115 ASD trios from three countries with a high frequency of consanguineous marriages (Pakistan, Iran, and Saudi Arabia).

## Materials and Methods

### Trio Family Ascertainment

Institutional Research Ethics Board approval was received from the Centre for Addiction and Mental Health (CAMH) and the recruitment sites. A summary of the cohorts, collaborator contributions, ascertainment, and assessment tools, is in Table 1. Overall, we have collected DNA from 115 trios: 62 trios from Pakistan, 40 trios from Iran, and 13 trios from Saudi Arabia, comprised of 345 individuals.

**Table 1:**
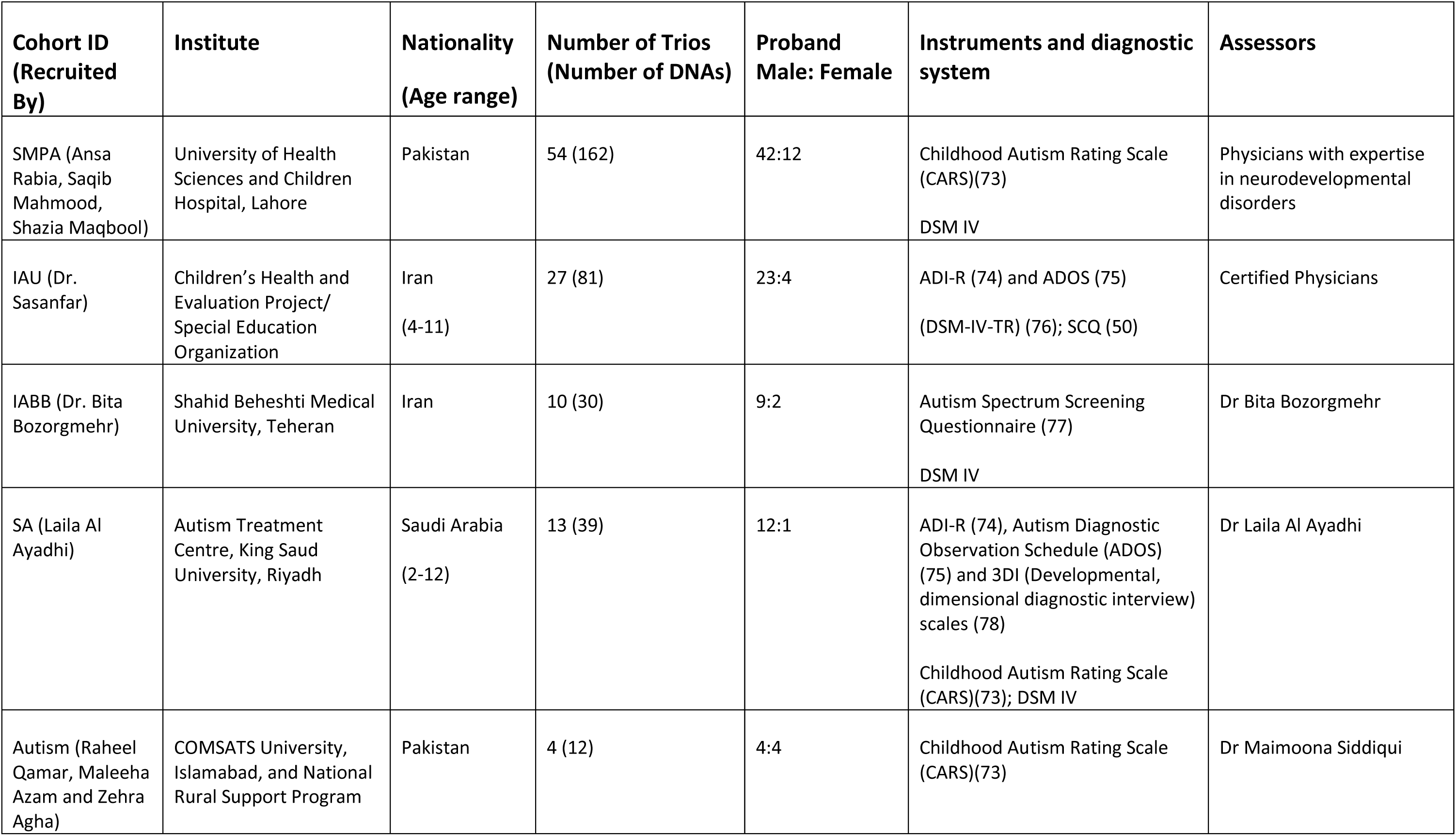
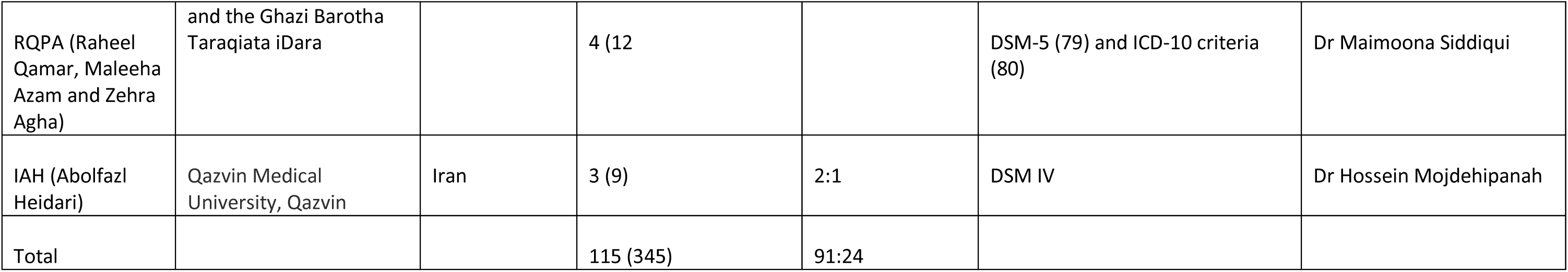
Cohort Description and clinical assessment.

| **Cohort ID**<br>**(Recruited**<br>**By)** | **Institute** | **Nationality**<br>**(Age range)** | **Number of Trios**<br>**(Number of DNAs)** | **Proband**<br>**Male: Female** | **Instruments and diagnostic**<br>**system** | **Assessors** |
| --- | --- | --- | --- | --- | --- | --- |
| SMPA (Ansa Rabia, Saqib Mahmood, Shazia Maqbool) | University of Health Sciences and Children Hospital, Lahore | Pakistan | 54 (162) | 42:12 | Childhood Autism Rating Scale (CARS)(73)<br>DSM IV | Physicians with expertise in neurodevelopmental disorders |
| IAU (Dr. Sasanfar) | Children's Health and Evaluation Project/ Special Education Organization | Iran<br>(4-11) | 27 (81) | 23:4 | ADI-R (74) and ADOS (75)<br>(DSM-IV-TR) (76); SCQ (50) | Certified Physicians |
| IABB (Dr. Bita Bozorgmehr) | Shahid Beheshti Medical University, Teheran | Iran | 10 (30) | 9:2 | Autism Spectrum Screening Questionnaire (77)<br>DSM IV | Dr Bita Bozorgmehr |
| SA (Laila Al Ayadhi) | Autism Treatment Centre, King Saud University, Riyadh | Saudi Arabia<br>(2-12) | 13 (39) | 12:1 | ADI-R (74), Autism Diagnostic Observation Schedule (ADOS) (75) and 3DI (Developmental, dimensional diagnostic interview) scales (78)<br>Childhood Autism Rating Scale (CARS)(73); DSM IV | Dr Laila Al Ayadhi |
| Autism (Raheel Qamar, Maleeha Azam and Zehra Agha) | COMSATS University, Islamabad, and National Rural Support Program | Pakistan | 4 (12) | 4:4 | Childhood Autism Rating Scale (CARS)(73) | Dr Maimoona Siddiqui |
| RQPA (Raheel Qamar, Maleeha Azam and Zehra Agha) | and the Ghazi Barotha Taraqiata iDara |  | 4 (12 |  | DSM-5 (79) and ICD-10 criteria (80) | Dr Maimoona Siddiqui |
| IAH (Abolfazl Heidari) | Qazvin Medical University, Qazvin | Iran | 3 (9) | 2:1 | DSM IV | Dr Hossein Mojdehipanah |
| Total |  |  | 115 (345) | 91:24 |  |  |

### Whole Exome Sequencing, Alignment and Variant Calling

Whole Exome Sequencing (WES) was performed on all samples using the Thruplex DNA-Seq (Rubicon Genomics) Library Preparation Kit with the Agilent SureSelect V5 Exome Capture kit. DNA was sheared using a Covaris ME220 Focused-Sonicator 200 bp, which was verified using a Agilent 2100 Bioanalyzer System for fragment length distribution and quantification. All trios were sequenced on Illumina HiSeq 2500 or NovaSeq sequencing systems. Details of read QC, alignment, variant calling and prioritization are provided in Supplementary Materials, and summarized in Figure 1. Fifteen trios were previously run using the Ion Proton platform/Ion Ampliseq™ Exome kit (Life Technologies)-all were re-run using the Illumina platform except trio IABB3, for which a causative frameshifting deletion was detected in the VPS13B/Cohen syndrome gene.

**Figure 1:**
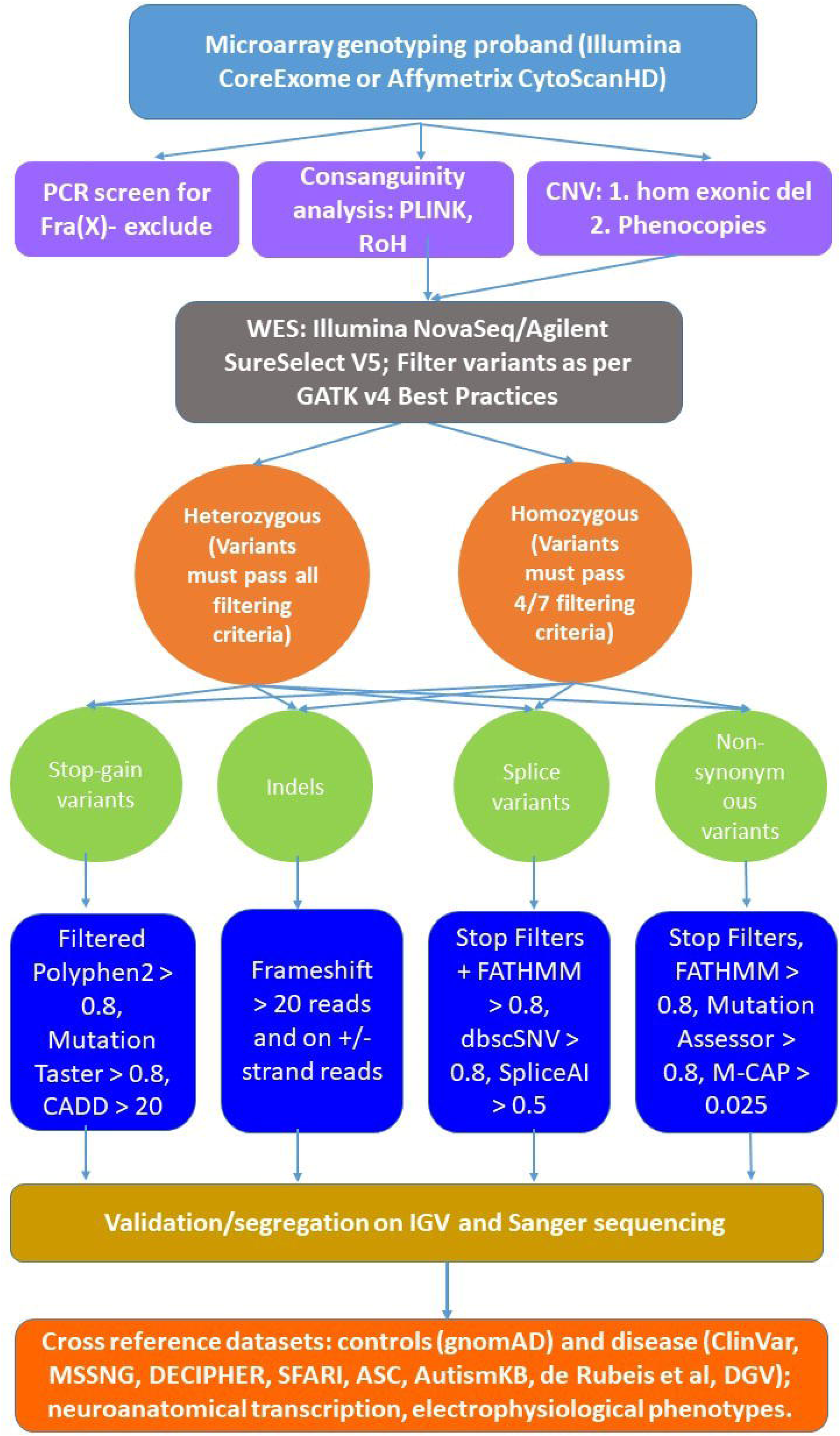
Variant prioritization methods where variants that pass GATK best practices would be filtered for MAF less than or equal to 10^-4^ and then categorized into recessive or homozygous variants. Variants are then prioritized based on assessment of potential damage to the protein, and then filtered based on scoring algorithms to predict pathogenicity.

### Microarray genotyping and analysis

Microarrays were run using DNA from 104 of the 115 ASD probands (where sufficient DNA was available), including 13 using Affymetrix CytoScanHD, 87 using Illumina CoreExome, and four with both arrays. CoreExome data in PLINK format was used for runs-of-homozygosity (RoH) analysis, using PLINK 1.9 (http://pngu.mgh.harvard.edu/purcell/plink). RoH data was used to generate an F-coefficient of consanguinity for the probands (and affected siblings), F_RoH_, as described in McQuillan et al, 2008 (41), where F_roh_ = ∑ L_roh_/L_auto_. Autosome size L_auto_ was estimated at 2,673,768 Kb. The CytoScanHD data was analysed using Chromosome Analysis Suite (ChAS), and a loss of heterozygosity (LoH) score was generated. For four samples that were run on both Illumina CoreExome and Affymetrix CytoScanHD arrays (probands from trios IABB2, IABB3, IABB4, and IABB5), we were able to generate both F_roh_ and LoH scores, which, as the relationship was almost linear (Pearson R^2^ = 0.9971) allowed us to convert LoH scores from CytoScanHD microarrays to F_roh_ (Supplementary Materials). Comparison statistics were performed using GraphPad (https://www.graphpad.com/).

### Copy Number Variant (CNV) analysis

CNV analysis of the CytoScanHD array data was performed using ChAS and PennCNV; for CoreExome data by the Illumina Genome Suite/CNVpartition and PennCNV. Evidence of homozygous loss CNVs was cross-references with WES data, using the Integrated Genome Viewer v2.3.5 (IGV: https://software.broadinstitute.org/software/igv/; (42), and CNVs corroborated in this manner were then checked by PCR. Further analysis/prioritization/validation of CNVs, including using WES data, is given in Supplementary Materials.

### Cross-referencing with other datasets

In order to evaluate the strength of candidacy of the variants/genes identified here or to find supporting evidence, we cross-referenced our findings with other datasets listed below. We identified rare variants in the candidate genes we report here (Tables 2-4) that were either: 1. *De novo*, 2. Heterozygous LoF but status either de novo or unknown; 3. Biallelic (i.e. homozygous); 4. Putative biallelic (possibly compound heterozygous, but phasing unknown); 5. X-linked in males, maternally inherited. Data are shown in Supplementary File 1.

**Table 2:**
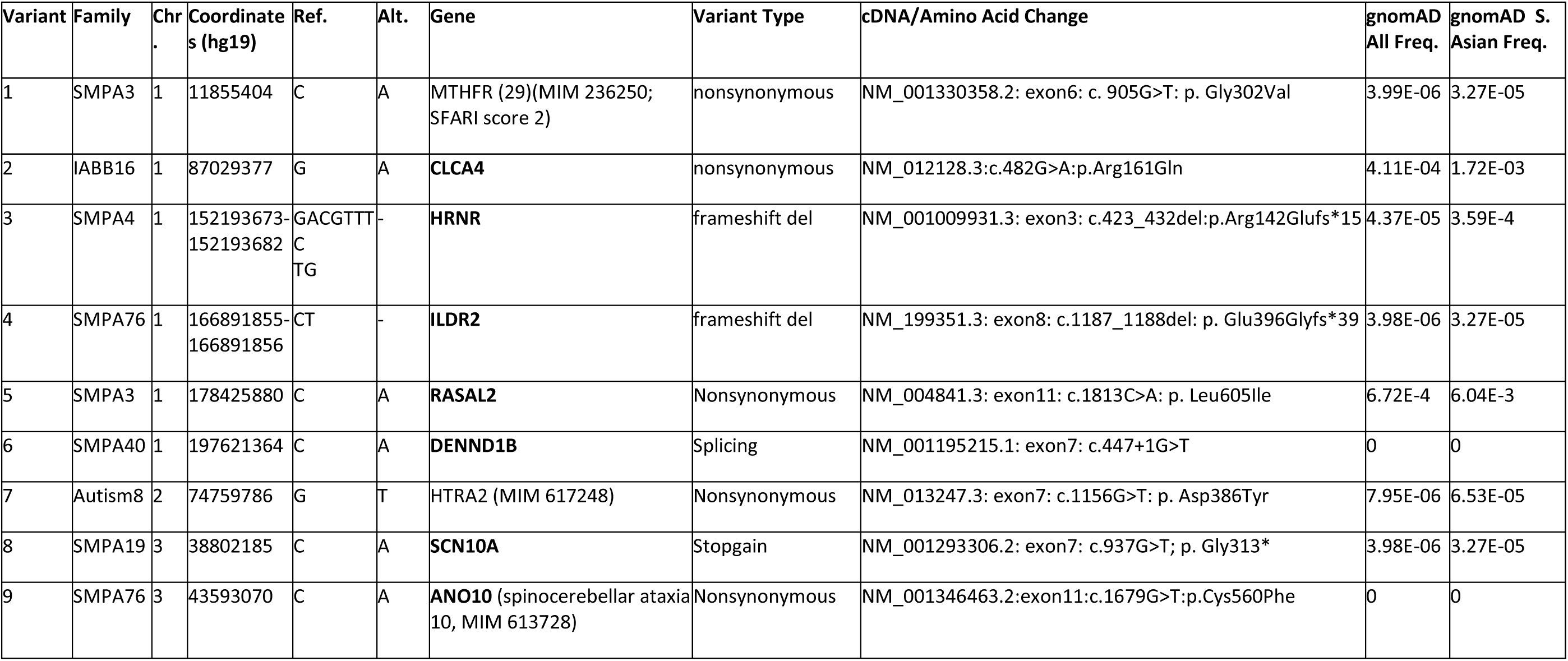
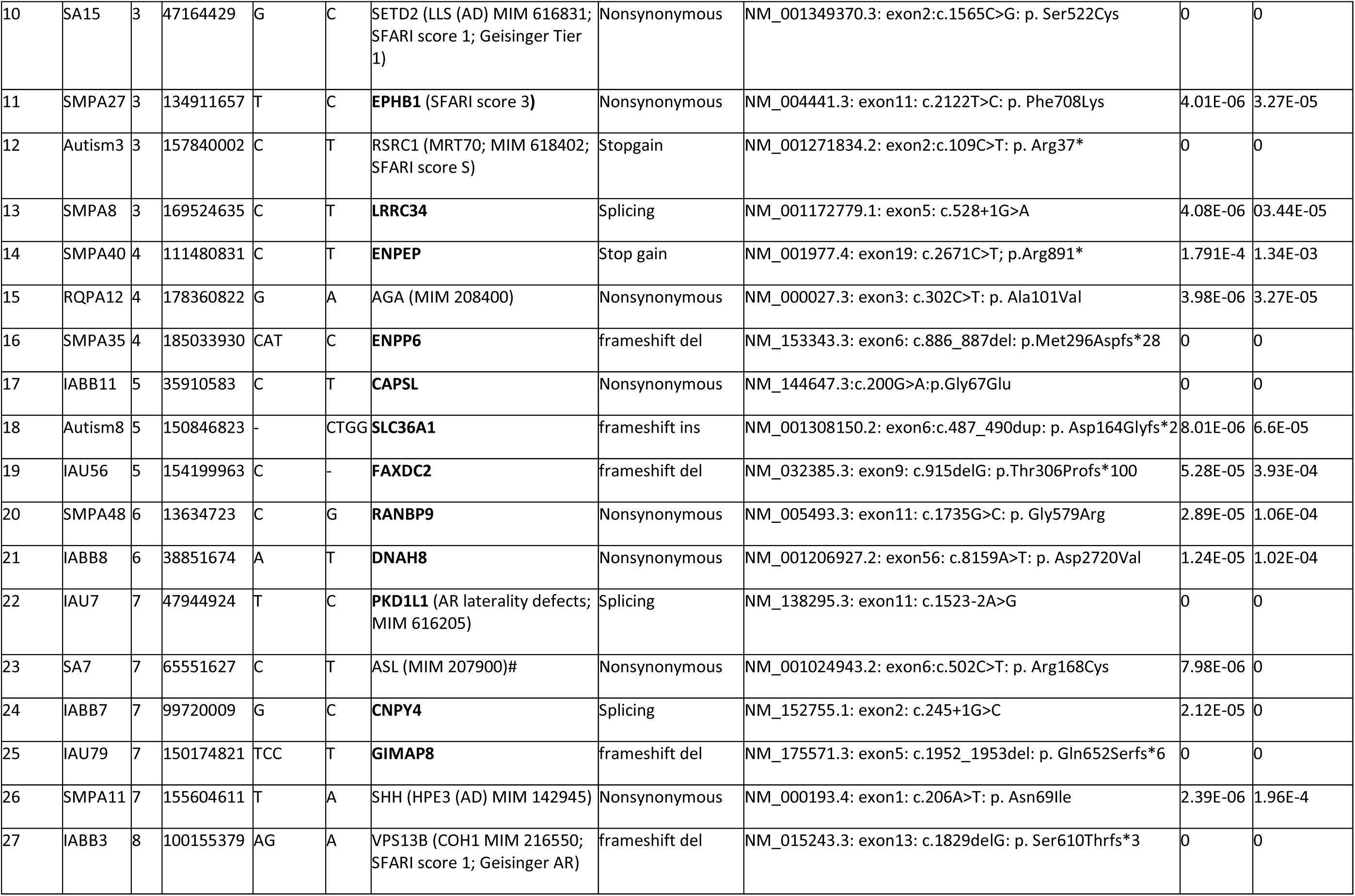
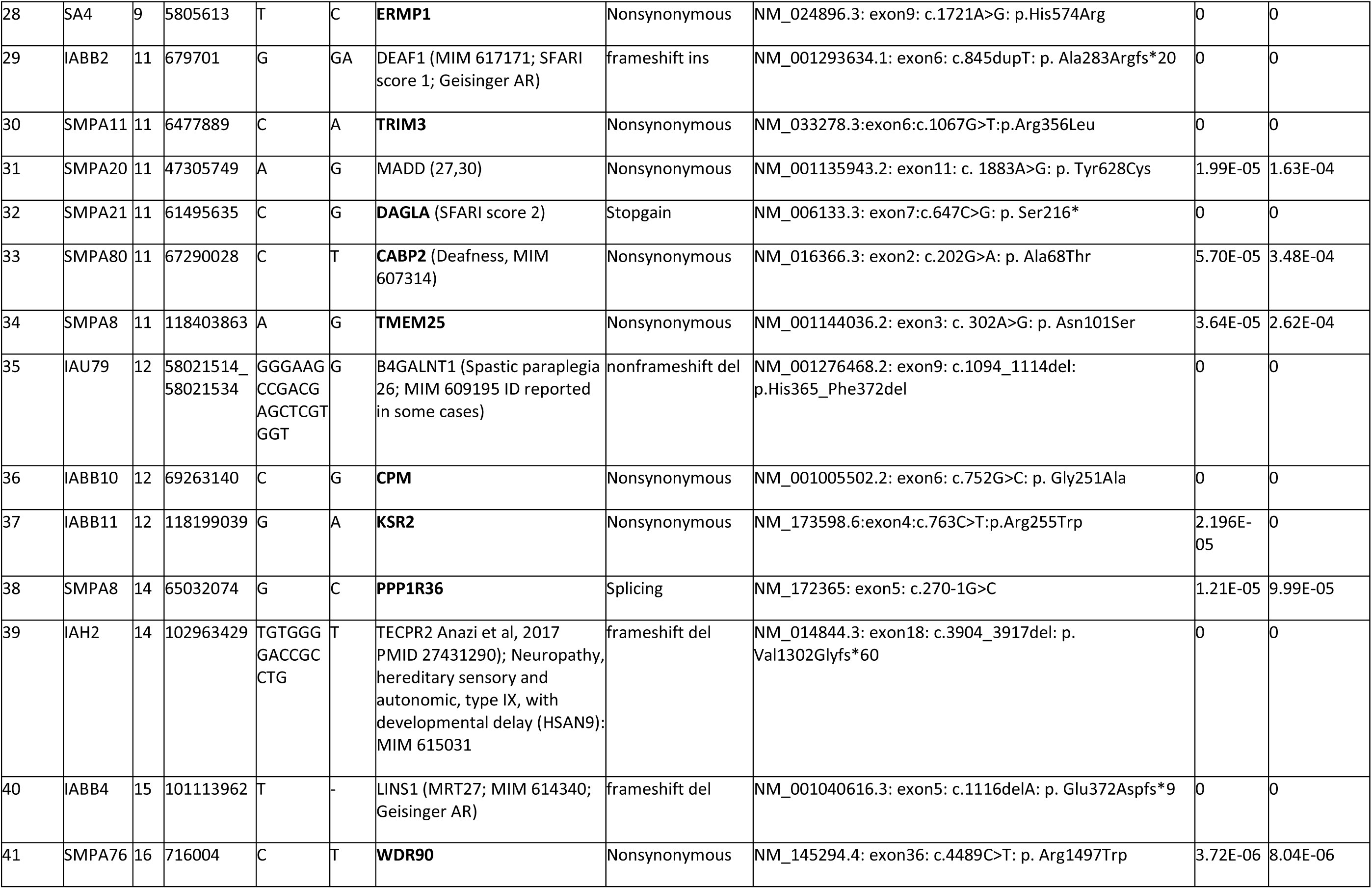
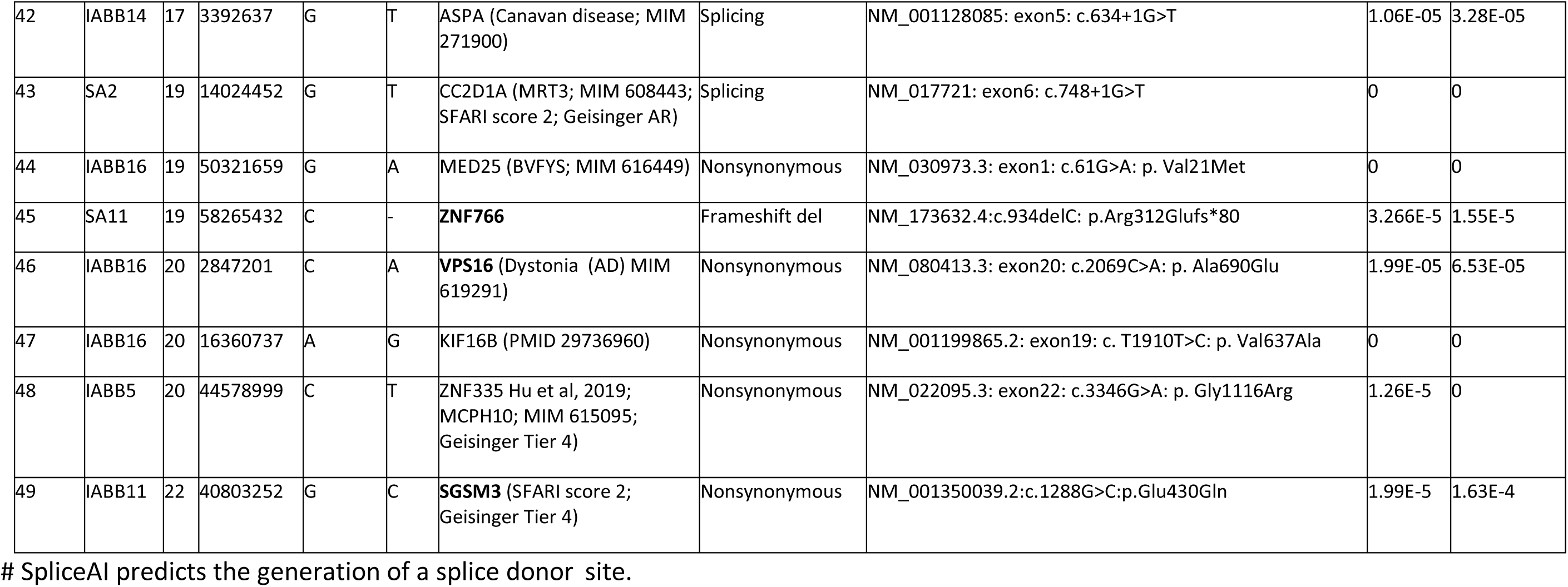
Biallelic/homozygous variants: Genes previously reported as pathogenic for ASD/ID or other disorder are indicated, with annotation using (in order of priority): OMIM: https://omim.org/; SFARI ASD genes https://gene.sfari.org/; DDD gene2phenotype genes: https://www.ebi.ac.uk/gene2phenotype/; Geisinger developmental brain disorder gene database: https://dbd.geisingeradmi.org/; NP*denovo*: http://www.wzgenomics.cn/NPdenovo/; Gene4denovo: http://www.genemed.tech/gene4denovo/search. Genes previously unreported for ASD or ID are in bold type. Control population frequencies show are from gnomAD (all, and for South Asian sub-population). All variants were validated by IGV (Integrated Genomics Viewer; Broad Institute) and by Sanger sequencing. Validations are shown in Supplementary Materials. Allele frequencies are from gnomAD v2.1.1 (www. https://gnomad.broadinstitute.org/; accessed June 2022).

**Table 3:**
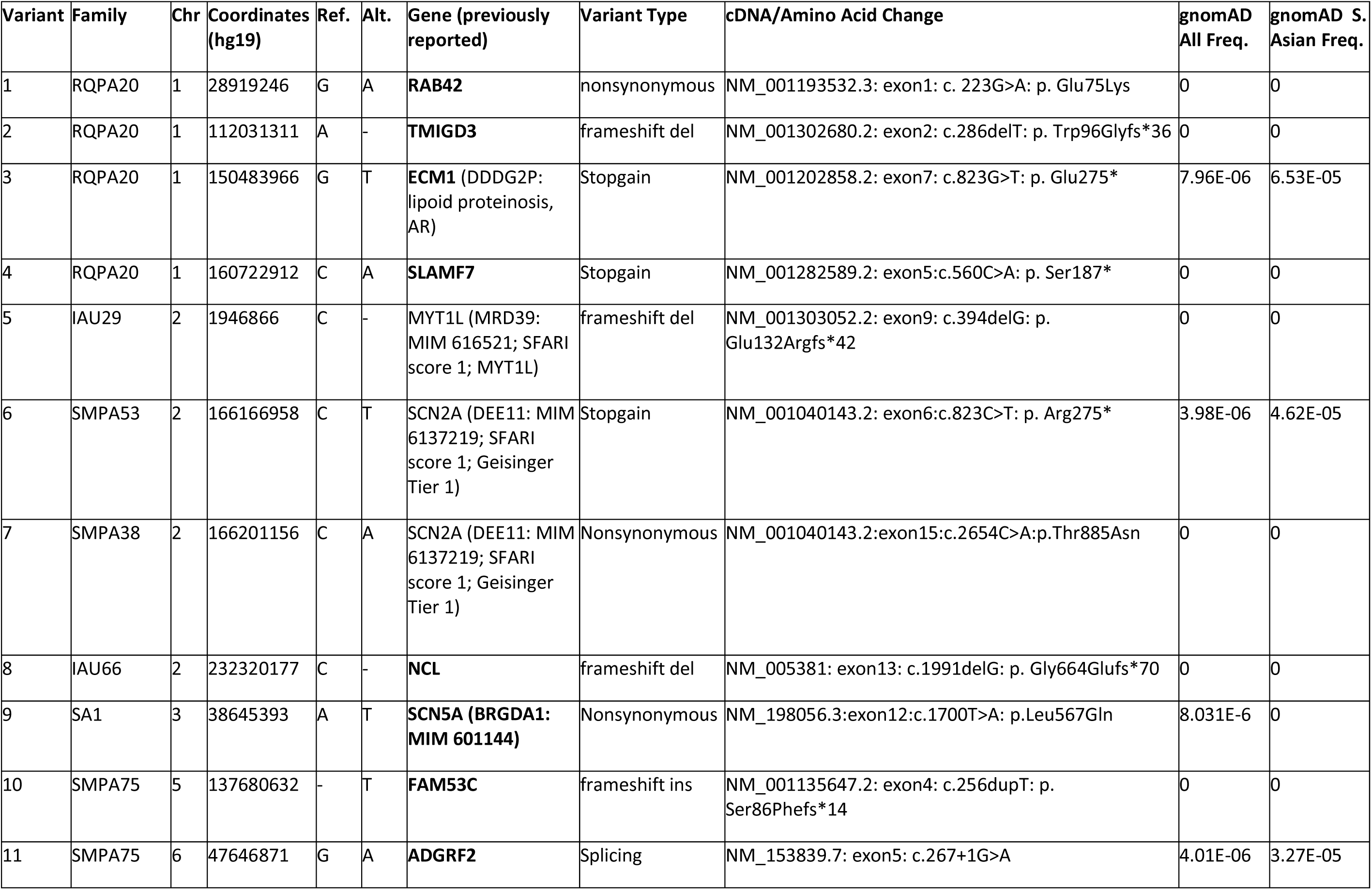
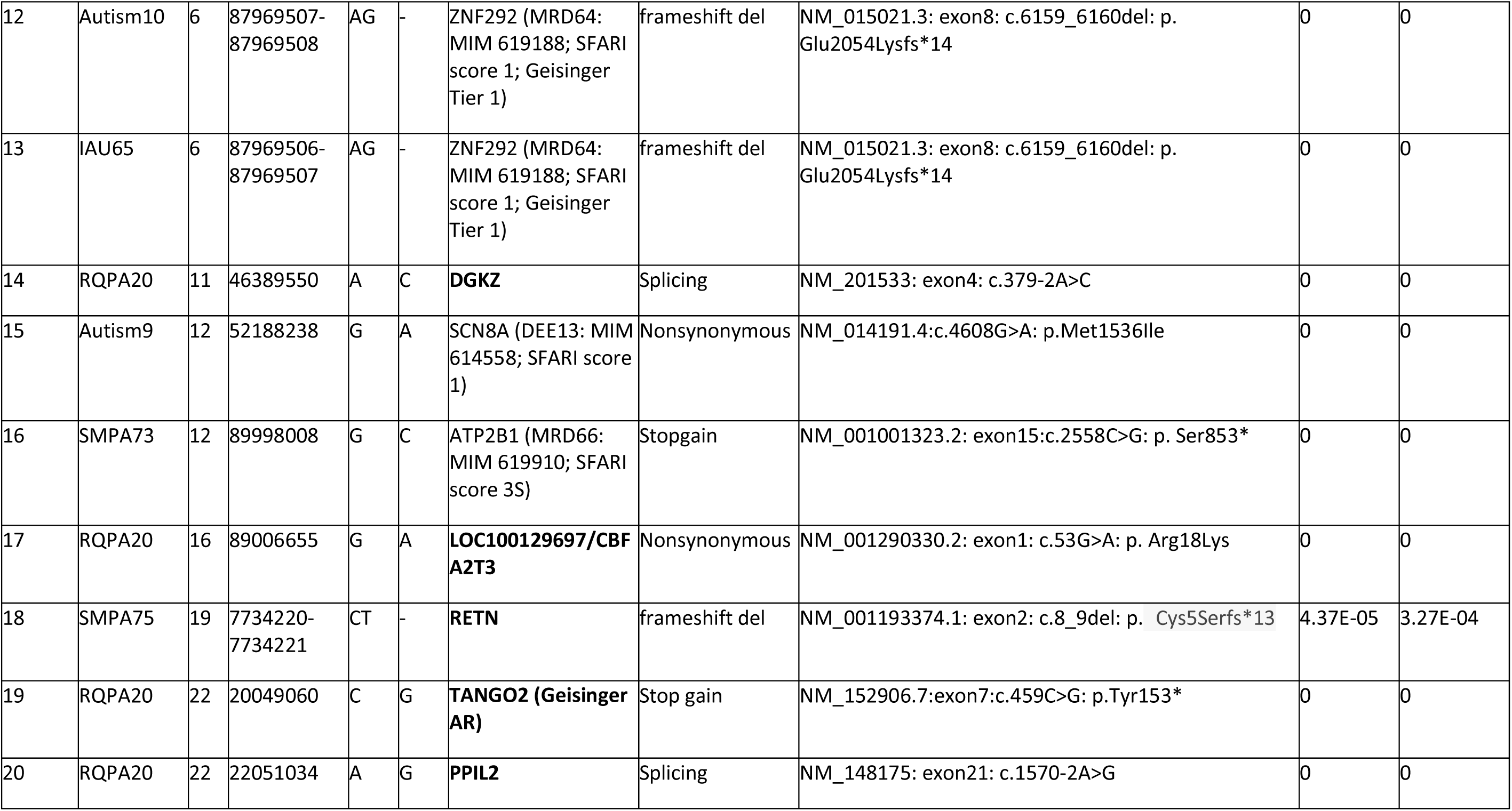
*De novo* variants. Genes previously reported as pathogenic for ASD/ID or other disorder are indicated, with annotation using (in order of priority): OMIM: https://omim.org/; SFARI ASD genes https://gene.sfari.org/; DDD gene2phenotype genes: https://www.ebi.ac.uk/gene2phenotype/; Geisinger developmental brain disorder gene database: https://dbd.geisingeradmi.org/; NP*denovo*: http://www.wzgenomics.cn/NPdenovo/; Gene4denovo: http://www.genemed.tech/gene4denovo/search. Genes previously unreported for ASD or ID are in bold type. Control population frequencies shown are from gnomAD (all, and for South Asian sub-population). Allele frequencies are from gnomAD v2.1.1 (www. https://gnomad.broadinstitute.org/; accessed June 2022). All variants were validated by IGV (Integrated Genomics Viewer; Broad Institute) and by Sanger sequencing. Validations are shown in Supplementary Materials.

**Table 4:**
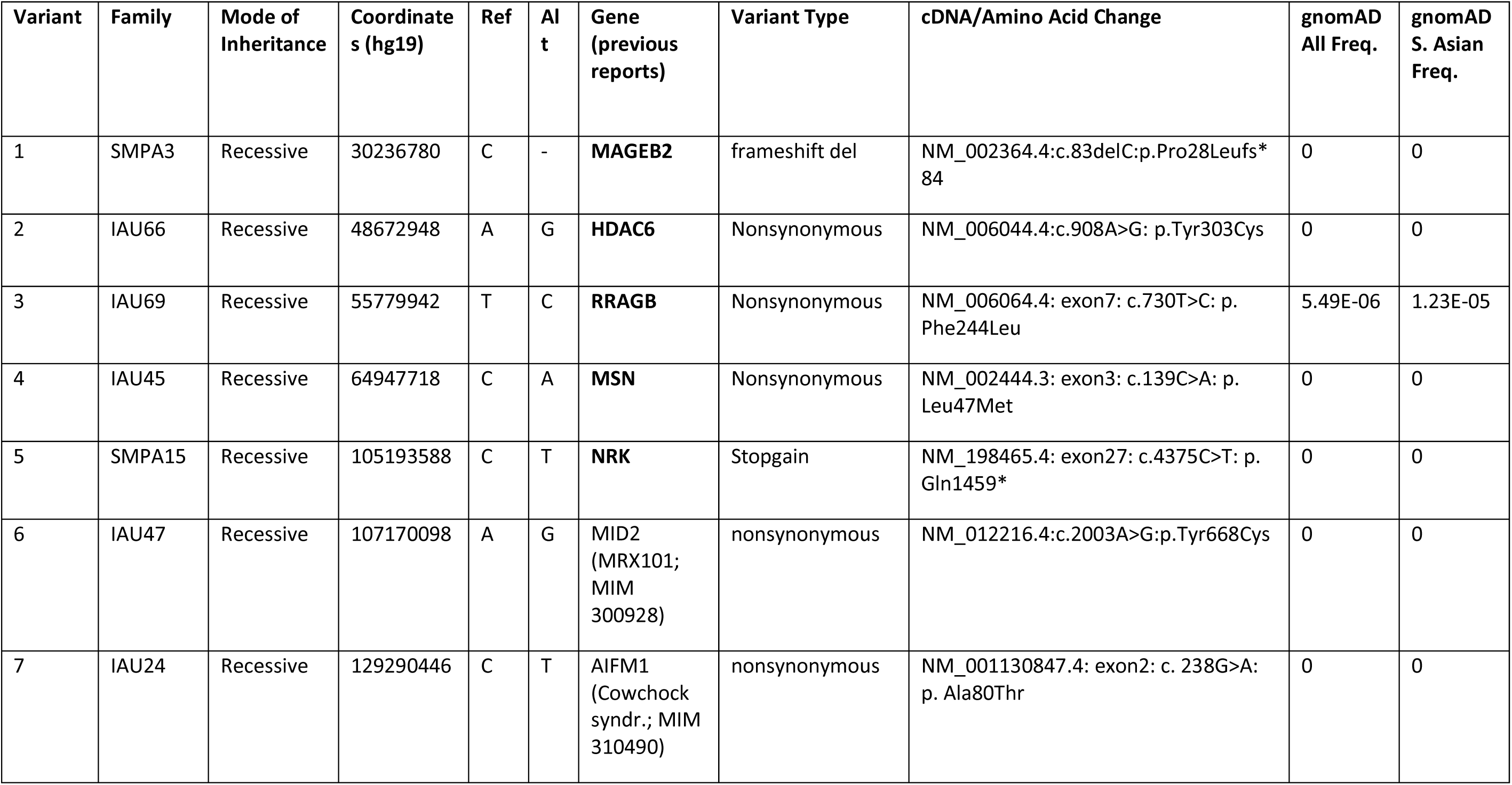
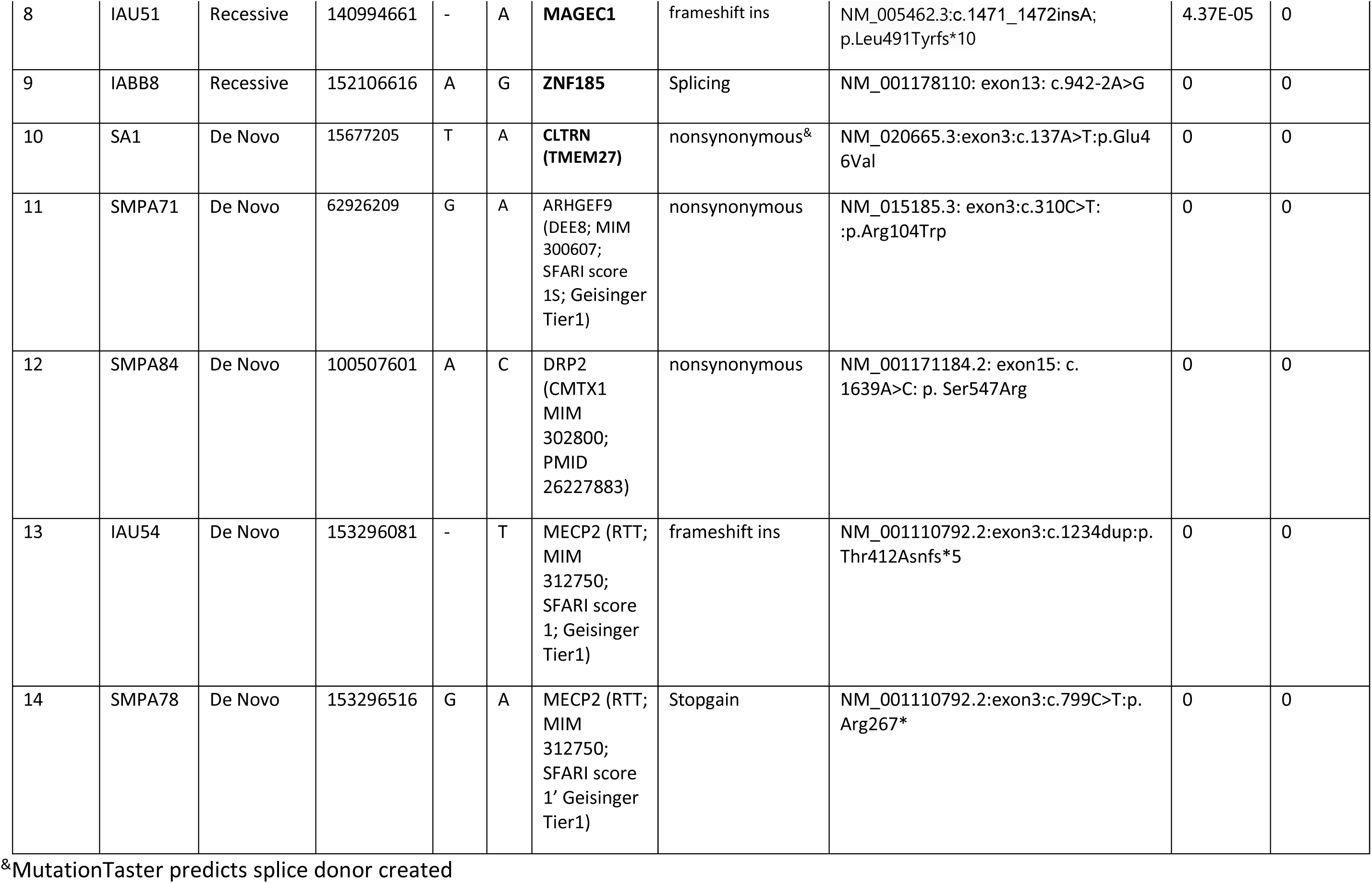
X-linked variants. Genes previously reported as pathogenic for ASD/ID or other disorder are indicated, with annotation using (in order of priority): OMIM: https://omim.org/; SFARI ASD genes https://gene.sfari.org/; DDD gene2phenotype genes: https://www.ebi.ac.uk/gene2phenotype/; Geisinger developmental brain disorder gene database: https://dbd.geisingeradmi.org/; NP*denovo*: http://www.wzgenomics.cn/NPdenovo/; Gene4denovo: http://www.genemed.tech/gene4denovo/search. Genes previously unreported for ASD or ID are in bold type. Control population frequencies shown are from gnomAD (all, and for South Asian sub-population). Allele frequencies are from gnomAD v2.1.1 (www. https://gnomad.broadinstitute.org/; accessed June 2022). All variants were validated by IGV (Integrated Genomics Viewer; Broad Institute) and by Sanger sequencing. Validations are shown in Supplementary Materials.

| Variant | Family | Mode of Inheritance | Coordinates (hg19) | Ref | Alt | Gene (previous reports) | Variant Type | cDNA/Amino Acid Change | gnomAD All Freq. | gnomAD S. Asian Freq. |
| --- | --- | --- | --- | --- | --- | --- | --- | --- | --- | --- |
| 1 | SMPA3 | Recessive | 30236780 | C | - | **MAGEB2** | frameshift del | NM_002364.4:c.83delC:p.Pro28Leufs*84 | 0 | 0 |
| 2 | IAU66 | Recessive | 48672948 | A | G | **HDAC6** | Nonsynonymous | NM_006044.4:c.908A>G: p.Tyr303Cys | 0 | 0 |
| 3 | IAU69 | Recessive | 55779942 | T | C | **RRAGB** | Nonsynonymous | NM_006064.4: exon7: c.730T>C: p. Phe244Leu | 5.49E-06 | 1.23E-05 |
| 4 | IAU45 | Recessive | 64947718 | C | A | **MSN** | Nonsynonymous | NM_002444.3: exon3: c.139C>A: p. Leu47Met | 0 | 0 |
| 5 | SMPA15 | Recessive | 105193588 | C | T | **NRK** | Stopgain | NM_198465.4: exon27: c.4375C>T: p. Gln1459* | 0 | 0 |
| 6 | IAU47 | Recessive | 107170098 | A | G | MID2 (MRX101; MIM 300928) | nonsynonymous | NM_012216.4:c.2003A>G:p.Tyr668Cys | 0 | 0 |
| 7 | IAU24 | Recessive | 129290446 | C | T | AIFM1 (Cowchock syndr.; MIM 310490) | nonsynonymous | NM_001130847.4: exon2: c. 238G>A: p. Ala80Thr | 0 | 0 |
| 8 | IAU51 | Recessive | 140994661 | - | A | **MAGEC1** | frameshift ins | NM_005462.3:c.1471_1472insA;<br>p.Leu491Tyrfs*10 | 4.37E-05 | 0 |
| 9 | IABB8 | Recessive | 152106616 | A | G | **ZNF185** | Splicing | NM_001178110: exon13: c.942-2A>G | 0 | 0 |
| 10 | SA1 | De Novo | 15677205 | T | A | **CLTRN**<br>**(TMEM27)** | nonsynonymous<sup>&</sup> | NM_020665.3:exon3:c.137A>T:p.Glu4<br>6Val | 0 | 0 |
| 11 | SMPA71 | De Novo | 62926209 | G | A | ARHGEF9<br>(DEE8; MIM<br>300607;<br>SFARI score<br>1S; Geisinger<br>Tier1) | nonsynonymous | NM_015185.3: exon3:c.310C>T:<br>:p.Arg104Trp | 0 | 0 |
| 12 | SMPA84 | De Novo | 100507601 | A | C | DRP2<br>(CMTX1<br>MIM<br>302800;<br>PMID<br>26227883) | nonsynonymous | NM_001171184.2: exon15: c.<br>1639A>C: p. Ser547Arg | 0 | 0 |
| 13 | IAU54 | De Novo | 153296081 | - | T | MECP2 (RTT;<br>MIM<br>312750;<br>SFARI score<br>1; Geisinger<br>Tier1) | frameshift ins | NM_001110792.2:exon3:c.1234dup:p.<br>Thr412Asnfs*5 | 0 | 0 |
| 14 | SMPA78 | De Novo | 153296516 | G | A | MECP2 (RTT;<br>MIM<br>312750;<br>SFARI score<br>1' Geisinger<br>Tier1) | Stopgain | NM_001110792.2:exon3:c.799C>T:p.<br>Arg267* | 0 | 0 |

<sup>&</sup>MutationTaster predicts splice donor created

#### MSSNG

More than 13,000 individuals from the Autism Genetic Research Exchange (AGRE) repository and other cohorts have been whole-genome sequenced, through Autism Speaks, and data made available through the MSSNG database (https://research.mss.ng; db7 (43). MSSNG data was accessed in May 2024.

#### SFARI: SSC biallelic

The Simons Simplex Collection (SSC) includes 2,600 simplex autism or related developmental disorder families. WES data is available for ∼2,500 of these families, biallelic variants for our gene list were searched using the GPF browser (https://gpf.sfari.org/gpf19/datasets/SSC/browser). Putative compound heterozygous variants criteria for minor allele frequency (MAF) in SSC exome, gnomAD exome and genome frequencies, of <0.001, or, for missense variants, MCP scores >1 and CADD scores >18.

#### SVIP biallelic

the autism Simons Variation in Individuals Project (SVIP) dataset was searched for putative biallelic variants through the browser gpf.sfari.org/gpf19/datasets/SVIP/browser. Putative compound heterozygous variants checked for criteria including minor allele frequency (MAF) in SSC exome, gnomAD exome and genome frequencies, of <0.001, or, for missense variants, MCP scores >1 and CADD scores >18.

#### Deciphering Developmental Disorders (DDD) study

includes sequence data for ∼14,000 children; this dataset was searched for variants in genes overlapping with our study through the known developmental genes list (https://decipher.sanger.ac.uk/ddd#ddgenes), as well as the list of research variants (https://decipher.sanger.ac.uk/ddd#research-variants), which are 2,723 variants of unknown significance from 4,293 children. For *de novo* variants, all are constitutive, unless specifically noted as mosaic here. CNV variants are not included here.

#### Autism Sequencing Consortium (ASC)

genes with *de novo* variants, either in the control set or case set, from ASC exome analysis browser (https://asc.broadinstitute.org/). This dataset lists numbers of *de novo* protein-truncating variants, as well as *de novo* missense variants with MCP scores either 1-2 or ≥2. The ASC dataset includes 6,430 probands, and includes the SSC data.

#### DeRubeis

genes with *de novo* variants reported in the DeRubeis et al, 2014 study, with either LoF or missense variants (44). As subject IDs were not reported, overlap with other studies recorded here is not possible.

#### AutismKB

additional studies where variants have been reported for the genes are taken from the Autism Knowledge Base (http://db.cbi.pku.edu.cn/autismkb_v2/quick_search.php), (45).

### Neuroanatomical Enrichment Analysis

The Allen Adult Human Brain Atlas (46) , Brainspan (47) , and a single-cell atlas of the mouse nervous system (48) were used to test for neuroanatomically specific expression. For each gene, expression was standardized across regions or cell types. For these compartments, each gene was then ranked from most specific to most depleted. The area under the receiver operating statistic (AUC) was used to quantify specific expression for the genes of interest within a region or cell cluster. The Mann-Whitney U test was used to test statistical significance, and the Benjamini–Hochberg procedure (49) was used to correct for multiple tests. In addition, in order to explore whether our candidate genes may contribute to electrophysiological and morphological phenotypes in neurons we cross-referenced gene expression correlates with electrophysiological phenotypes through a dataset published by Bomkamp et al, 2019 (50).

## Results

We ascertained, phenotyped, collect blood and/or saliva for DNA extraction, and performed whole exome sequencing (mean depth: 27.29; average % coverage at 30X=31.01; average % coverage at 100X=7.32; see Supplementary File 1) from 115 ASD proband (91M:24F)/mother/father trios (62 from Pakistan, 40 from Iran, and 13 from Saudi Arabia), for a total of 345 subjects (Table1). In addition, microarrays were run on 104 of the ASD probands, providing data for CNV and ROH analyses.

### Whole Exome Sequencing

We discovered 84 SNVs or indels through our analysis pipeline, including 49 homozygous autosomal variants, 20 autosomal *de novo* (and 1 inherited) variants, and 5 *de novo* and 9 maternally inherited X-linked variants.

### Biallelic mutations (Table 2)

We identified homozygous variants (7 LoF or pLoF; 9 nonsynonymous; 1 in-frame del) in 17 genes that have been previously reported to be associated with autosomal recessive non-syndromic ID, including *BTN3A2* (30), *CC2D1A* (MIM 608443; MRT3; (51)), *LINS1* (28) , *MADD* (30), *MTHFR* (29), *RSRC1* (52,53), *TECPR2* (30) and *ZNF335* (27), or for syndromic or metabolic forms of autosomal recessive ID, including *AGA* (aspartylglucosaminuria), *ASL* (arginosuccinic aciduria), *ASPA* (aspartoacylase deficiency/Canavan disease), and *HTRA2* (3-methylglutaconic aciduria). Three genes, *CC2D1A*, *DEAF1, and VPS13B* have also previously been associated with ASD or autistic features (94–96; (57)). Biallelic mutations in *DEAF1* are known to cause neurodevelopmental disorder with hypotonia, impaired expressive language, with or without seizures (MIM 617171). LoF variants were reported for new candidate genes ENPEP, *DAGLA*, *FAXDC2*, *GIMAP8*, *HRNR*, *ILDR2*, *SLC36A1*, *SCN10A*, *VPS16*, ZNF766, and pLoF (such as canonical splice site variants) in *CNPY4*, *DENND1B*, *LRRC34*, *PKD1L1*, and *PPP1R36*. *VPS16* has previously been reported for autosomal dominant dystonia (MIM 608559), and *PKD1L1* for AR visceral heterotaxy 8 (MIM 617205). Heterozygous missense and LoF mutations in *SHH* are known to be involved in autosomal dominant holoprosencephaly 3 (HPE3; MIM 142945), and so it is somewhat surprising to find a homozygous missense variant associated with a milder phenotype and with no obvious HPE3-related dysmorphic features. The Asn69Ile variant in *SHH* substitutes an asparagine for isoleucine at a residue that is conserved across vertebrates, is predicted to be damaging, and is found in heterozygous form in only 21 out of 1.6E6 gnomAD (v4.1.0) alleles, and no homozygotes.

In one trio (trio SMPA21) we report a homozygous stop-gain variant in *DAGLA*, for which rare heterozygous variants were reported to be associated with neurodevelopmental disorders, including autism (58). SFARI also lists this as a strong candidate for a ASD disease gene (gene score of 2; https://gene.sfari.org/). We also report homozygous missense variants in novel candidate genes *ANO10*, *CABP2*, *CAPSL*, *CLCA4*, *CPM*, *DNAH8*, *ERMP1*, *KSR2*, *RANBP9*, *RASAL2*, *TMEM25*, *TRIM3*, *VPS16*, and *WDR90*, plus in *SGSM3* and *EPHB1*, for which *de novo* variants in have been reported for ASD (59), and are listed respectively as strong candidate and suggestive evidence in SFARI (scores of 2 and 3, respectively).

Although not focused on consanguineous populations, WGS data from the MSSNG study, as well as WES studies such as DDD and SFARI, support a number of our candidate AR genes (see Supplementary File 1), with homozygous rare variants in *DNAH7* (MSSNG), *DNAH8* (MSSNG), *CAPSL* (MSSNG), and *CYP2A7* (MSSNG), and putative compound heterozygous damaging rare variants in *DNAH8* (in three MSSNG individuals), *SCN10A* (DDD), and *WDR90* (DDD)(see Supplementary File 1). There was also evidence of disease-causing biallelic variants in known AR neurodevelopmental genes *MTHFR* (three homozygous; DDD), *AGA* (one homozygous stop gain; DDD), *DEAF1* (two homozygous stop gain; MSSNG and DDD), *ZNF335* (one homozygous missense, DDD; one putative compound heterozygous, DDD), *ASL* (one homozygous missense; MSSNG), *CC2D1A* (one homozygous missense, MSSNG; two homozygous LoF DDD), and *VPS13B* (six homozygous, nine putative compound heterozygous).

### *De novo* mutations (Table 3 & 4)

*De novo* dominant mutations have been shown to be a major causal factor in ASD etiology. We identified 25 *de novo* variants (20 autosomal and 5 X-linked). Among these genes, variants in four of them have been identified in previous studies (*MECP2*, *MYT1L*, *SCN2A*, *ZNF292*). 14 of the 25 variants were LoF, three were putative LoF (splice site), and the remainder were nonsynonymous. The mutations include genes that have been implicated in other recent studies of ASD and/or ID. For instance, we report a *de novo* hemizygous nonsynonymous mutation in *CLTRN* (on chromosome X). Hemizygous mutations in *CLTRN* have recently been reported in several males with neutral aminoaciduria accompanied with autistic features, anxiety, depression, compulsions and motor tics (60)-a clinical picture reminiscent of the autosomal recessive Hartnup disorder (MIM 234500). There were also several other genes with *de novo* variants that have not been associated with ASD (or ID) previously, and may represent novel putative genes for ASD, and for other trios there are variants in genes linked with other nervous system disorders, such as *DRP2* in Charcot-Marie-Tooth disease (61).

Our study identified a *de novo* LoF mutation in *ZNF292* (NM_015021:c.6159_6160del; p.Glu2054Lysfs*14) in two trios (IAU-65 (Iranian) and Autism-10 (Pakistani)). These two trios and 26 additional families (from multiple studies and cohorts) with mutations in *ZNF292* were reported recently (62).

Cross-referencing with other datasets, there was also support for new candidate dominant/*de novo* autosomal genes, including *ECM1*, *SLAMF7*, *NCL*, *FAM53C*, *ADGRF2*, *TANGO2*, *DGKZ*, *ATP2B1* (5 individuals), *CBFA2T3*, *RETN*, and *PPIL2* (see Supplementary File 1). There was support for X-linked recessive (in males, maternally inherited or *de novo*) candidate gene *NRK* (three individuals), *MAGEB2* (three individuals), *MAGEC1* (11 individuals), *MSN* (five individuals), and *ZNF185* (one individuals).

### X-linked (inherited; Table 4)

Two mutations were identified in well-established ASD or ID genes, namely, *AIFM1* (Cowchock syndrome; MIM 310490), and *MID2* (XLID101: MIM 300928).

### Splice site variants

In addition to the detection of variants at canonical splice donor and splice acceptor sites (Tables 2-4), use of SpliceAI suggests that, for the biallelic nonsynonymous variant in the known ASD/ID gene *ASL* (trio SA7), an alternative explanation could be the generation of an alternative splice donor by the cytosine to thymine transition at this site. However, other splice algorithms such as https://www.fruitfly.org/seq_tools/splice.html do not support this, and molecular experimental procedures may be needed to corroborate or refute this prediction.

### Copy Number Variants (Table 5 & 6)

Eight autosomal biallelic loss CNVs were identified among five trios using microarray data, that were subsequently validated through inspection of the WES data using IGV, and through molecular methods, using PCR and/or Sanger sequencing. Of these implicated genes, mutations/knock-outs of both *DHRS4* and *KLK15* have a neurobehavioral phenotype in mouse models (Supplementary File 1). *DNAH7* is highly expressed in brain, and both SHPK (sedoheptulokinase) and WDR73 have high expression in cerebellum. Of the biallelic CNVs identified, *WDR73* has previously been associated with the autosomal recessive Galloway-Mowat syndrome 1 (MIM 251300), which includes microcephaly, delayed psychomotor development and cerebellar atrophy. Determining the ASD causative genetic variation in many of the probands with CNVs is difficult. The proband with the *WRD73*, *CYP2A7*, and *KLK15* CNVs (trio IABB14) also has a biallelic splice mutation in the Canavan disease gene *ASPA*. Additionally, the proband with the *SHPK* CNV (trio IABB2) also has a biallelic LoF mutation in the known ID gene *DEAF1*, and the proband with the *SIRPB1* CNV (trio IAU79) has a biallelic non-frameshift deletion in the known ID/spastic paraplegia gene *B4GALNT1*, complicating identification of the molecular causation (see Supplementary File 1). A biallelic CNV loss implicates the gene *SEMG1* in trio SMPA4, however expression of this gene is restricted to the seminal vesicle (see Supplementary File 1), and thus unlikely to be related to the ASD phenotype.

**Table 5:**
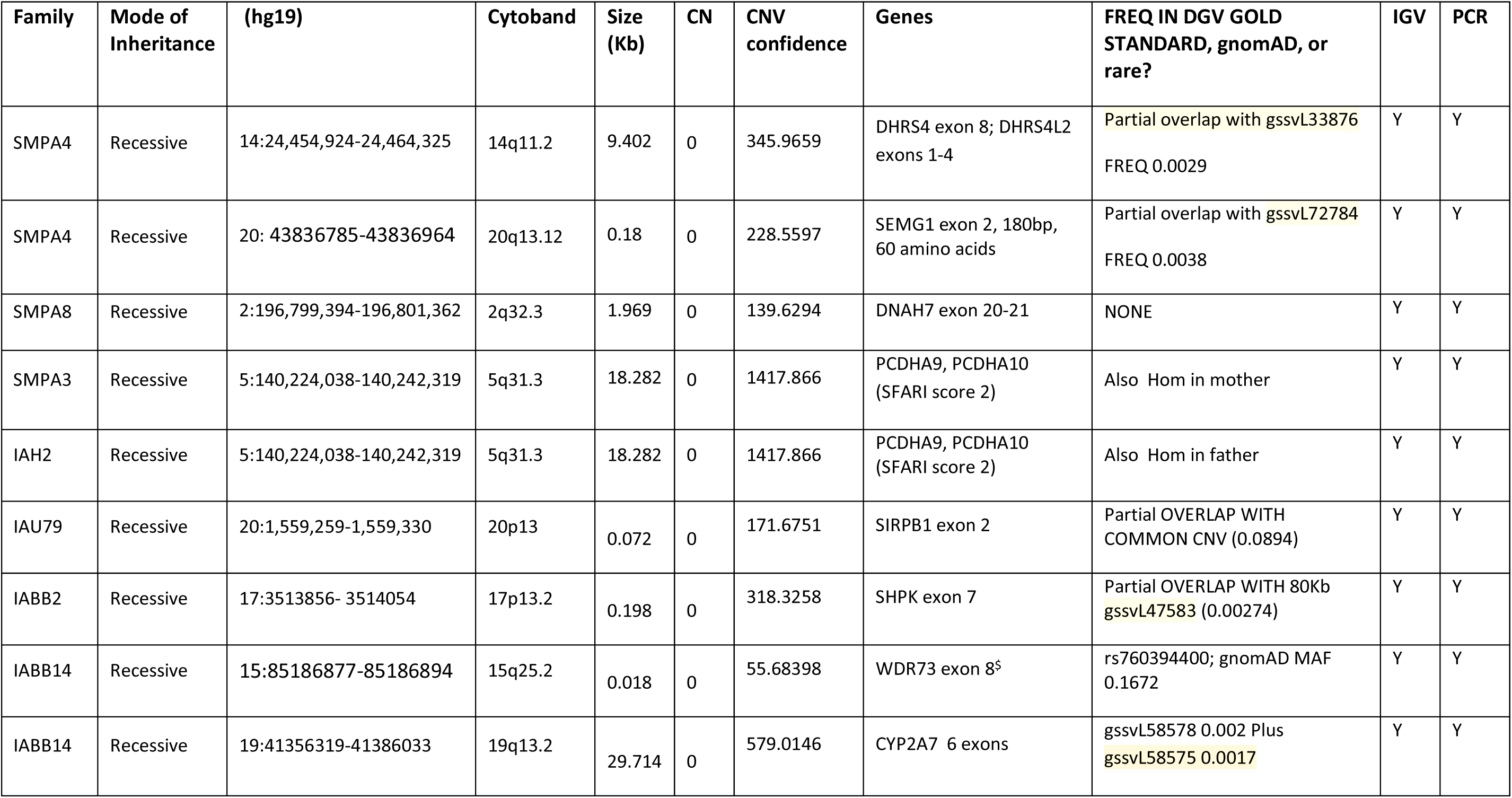
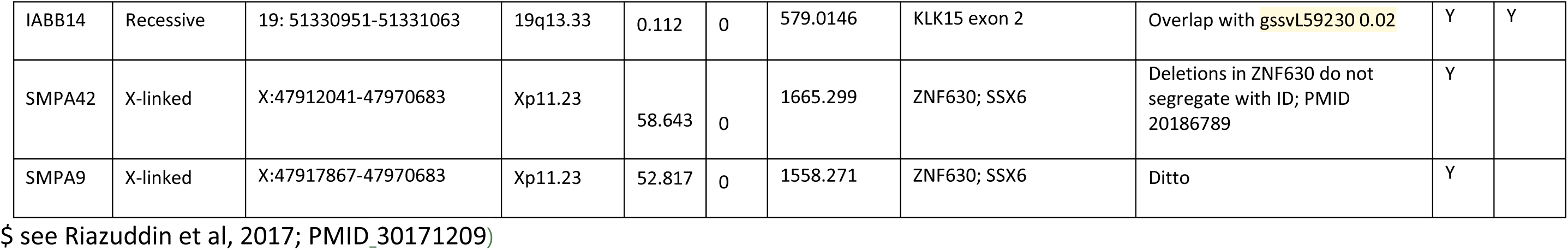
CNV summary: validated biallelic loss CNVs called by CNVpartition or ChAS. Biallelic losses called from microarray data by PennCNV and from WES data by CLAMMS (81)) did not validate using IGV, and hence are not included. Comparison with control population CNVs used the Database of Genomic Variants (DGV) Gold, accessed through the Decipher genomic browser (www.deciphergenomics.org), also through gnomAD. Coordinates given using hg19.

| Family | Mode of Inheritance | (hg19) | Cytoband | Size (Kb) | CN | CNV confidence | Genes | FREQ IN DGV GOLD STANDARD, gnomAD, or rare? | IGV | PCR |
| --- | --- | --- | --- | --- | --- | --- | --- | --- | --- | --- |
| SMPA4 | Recessive | 14:24,454,924-24,464,325 | 14q11.2 | 9.402 | 0 | 345.9659 | DHRS4 exon 8; DHRS4L2 exons 1-4 | Partial overlap with gssvL33876<br>FREQ 0.0029 | Y | Y |
| SMPA4 | Recessive | 20: 43836785-43836964 | 20q13.12 | 0.18 | 0 | 228.5597 | SEMG1 exon 2, 180bp, 60 amino acids | Partial overlap with gssvL72784<br>FREQ 0.0038 | Y | Y |
| SMPA8 | Recessive | 2:196,799,394-196,801,362 | 2q32.3 | 1.969 | 0 | 139.6294 | DNAH7 exon 20-21 | NONE | Y | Y |
| SMPA3 | Recessive | 5:140,224,038-140,242,319 | 5q31.3 | 18.282 | 0 | 1417.866 | PCDHA9, PCDHA10 (SFARI score 2) | Also Hom in mother | Y | Y |
| IAH2 | Recessive | 5:140,224,038-140,242,319 | 5q31.3 | 18.282 | 0 | 1417.866 | PCDHA9, PCDHA10 (SFARI score 2) | Also Hom in father | Y | Y |
| IAU79 | Recessive | 20:1,559,259-1,559,330 | 20p13 | 0.072 | 0 | 171.6751 | SIRPB1 exon 2 | Partial OVERLAP WITH COMMON CNV (0.0894) | Y | Y |
| IABB2 | Recessive | 17:3513856- 3514054 | 17p13.2 | 0.198 | 0 | 318.3258 | SHPK exon 7 | Partial OVERLAP WITH 80Kb gssvL47583 (0.00274) | Y | Y |
| IABB14 | Recessive | 15:85186877-85186894 | 15q25.2 | 0.018 | 0 | 55.68398 | WDR73 exon 8<sup>s</sup> | rs760394400; gnomAD MAF 0.1672 | Y | Y |
| IABB14 | Recessive | 19:41356319-41386033 | 19q13.2 | 29.714 | 0 | 579.0146 | CYP2A7 6 exons | gssvL58578 0.002 Plus gssvL58575 0.0017 | Y | Y |
| IABB14 | Recessive | 19: 51330951-51331063 | 19q13.33 | 0.112 | 0 | 579.0146 | KLK15 exon 2 | Overlap with gssvL59230 0.02 | Y | Y |
| SMPA42 | X-linked | X:47912041-47970683 | Xp11.23 | 58.643 | 0 | 1665.299 | ZNF630; SSX6 | Deletions in ZNF630 do not segregate with ID; PMID 20186789 | Y |  |
| SMPA9 | X-linked | X:47917867-47970683 | Xp11.23 | 52.817 | 0 | 1558.271 | ZNF630; SSX6 | Ditto | Y |  |

\$ see Riazuddin et al, 2017; PMID\_30171209)

There are also three large, multi-genic *de novo* CNV losses among the trios, which may also be pathogenic (Supplementary Table S1).

### Neuroanatomical Enrichment Analysis

Testing for regional and cell-type specific expression did not indicate clear anatomical targets with higher expression of the candidate genes. Enrichment in the cerebellum and cerebral cortex was observed in the BrainSpan human developmental atlas, but this was not supported by either the mouse or adult human atlases. We found no consistent neuroanatomical expression pattern for the identified genes, suggesting heterogeneity of neural circuits disrupted.

### High consanguinity coefficient correlates with identification of biallelic variants

Using microarray data we calculated the consanguinity coefficient F_RoH_ using autosomal RoH for 104 of the 115 trios. This included 13 trios for which LoH scores were converted to F_RoH_ using the equation F_RoH_=(LoH – 0.0111)/1.034 (see Methods and Supplementary Materials). Comparison of the degree of consanguinity (using F_RoH_) between 33 trios with biallelic variants versus 20 trios with *de novo* and X-linked variants indicated a strongly significant shift (unpaired *t* test (two-tailed): *p* < 0.0001; df=51; mean F_RoH_ for biallelic variants= 0.09003, S.E.M.=0.00672, N=33; mean F_RoH_ for autosomal *de novo* and X-linked = 0.02955, S.E.M. = 0.00737, N=20). This is also true for variants identified in known ASD/ID genes (*p* =0.0045; df=22; mean F_RoH_ for biallelic 0.08146, N=13, S.E.M.=0.01019; mean F_RoH_ for autosomal *de novo* and X-linked = 0.03282, N=11, S.E.M.=0.01159).(Figure 2A). Comparison also showed clear differences in F_RoH_ levels between different cohorts, and country of ascertainment (Figure 2B & C).

**Figure 2:**
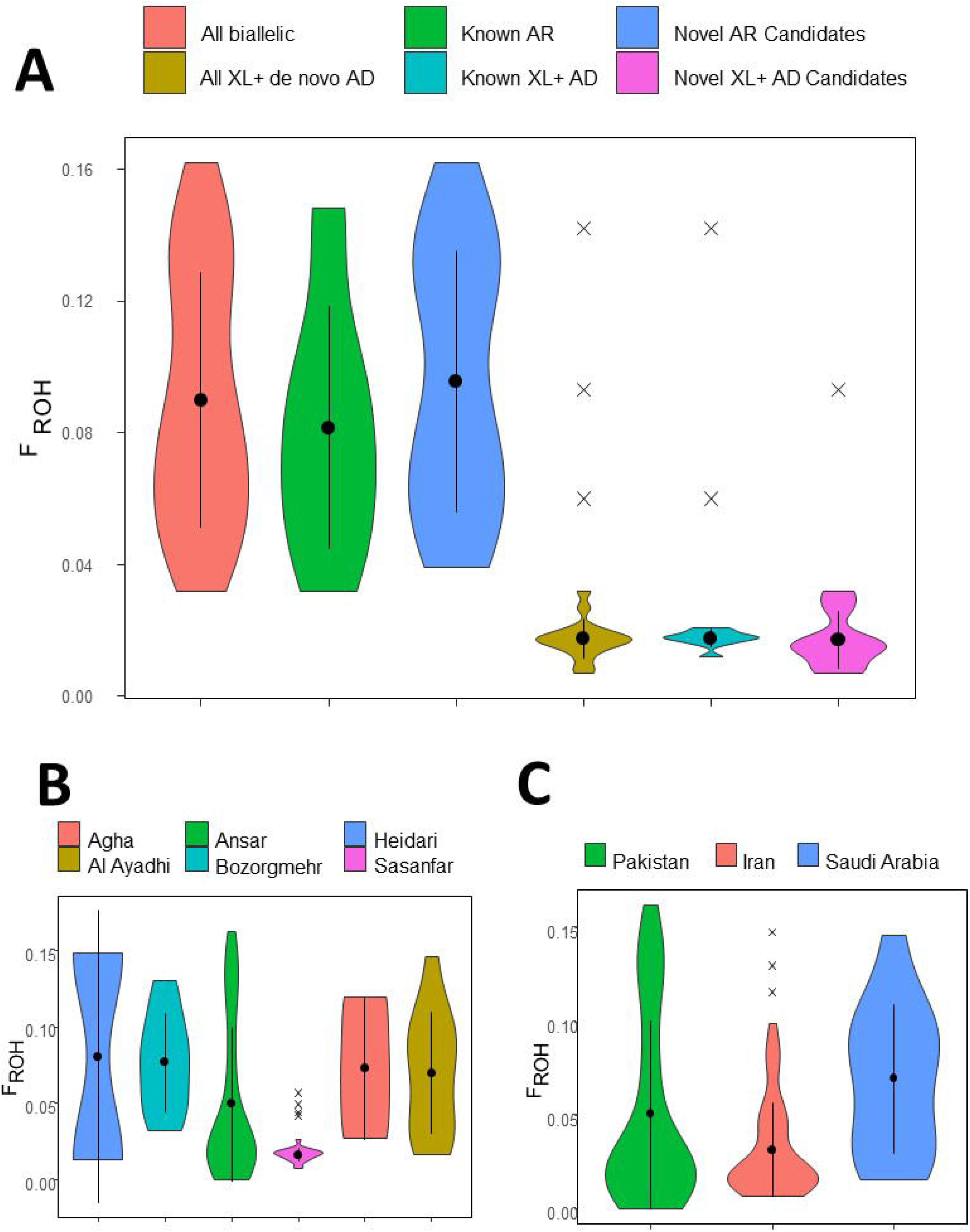
Violin plots comparing consanguinity coefficient F_roh_, for A: different categories of variant identified: All biallelic variants (N=27 observations), variants in known AR ID and/or ASD genes (N=10 observations), variants in novel candidate genes (N=17 observations), all X-linked (XL) variants plus *de novo* autosomal dominant (AD) variants (N=18 observations), variants in known XL and AD genes (N=11 observations), and variants in novel candidate XL and AD genes (N=7 observations). Unpaired *t* test (2-tailed) for comparison of means showed comparison for i) all biallelic versus all *de novo* autosomal plus X-linked to be extremely significant (p<0.0001, *t*=5.8309, d.f.=51), ii) known AR versus AD ID and/or ASD genes to be strongly significant (p=0.0045, *t*=3.1642, d.f.=22), iii) novel candidate AR versus AD genes to be strongly significant (p<0.0001, *t*=5.4795, d.f.=26). No significant difference in mean F_roh_ was seen for comparisons between known and novel candidate AR genes, or between known and novel AD plus XL genes; **B:** different cohorts, categorized by last name of principal investigator for each collection (Agha, N=3, Ansar, N=52, Heidari, N=2, Al Ayadhi, N=13, Bozorgmehr, N=10, Sasanfar, N=24 observations). While F_roh_ distribution was similar for most cohorts, the Iranian cohort from Sasanfar had a significantly lower mean than most other cohorts; **C:** grouped by country of origin (Pakistan, N=55, Iran, N= 36, Saudi Arabia=13 observations). Comparison of mean F_roh_ was non-significant for Pakistan versus Iran and versus Saudi Arabia, but significant for Iran versus Saudi Arabia, but non-significant after correction for multiple testing (p=0.0206, t=2.396, d.f.=36). Plots were prepared using R software using ggplot2, and show mean, standard deviation, and outliers marked by “X”.

## Discussion

Many recent studies involving NGS in ASD have involved large cohorts, focusing predominantly on dominant/*de novo* inheritance. This focus is largely driven by the fact that autosomal recessive variants in novel genes are more difficult to identify in the outbred population. Identification of candidate AR ASD genes through the study of consanguineous families should help streamline the identification of biallelic mutations through clinical genetic screening in outbred populations. Using whole exome trio analysis in consanguineous families, we have enriched for recessive variants to assess the role these variants play in ASD in populations where endogamy is common. This study of 115 trios has identified WES variants for 32 trios in genes previously associated with ASD or other neurodevelopmental disorders, resulting in a diagnostic yield of 28% (18 autosomal recessive, seven autosomal *de novo* plus one maternally inherited, and six X-linked variants in known genes). With a further three trios with large, multi-genic, loss CNVs validated experimentally as *de novo* and putatively pathogenic (Table 6), this yield increases to 30%. There were many other variants identified that met filtering criteria which could potentially represent novel ASD genes or targets. We present 32 new candidate autosomal recessive variants/genes, including a biallelic variant in *SHH*, a gene associated with autosomal dominant holoprosencephaly. Within this cohort we identified variants in five genes that are associated with known metabolic syndromes (4.3% of the diagnosis cohort): *AGA*, *ASL*, *HTRA2*, *ASPA,* and *MTHFR*. These genes may represent better clinical management opportunities for patients, and potentially better therapies.

A number of the known ID genes identified among the 115 ASD trios have also previously been reported for ASD or ASD-like features, e.g. *CC2D1A* (55,62,63), *VPS13B* (33), *DEAF1* (54), *ZNF335* (64), *ZNF292* (65,66), *MYT1L* (66,67), and *SCN2A* (59,68,69). Of the candidate genes identified here, MSSNG, SFARI, and other datasets provide putative support for the biallelic genes, with homozygous variants in *DNAH7*, *ANO10*, *DNAH8*, *CAPSL*, *CYP2A7*, and *SIRPB1*, and putative compound heterozygous variants in *DNAH7*, *DNAH8, SCN10A*, and *WDR90*, and for *de novo*/dominant genes *NCL*, *SLAMF7*, *ADGRF2*, *DGKZ*, *ATP2B1*, *CBFA2T3*, *RETN*, and *PPIL2* (Supplementary File 1).

For three of the known ASD or ID genes reported here, namely *ZNF292*, *SCN2A*, and *MECP2,* mutations have been identified in two unrelated trios from our cohort. Mutations in the known ASD gene *SCN2A* (68) were identified in two trios-both *de novo*. In addition, mutations were identified in three other voltage-gated sodium channel members, including a biallelic stop gain mutation in *SCN10A*, in which heterozygous gain of function missense mutations have previously been linked to familial episodic pain syndrome 2 (FEPS2; MIM 615551)(70), as well as *de novo* missense mutations in *SCN5A* and *SCN8A*. The *SCN8A* variant is predicted as damaging by all algorithms tested, and mutations in this gene have previously been linked to AD developmental and epileptic encephalopathy 13 (DEE13; MIM 614558). The *SCN5A* variant was predicted as benign by several of the algorithms and is less likely to be related to ASD in this trio. The Arg267* mutation identified in *MECP2* in trio SMPA78 has been frequently reported in cases of Rett syndrome. However more C-terminal truncating mutations such as the Thr412Asnfs*5 mutation in trio IAU54 typically have a significantly milder phenotype(71).

For some of the autosomal recessive candidate genes, there is additional support from animal models. For instance, *Rasal2* (MGI:2443881) and *Vps1*6 (MGI:2136772) knockout mice have behavioral/neurological and nervous system phenotypes (http://www.informatics.jax.org/; Supplementary File 1). Additional discussion of mouse models relevant to the candidate genes is provided in Supplementary Materials.

There are several trios for which there are two or more candidate variants (Supplementary File 1). For some of these, there may be a variant that clearly delivers a more plausible etiopathological explanation. For instance, for trio SMPA3 the biallelic change in *MTHFR*; for trio IAH2, the biallelic LoF change in *TECPR2* (SPG49; MIM 615031; ID (30); hereditary sensory neuropathy with ID (72)); for trio IABB2, the biallelic LoF variant in (NEDHELS; MIM 617171); for trio IABB14, the biallelic splice mutation in ASPA (Canavan disease; MIM 271900).

Since our initial methodology used relatively stringent MAF cut-off criteria for biallelic variants (<0.001), we attempted to justify this level by reiterating the analysis but with lower thresholds (<0.01). Using the more relaxed criteria resulted in just two additional candidate variants, in the genes *CLCA4* (gnomAD South Asian MAF=1.72E-3), and *RASAL2* (gnomAD South Asian MAF=6E-3), but none in known neurodevelopmental genes. Our methodology was particularly stringent regarding *de novo* nonsynonymous variants, owing to the large number of likely spurious calls, and the inclusion of only variants predicted as damaging by all algorithms used may have excluded some *bona fide* mutations. Other variants, particularly intronic or intergenic, would also likely be missed by our approach. Also, a proportion of exons, and particularly in GC-rich regions, are either missed or have low coverage in WES. Follow-up with whole genome sequencing could be considered as a possible next step. However, parsing and assessing rare non-coding variants is particularly challenging.

It was recently estimated in the DDD cohort that recessive variants make up 3.6% of diagnoses, whereas *de novo* variants contribute 48.6% of variants (38). The same study also compared homozygous variants in a narrowly defined group of Pakistani Ancestry in the British Isles (PABI), which is most like our cohort (38). The PABI cohort had 356 (333 undiagnosed) probands, 110 of whom (30.9%) had biallelic coding candidate variants, approximately half of which are in known DD genes and half in novel candidates (38). For comparison, our study reports very similar yields for recessive variants, with 15% of probands with biallelic variants in known ASD/ID genes (17/115), or 30% including those with biallelic variants in novel candidate genes (35/115). We report 28 *de novo* variants (20 autosomal, five X-linked, three CNVs) in 17 of 115 individuals (15%), which is somewhat lower than those reported for the PABI cohort (29.8%). This difference is most likely due to differences in the methodologies or stringency used for calling *de novo* variants in the respective studies.

Comparison of consanguinity (F_RoH_) between trios with biallelic variants (either known or candidates) versus trios with *de novo* and X-linked variants indicated a strongly significant shift.. Thus, unsurprisingly, the chances of finding biallelic rare variants is strongly associated with higher consanguinity, and should be a consideration for diagnostic genetic analysis.

Overall, the findings demonstrate the importance of autosomal recessive mutations in ASD in countries, or in cohorts, with high rates of consanguinity. In countries where consanguinity is rare, when screening ASD individuals’ genomes attention should still be paid to the relatedness of the parents, and, for those with a high F-coefficient, attention should be focused on biallelic variants. Additionally, the candidate genes reported here should be examined for both autozygous *and* allozygous mutations in outbred populations, just as would be done for more established autosomal recessive gene disorders.

## Supporting information

Supplementary Materials_Variant Validations

## Data Availability

All data produced in the present study are available upon reasonable request to the authors

## Declaration of Interests Statement

L.F. owns shares in Quince Therapeutics and has received consulting fees from PeopleBio Co., GC Therapeutics Inc., Cortexyme Inc., and Keystone Bio. M.A. is co-founder and director of Institute of Omics and Health Research, Lahore, a research organization. The organization was not involved in the research on which this manuscript is based. All other authors declare no competing interests.

## Acknowledgements

The authors thank the families for their participation in this study. We acknowledge the assistance of Natalie Freeman with PLINK microarray analysis, and of Stephen Pastore with the preparation of violin plots using R software. This work was supported by grants from the Canadian Institutes of Health Research to JBV (#MOP-102758 and #PJT-156402), also a Brain & Behavior Research Foundation (BBRF/NARSAD) Young Investigator Award to NV. RH was supported by a Peterborough K.M. Hunter Charitable Foundation Graduate Scholarship. AR and AH were supported by International Research Fellowship Program scholarships from the Pakistani Higher Education Commission. CS was supported by a Summer University Research Program award from the Institute of Medical Science, University of Toronto. Computations were performed on the CAMH Specialized Computing Cluster. The SCC is funded by The Canada Foundation for Innovation, Research Hospital Fund. Additional acknowledgements are included in Supplementary Materials

## Data and code availability

New variants reported here are available through ClinVar SUB14665852, and SCV accession numbers are included in Supplementary File 1. Microarray genotype data and whole exome sequence data will be made available from the corresponding author upon request.

## Notes

### Author Declarations

Institutional Research Ethics Board approval was received for this study through the Centre for Addiction and Mental Health (CAMH) and other institutional recruiting sites.

### Summary of Updates

Added F-coefficient data and analysis Shortened intro and discussion

