## Supplementary Materials for "Autism spectrum disorder trios from consanguineous populations are enriched for rare biallelic variants, identifying 32 new candidate genes"

**Contents:**

1. **Methods**
2. **Conversion of LoH (Affymetrix CytoScanHD) to Σroh (Illumina CoreExome/PLINK)**
3. **Figure S1**
4. **Additional Discussion**
5. **Additional acknowledgements**
6. **Supplementary References**

### Methods

#### Whole Exome Sequencing, Alignment and Variant Calling

After sequencing, FastQC was used to assess the quality of sequences and to determine the adapter contamination and read quality. Unligated adapters and low quality sequences were removed using FastX Toolkit (Updated Aug 4th 2017: https://github.com/agordon/fastx_toolkit) (1) and Trimmomatic (v 0.33: Updated Mar. 10th, 2015: https://github.com/timflutre/trimmomatic) (2) . Cleaned fastq files were then aligned to the hg19 reference genome using BWA 0.7.17 (3) and variants were called using the GATK best practices for variant calling and quality control (4). Briefly, after alignment, duplicate reads were removed using Samtools version 1.4 (5) and the GATK v4 (6) was used for base recalibration, indel realignment and then variant calling using the Global Haplotyper (6). This created raw Variant Call Format (VCF) files. The variants called per sample were then checked for their transition-transversion ratio which was approximately 2.5.

#### Annotation and Variant Prioritization

VCF files were annotated using ANNOVAR (7), integrating allele frequencies, from gnomAD v2.1.1 (8), The Greater Middle Eastern Variome Server (9) and ExAC (10) , as well as functional pathogenicity scores from Polyphen2 (11) , SIFT (12) and MutationTaster (13) , MutationAssessor (14), FATHMM (15) , and CADD (16). Variants were assessed, firstly, if they passed the quality metrics as set by the Convolutional Neural Network scoring algorithm by GATK (6). Briefly, variants were compared against known variants from dbSNP and a convolutional neural network was trained to filter out poor variant calls based on several metrics (6) and minor allele frequency (MAF) in gnomAD (https://gnomad.broadinstitute.org/) of equal to or lower than $1 \times 10^{-3}$ for autosomal recessive, or $1 \times 10^{-5}$ for autosomal dominant/*de novo*, with a preference for variants that had no homozygotes in the gnomAD non-Neuro Cohort. For autosomal recessive variants, as the MAF used was on the conservative side, reanalysis was also performed using less stringent values ($1 \times 10^{-3}$, $1 \times 10^{-2}$), to see if any variants in known ASD/ID genes were missed under the stringent analysis. Variants were then split into either homozygote or heterozygote categories and then prioritized by the functional impact of mutation type. For example, LoF variants would carry more weight than missense variants leading to assessing stopgain, frameshift, splicing, and missense variants, in that order. The LoF mutations were given highest priority as it was assumed that LoF variants will disrupt protein function more severely and, depending on the position within the transcript, cause nonsense mediated mRNA decay (NMD). The variants were then assessed for their functional pathogenicity score, where variants that met at least two functional prediction scores between Polyphen2 (11), and MutationTaster (13) and CADD (16) were further investigated. Special attention was given to splicing with functional annotation scores being used for splicing events with the addition of FATHMM (15) and dbscSNV (17). Due to the large number of missense variants for autosomal dominant/*de novo* inheritance, stricter criteria were used where a variant would only be considered if it was predicted as pathogenic by all software used including Polyphen2 (11) , MutationTaster (13), MutationAssessor (14), FATHMM (15), CADD(16) and M-CAP(18). The overall prioritization method is outlined in Figure 1. Variants were also analyzed to determine if they are in genes associated with any previous studies reporting an association with ASD or ID. Variants in genes in pathways such as neuron growth and guidance were also considered as a higher priority. Finally, variants were Sanger-sequenced, and segregation checked with both parents for validation.

A second round of annotation was performed independently, on the same vcf file as above, using VarSeq™ (Golden Helix Inc.; Bozeman, MT), with similar filtering steps.

**Splice site annotation**

Standard annotation software has been noted to be suboptimal for splicing variants and cryptic splice sites (19). To improve detection of splicing variants, particularly in neurodevelopmental disorders, SpliceAI was used to identify splicing variants or cryptic splice sites (20). All VCF files were run through the standard SpliceAI algorithm as per documentation and filtered for variants with a SpliceAI score greater than 0.5. Once variants were extracted, they were annotated with ANNOVAR (21) to add additional metadata for biological interpretation. Variants were then binned into heterozygous, X-linked and homozygous and filtered by gnomAD v2.1.1 (22) allele frequency (MAF < 0.01). Variants that survived filtering were then investigated for appropriate inheritance patterns (i.e. *de novo* for heterozygous variants and both parents contributing one allele for homozygous variants).

**Copy Number Variant (CNV) analysis**

In addition to microarray CNV analysis, for WES data we performed CNV analysis using CLAMMS (23), XHMM (24) and CoNIFER (25). The analysis of exome sequencing data for CNVs has drawbacks due to the capture technology used. A majority of these methods employ a read-based exome calling strategy after normalizing for the depth of sequencing across different regions. CLAMMS was used because it offered the ability to create subsets of the samples and construct more specific references through principal component analysis (PCA) of sequencing metrics. All softwares used were run according to standard procedures described in their respective documentation. Filtering was done for quality metrics outlined in the documentation. Once CNVs were called they were also filtered against the gnomAD structural variation call set (26) and the Database of Genomic Variants (DGV: http://dgv.tcag.ca/dgv/app/home). CNVs that overlapped more than 50% with gnomAD or DGV SVs were filtered out. Once the final call set after filtration was created, CNVs were then binned into homozygous, heterozygous or X-linked variants and further validated with qPCR to check whether they followed expected inheritance patterns for their respective zygosity. For instance, homozygous CNVs in the proband would have to have both parents that were heterozygous, and heterozygous proband variants would have to be *de novo*. X-linked variants were also further validated in males, as they lack a second copy of the X chromosome and are thus more likely to be damaging. CNV validations were also performed for family members, to confirm the inheritance pattern of candidate variants.

**Conversion of LoH (Affymetrix CytoScanHD) to Σroh (Illumina CoreExome/PLINK)**

| Trio | Σroh | LoH |
|---|---|---|
| IABB2 | 0.077844 | 0.093851 |
| IABB3 | 0.043444 | 0.056334 |
| IABB4 | 0.032037 | 0.043235 |
| IABB5 | 0.098961 | 0.111812 |

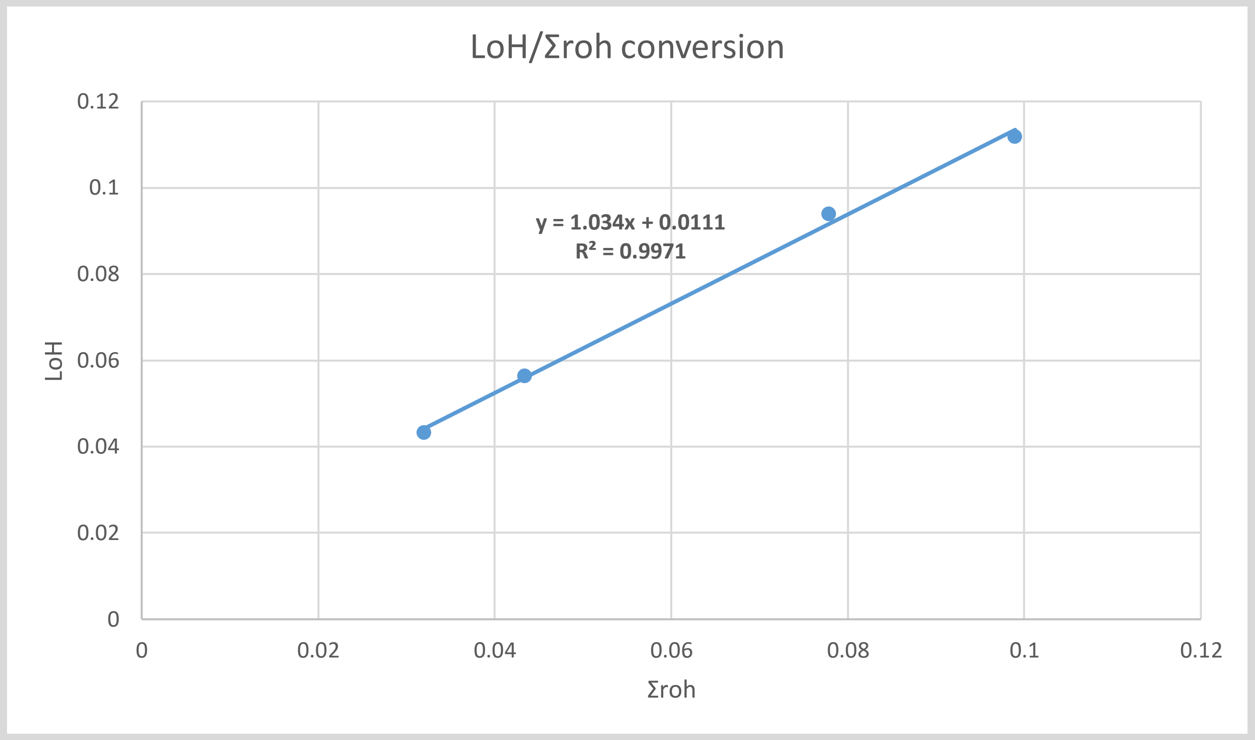


**Figure S1 Additional violin plots (supplementary to Figure 1) with data points shown, comparing consanguinity coefficient $F_{roh}$, for A:** different categories of variant identified: All biallelic variants (N=27 observatons), variants in known AR genes (N=10 observations), variants in novel candidate genes (N=17 observations), all X-linked (XL) variants plus *de novo* autosomal dominant (AD) variants (N=18 observations), variants in known XL and AD genes (N=11 observations), and variants in novel candidate XL and AD genes (N=7 observations). Unpaired *t* test (2-tailed) for comparison of means showed comparison for i) all biallelic versus all *de novo* autosomal plus X-linked to be extremely significant ($p<0.0001$, $t=5.8309$, d.f.=51), ii) known AR versus AD genes to be very significant ($p=0.0045$, $t=3.1642$, d.f.=22), iii) novel candidate AR versus AD genes to be extremely significant ($p<0.0001$, $t=5.4795$, d.f.=26). No significant difference in mean $F_{roh}$ was seen for comparisons between known and novel candidate AR genes, or between known and novel AD plus XL genes; **B:** different cohorts, categorized by last name of principal investigator for each collection (Agha, N=3, Ansar, N=52, Heidari, N=2, Al Ayadhi, N=13, Bozorgmehr, N=10, Sasanfar, N=24 observations). While $F_{roh}$ distribution was similar for most cohorts, the Iranian cohort from Sasanfar had a significantly lower mean than most other cohorts; **C:** grouped by country of origin (Pakistan, N=55, Iran, N= 36, Saudi Arabia=13 observations). Comparison of mean $F_{roh}$ was non-significant for Pakistan versus Iran and versus Saudi Arabia, but significant for Iran versus Saudi Arabia, but non-significant after correction for multiple testing (p=0.0206, t=2.396, d.f.=36). Plots were prepared using R software using ggplot2, and show mean, standard deviation, and outliers marked by “X”. D: all biallelic compared with all XL plus all *de novo* (DN) AD, grouped by country of origin.

A

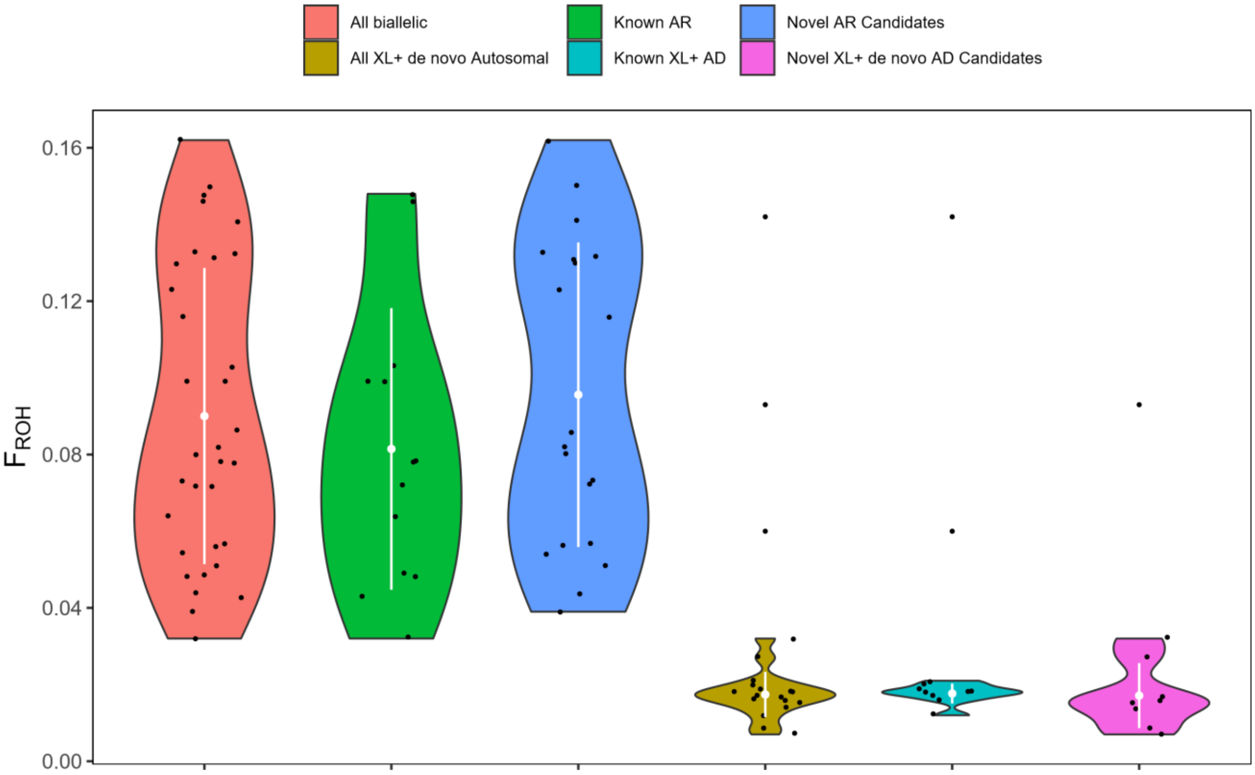

All biallelic
Known AR
Novel AR Candidates
All XL+ de novo Autosomal
Known XL+ AD
Novel XL+ de novo AD Candidates
$F_{ROH}$
0.16
0.12
0.08
0.04
0.00

**B**

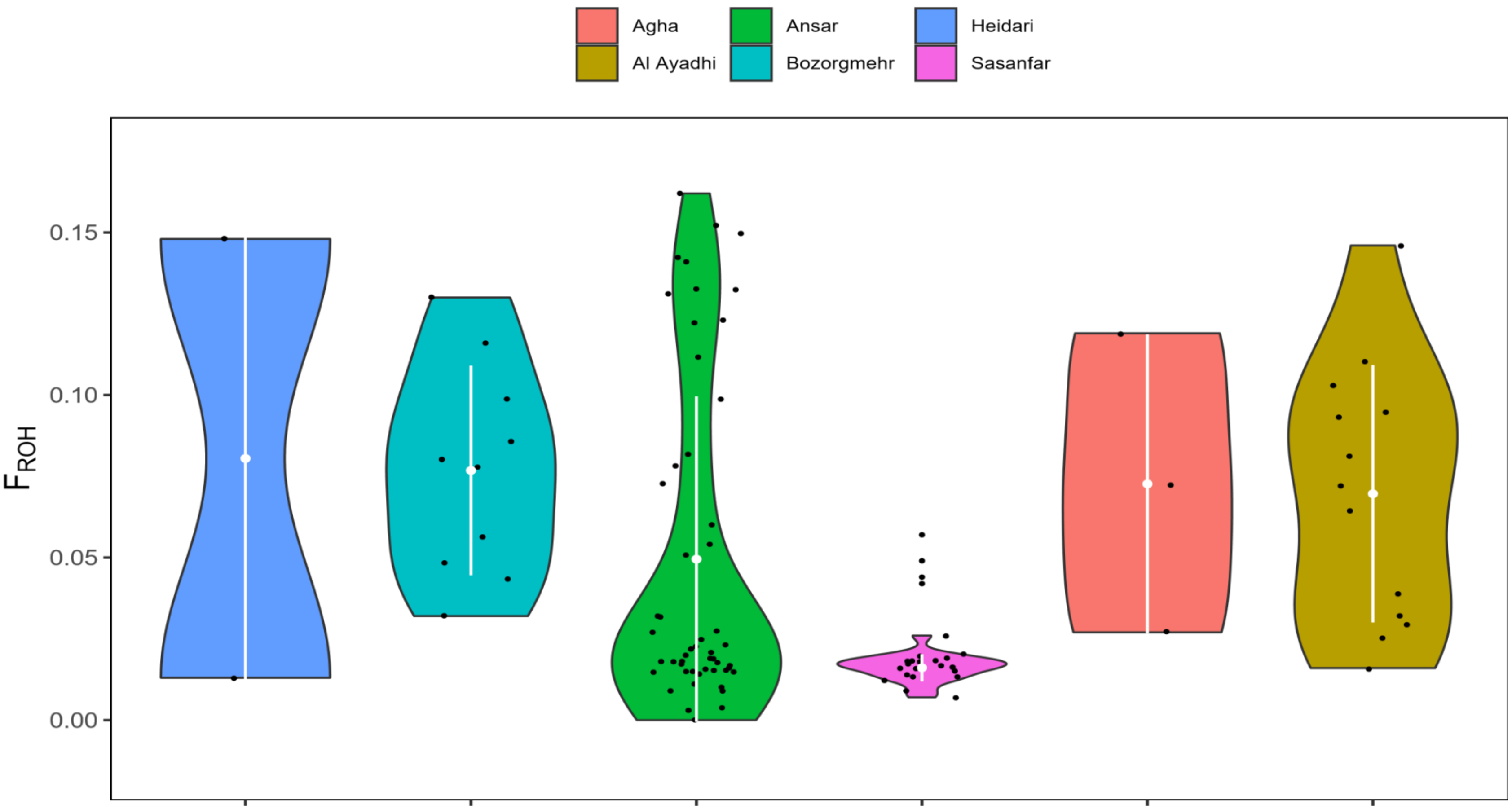
Agha
Al Ayadhi
Ansar
Bozorgmehr
Heidari
Sasanfar
$F_{ROH}$
0.15
0.10
0.05
0.00

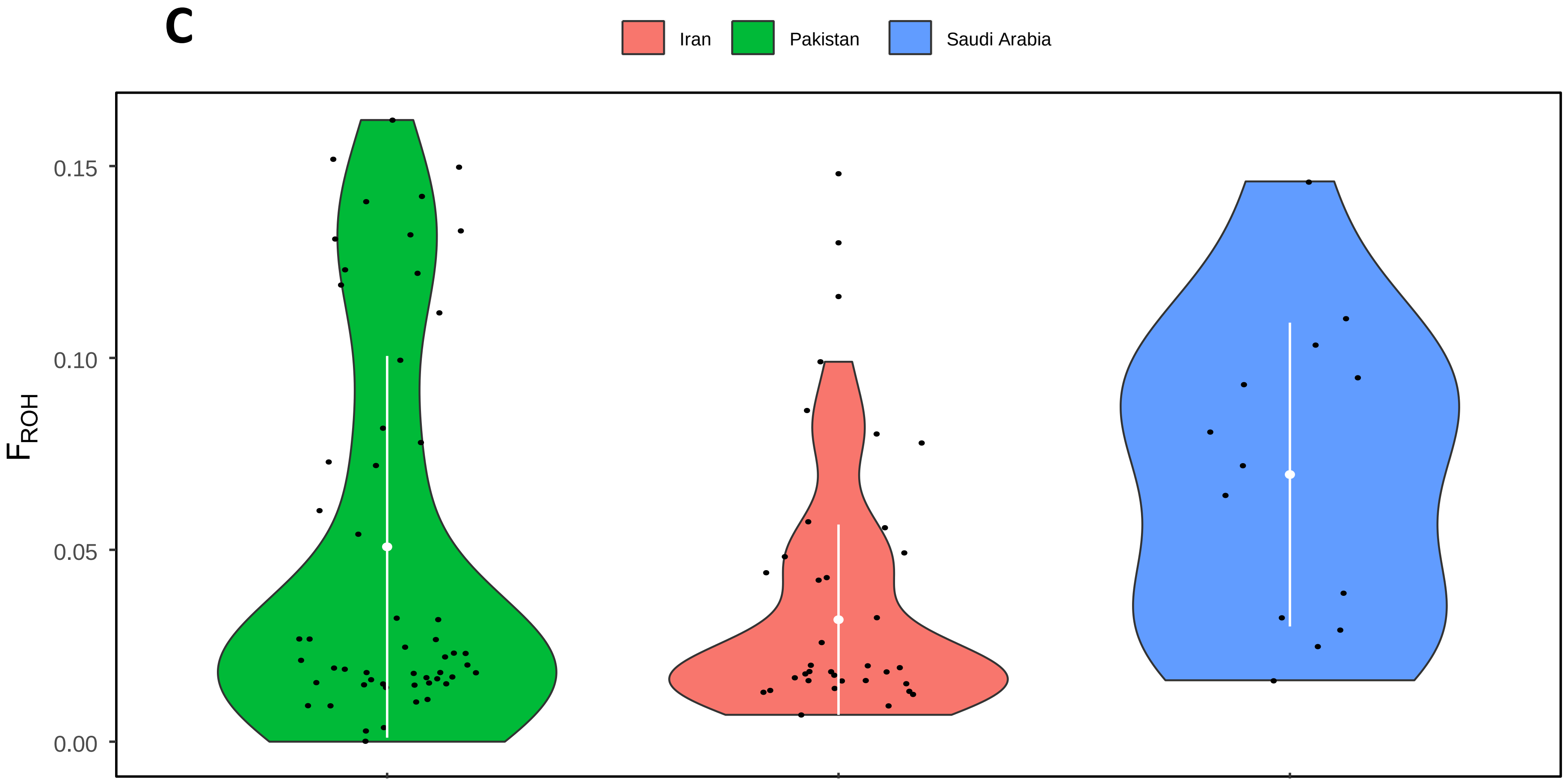
C
Iran
Pakistan
Saudi Arabia
0.15
0.10
0.05
0.00
F_ROH

**D**

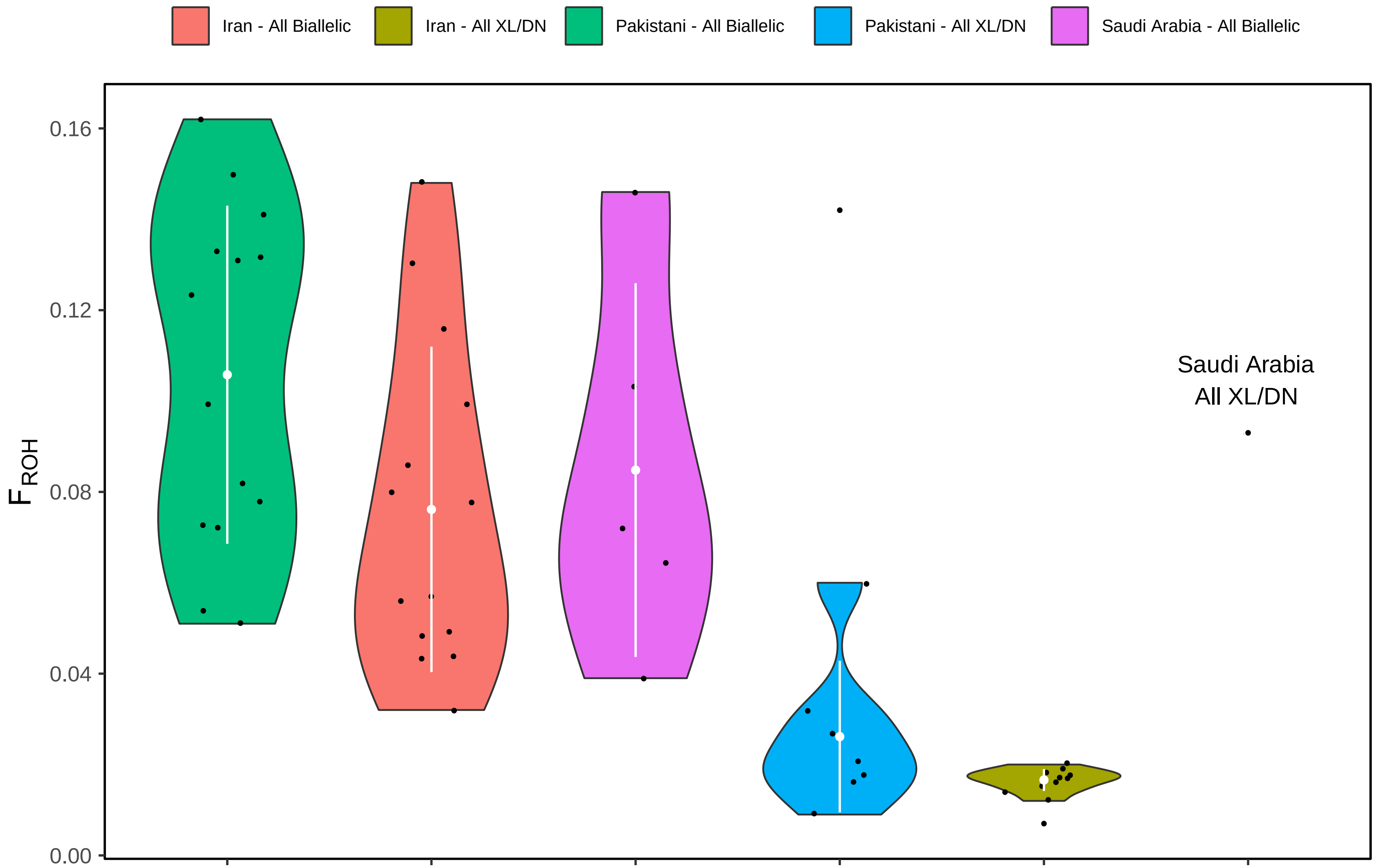

Iran - All Biallelic
Iran - All XL/DN
Pakistani - All Biallelic
Pakistani - All XL/DN
Saudi Arabia - All Biallelic
0.16
0.12
0.08
0.04
0.00
$F_{ROH}$
Saudi Arabia
All XL/DN

### Additional Discussion Points

#### Calcyphosine–related genes

We identified a biallelic nonsynonymous mutation in calcyphosine-like protein gene, *CAPSL*. Biallelic mutations have previously been reported for ID in both of the calcyphosine–related genes, *CAPS* (Harripaul et al, 2018) and *CAPS2* (Anazi et al, 2017). Other than being calcium-binding EF-hand domain proteins, remarkably little is currently known about the role of calcyphosine. Given the possible involvement of all three calcyphosine-related genes in neurodevelopmental disorders, further investigation into this biological pathway is clearly warranted.

#### Animal models

For some of the autosomal recessive candidate genes, there is additional support from animal models. *Rasal2* (MGI:2443881) and *Vps16* (MGI:2136772) knockout mice have behavioral/neurological and nervous system phenotypes (http://www.informatics.jax.org/; Supplementary File 1). Note that VPS16 encodes a vesicle-mediated trafficking protein similar to *VPS13B*, the Cohen syndrome gene. Homozygous knockout of *Ephb1* results in impaired contextual and cued conditioning, as well as abnormal freezing behaviour, and homozygous knockout of *Slc36a1* results in embryonic growth retardation, decreased freezing behaviour, and preweaning lethality (www.mousephenotypes.org; Supplementary File 1). Biallelic knockout of *Dagla* results in decreased brain size, hypoactivity, abnormal behaviour, and decreased thigmotaxis (Supplementary File 1). In general, there is a high rate of neurodevelopmental phenotypes in knockout mice available for the genes with biallelic variants (~70%), and in biallelic or hemizygous knockouts for the X-linked genes (~58%). However, it is a remarkably contrasting story for the *de novo*/dominant variants, for which there is little or no support by way of neurodevelopmental or behavioural phenotypes in heterozygous mouse models (0%). Moreover, where there are biallelic knockout mouse models for genes from the *de novo*/dominant set, the phenotypes described are mainly unrelated to the CNS and behaviour (<10%; Supplementary File 1). This would suggest that the autosomal biallelic (and X-linked) mutations among our cohort are much more likely to be etiopathologically relevant to ASD.

**Additional acknowledgements**

The authors wish to acknowledge the resources of MSSNG (research.mss.ng), Autism Speaks and The Centre for Applied Genomics at The Hospital for Sick Children, Toronto, Canada. We also thank the participating families for their time and contributions to this database, as well as the generosity of the donors who supported this program.

DDD: The DDD study presents independent research commissioned by the Health Innovation Challenge Fund [grant number HICF-1009-003], a parallel funding partnership between Wellcome and the Department of Health, and the Wellcome Sanger Institute [grant number WT098051]. The views expressed in this publication are those of the author(s) and not necessarily those of Wellcome or the Department of Health. The study has UK Research Ethics Committee approval (10/H0305/83, granted by the Cambridge South REC, and GEN/284/12 granted by the Republic of Ireland REC). The research team acknowledges the support of the National Institute for Health Research, through the Comprehensive Clinical Research Network.
