## Supplementary Materials_Variant Validations for "Autism spectrum disorder trios from consanguineous populations are enriched for rare biallelic variants, identifying 32 new candidate genes"

**Supplementary Materials: Supporting documentation for variants:**

**Variants from Table 2. Recessive**

1. SMPA3: *MTHFR*

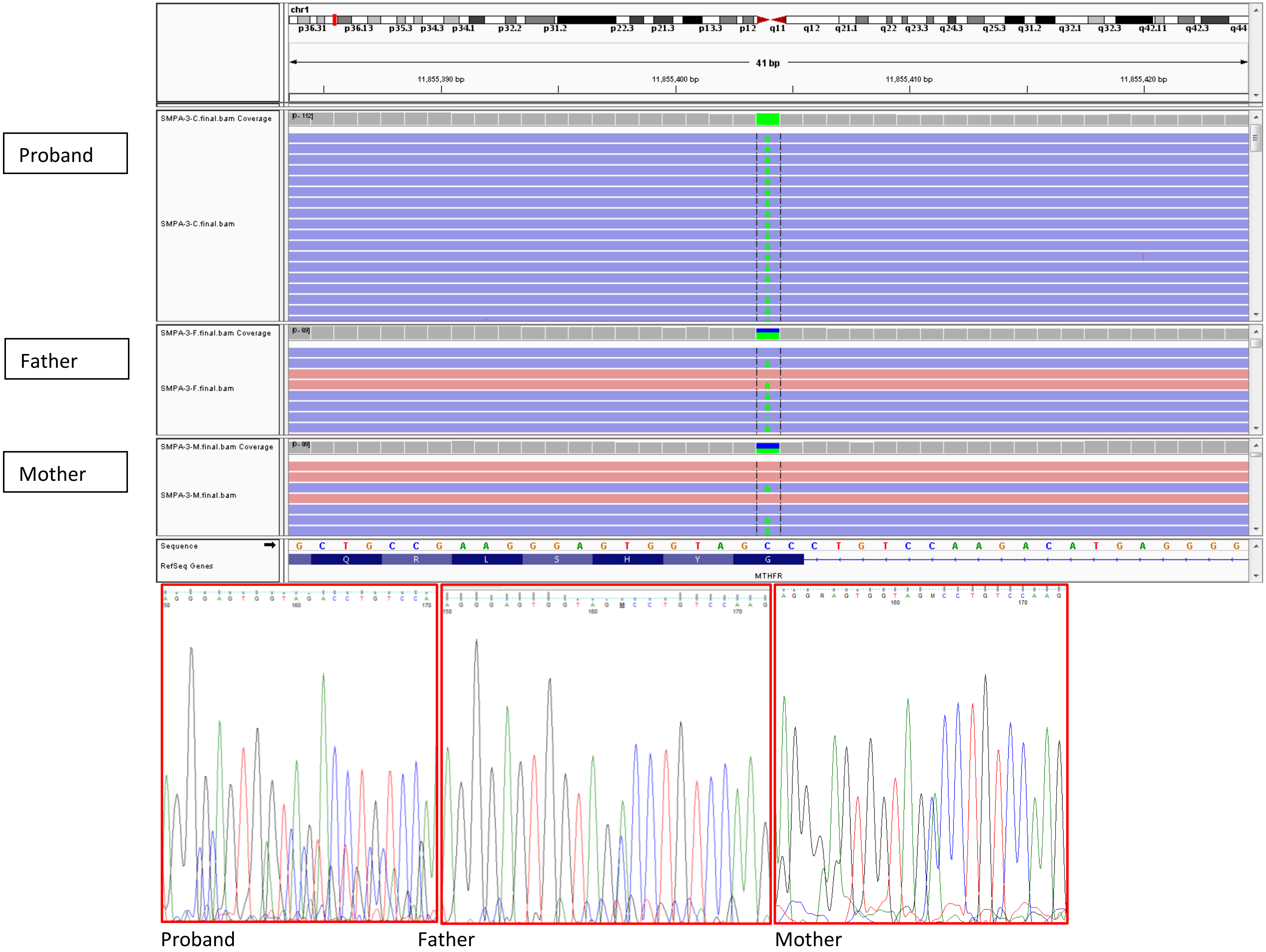

2. IABB16: *CLCA4*

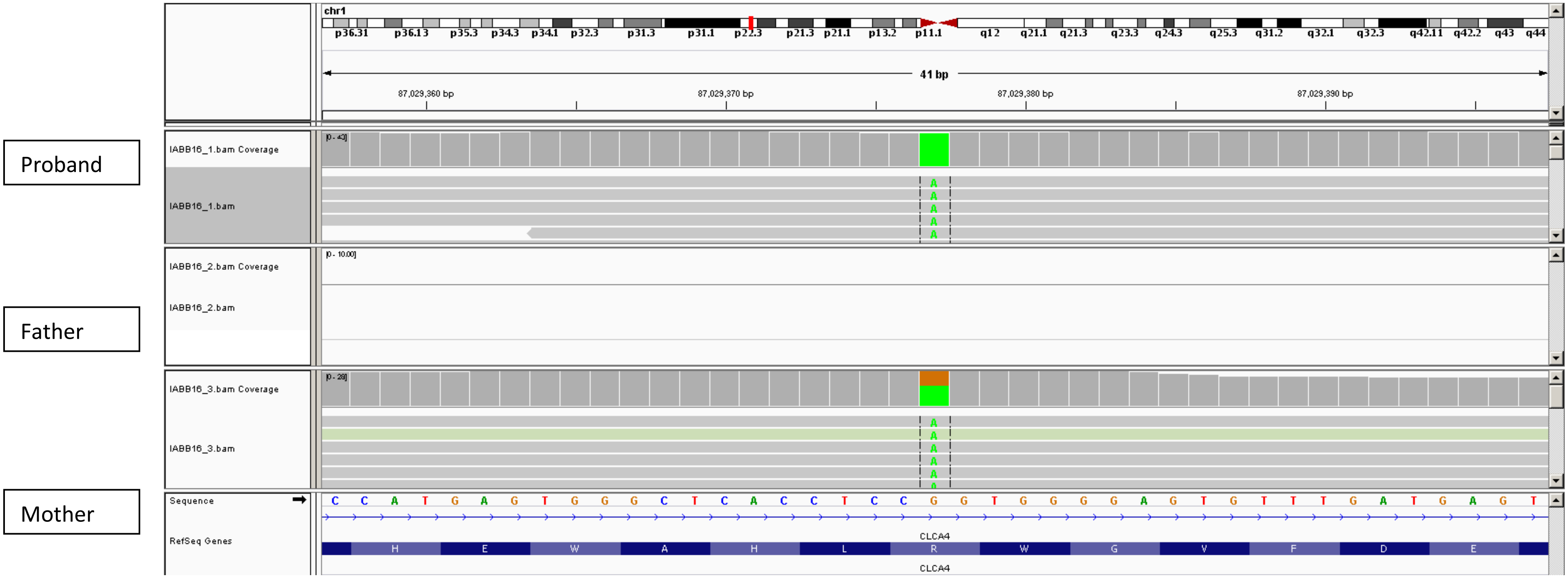


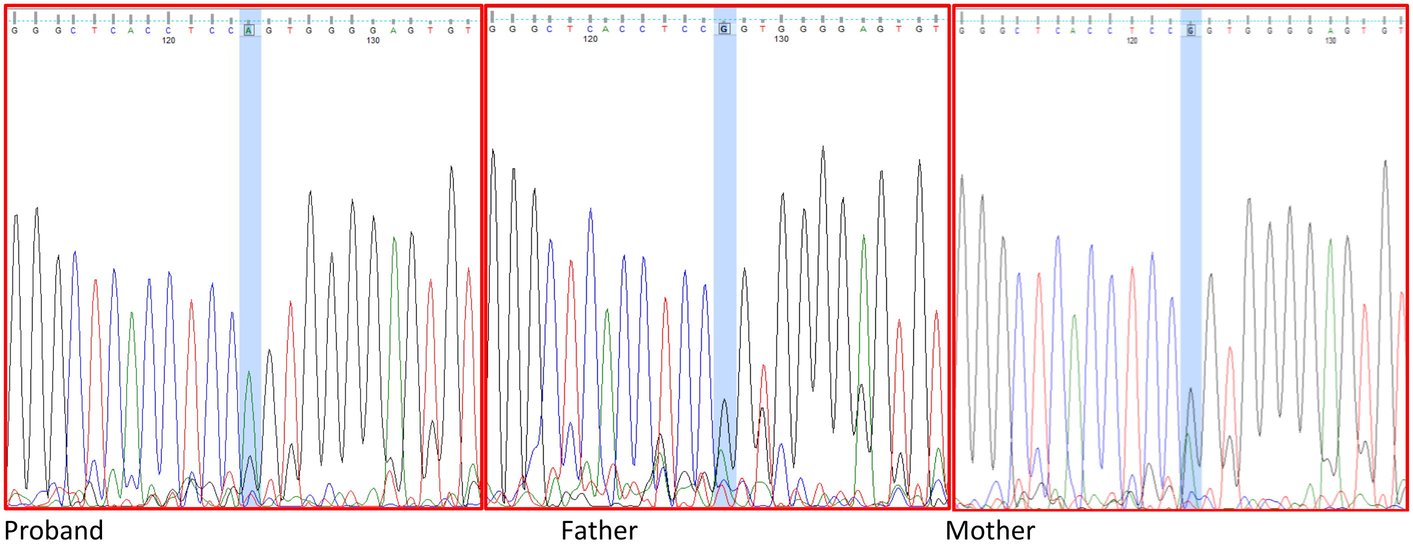

3. SMPA4: *HRNR*

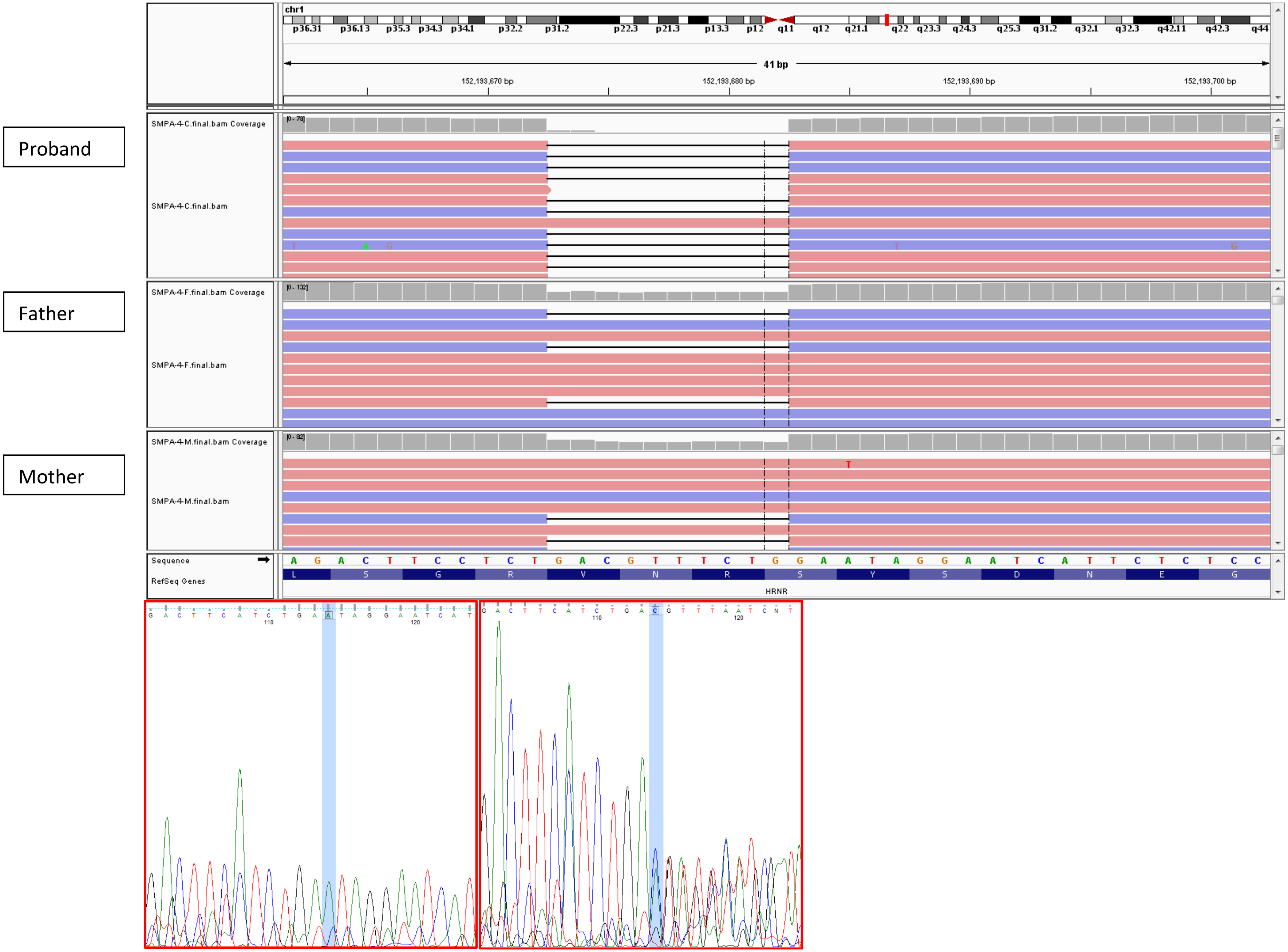


Proband Mother

4. SMPA76: *ILDR2*

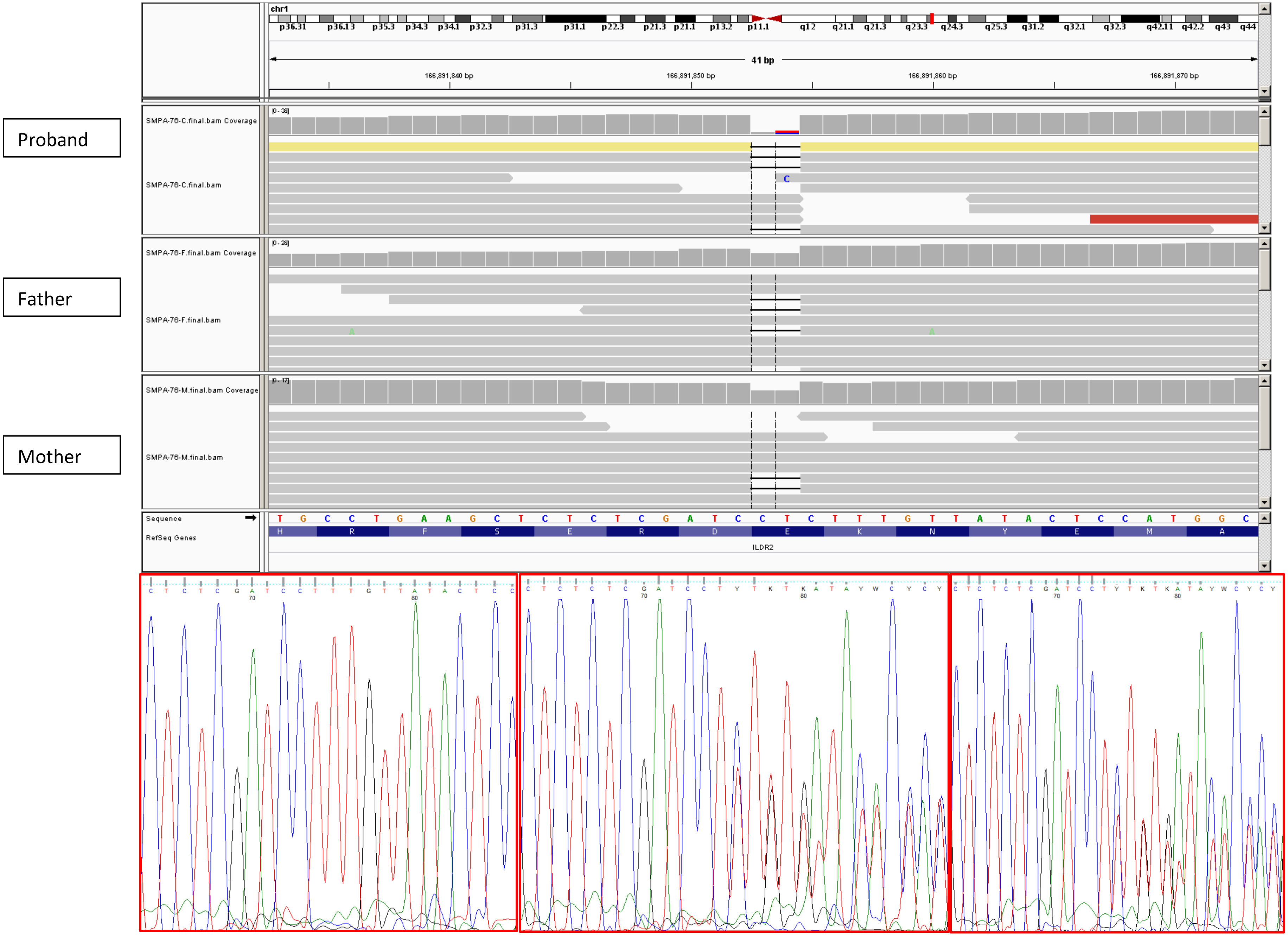


Proband Father Mother

5. SMPA3: *RASAL2*

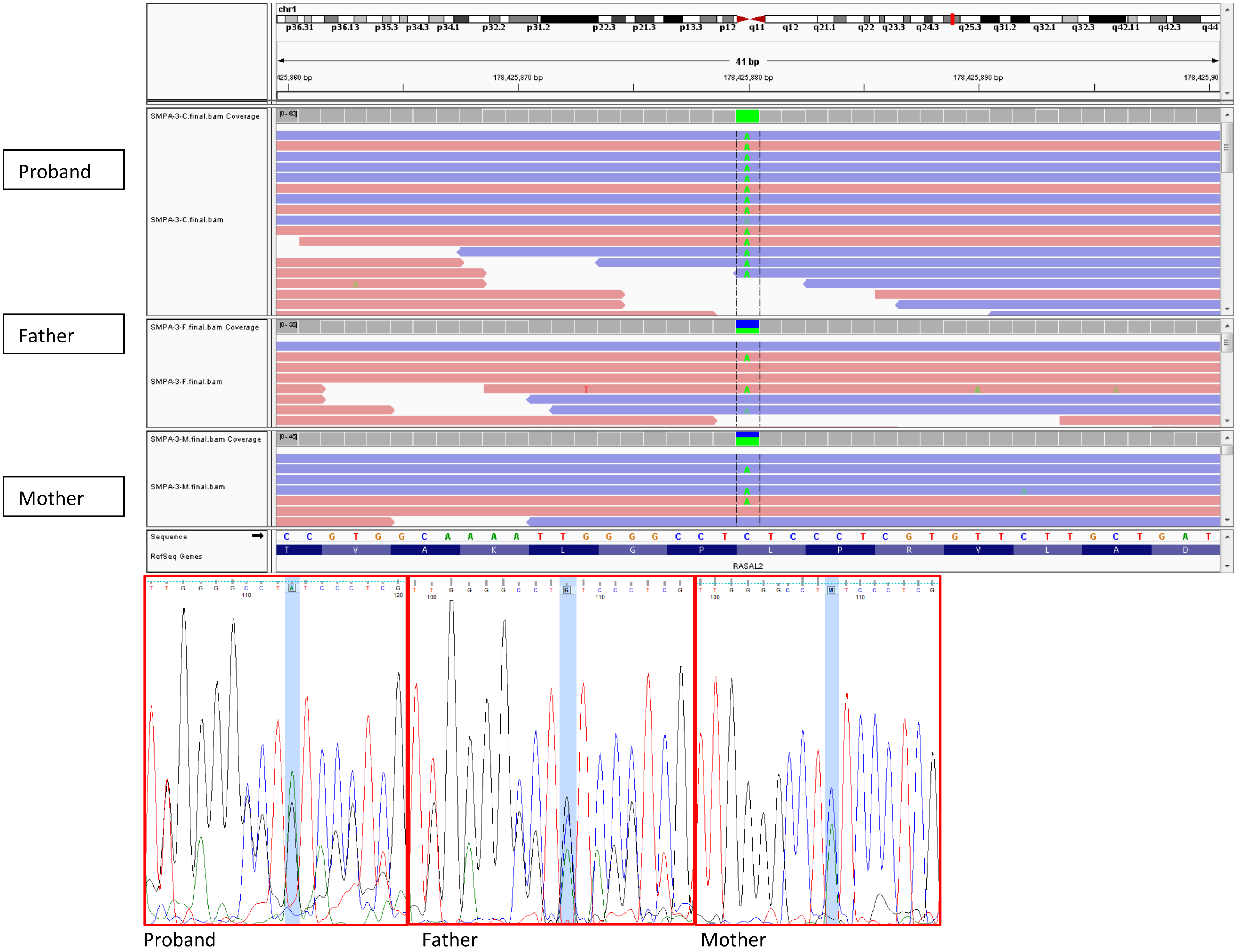

6. SMPA4: *DENND1B*

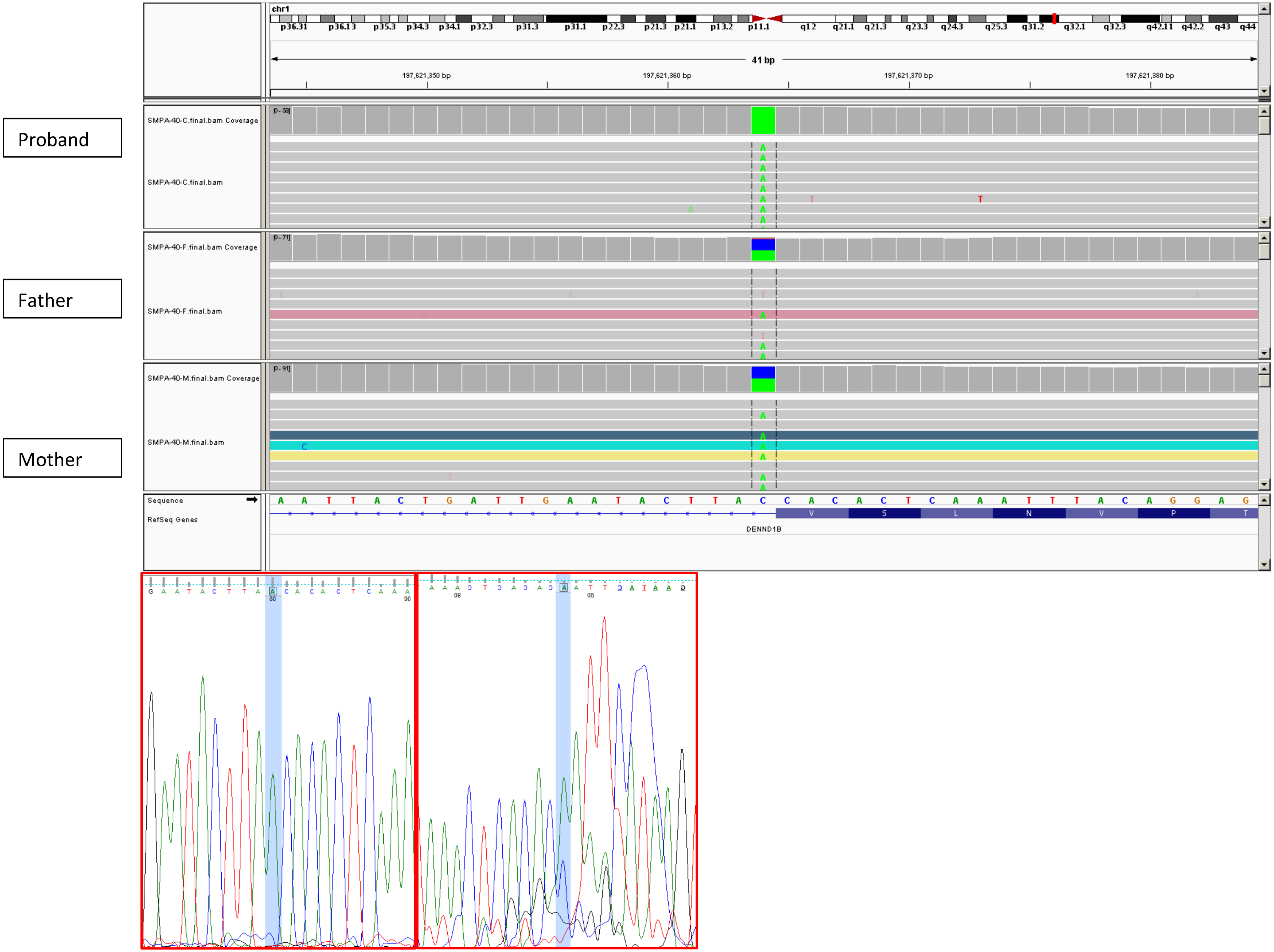


Proband Father

7. Autism8: *HTRA2*

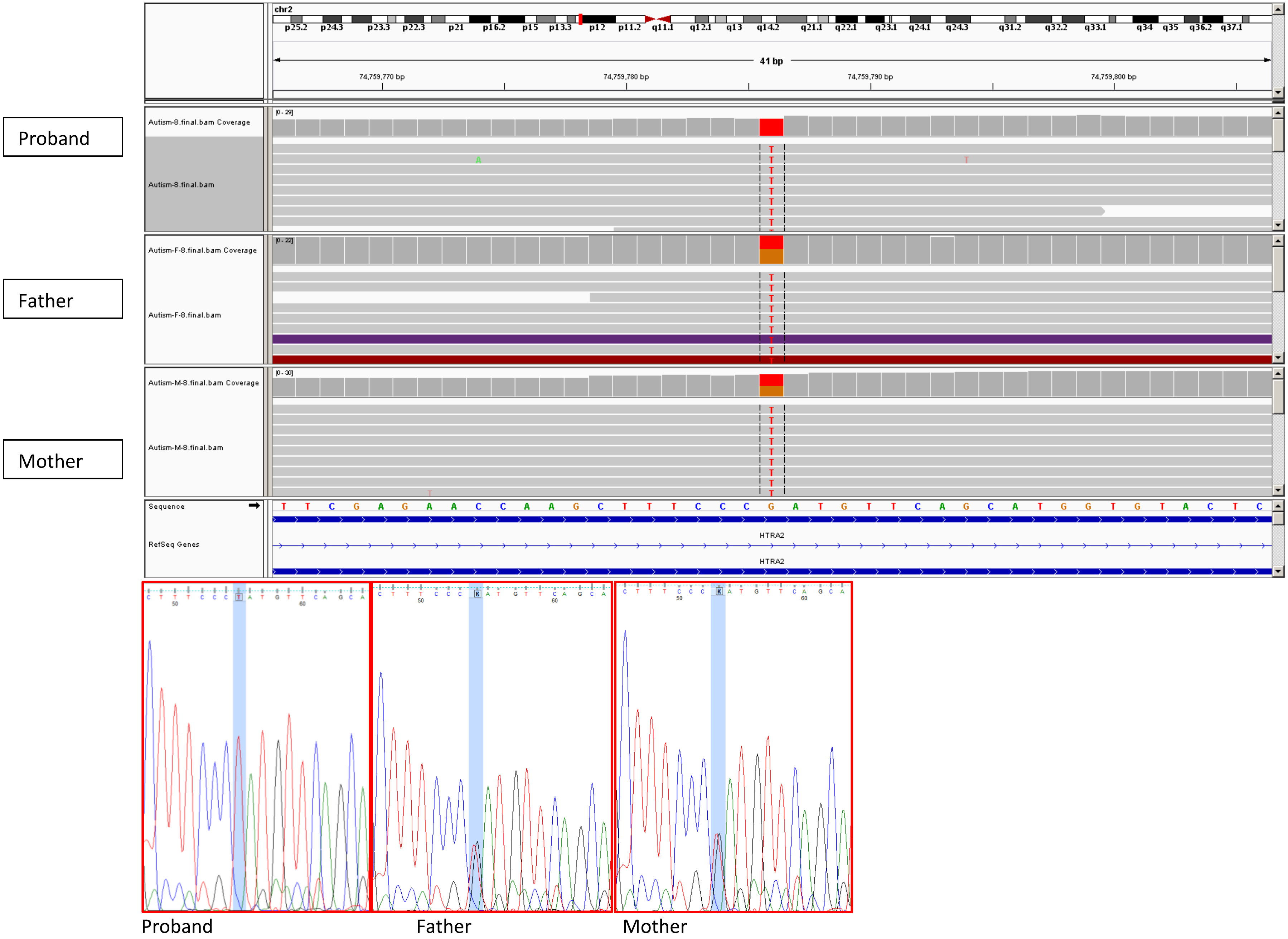

8. SMPA19: *SCN10A*

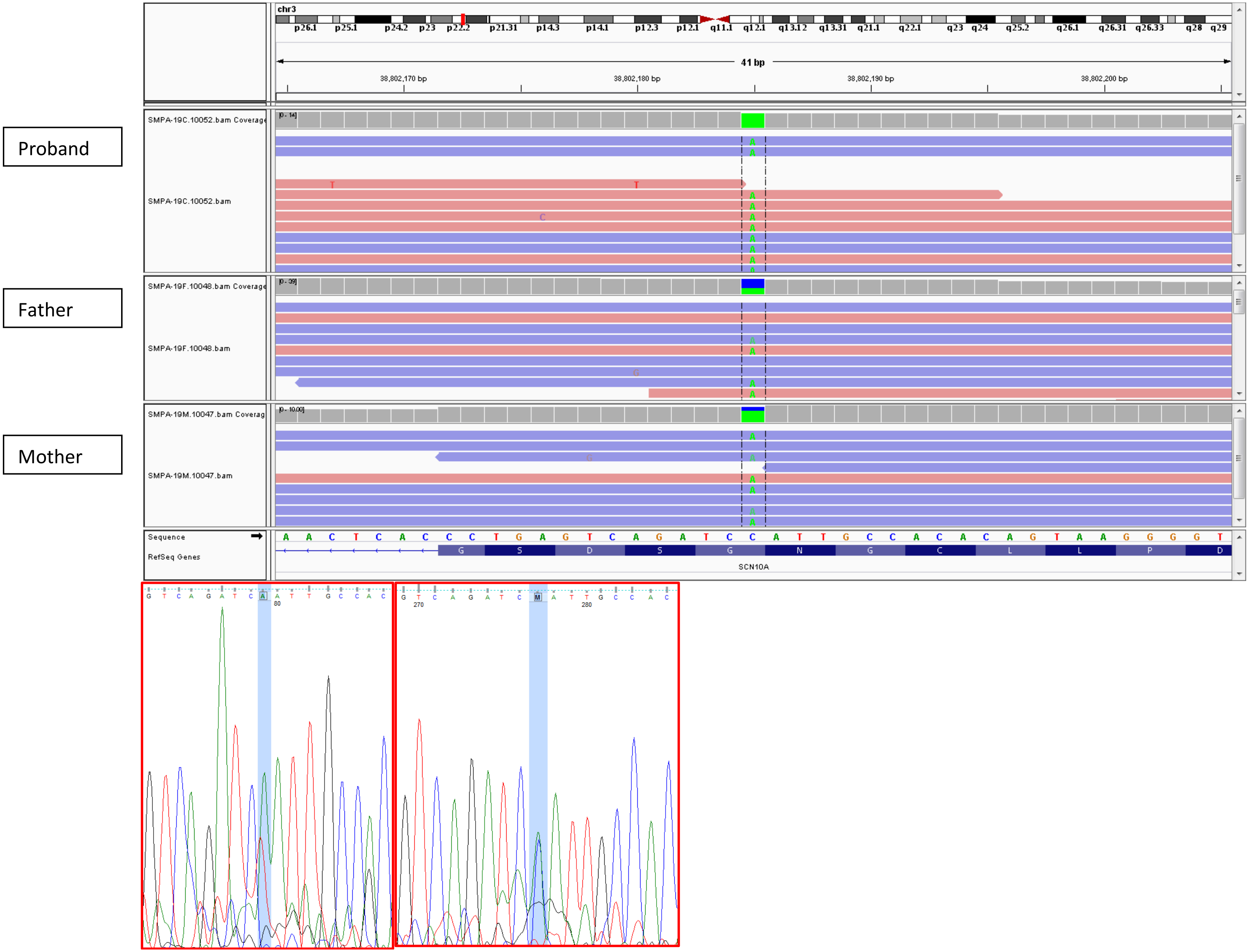


Proband Mother

9. SMPA76: *ANO10*

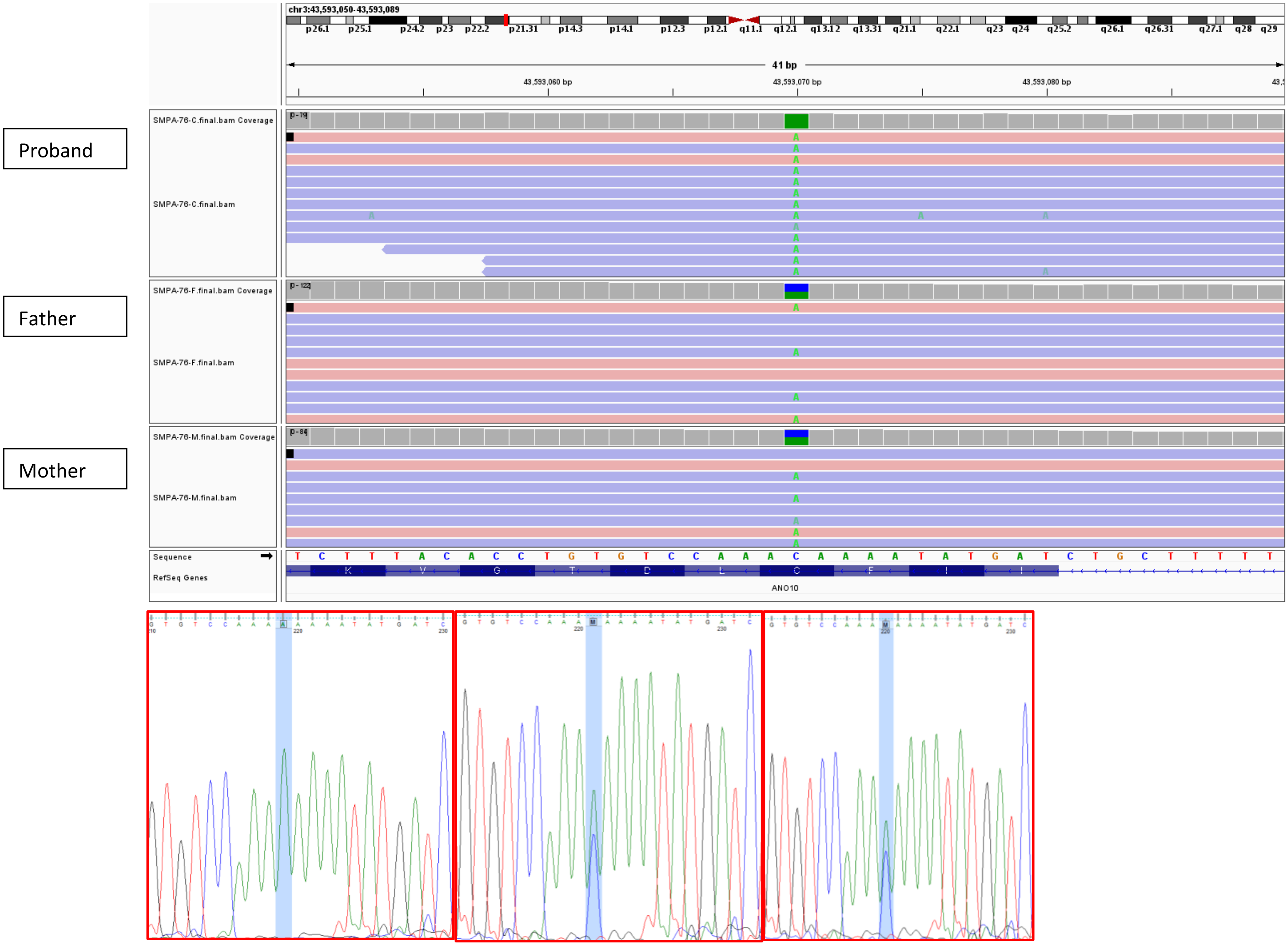


Proband Father Mother

10. SA15: *SETD2*

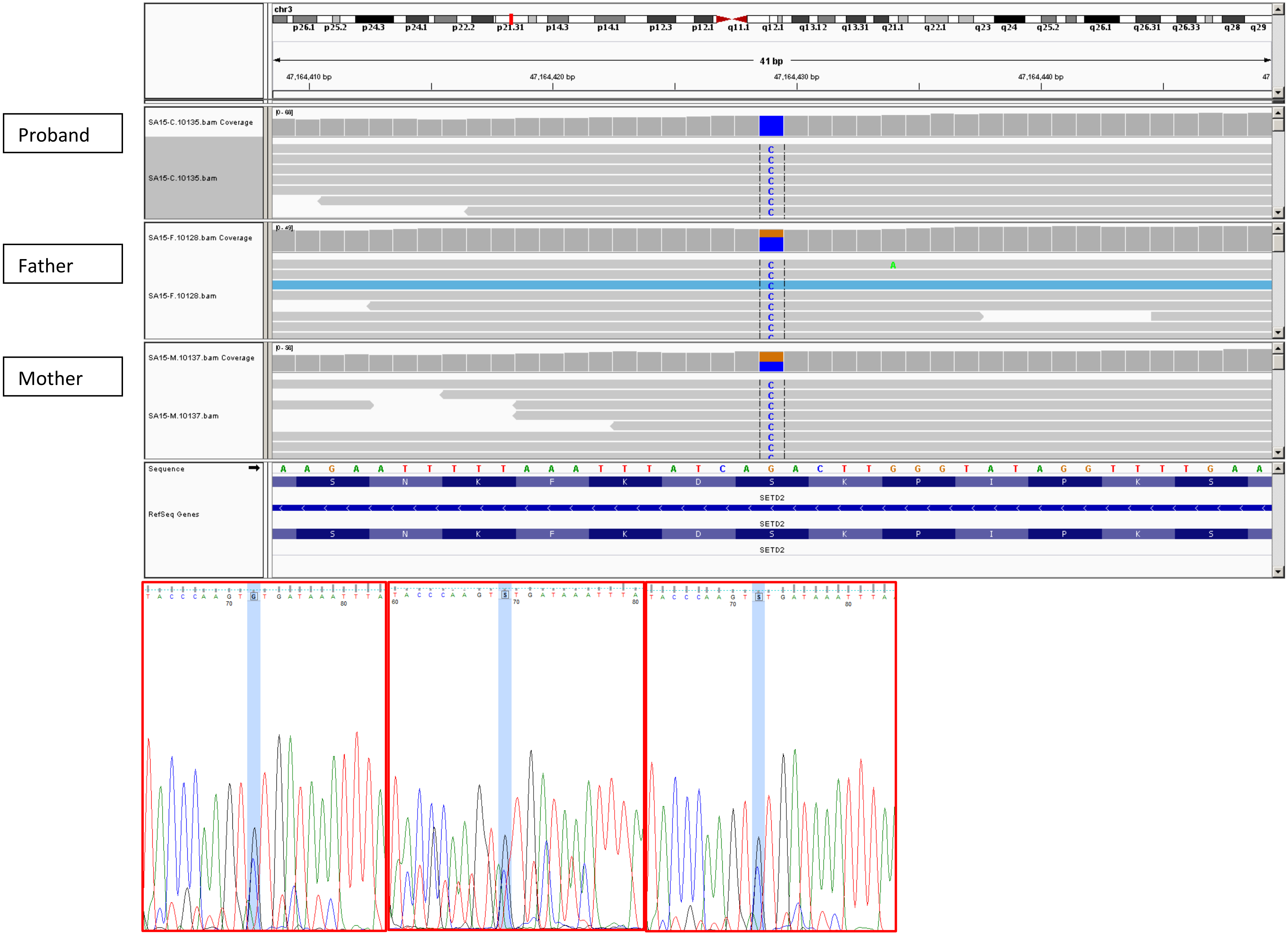


Proand Father Mother

11. SMPA27: *EPHB1*

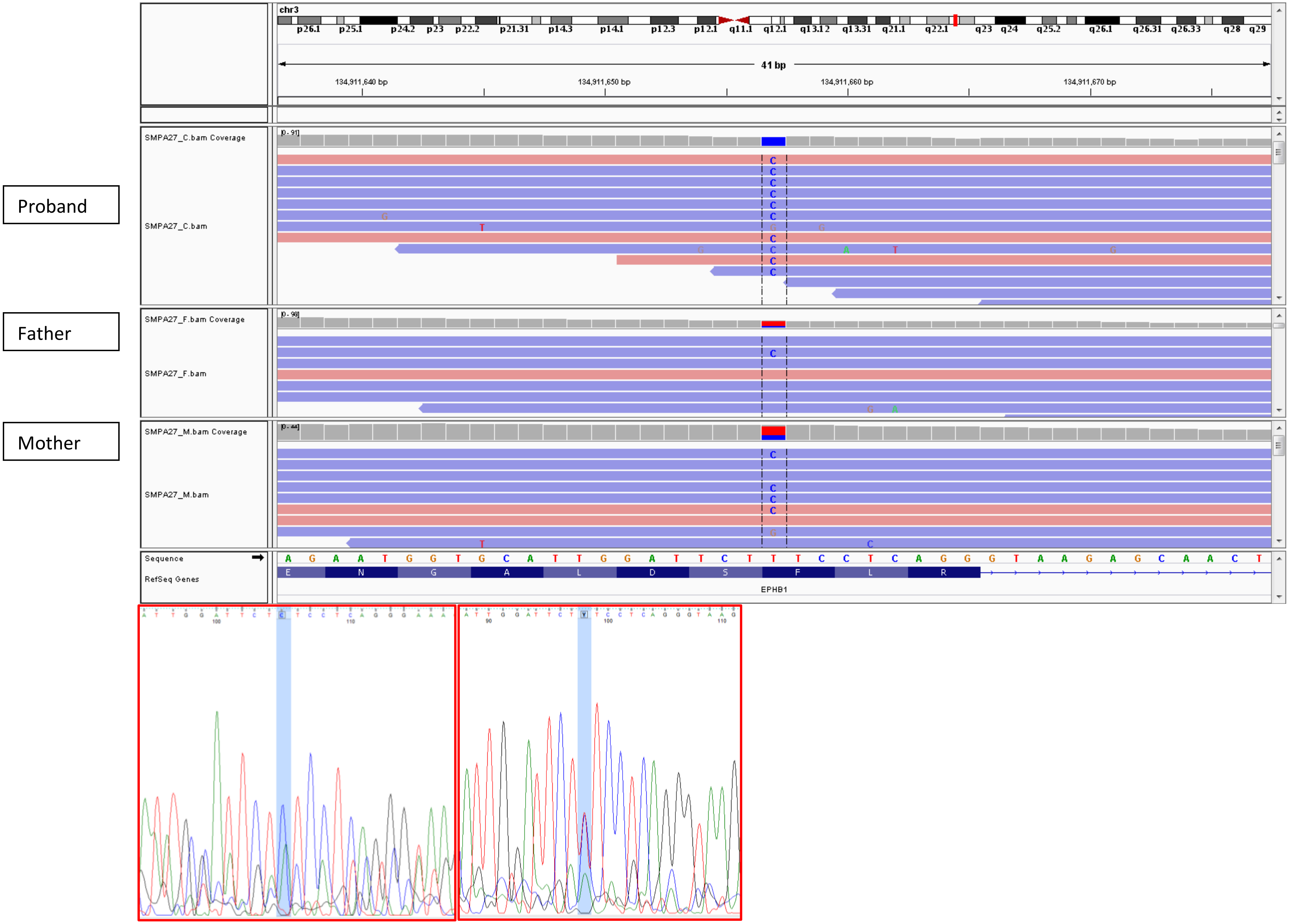


Proband Mother

12. Autism3: *RSRC1*

Proband

Father

Mother

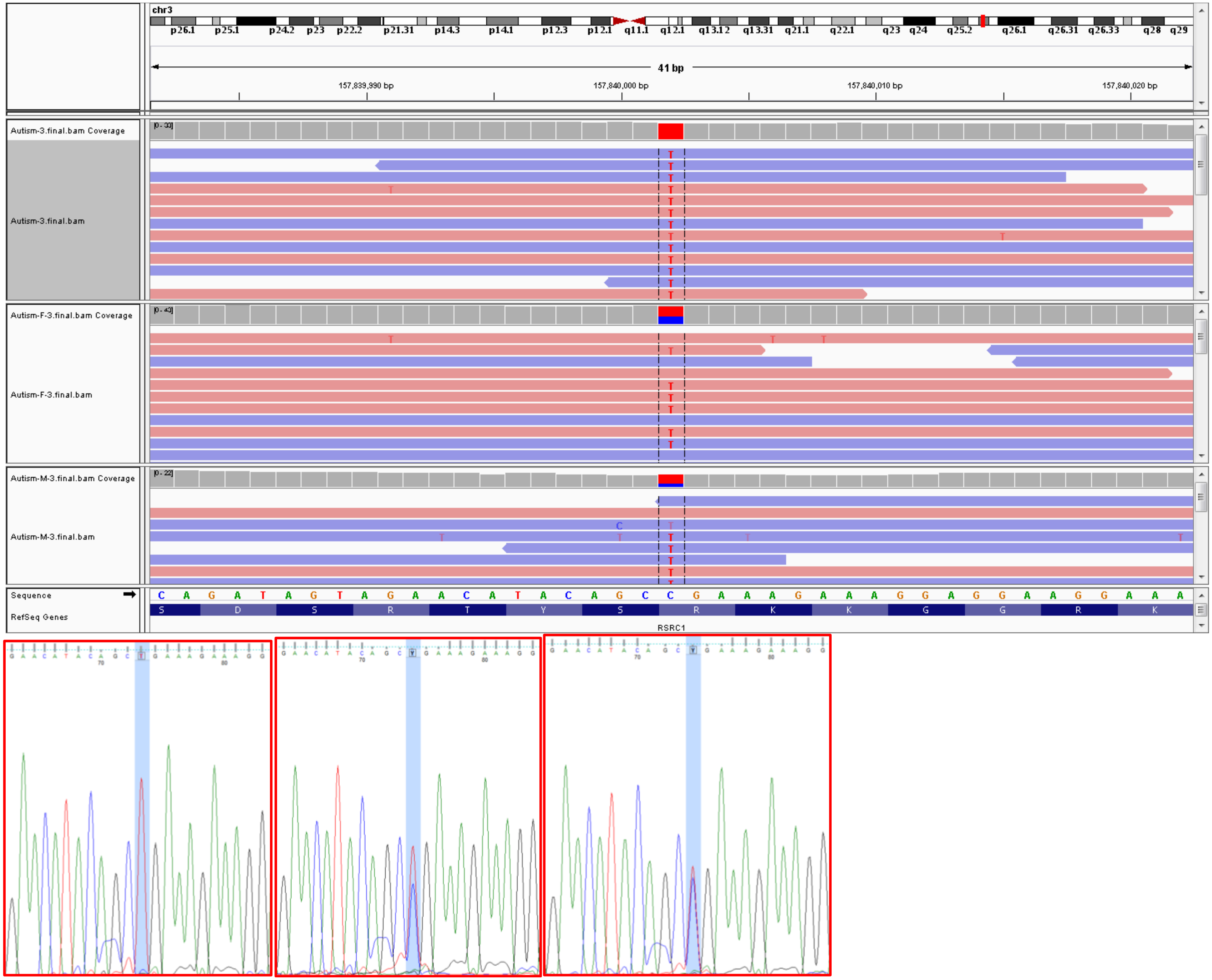


Proband

Father

Mother

13. SMPA8: *LRRC34*

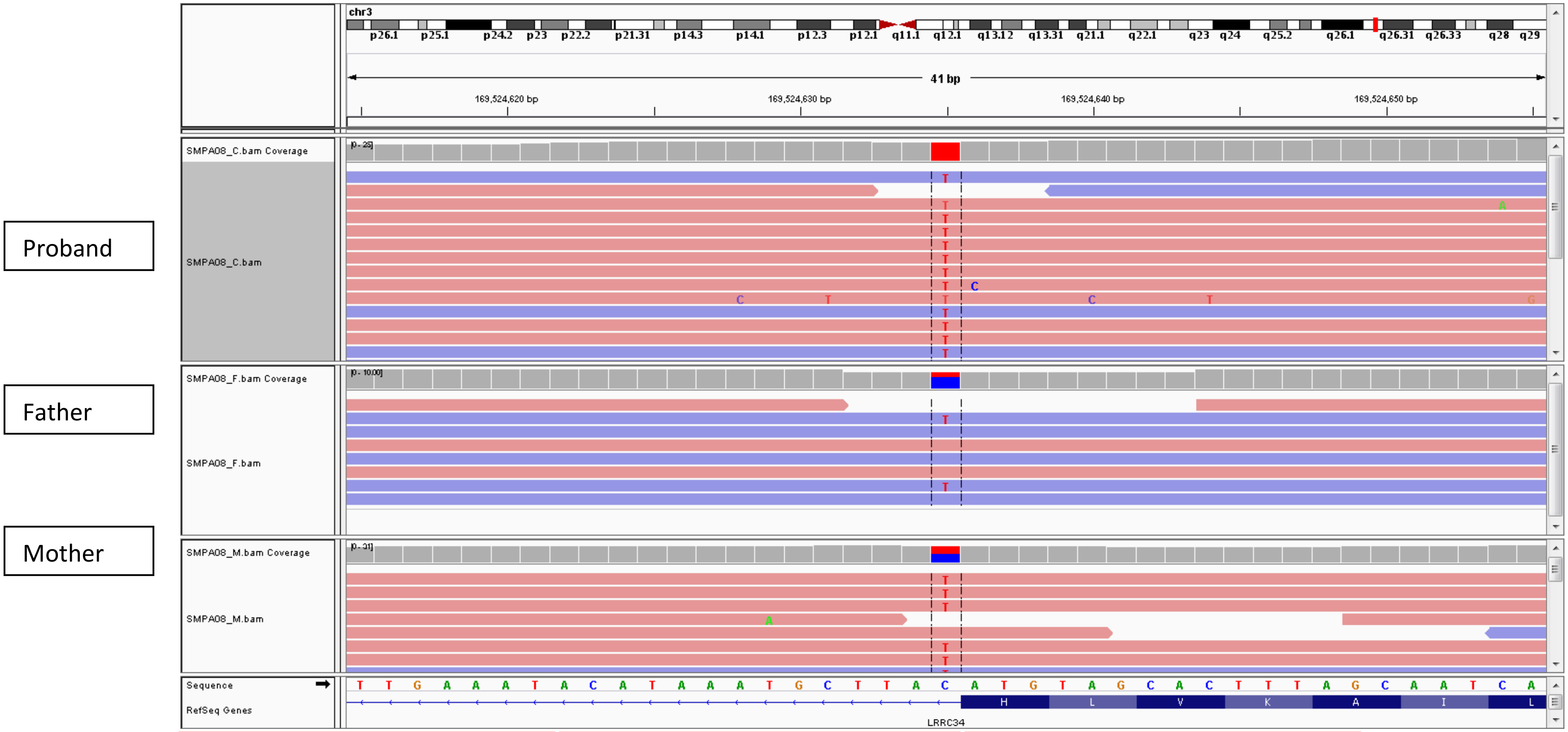


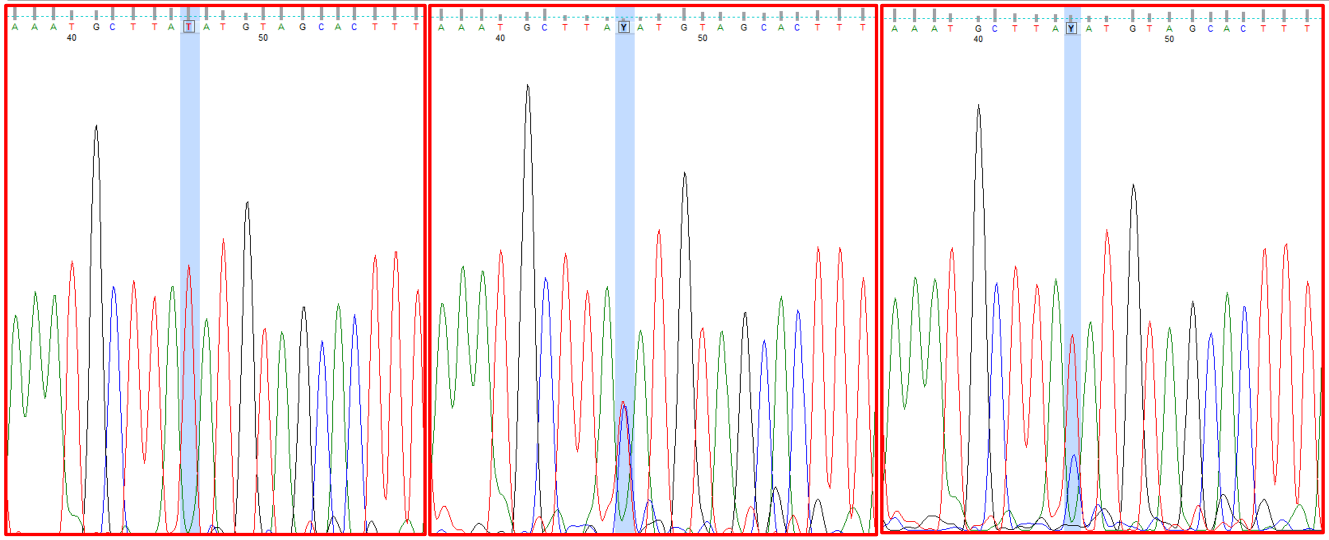

Proband Father Mother

14. SMPA40: *ENPEP*

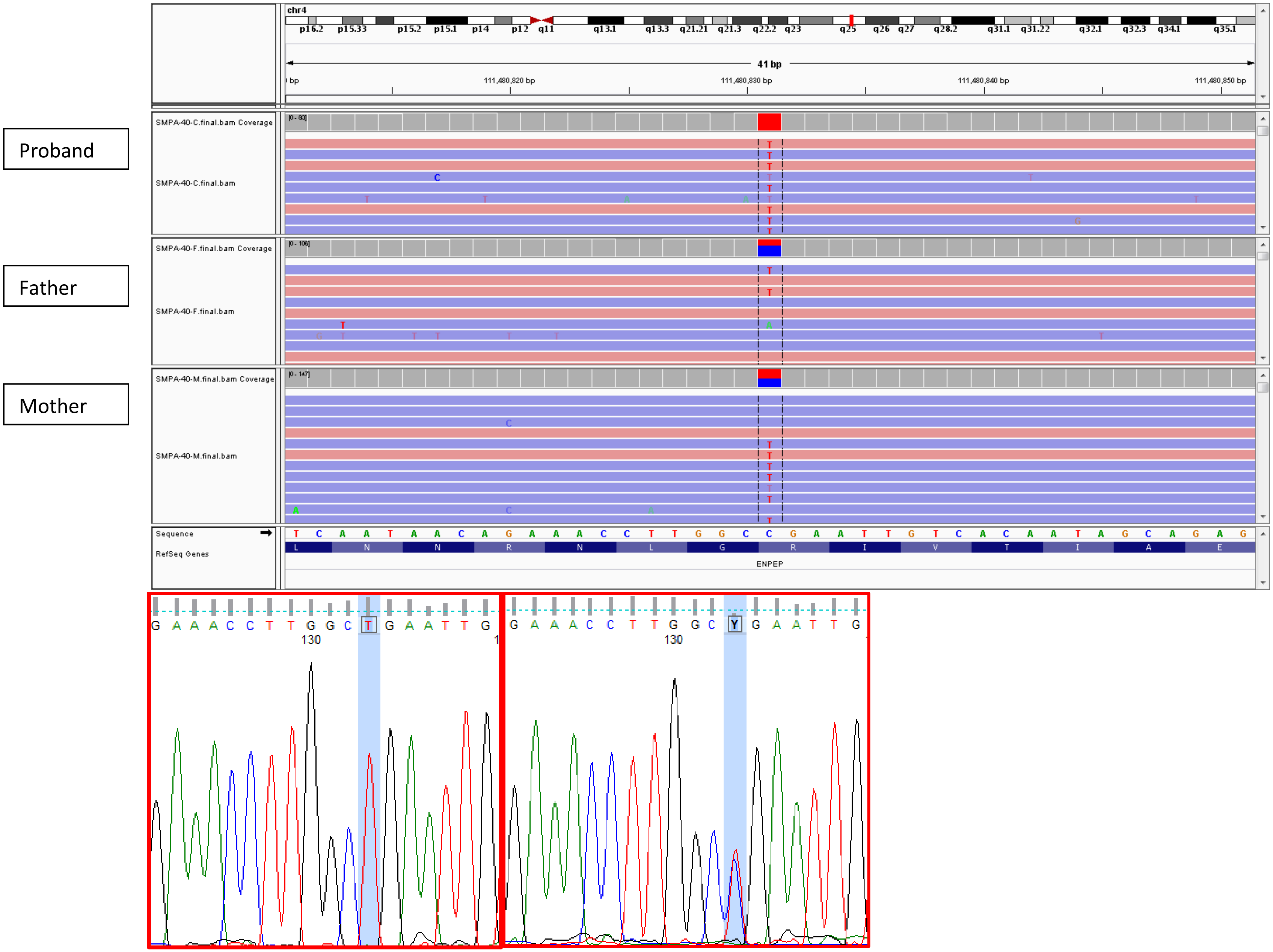


Proband Father

15. RQPA12: *AGA*

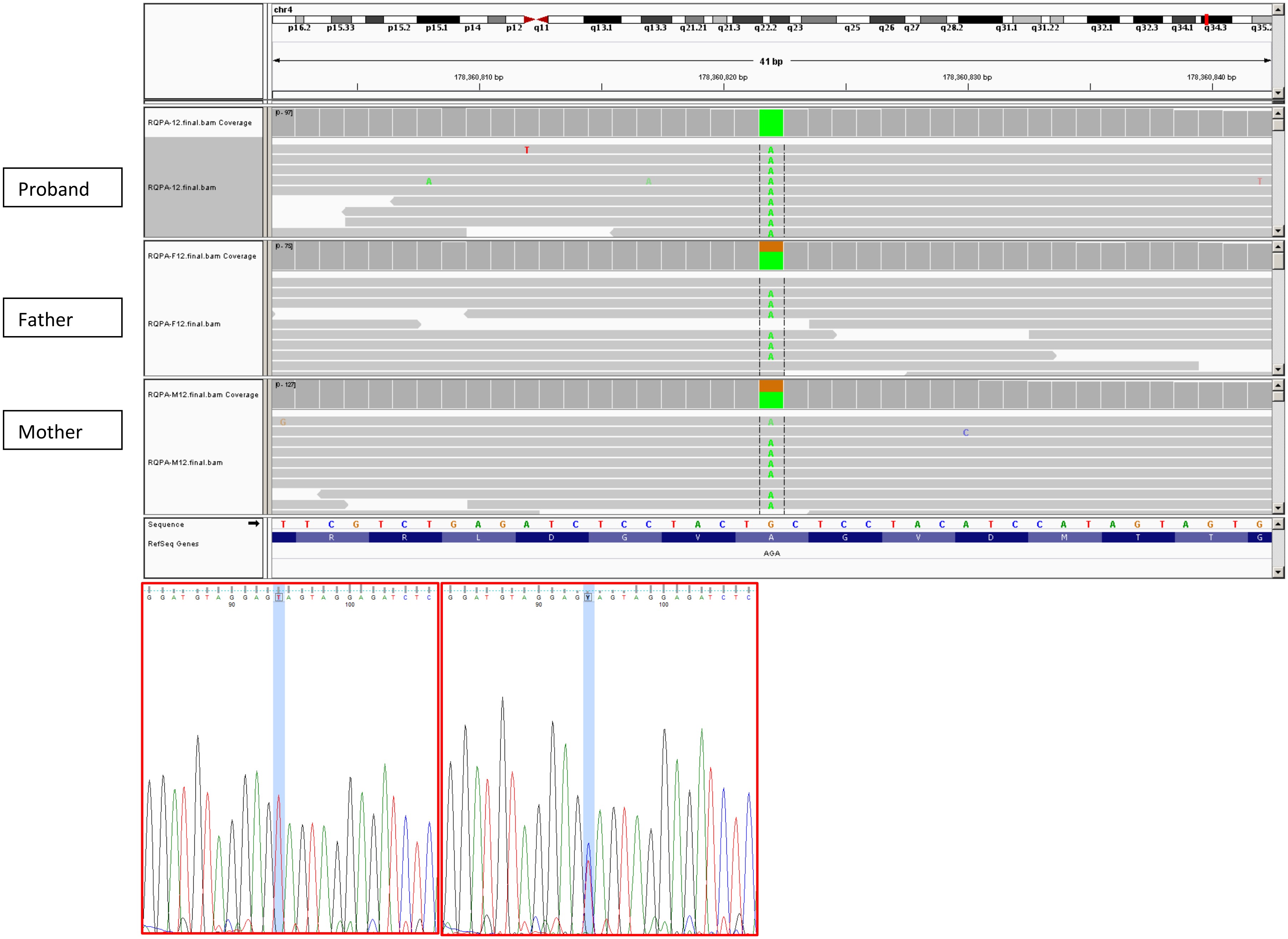


Proband

Mother

16. SMPA35: *ENPP6*

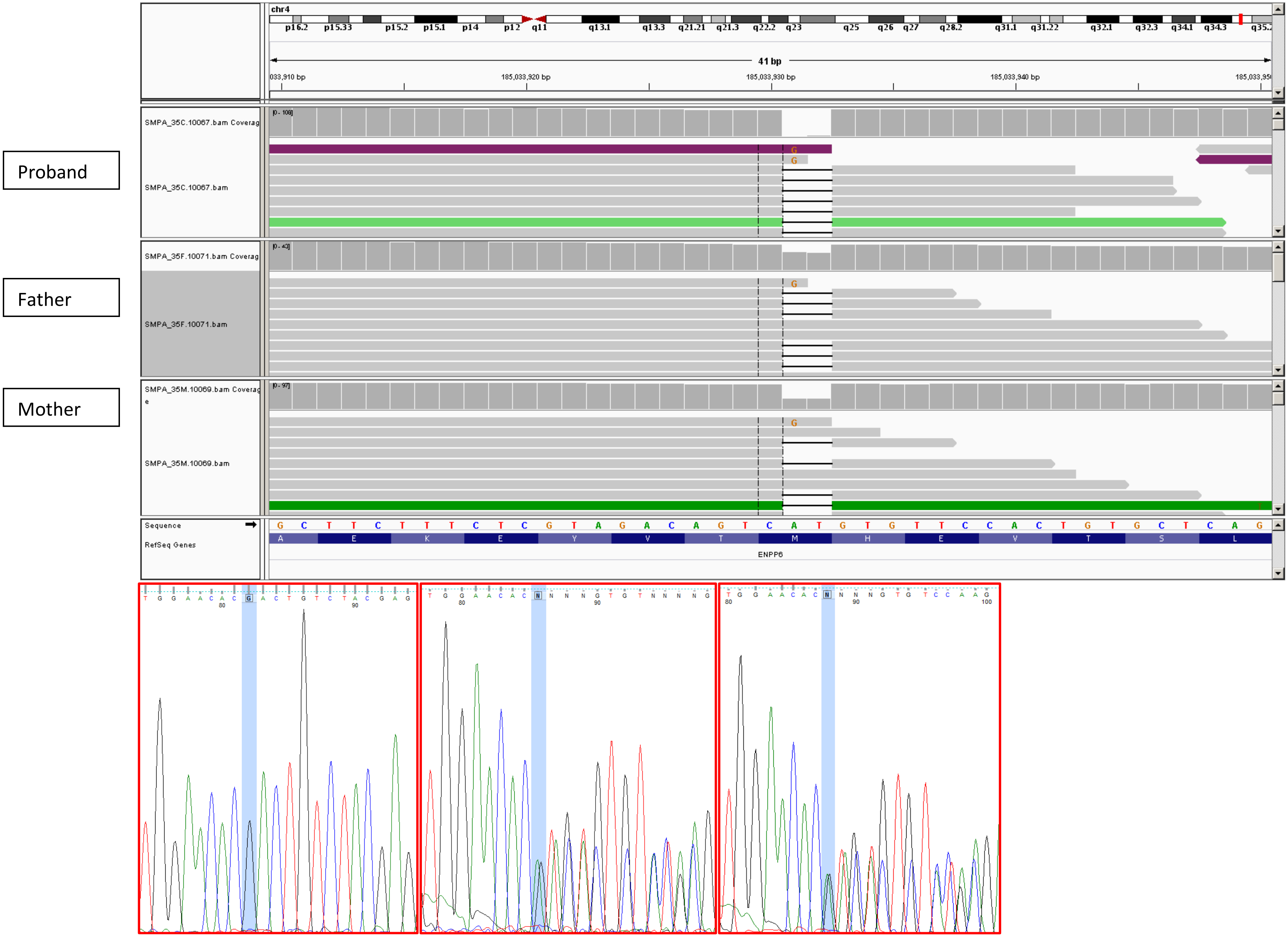


Proband Father Mother

17. IABB1: *CAPSL*

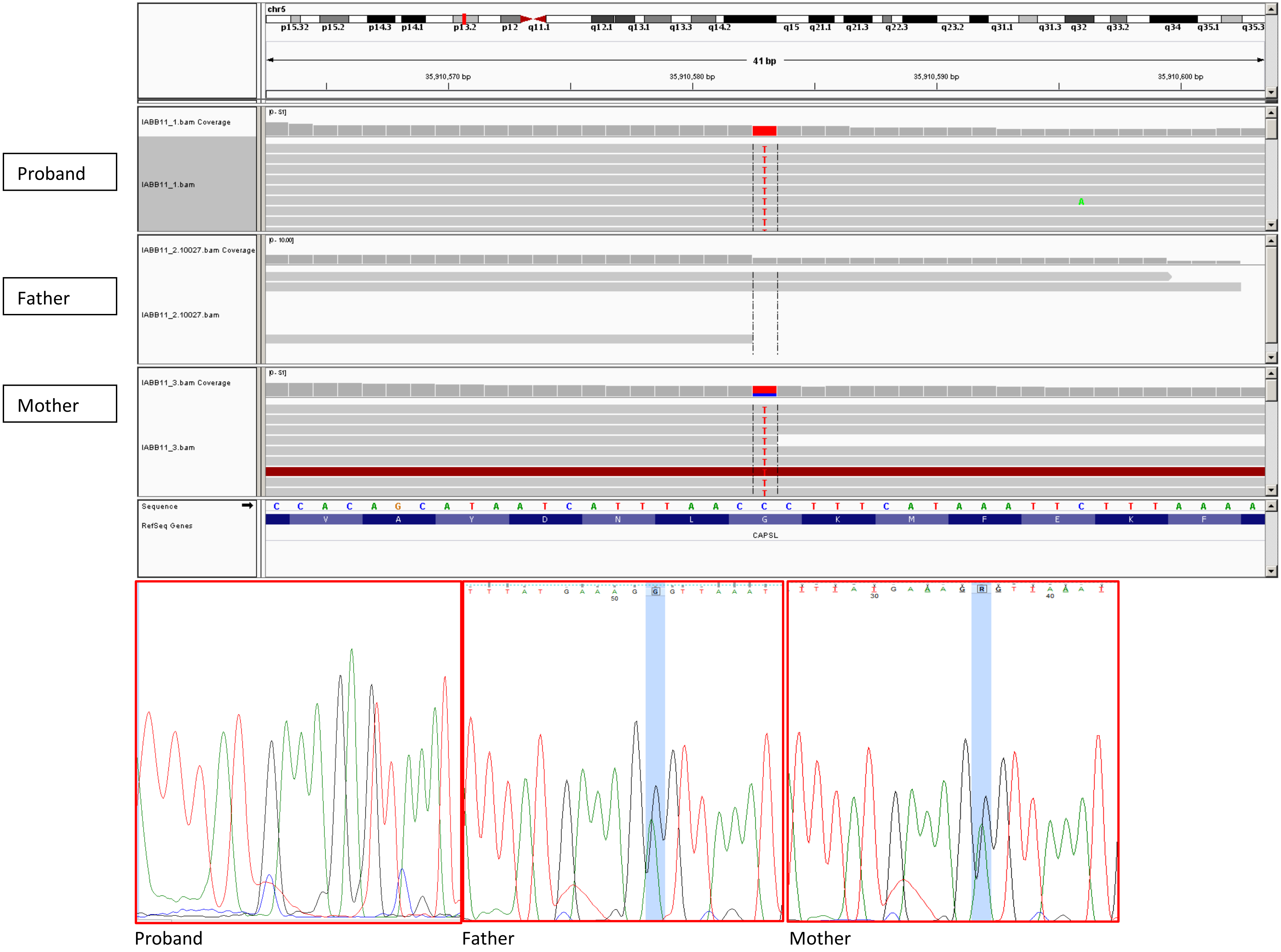

18. Autism8: *SLC36A1*

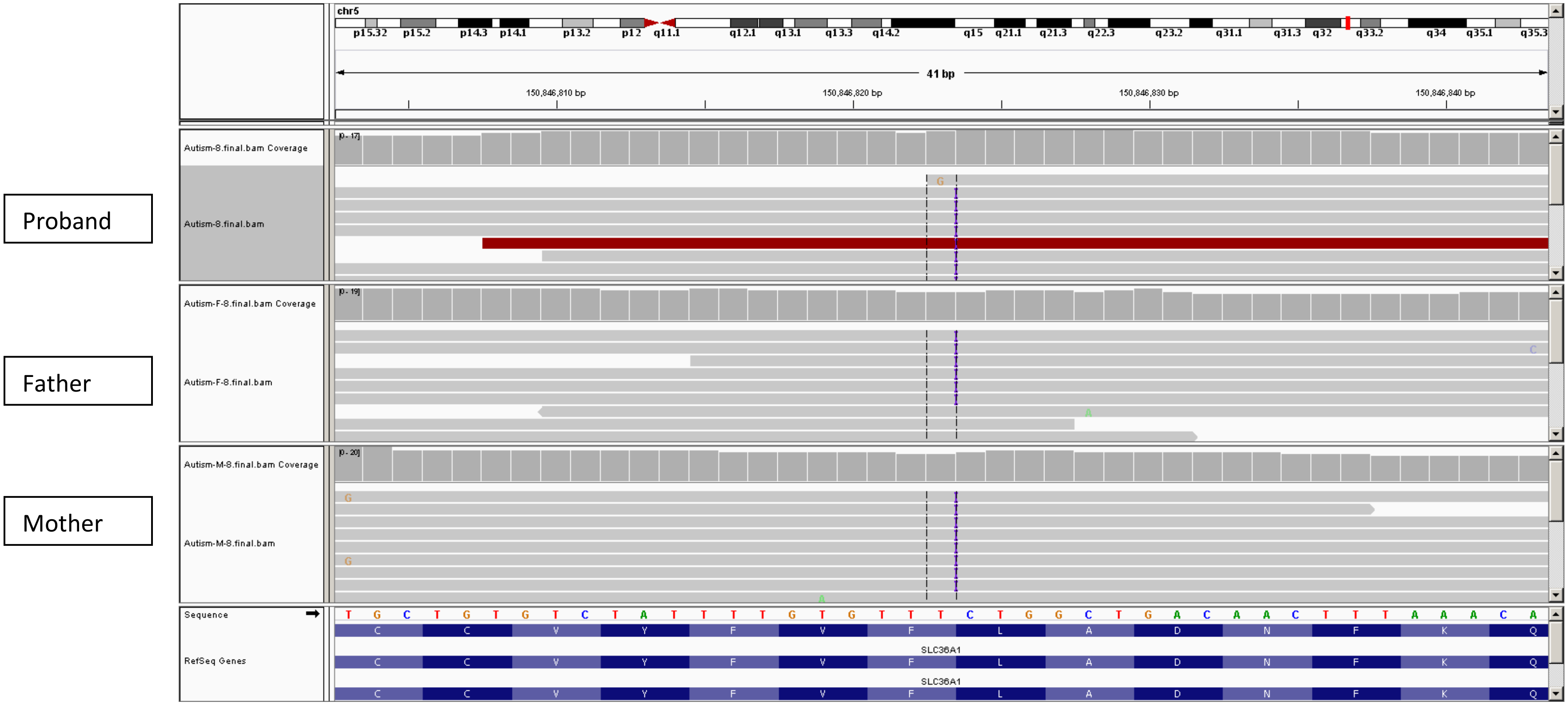


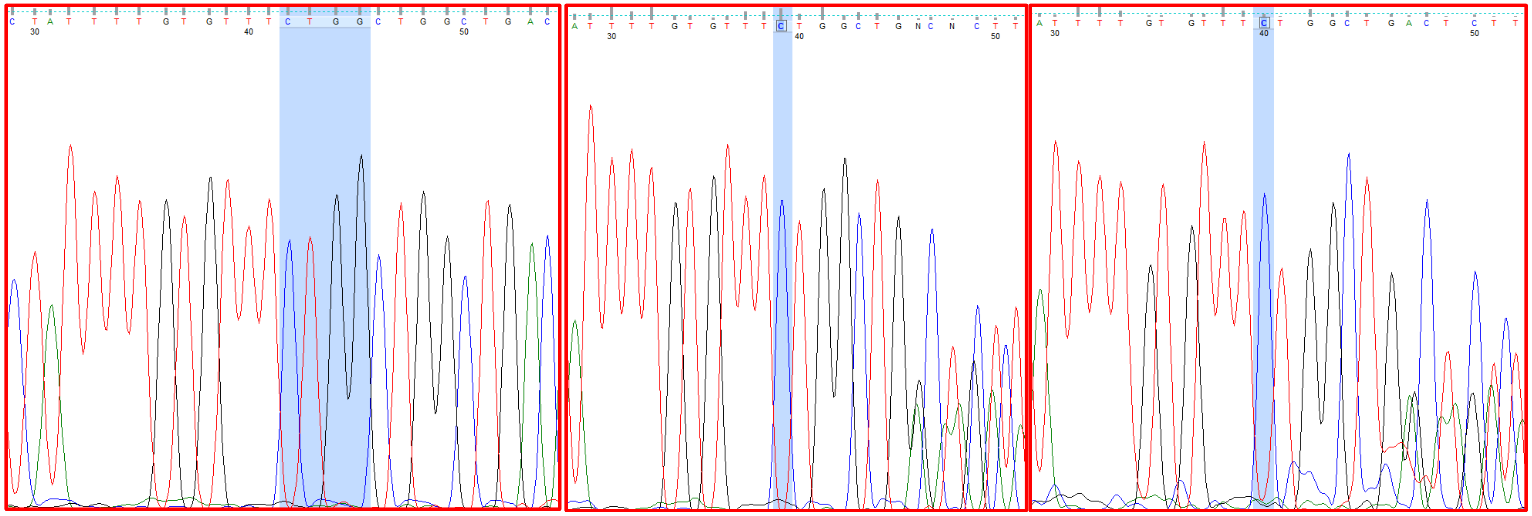

Proband Father Mother

19. IAU56: *FAXDC2*

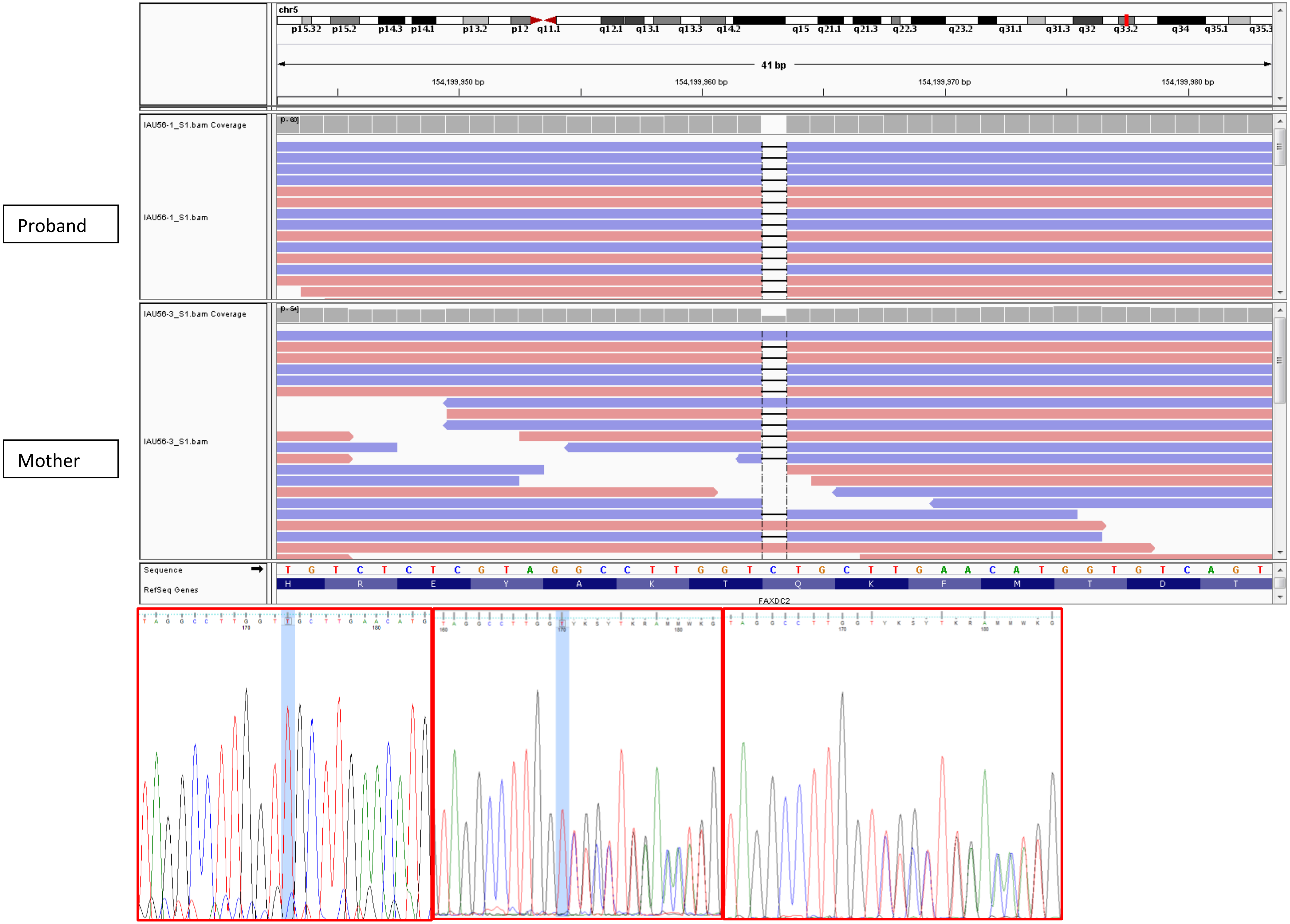


Proband Father Mother

### 20. SMPA48: *RANBP9*

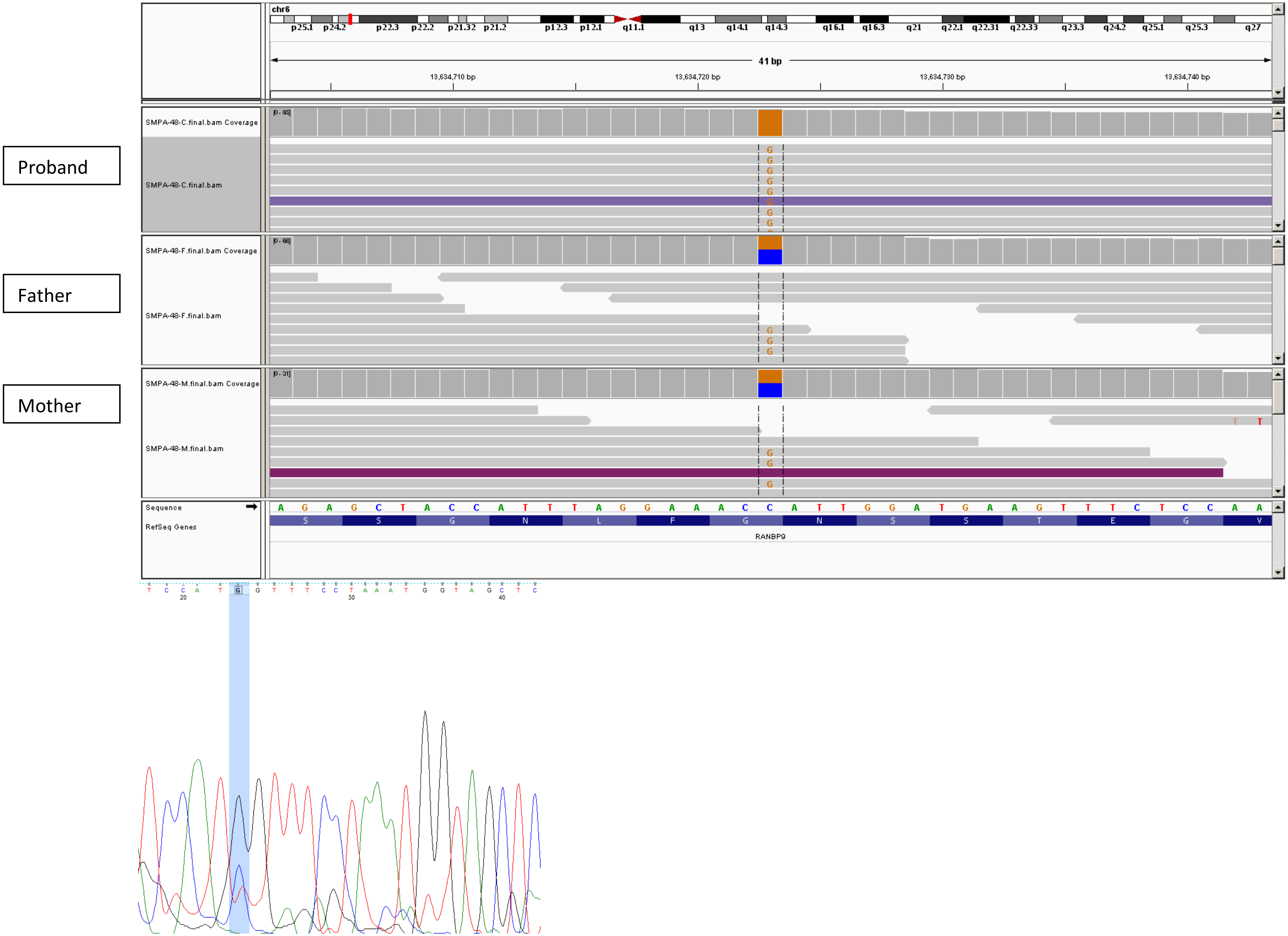


Mother (Father and proband DNA exhausted)

21. IABB8: *DNAH8*

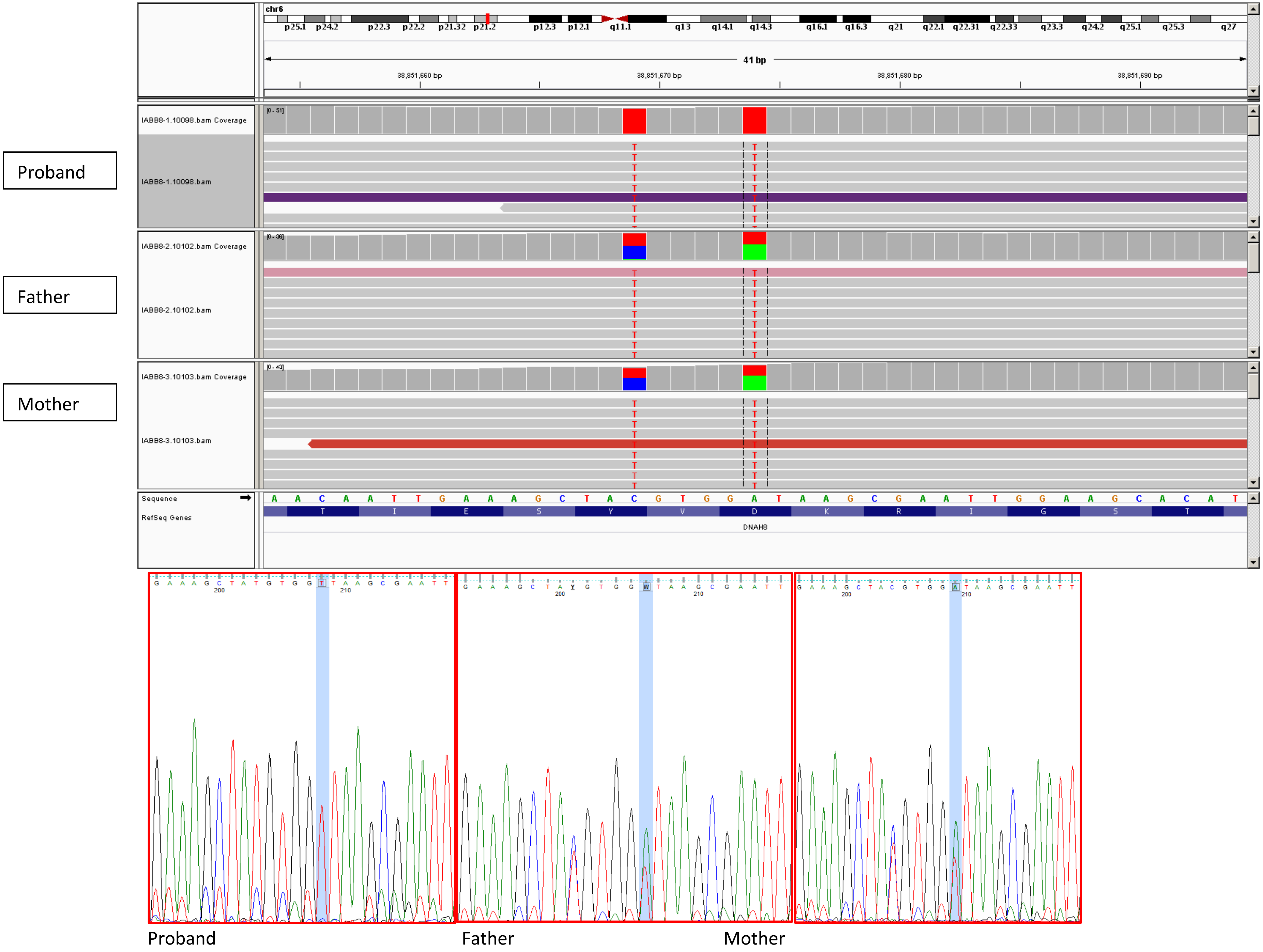

22. IAU7: *PKD1L1*

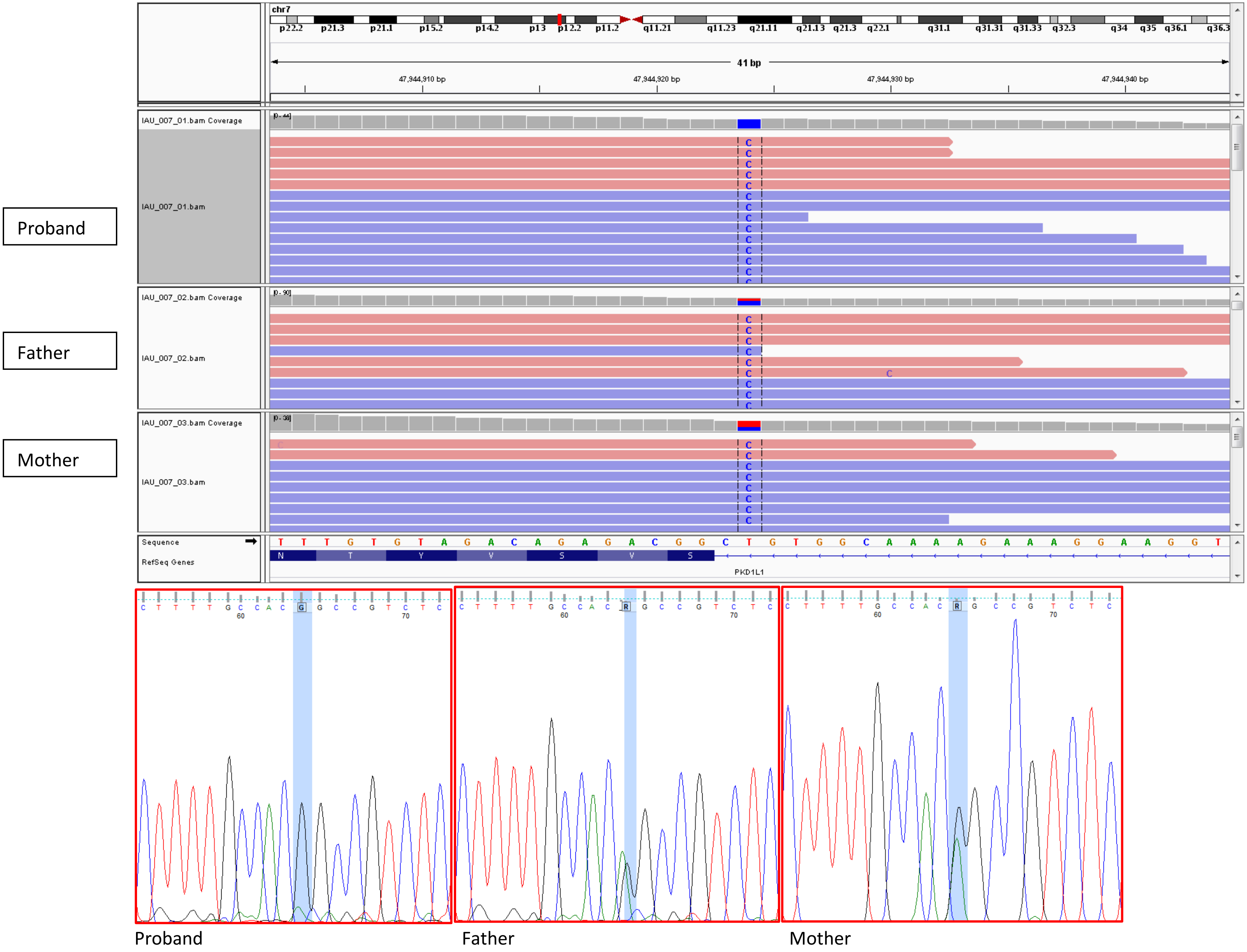

23. SA7: *ASL*

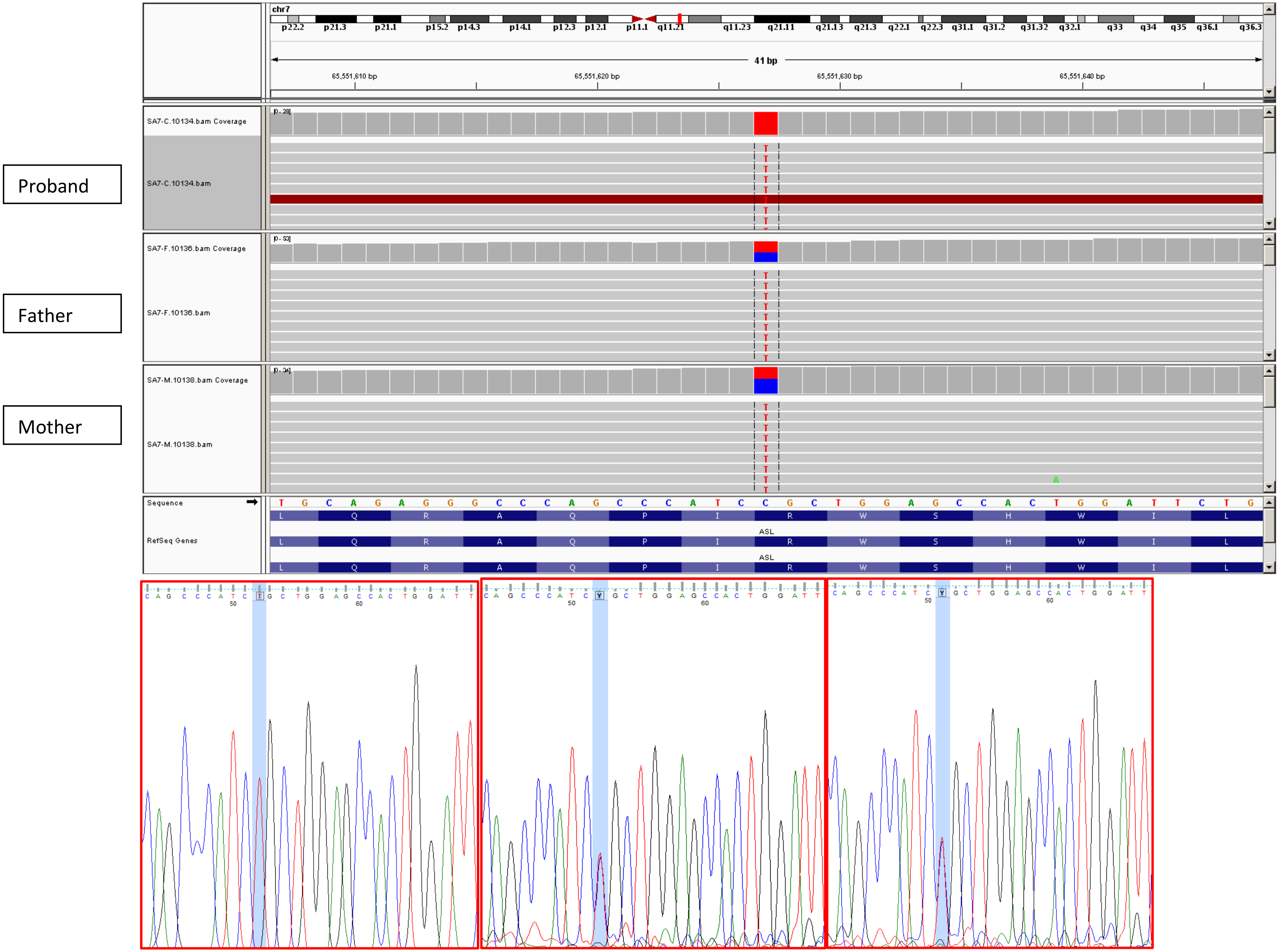


Proband Father Mother

24. IABB7: *CNPY4*

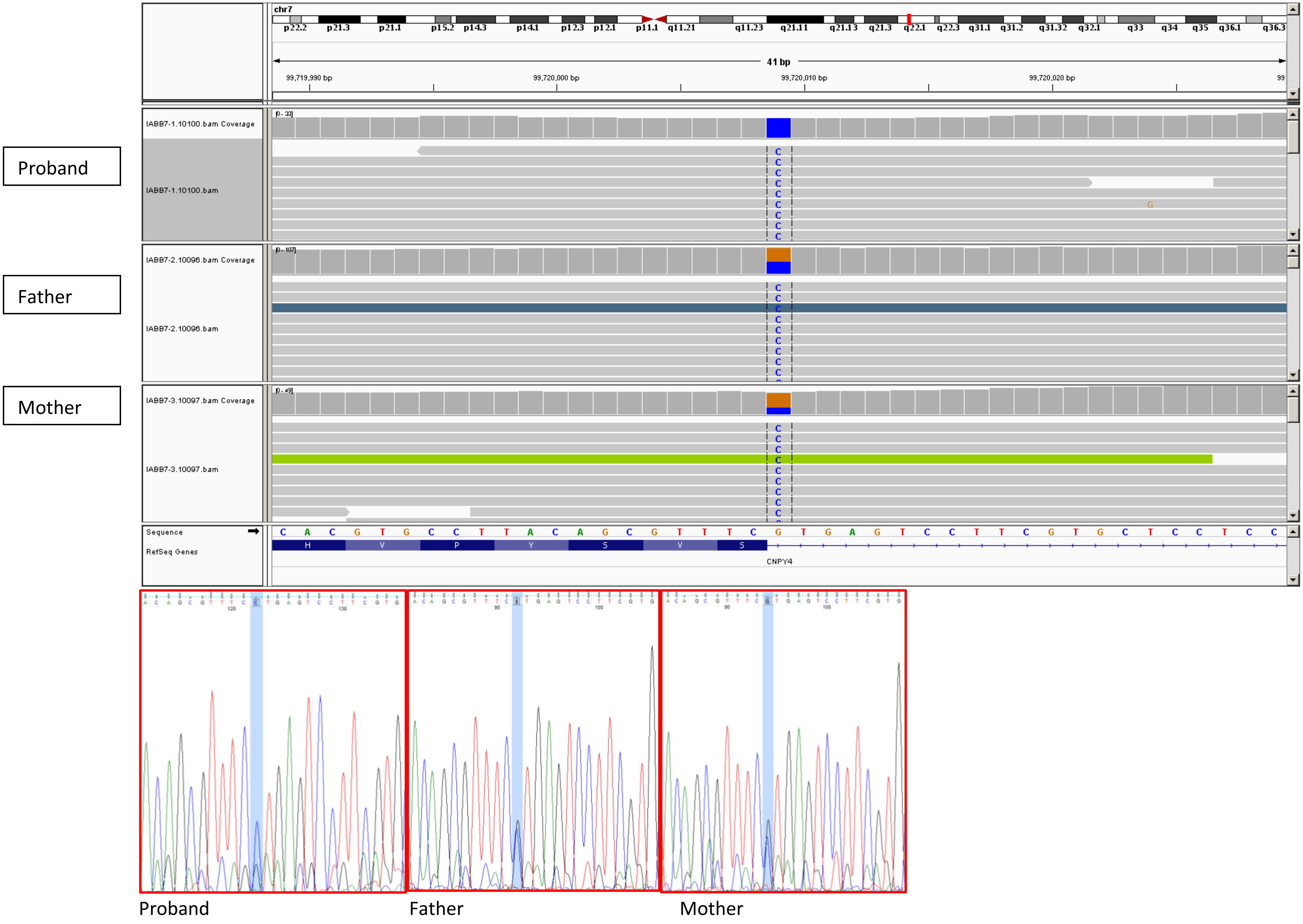

25. IAU79: *GIMAP8*

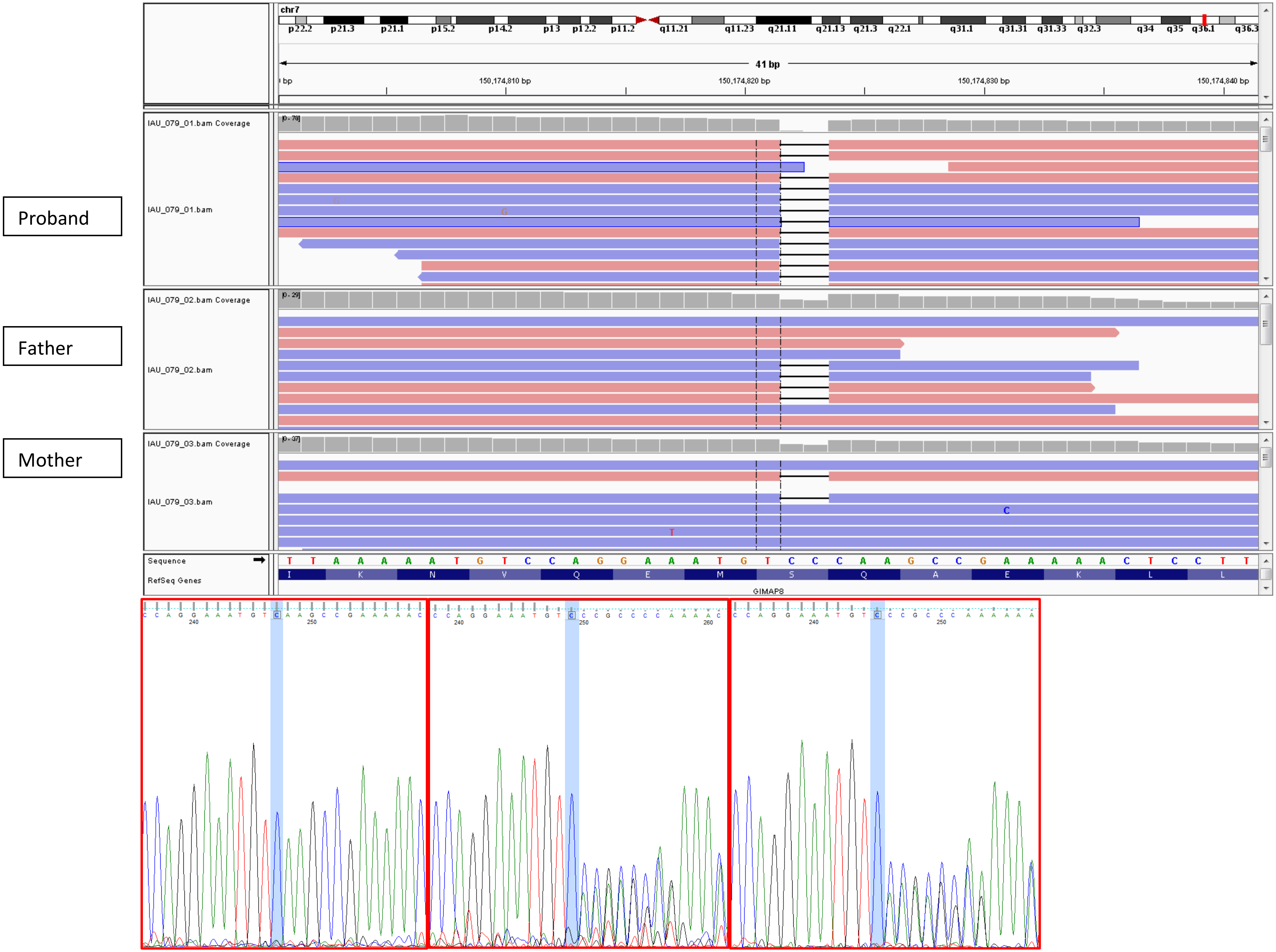


Proband Father Mother

26. SMPA11: *SHH*

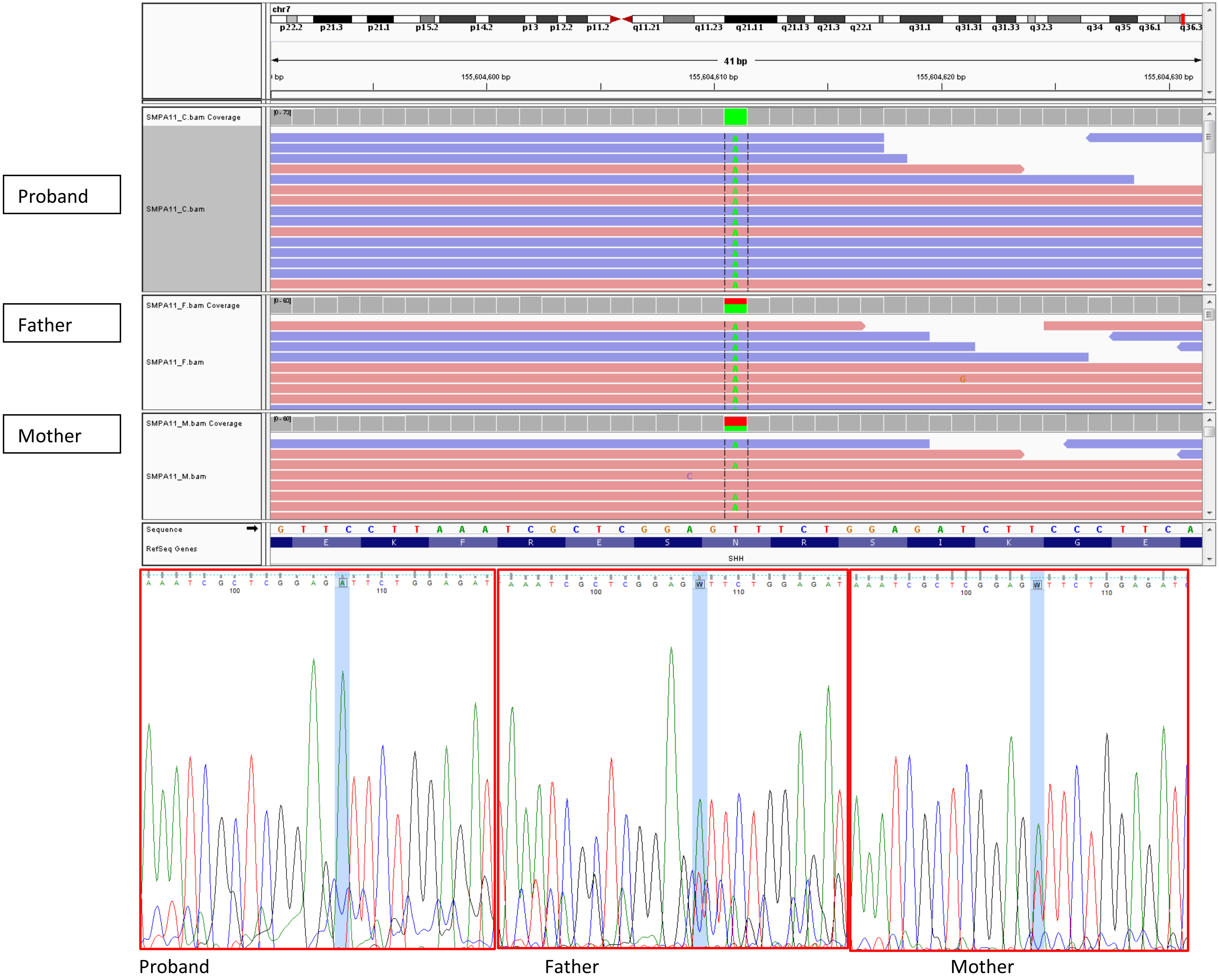

27. IABB3: *VPS13B*

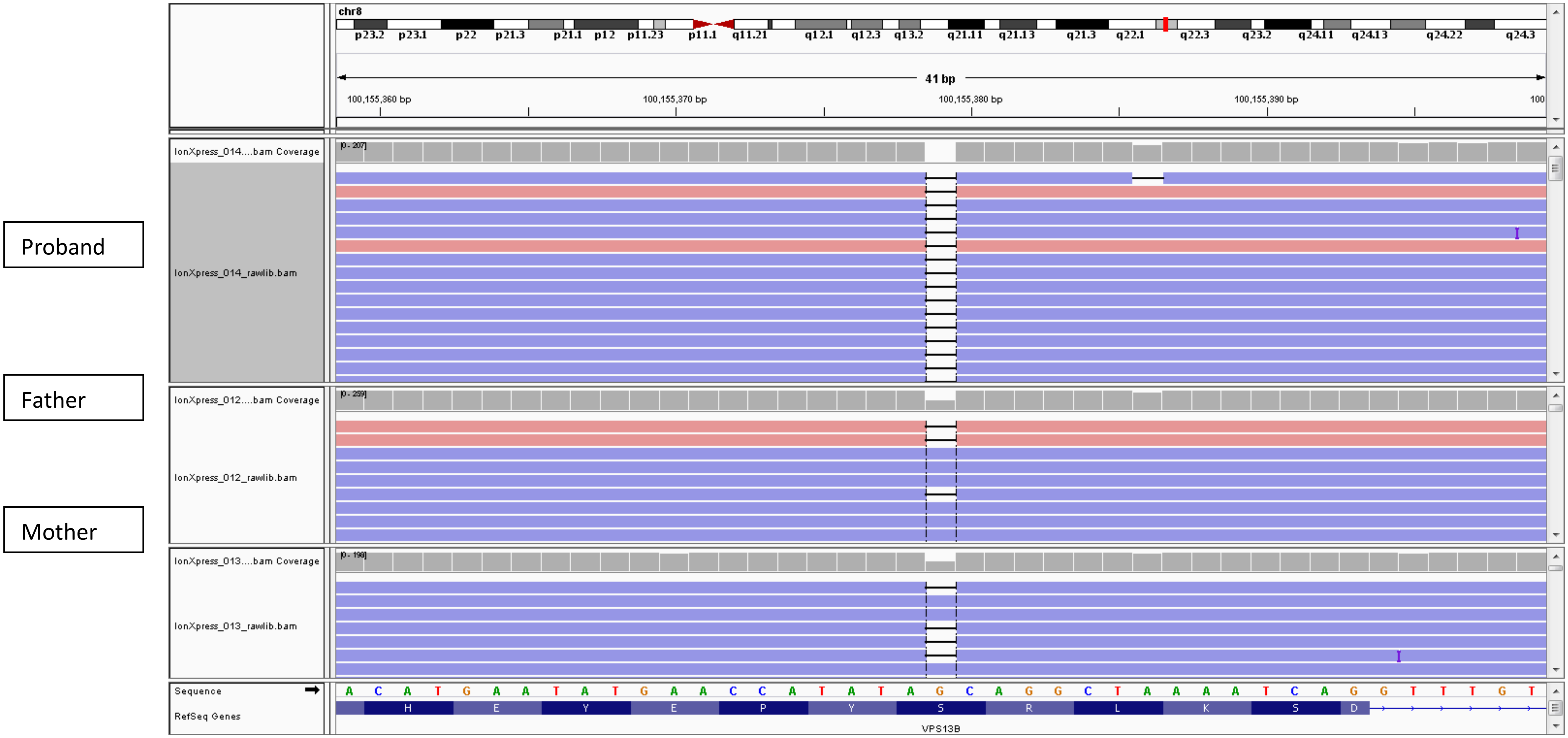


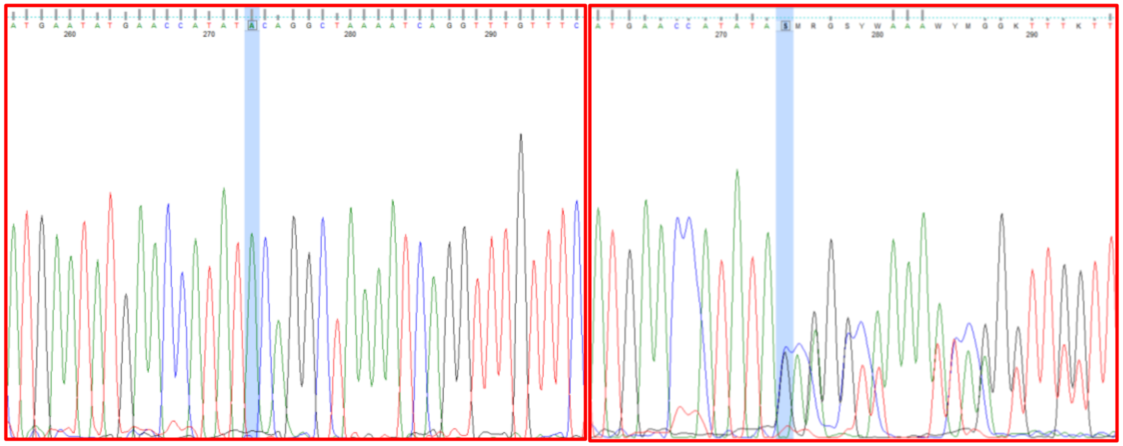

Proband Mother

28. SA4: *ERMP1*

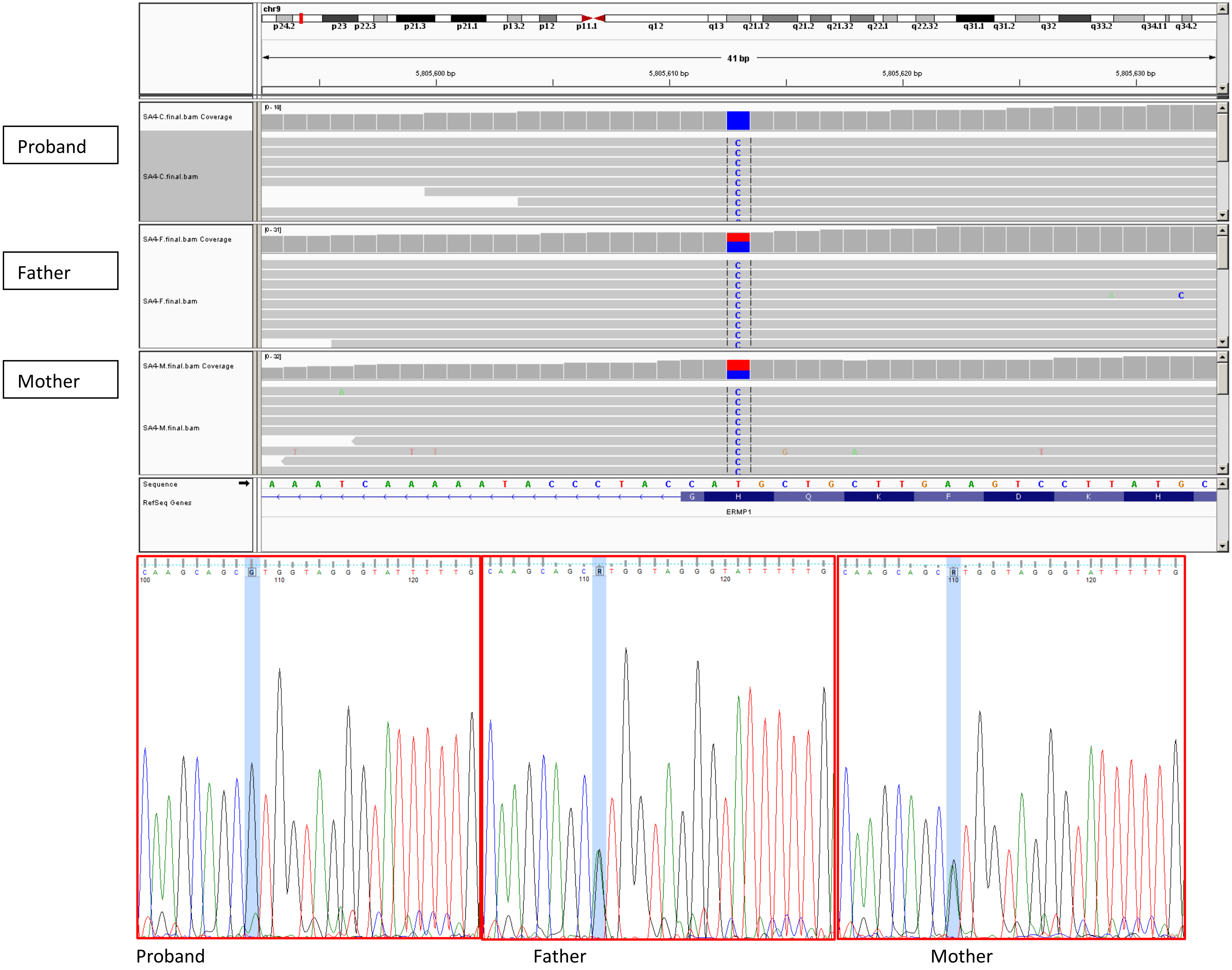
Proband
Father
Mother
chr9
41 bp
5,805,600 bp
5,805,610 bp
5,805,620 bp
5,805,630 bp
SA4-C.final.bam Coverage
SA4-C.final.bam
SA4-F.final.bam Coverage
SA4-F.final.bam
SA4-M.final.bam Coverage
SA4-M.final.bam
Sequence
RefSeq Genes
ERMP1
Proband
Father
Mother

29. IABB2: *DEAF1*

Father

Mother

Probad

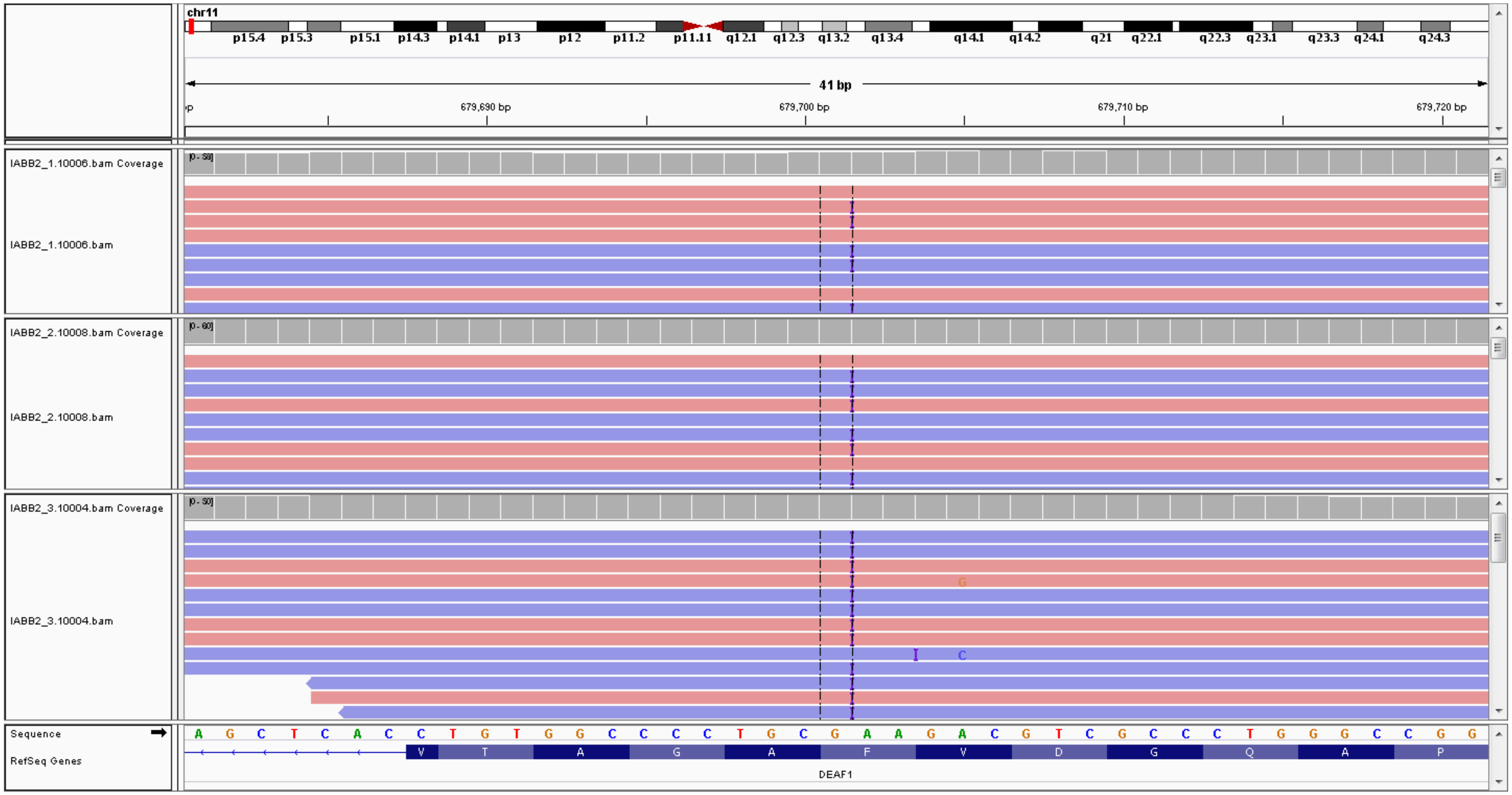


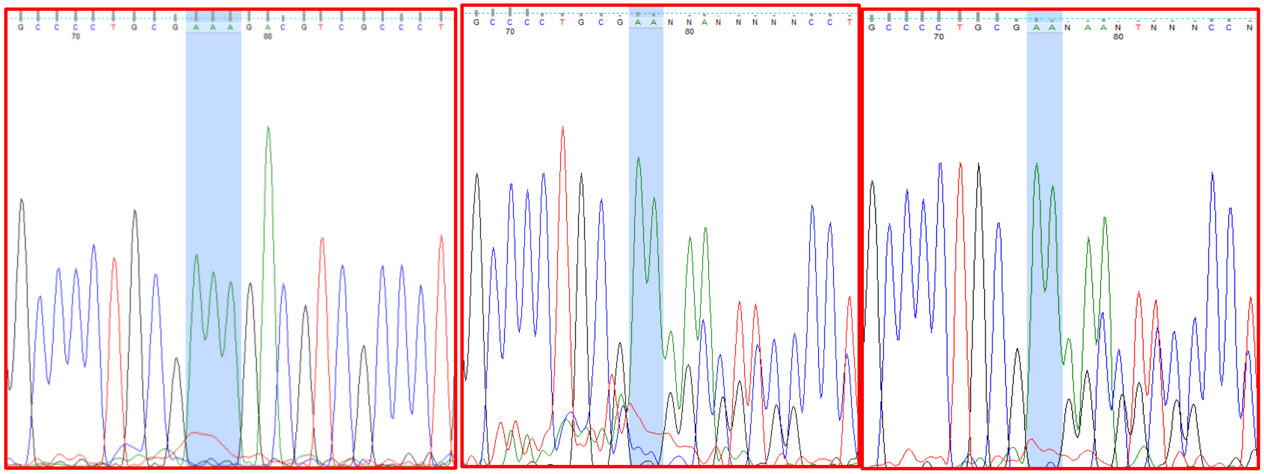

Proband

Father

Mother

30. SMPA11: *TRIM3*

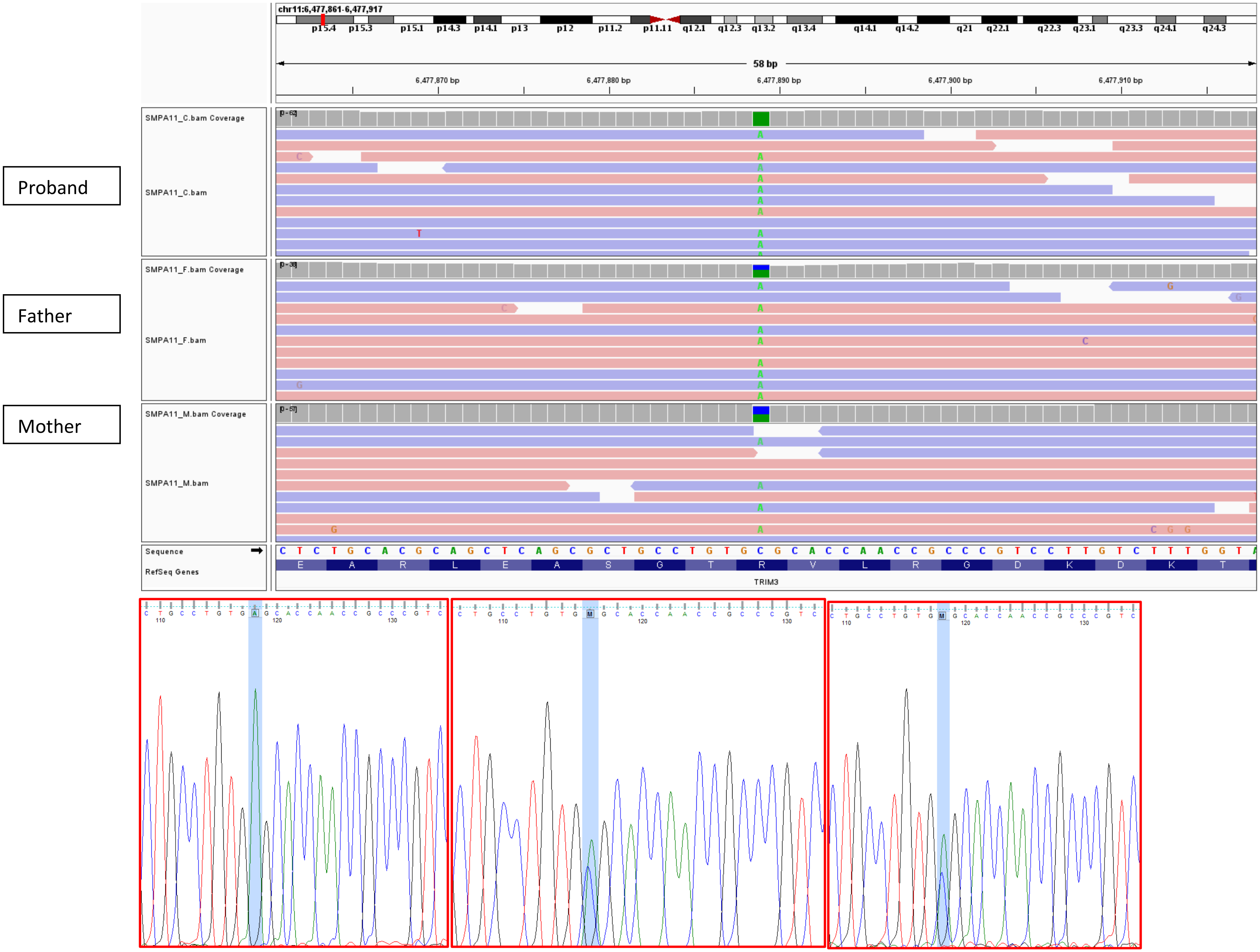


Proband Father Mother

31. SMPA20: *MADD*

Proband

Father

Mother

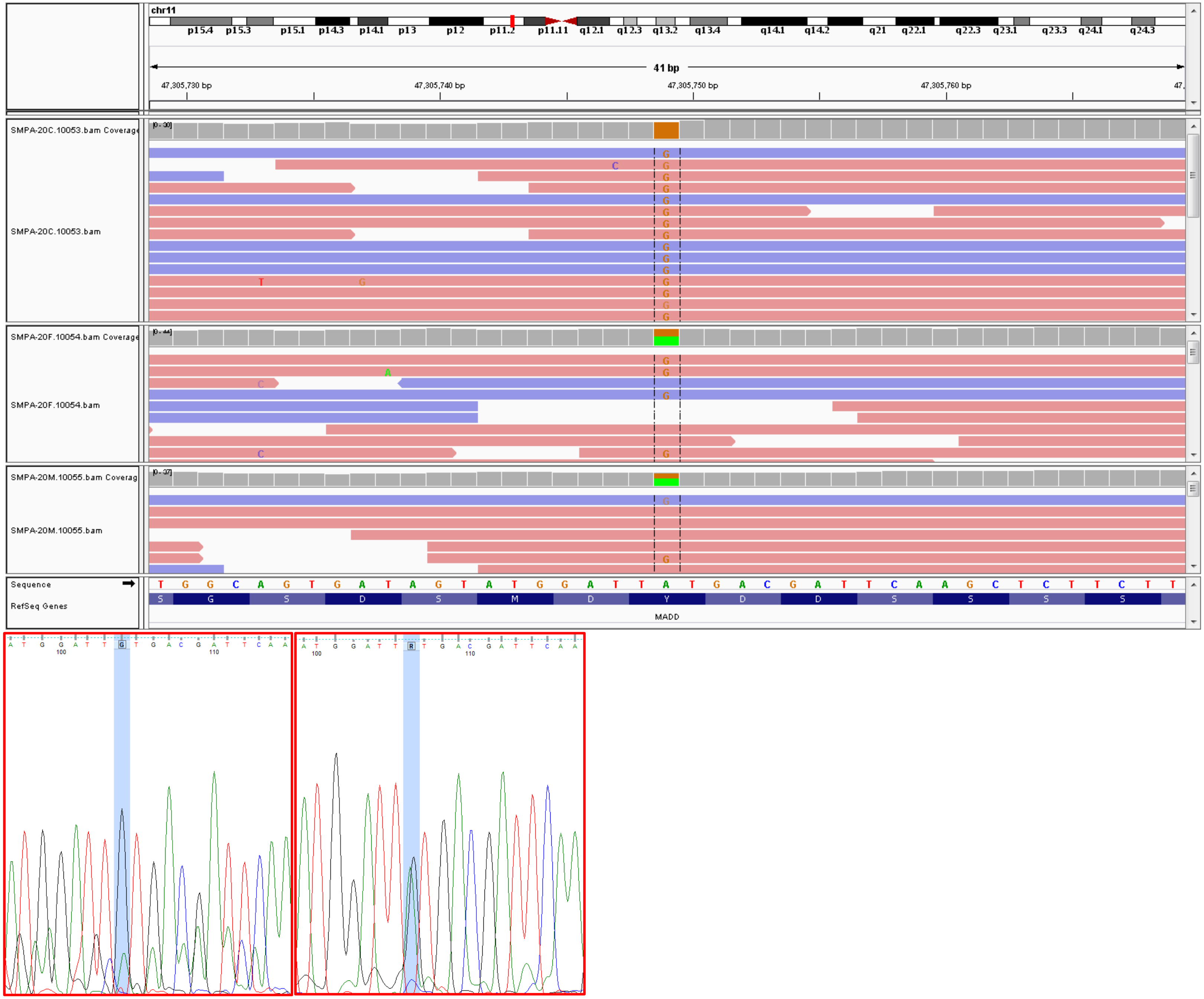


Proband

Father

32. SMPA21: *DAGLA*

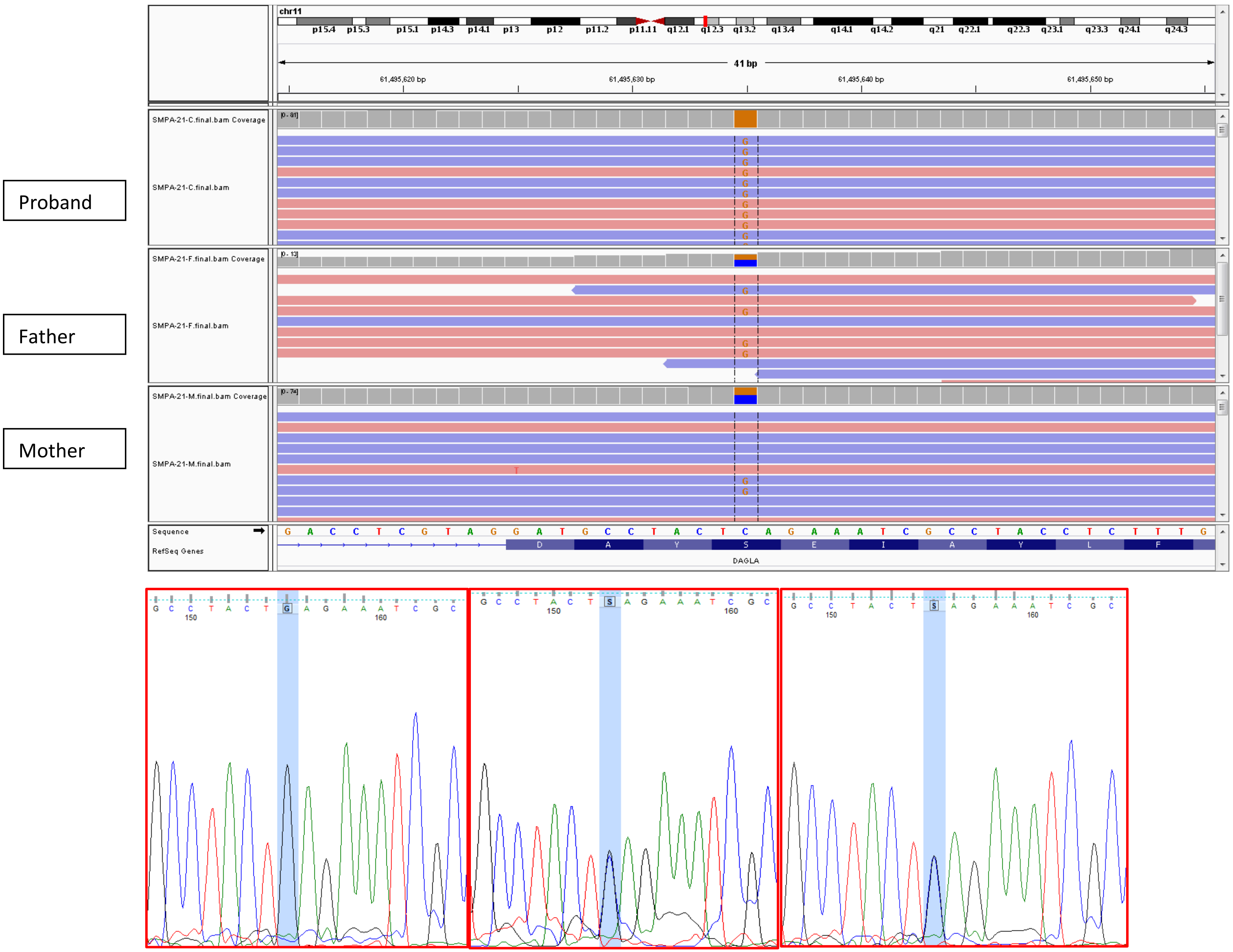


Proband Father Mother

33. SMPA80: *CABP2*

Proband

Father

Mother

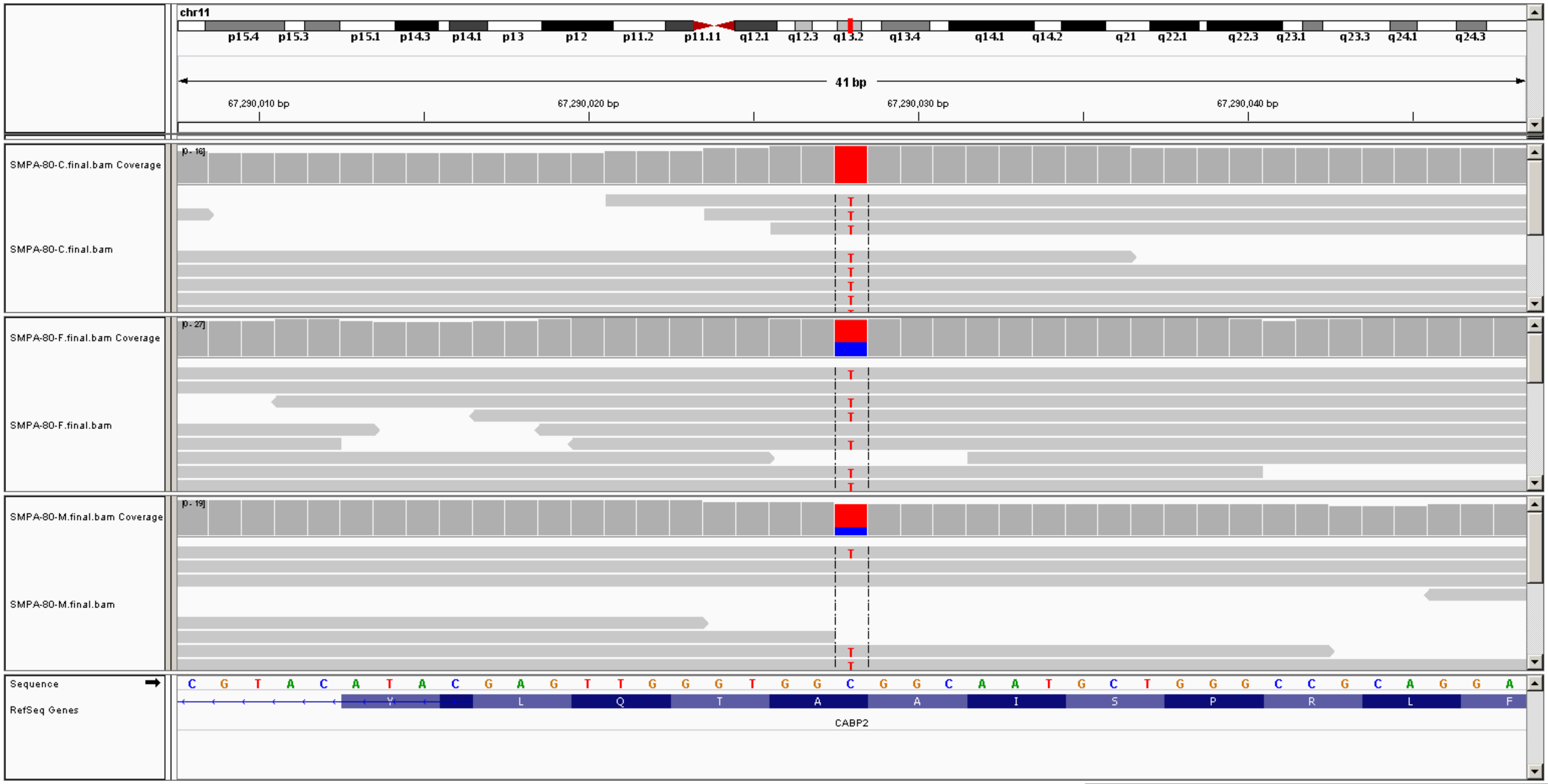


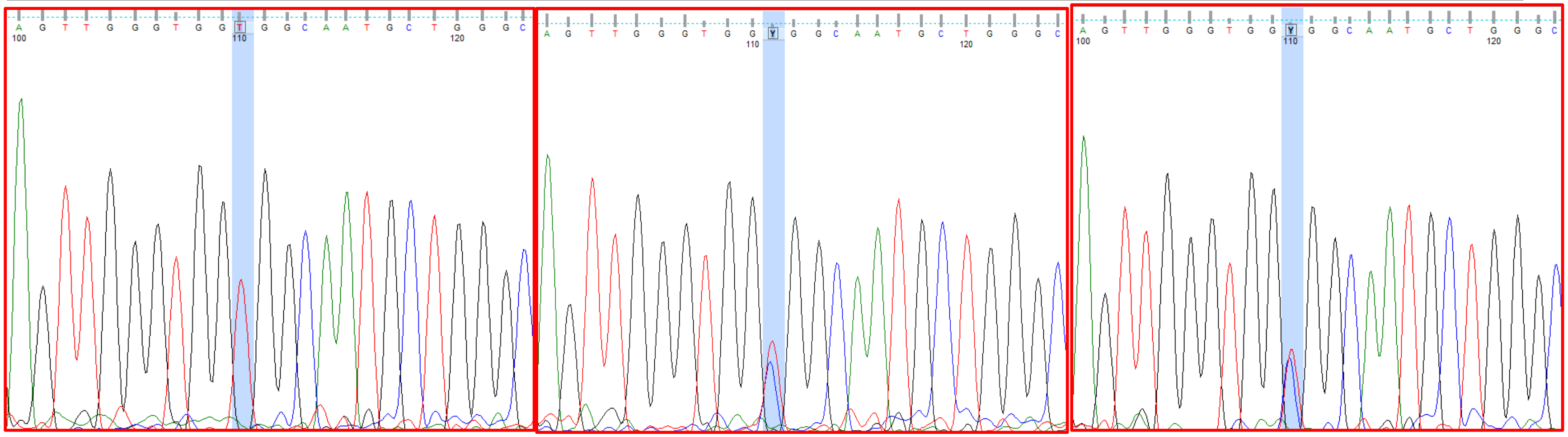

Proband Father Mother

34. SMPA8: *TMEM25*

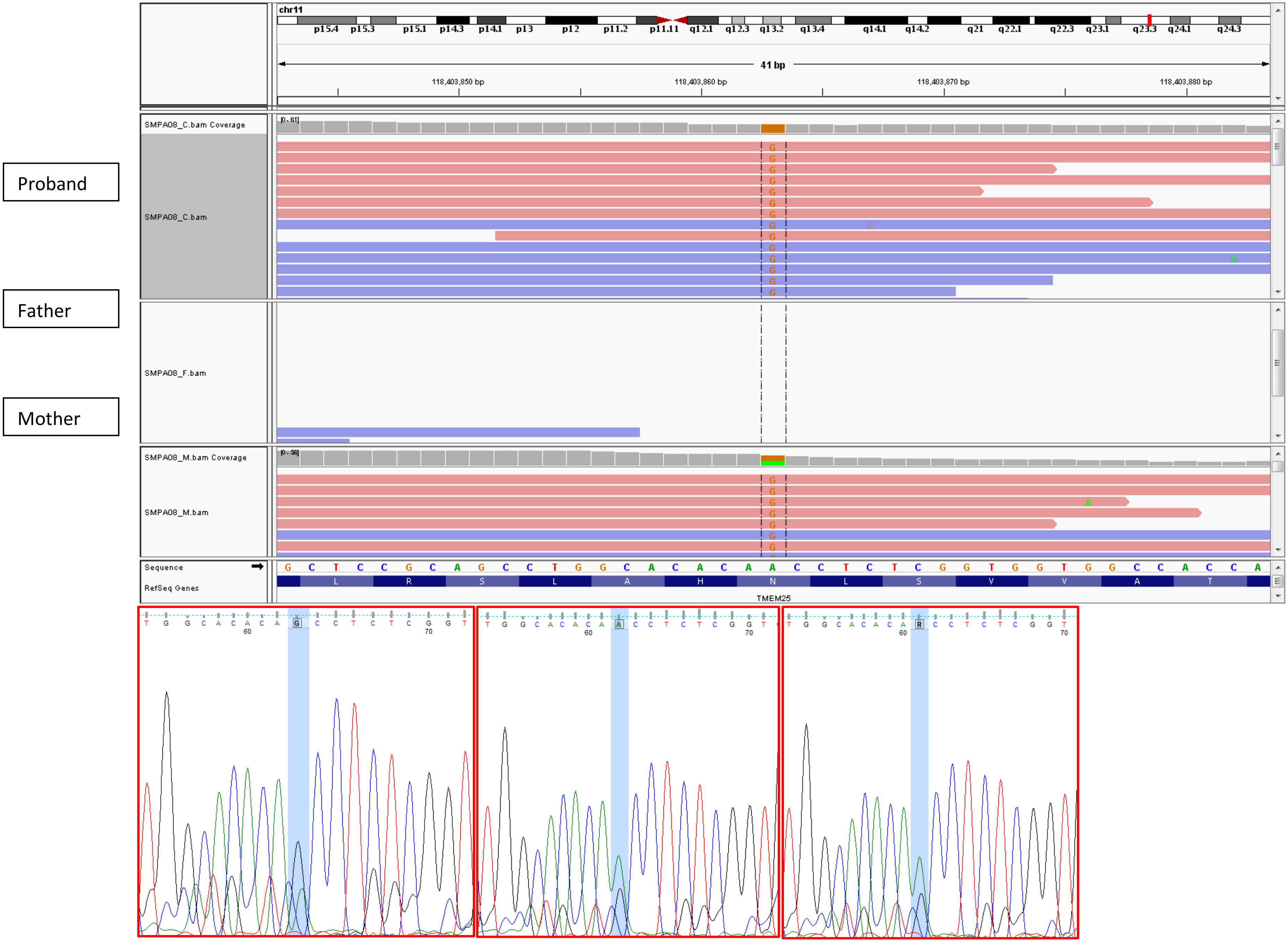


Proband Father Mother

35. IAU79: *B4GALNT1*

Proband

Father

Mother

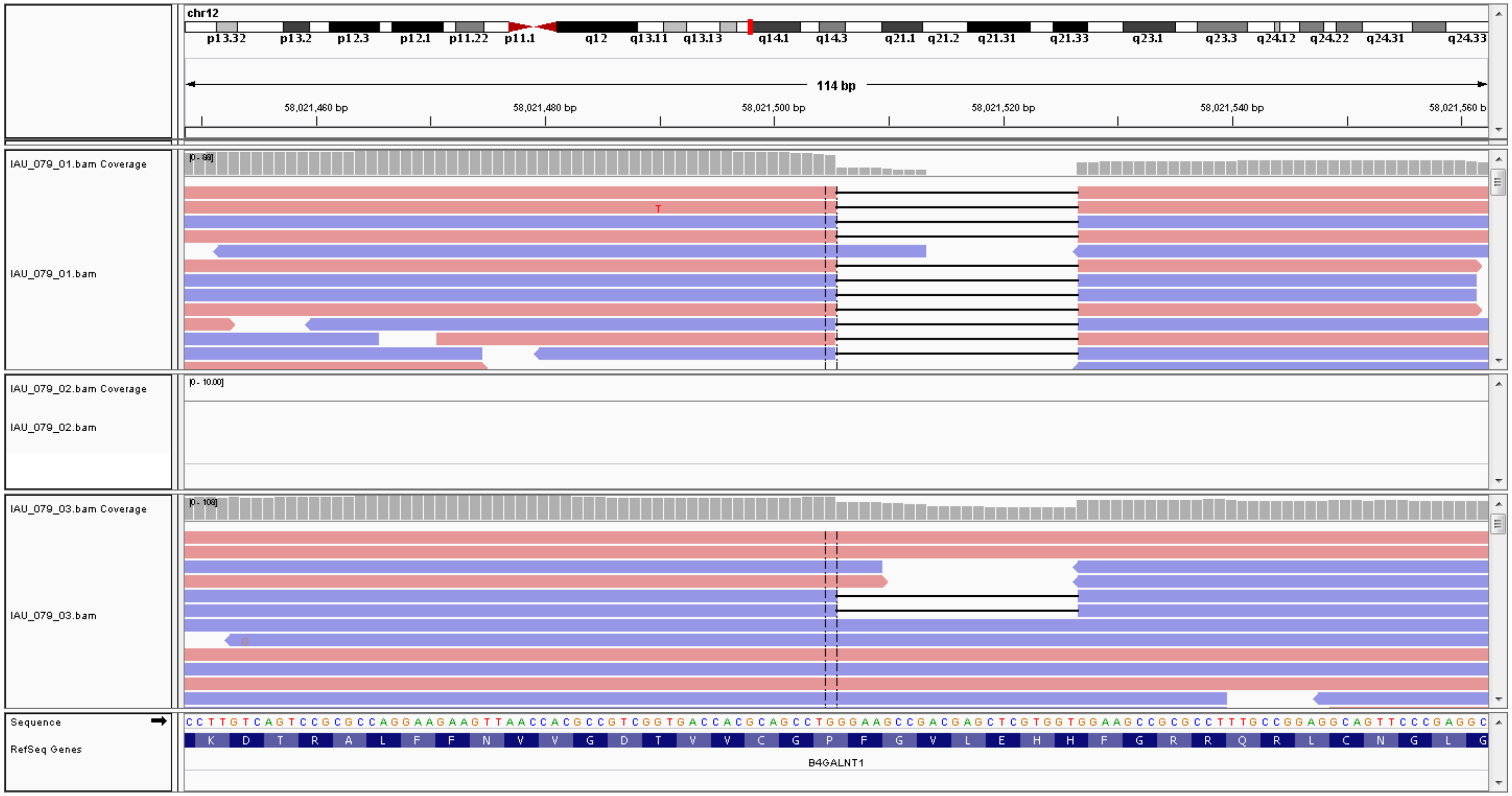


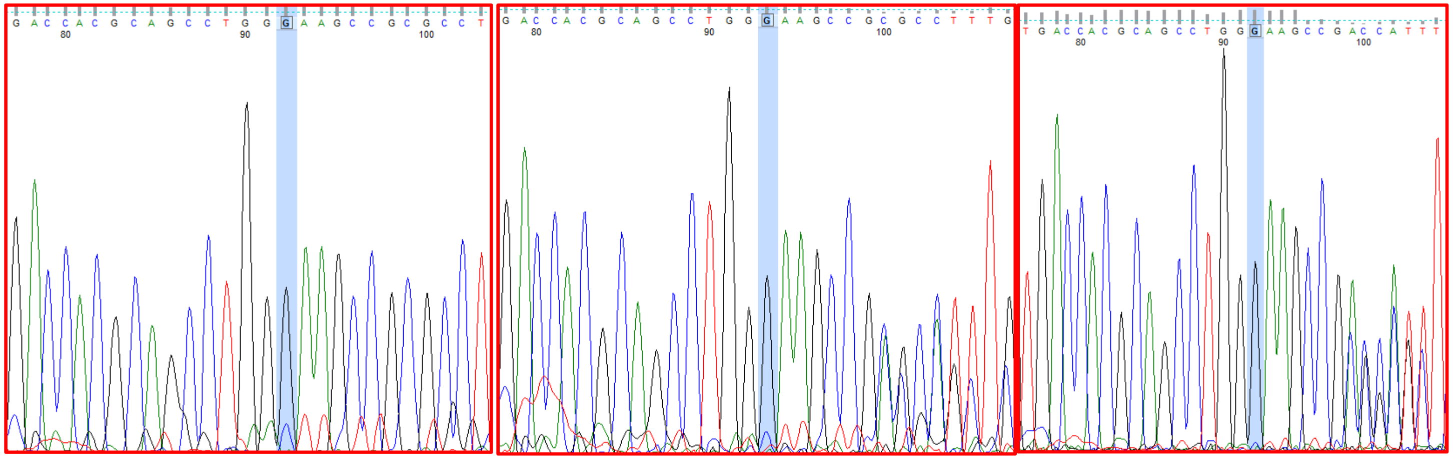

Proband Father Mother

36. IABB10: *CPM*

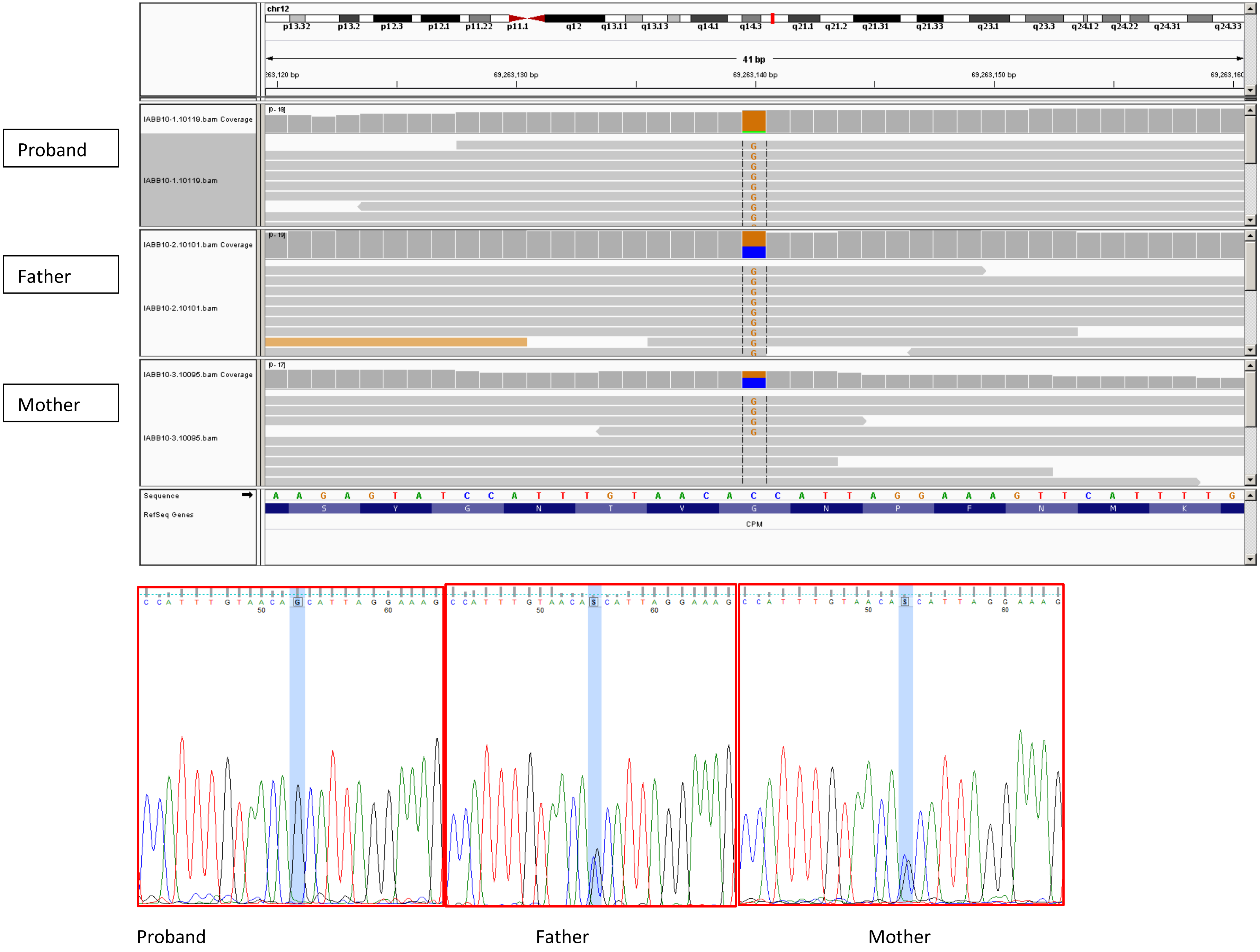

37. IABB11:*KSR2* (proband DNA exhausted)

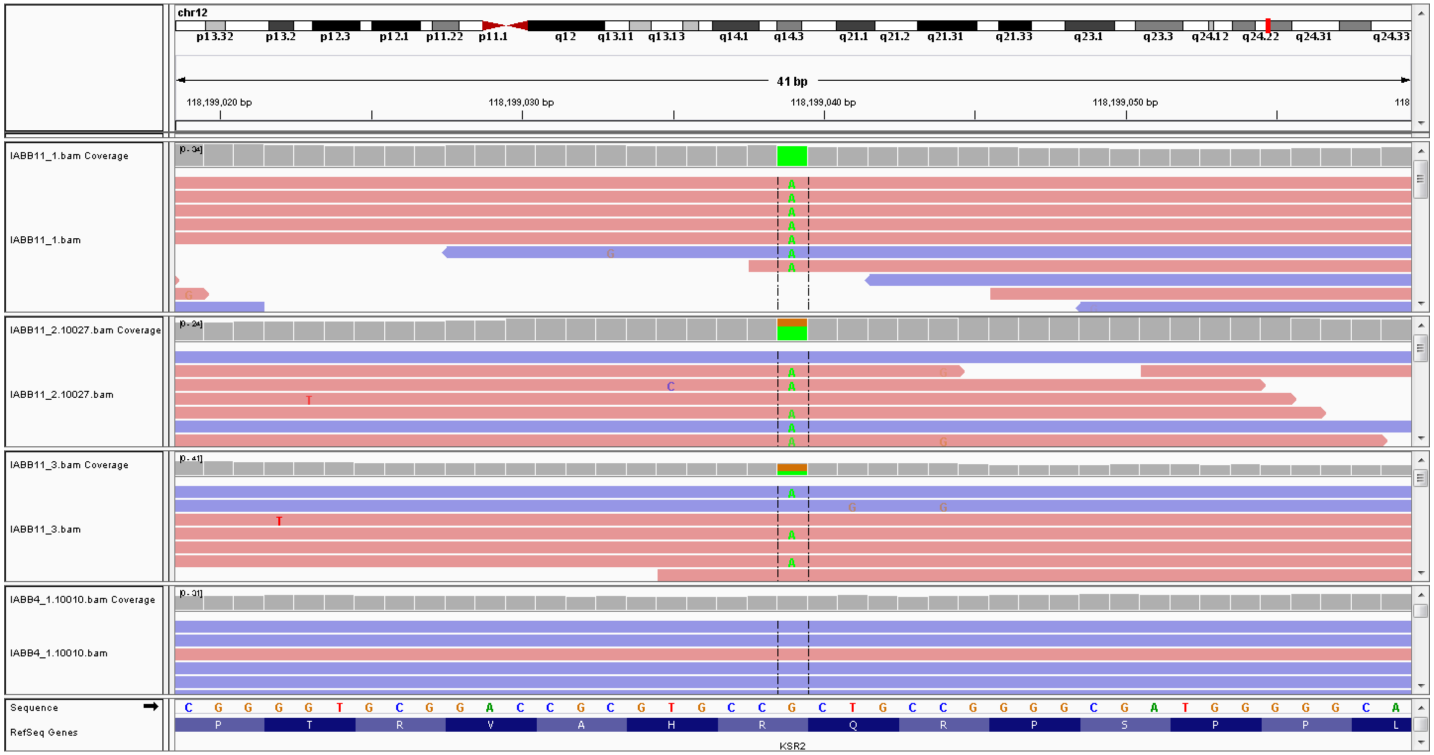


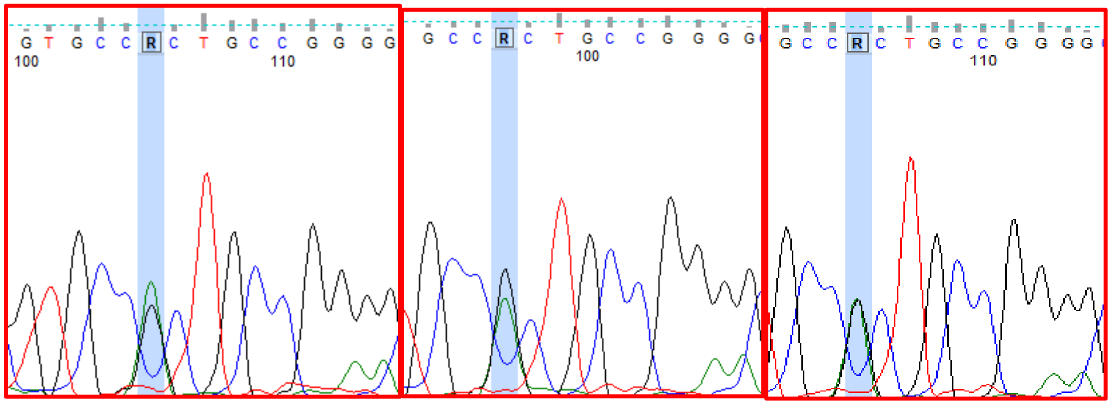

IABB11-3 IABB11-2 IABB11-4 (unaffected sibling)

38. SMPA8: *PPP1R36*

Proband

Father

Mother

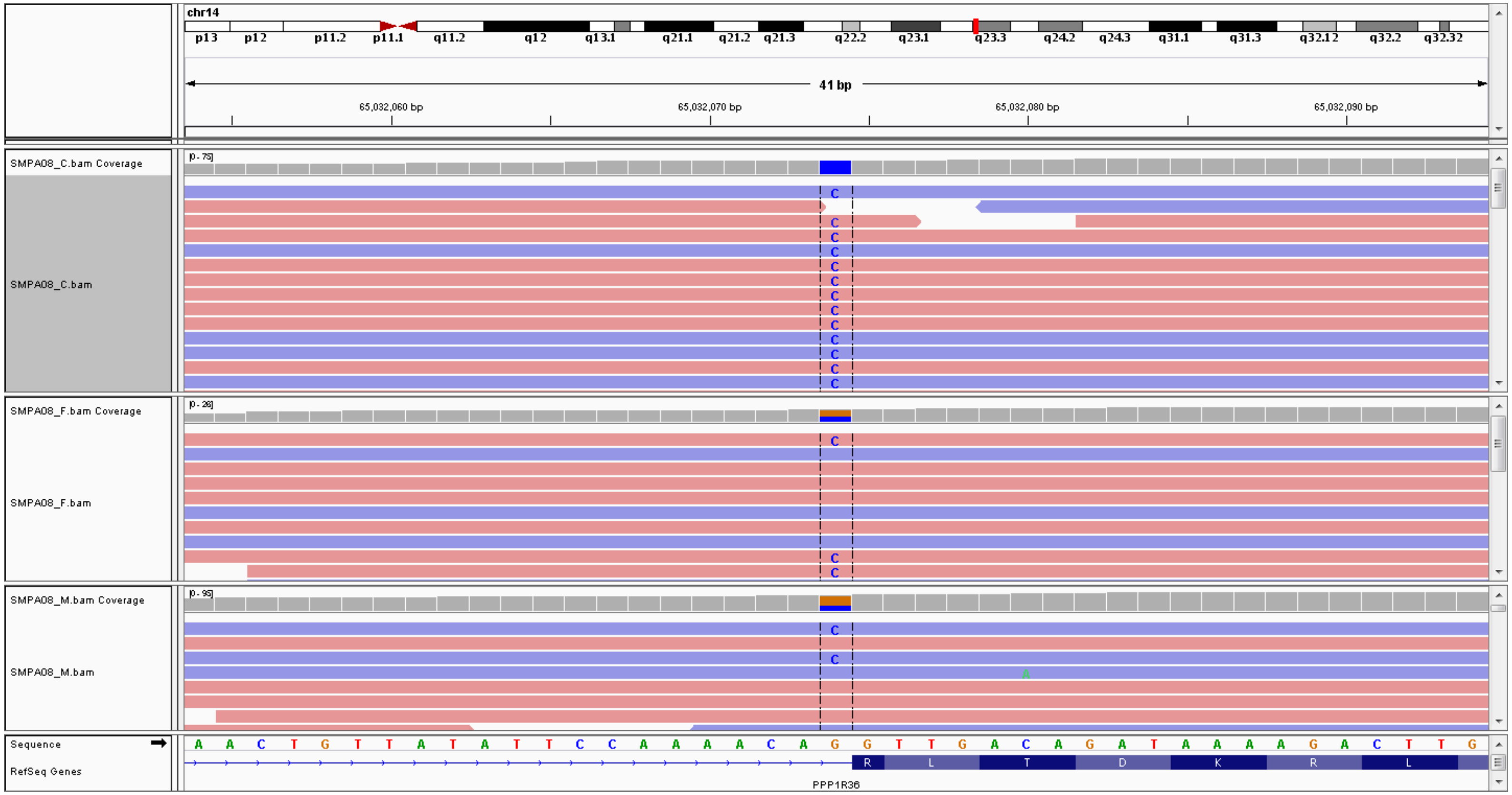


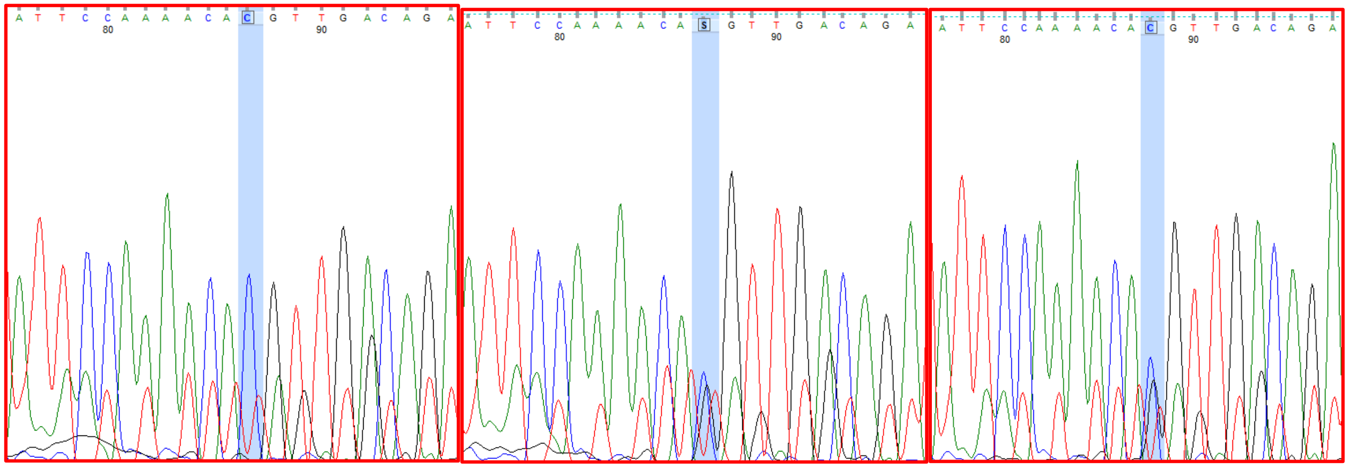

Proband

Father

Mother

39. IAH2: *TECPR2*

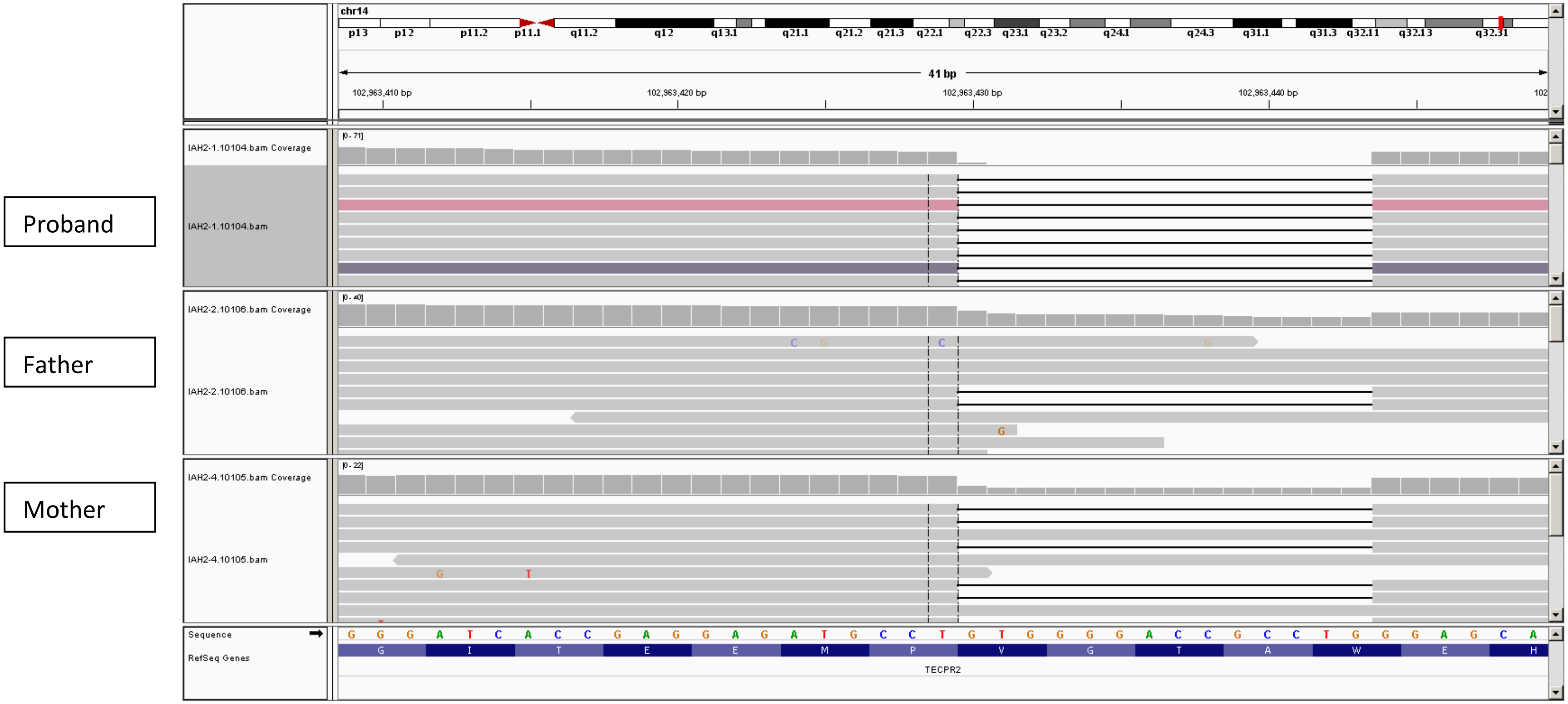


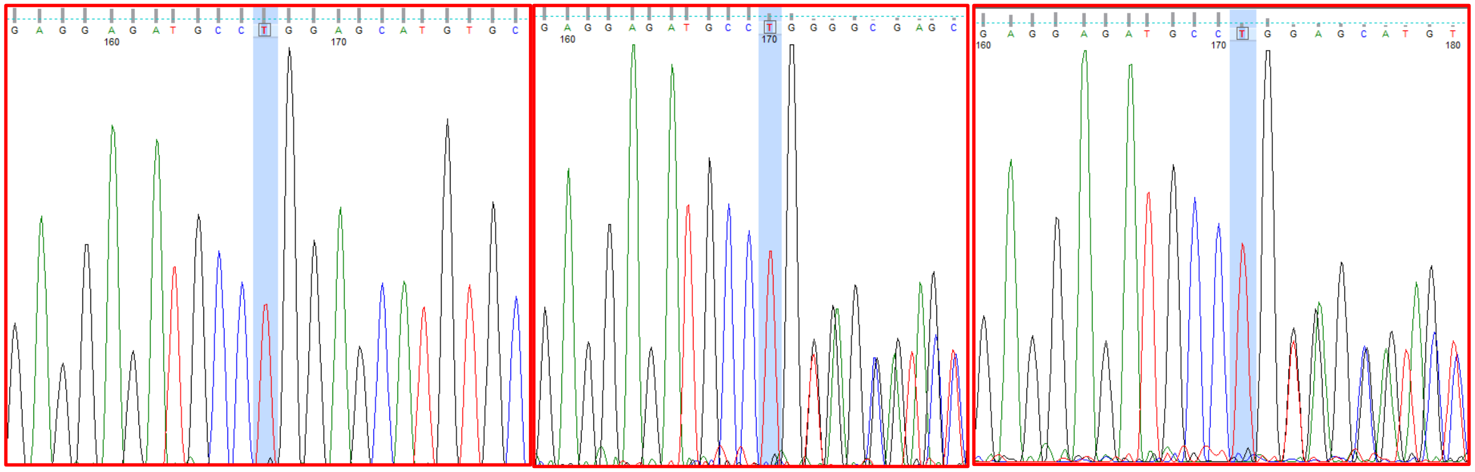

40. IABB4: *LINS1*

Father

Mother

Proband

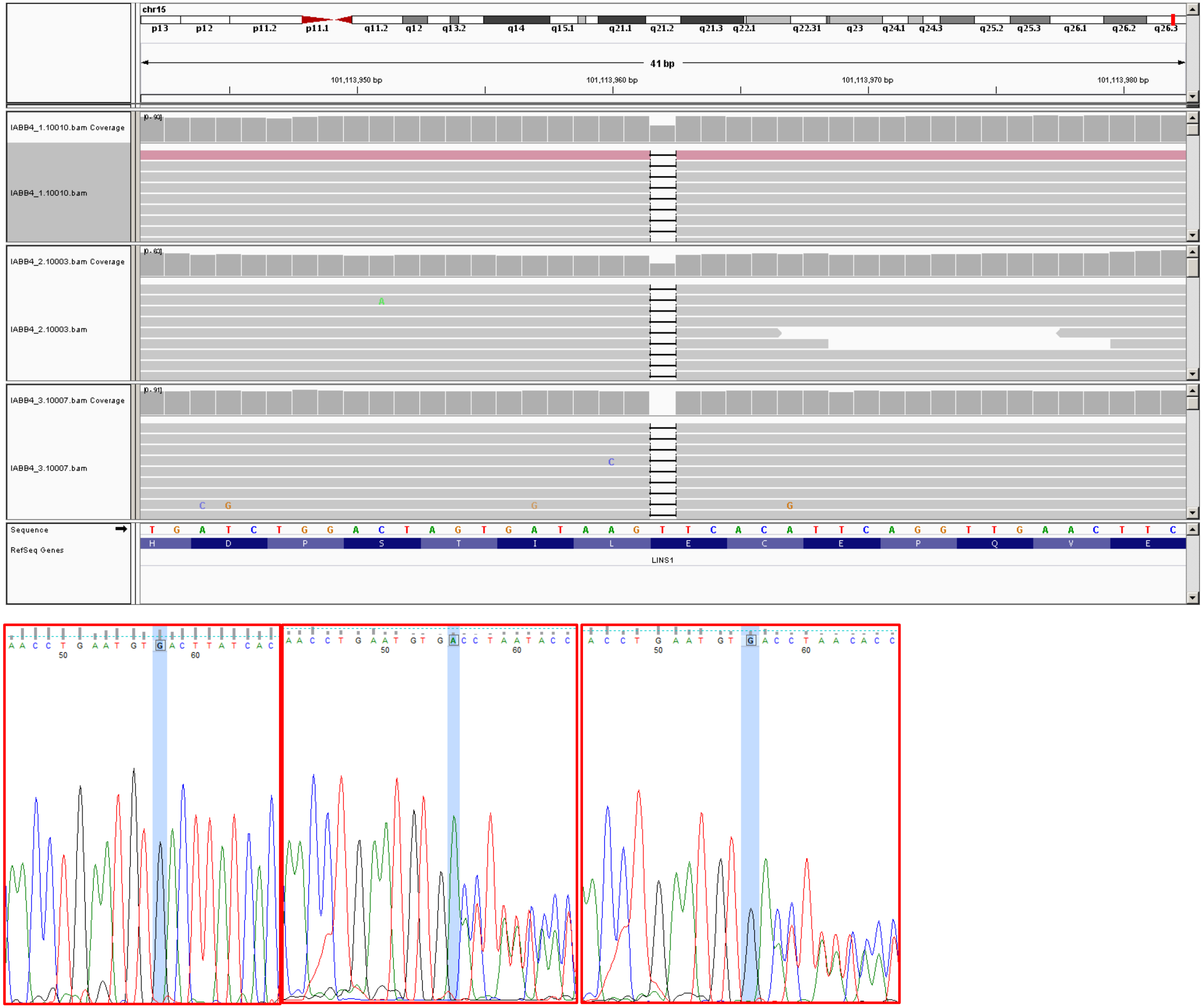


Proband

Father

Mother

41. SMPA76: *WDR90*

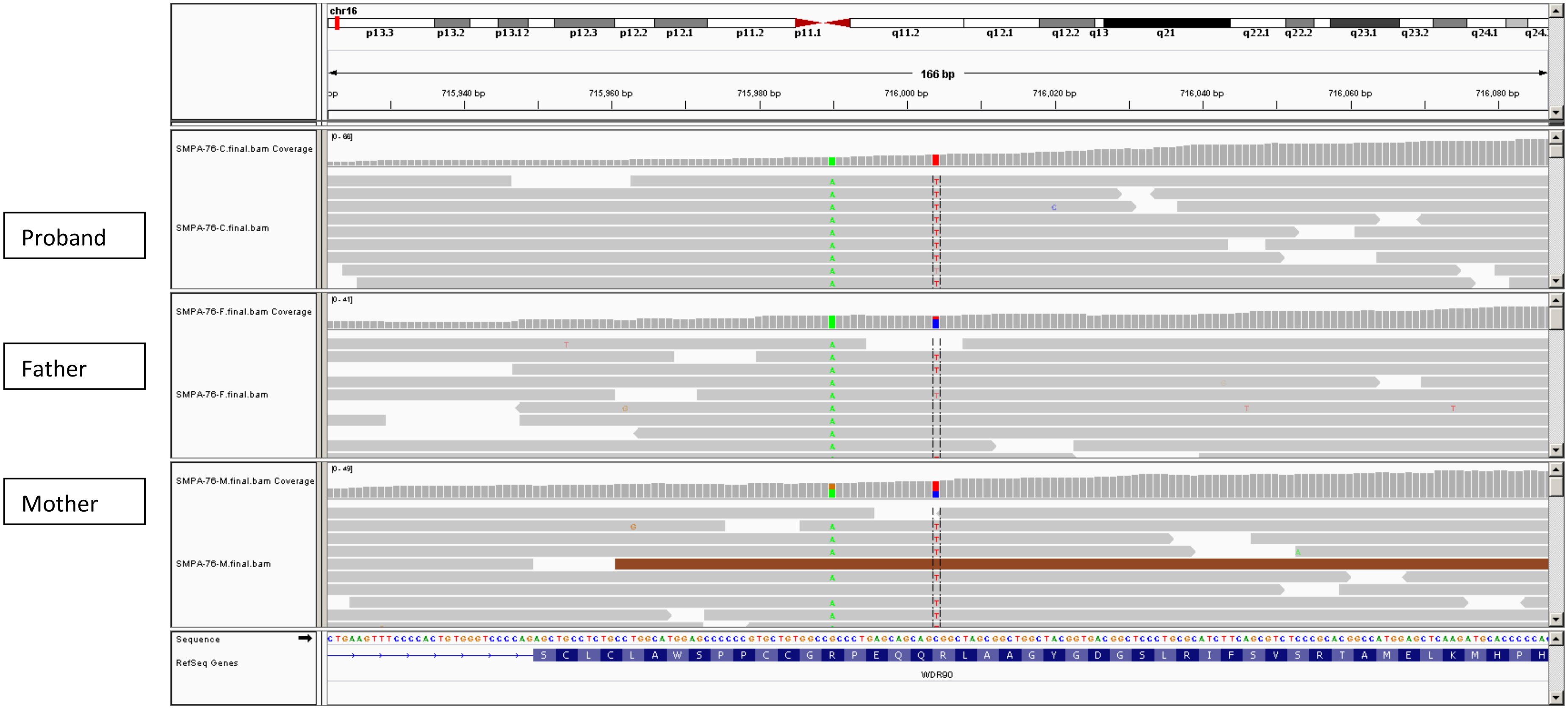


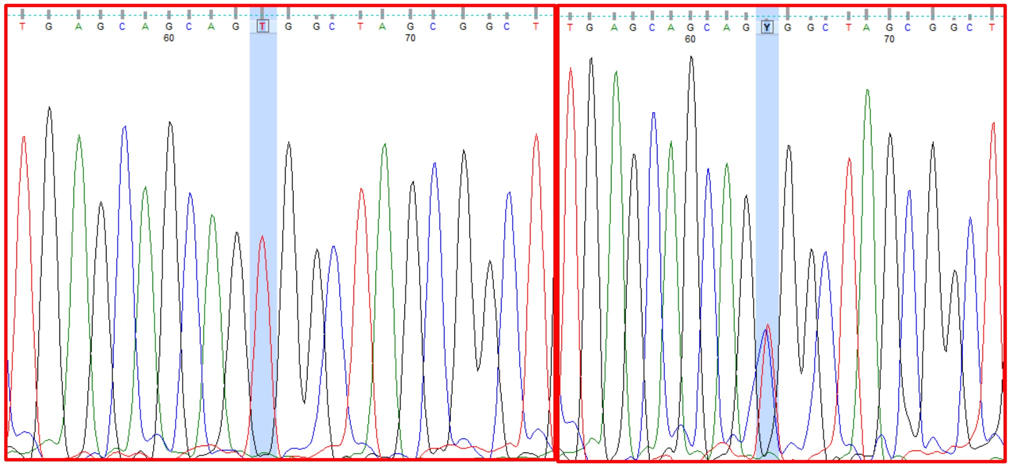

Proband

Father

42. IABB14: *ASPA*

Proband

Father

Mother

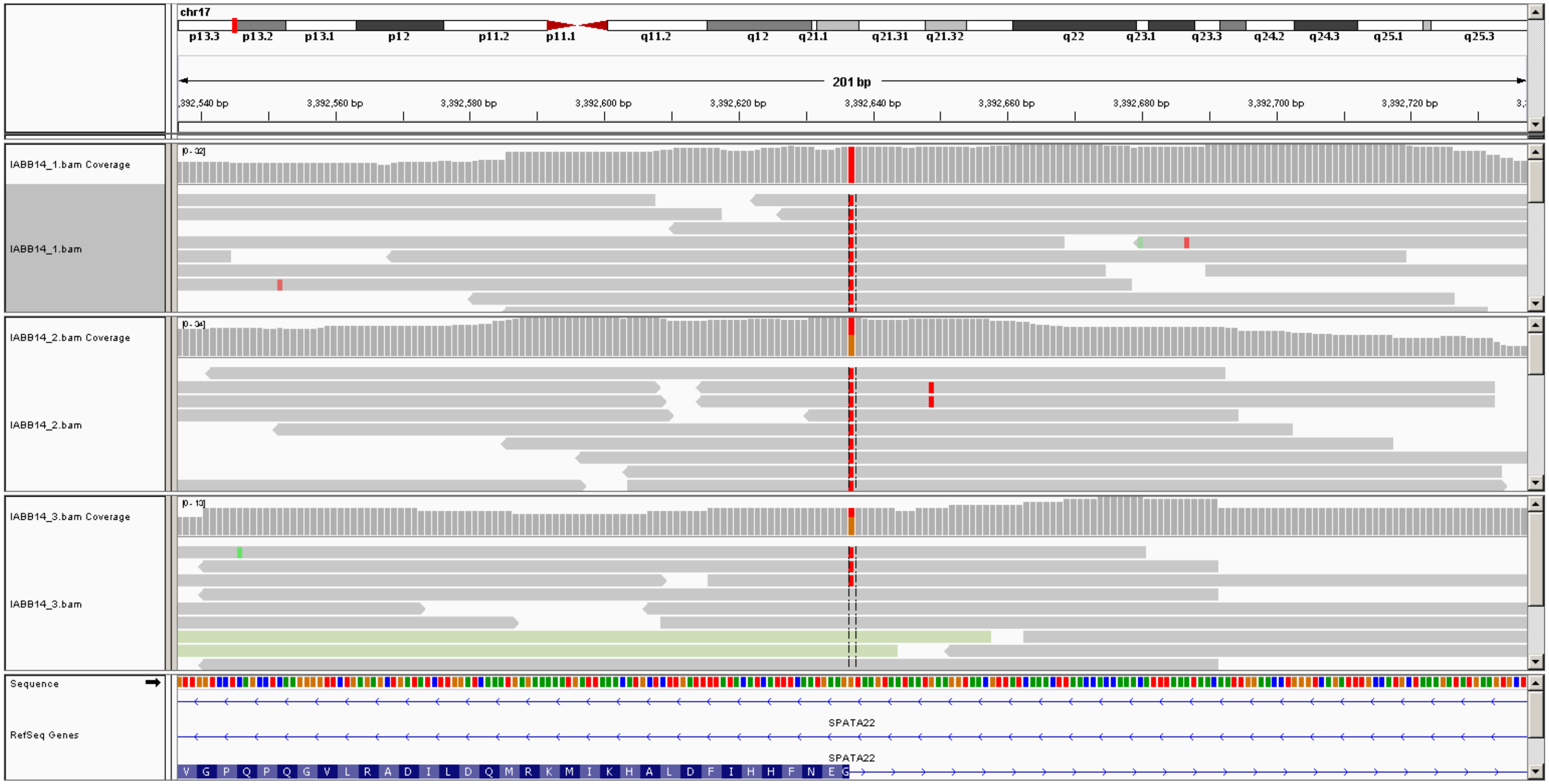


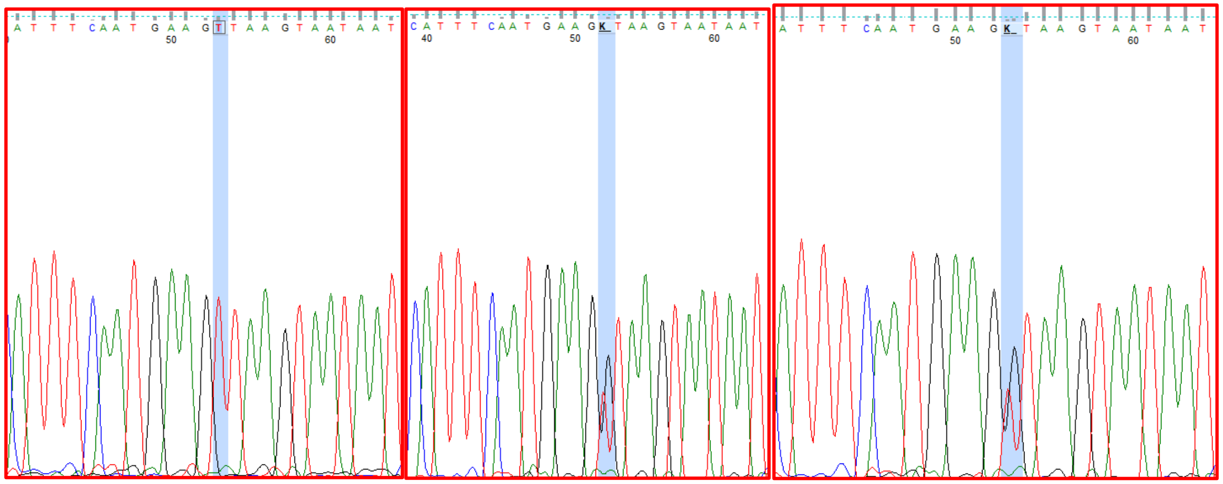

Proband

Father

Mother

43. SA2: *CC2D1A*

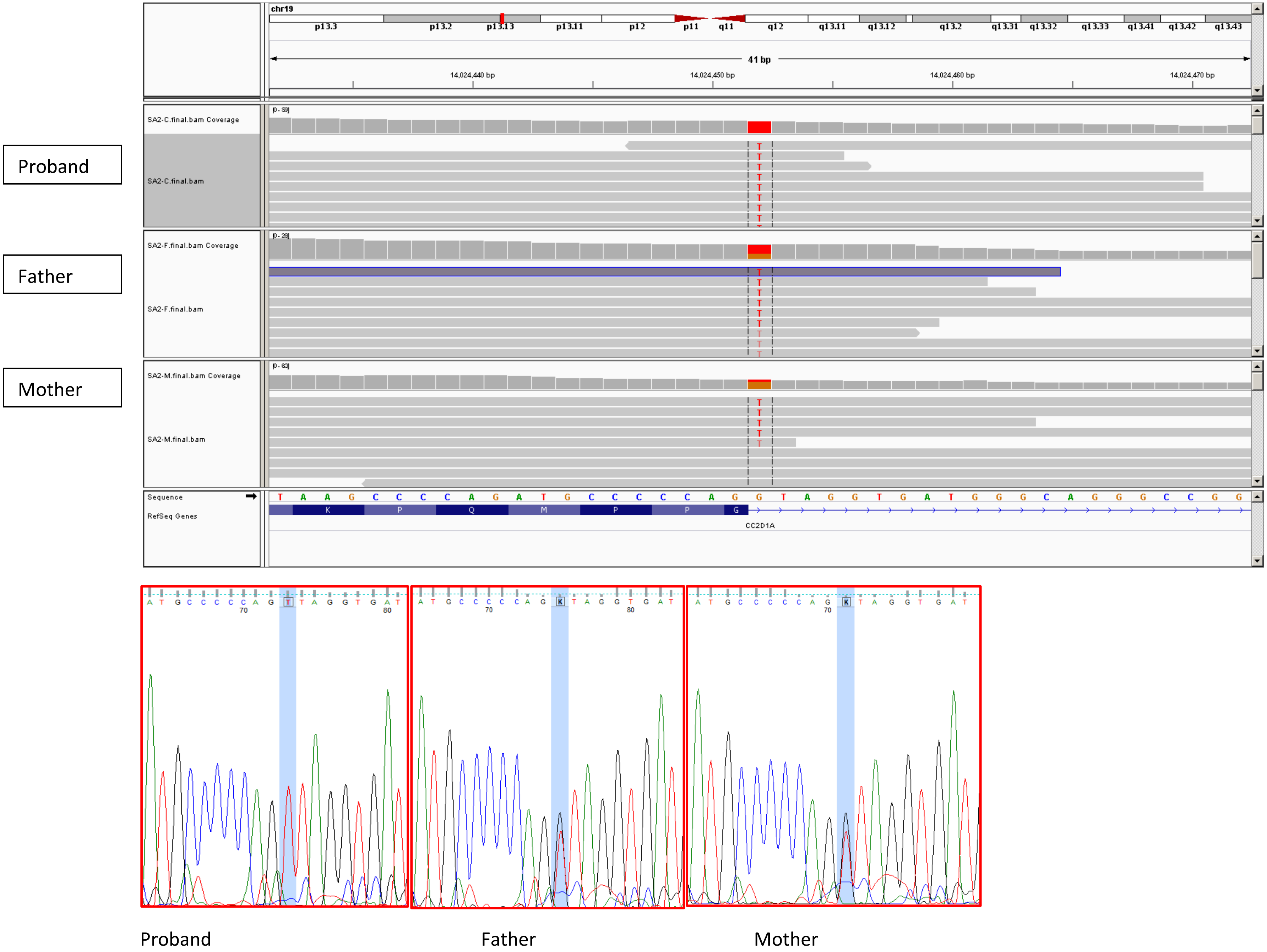

44. IABB16: *MED25*

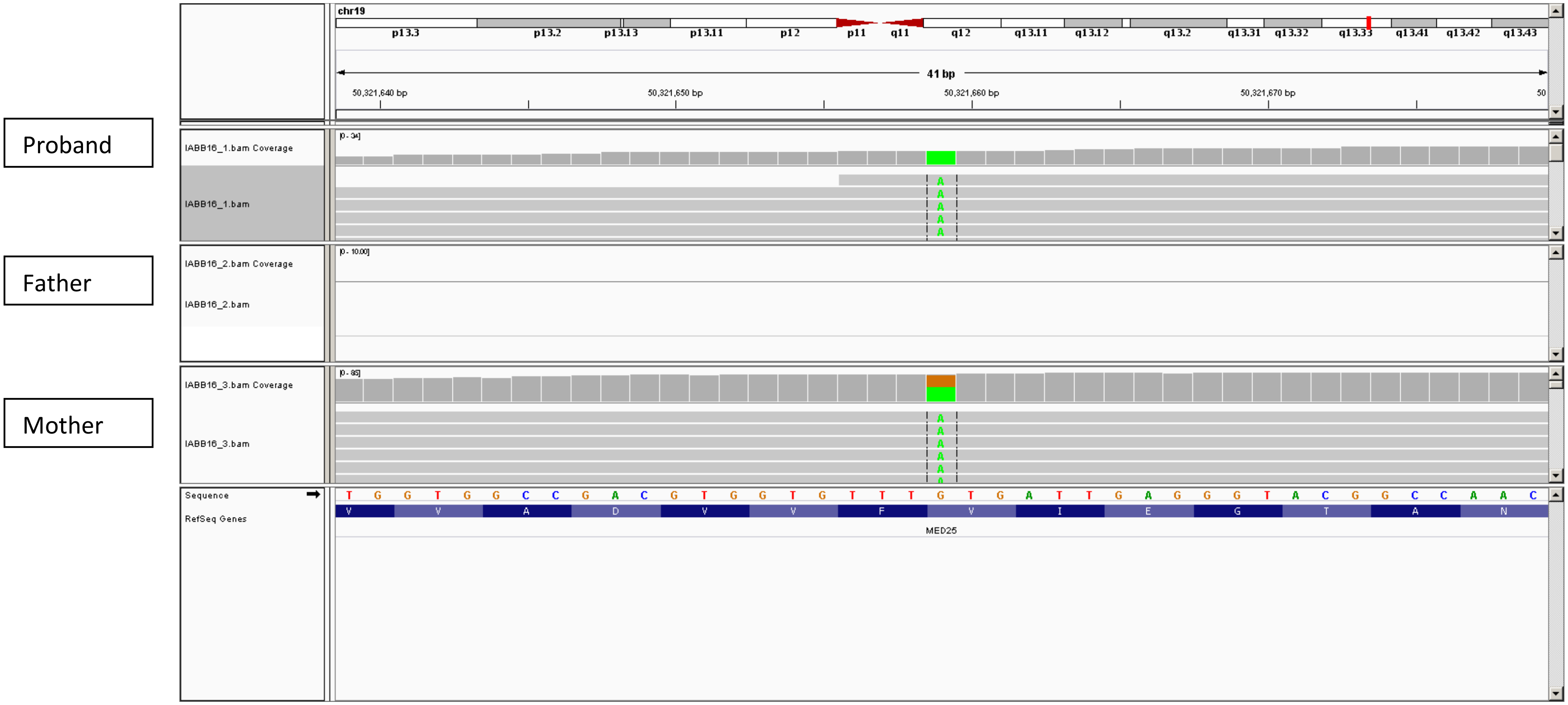


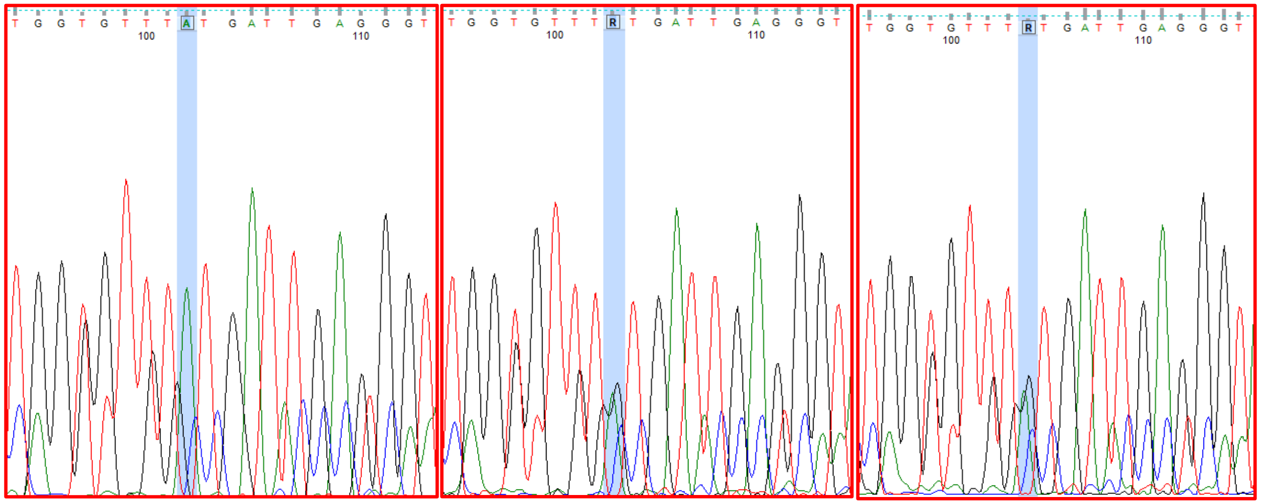

Proband Father Mother

45. SA11: ZNF766

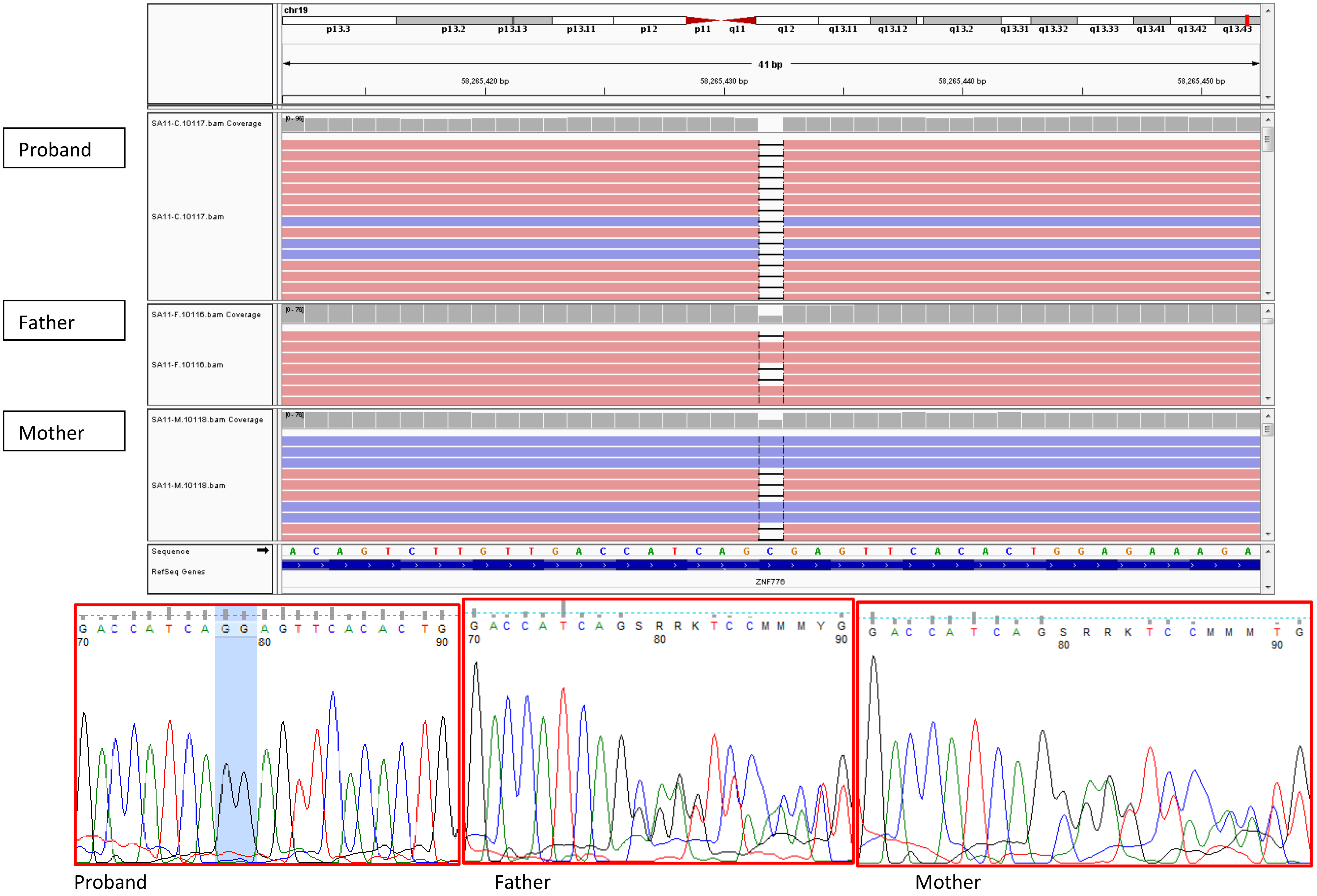

46. IABB16: *VPS16*

Proband

Father

Mother

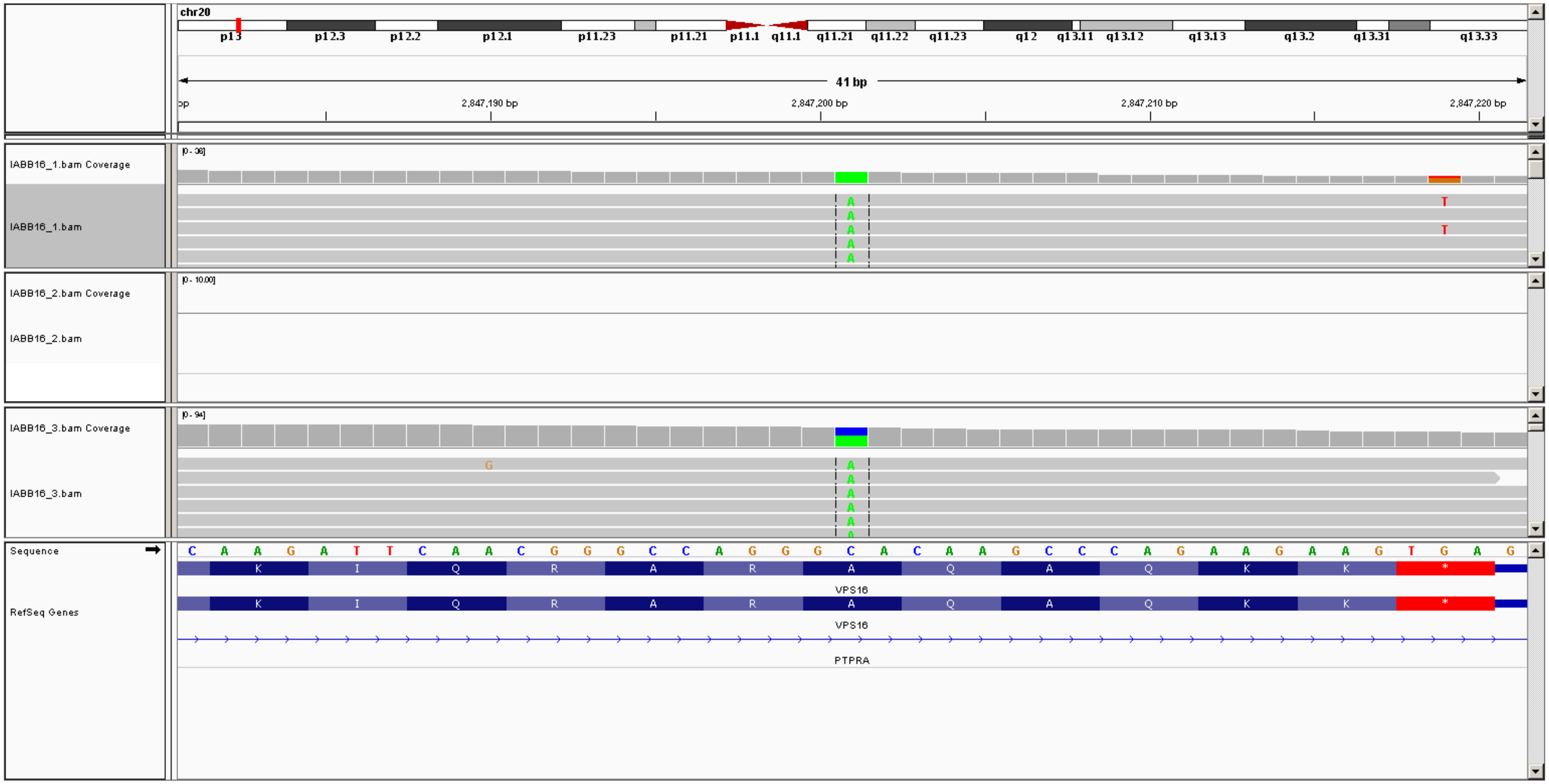


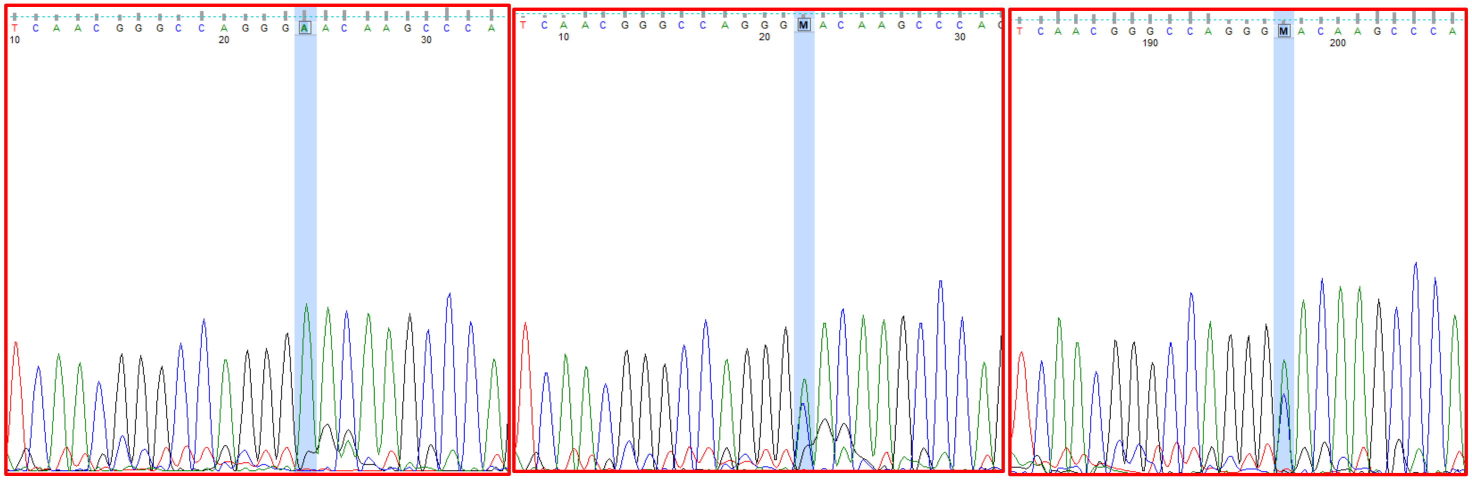

Proband

Father

Mother

47. IABB16: *KIF16B*

Proband

Father

Mother

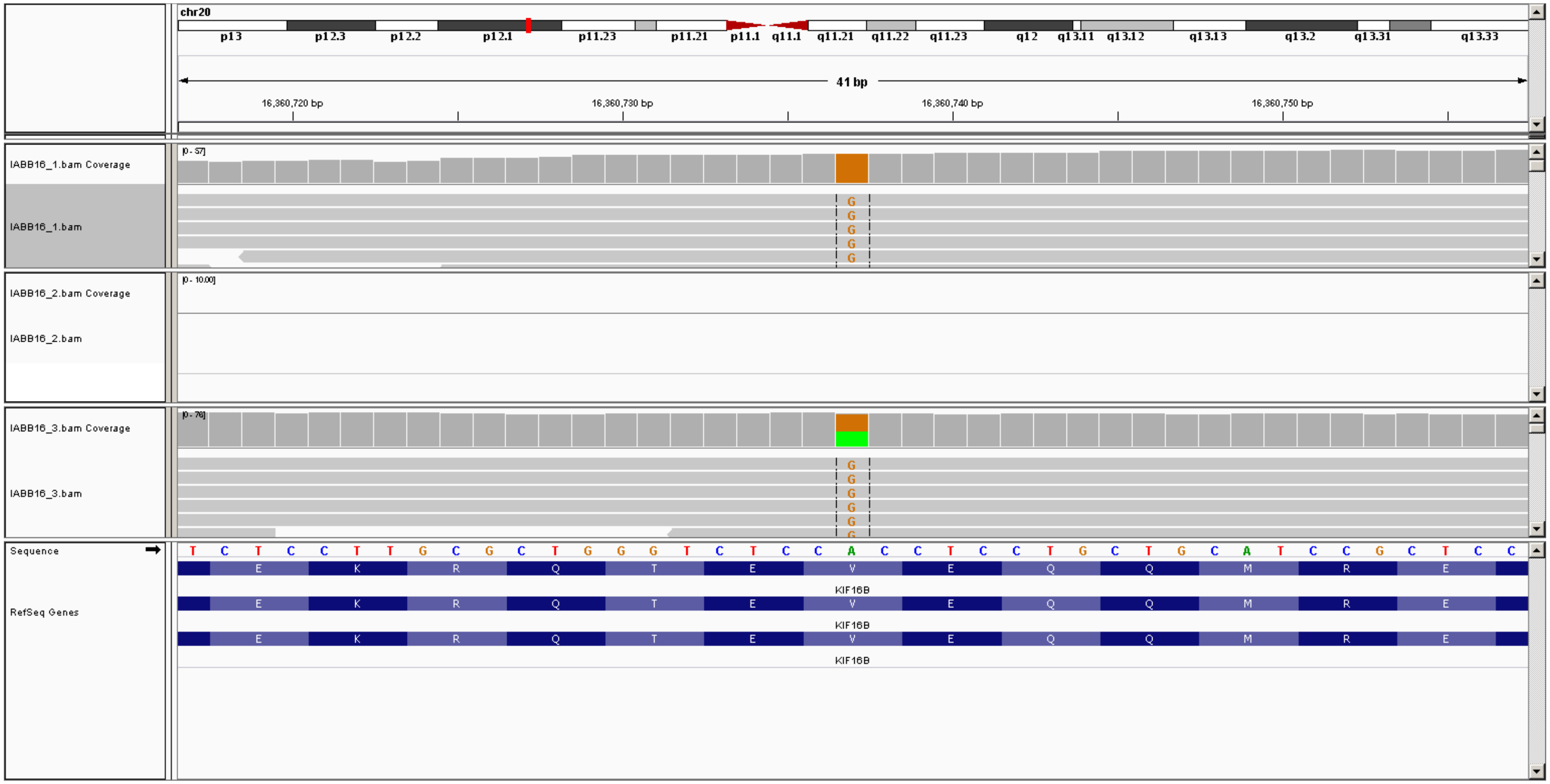


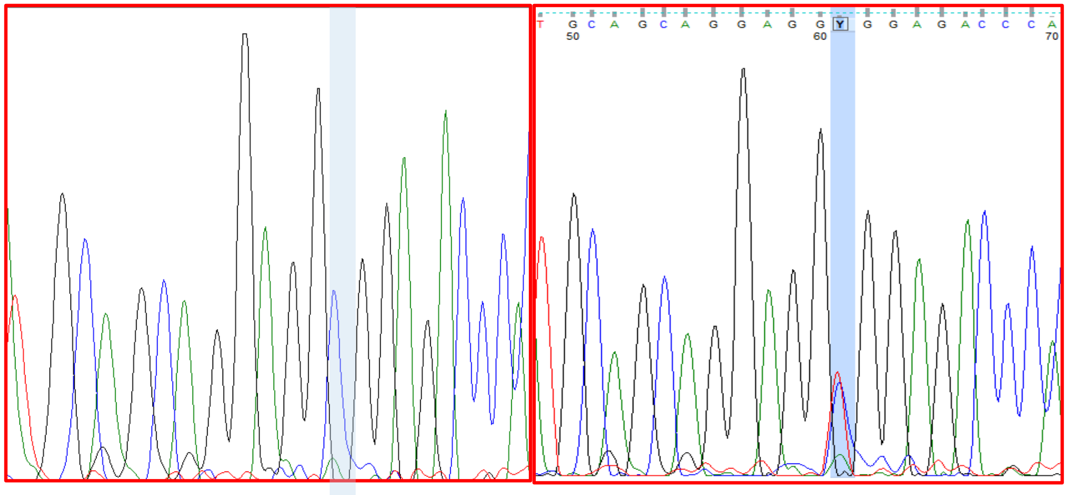

Proband Mother

48. IABB5: *ZNF335*

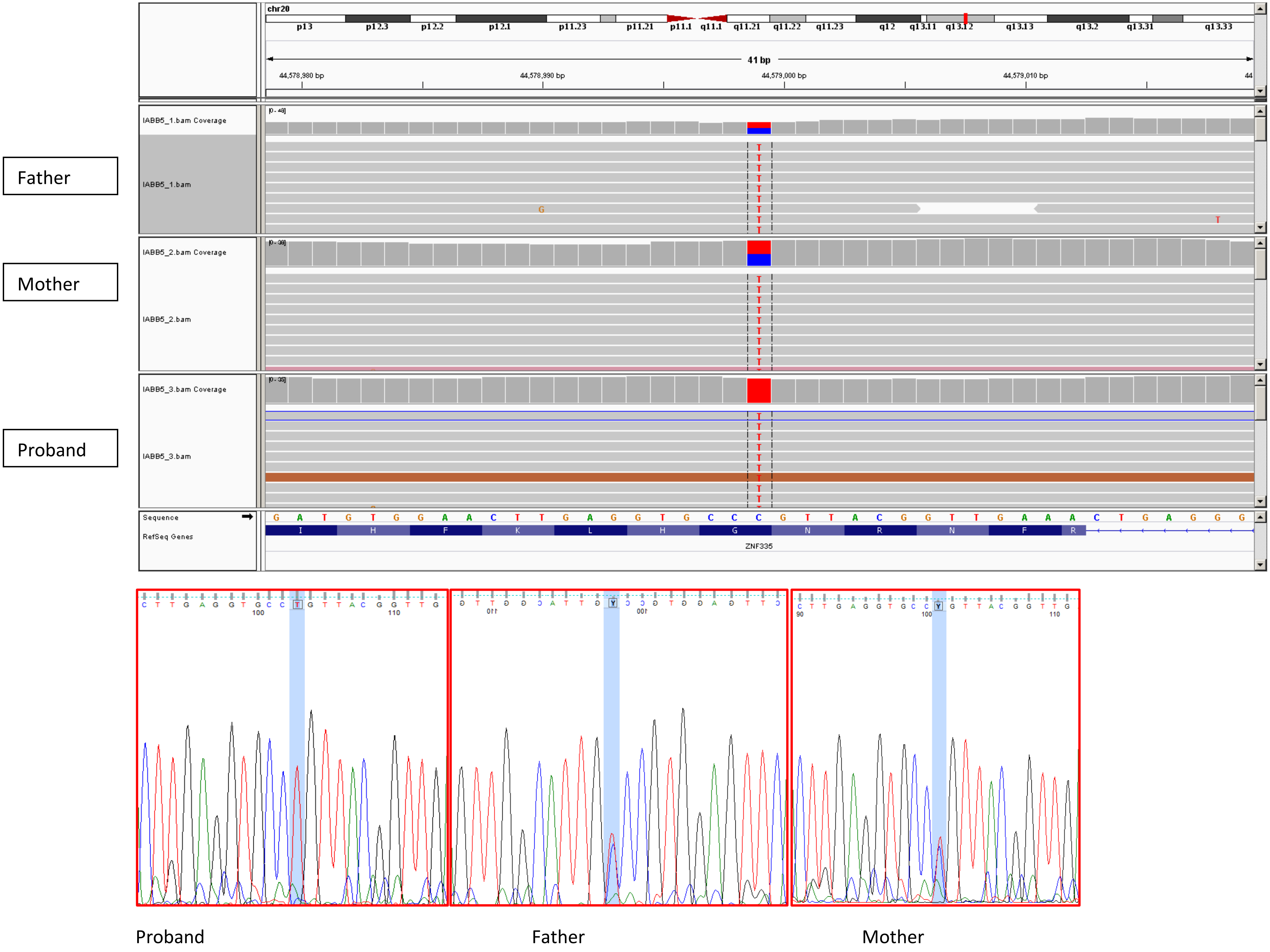

49. IABB11: *SGSM3*

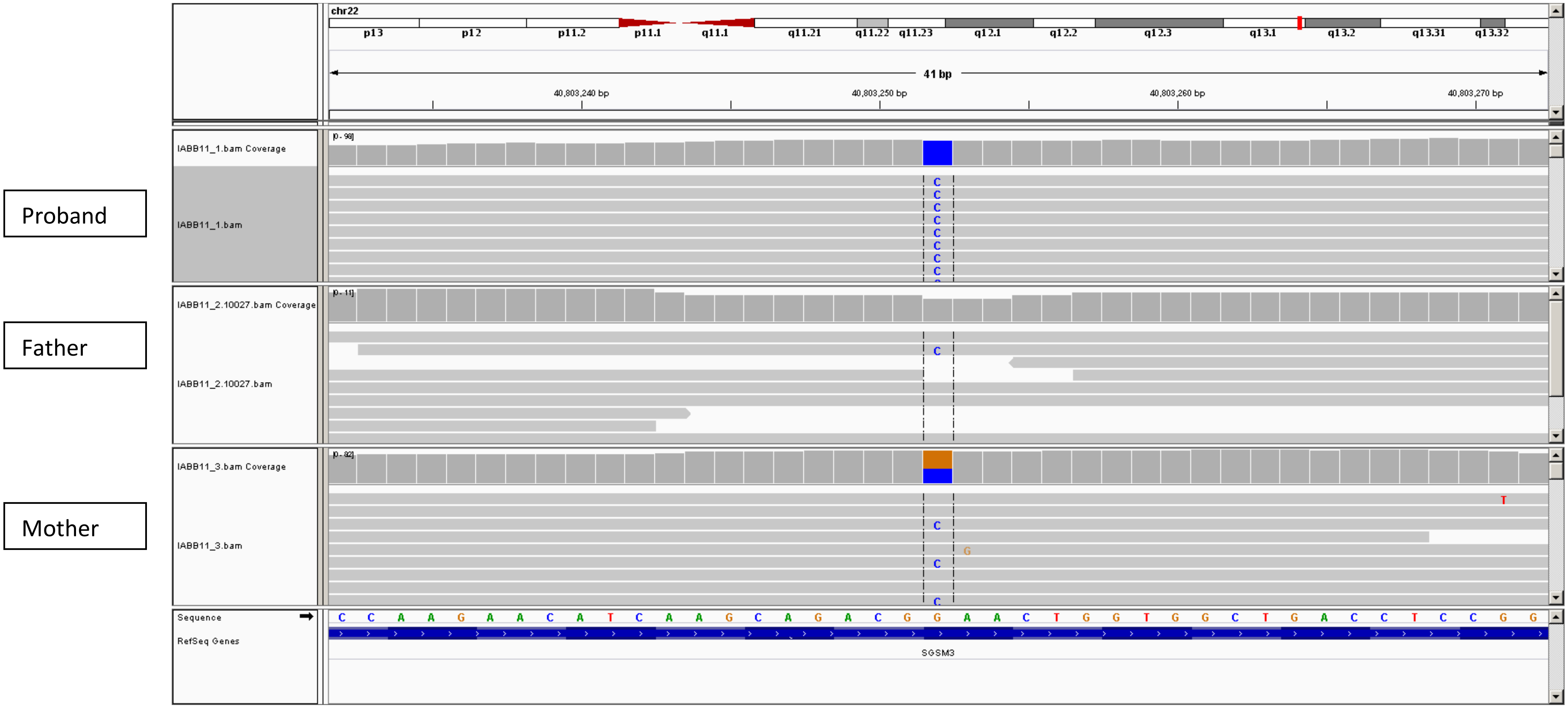


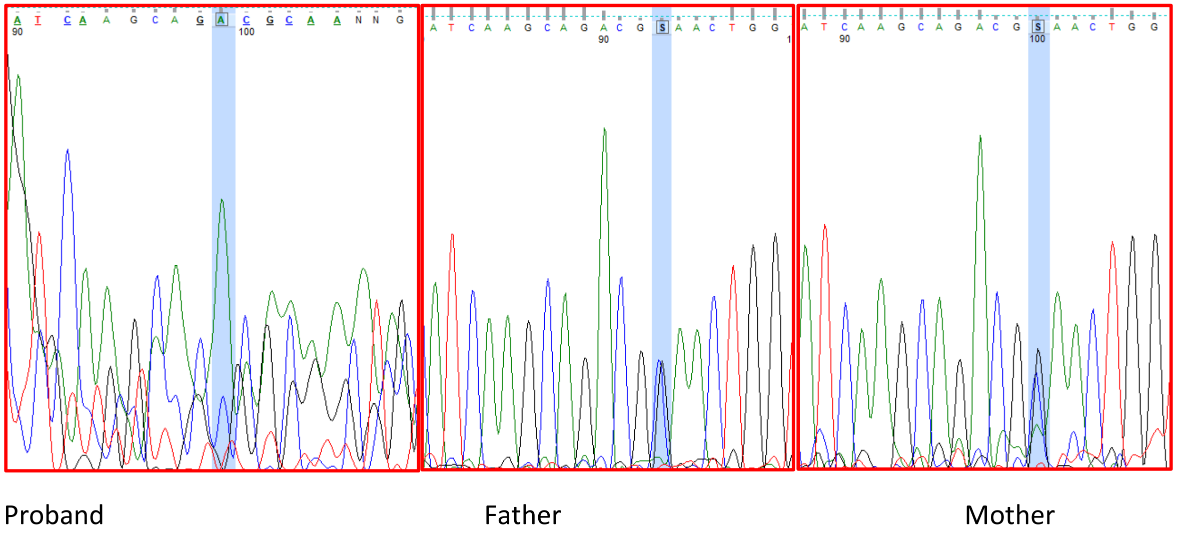

**Table 3: De novo/dominant**

1. RQPA20: *RAB42*

Proband

Father

Mother

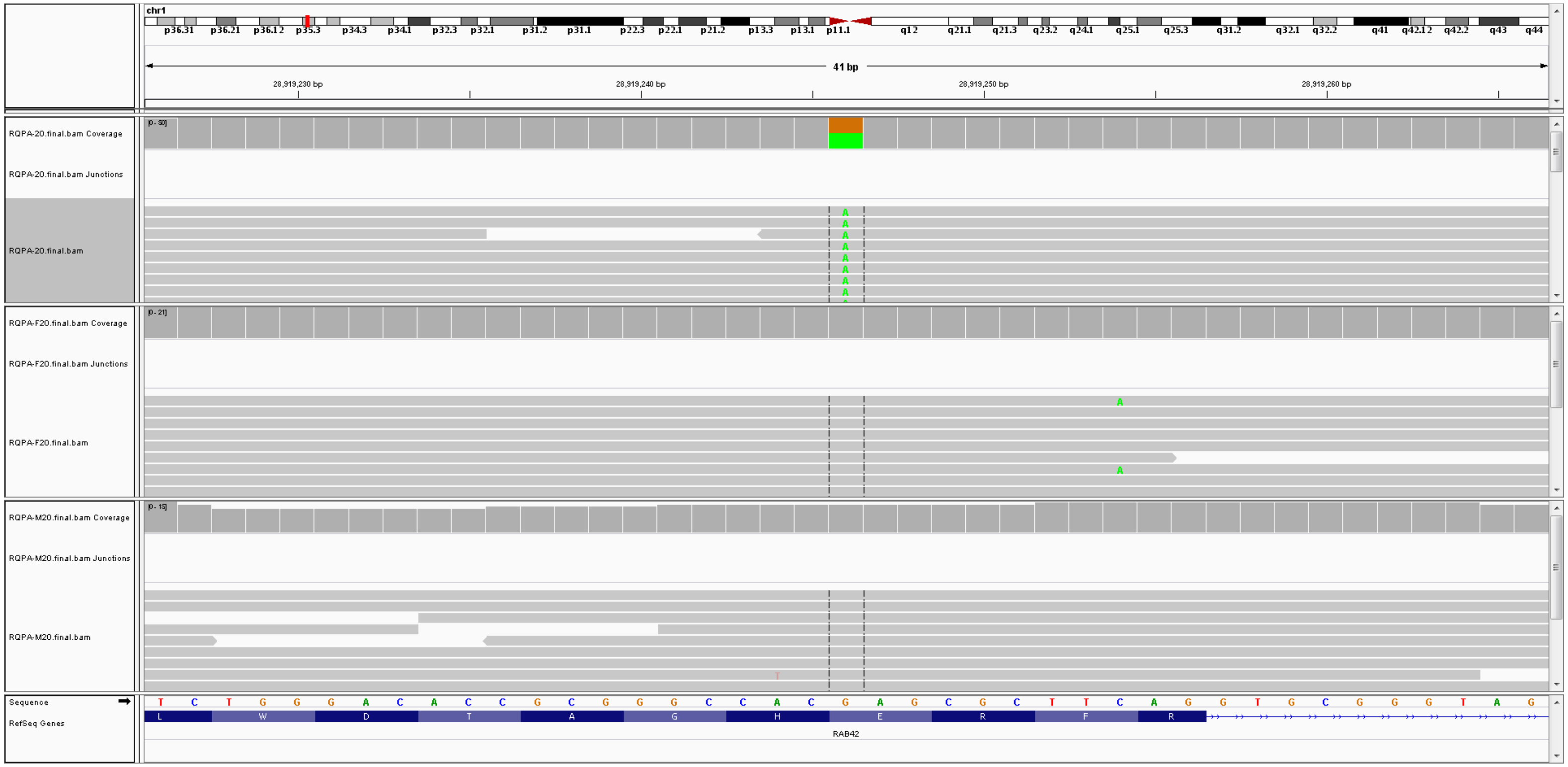


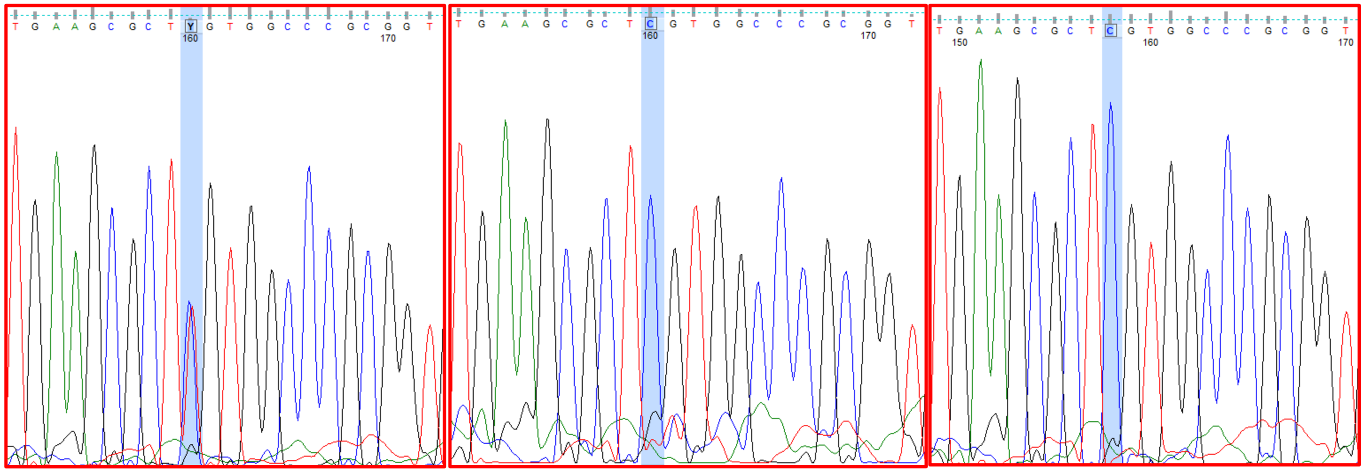

Proband Father Mother

2. RQPA20: *TMIGD3*

Proband

Father

Mother

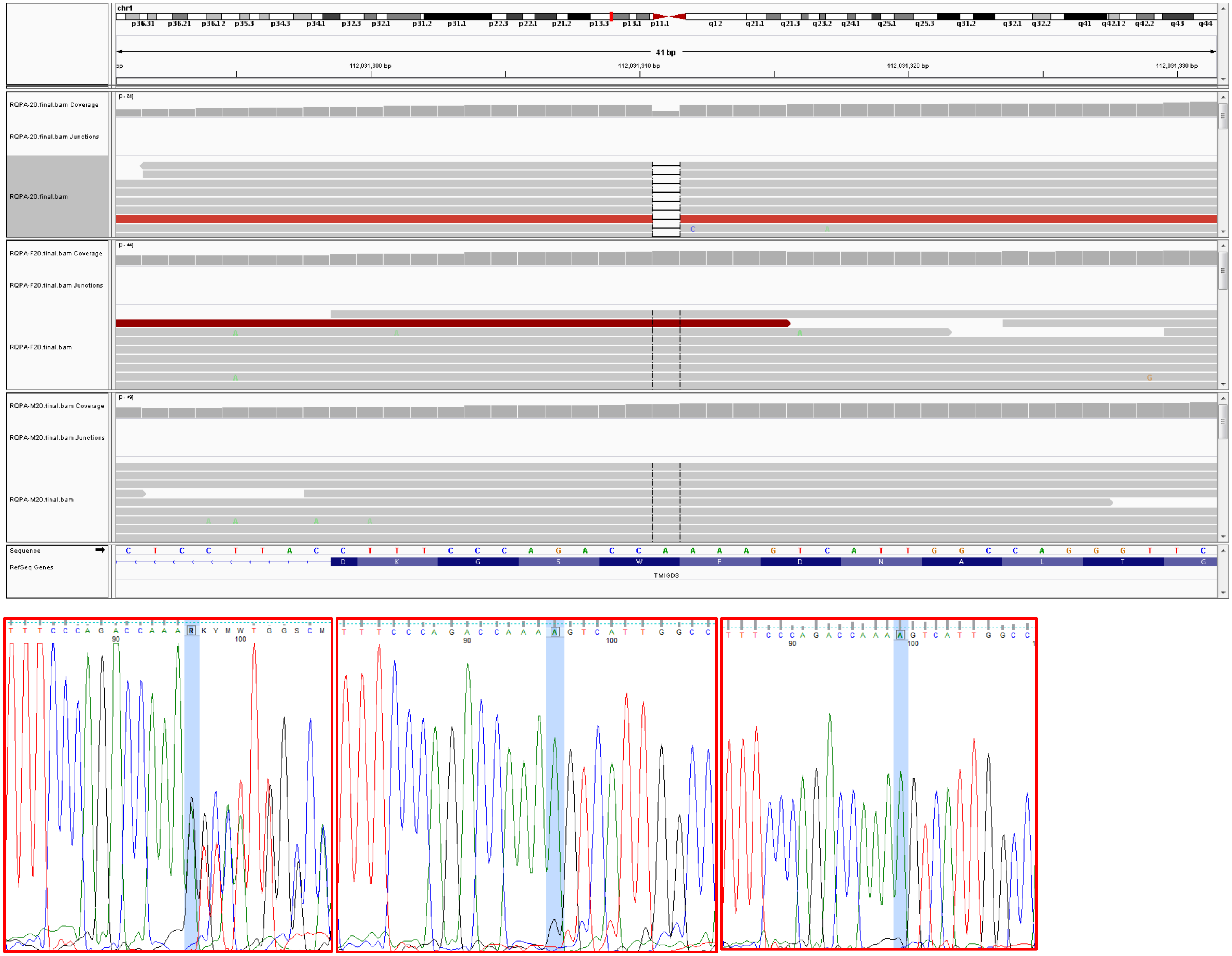


Proband Father Mother

3. RQPA20: *ECM1*

Proband

Father

Mother

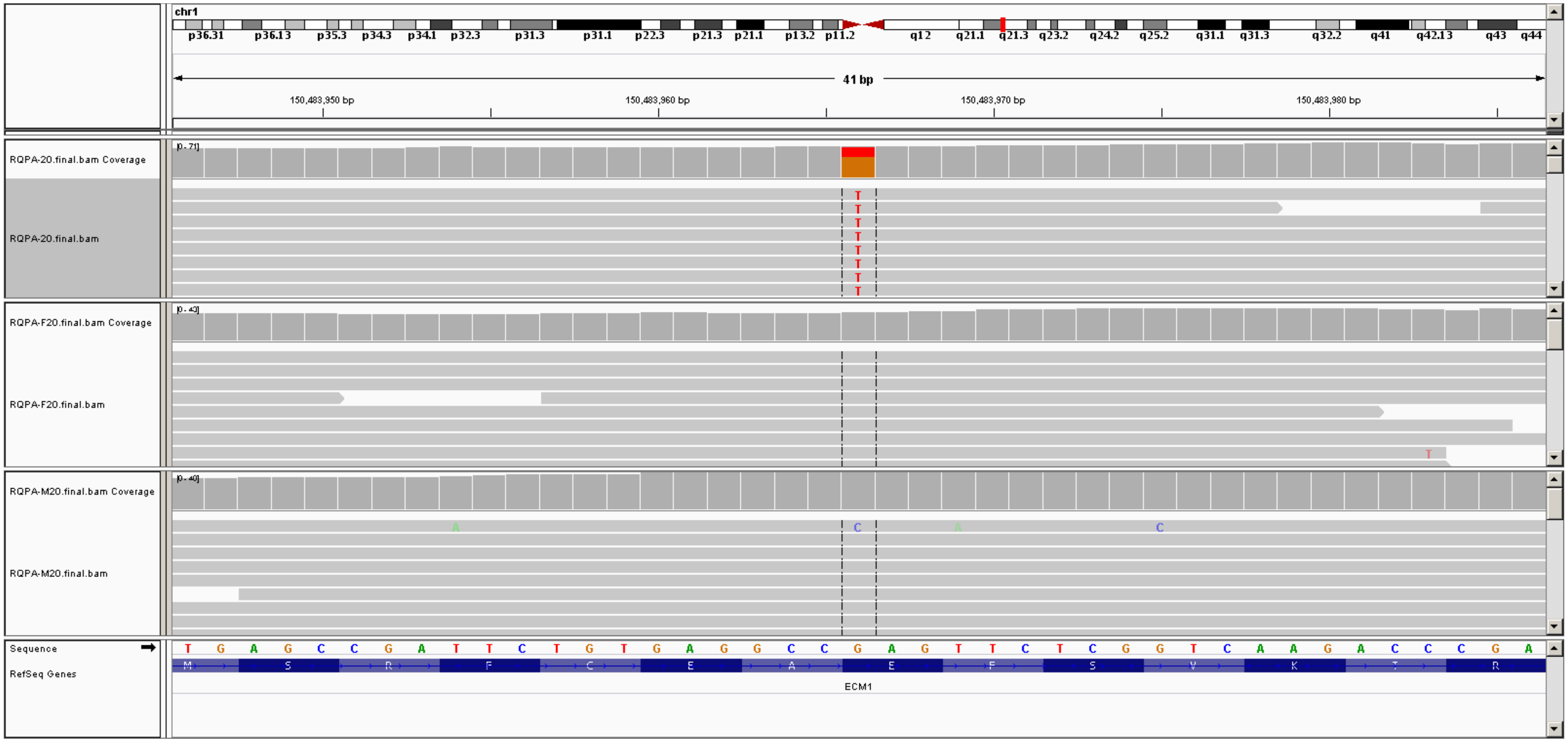


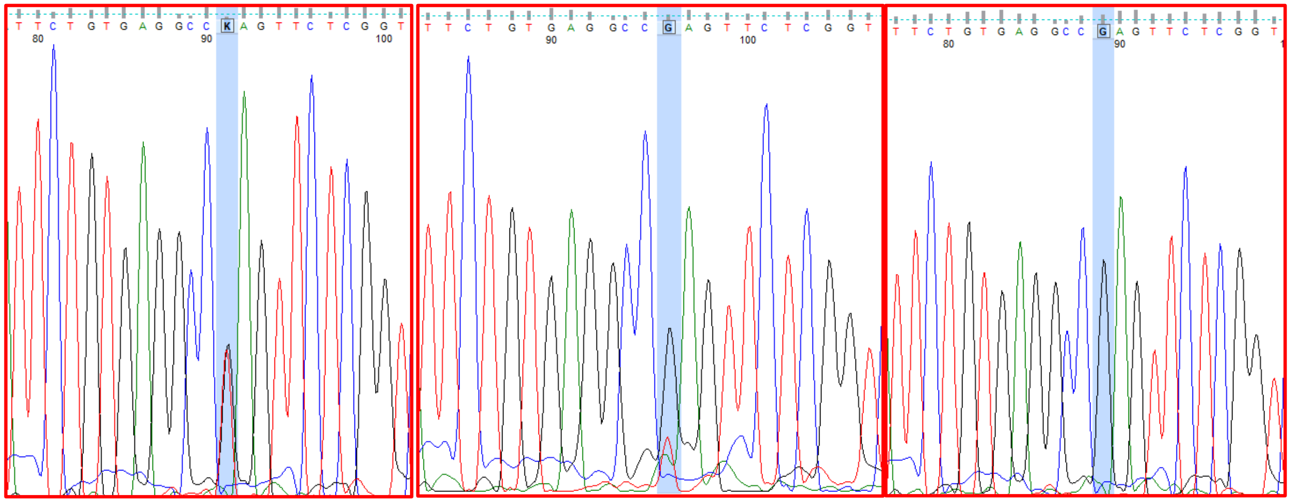

Proband Father Mother

4. RQPA20: *SLAMF7*

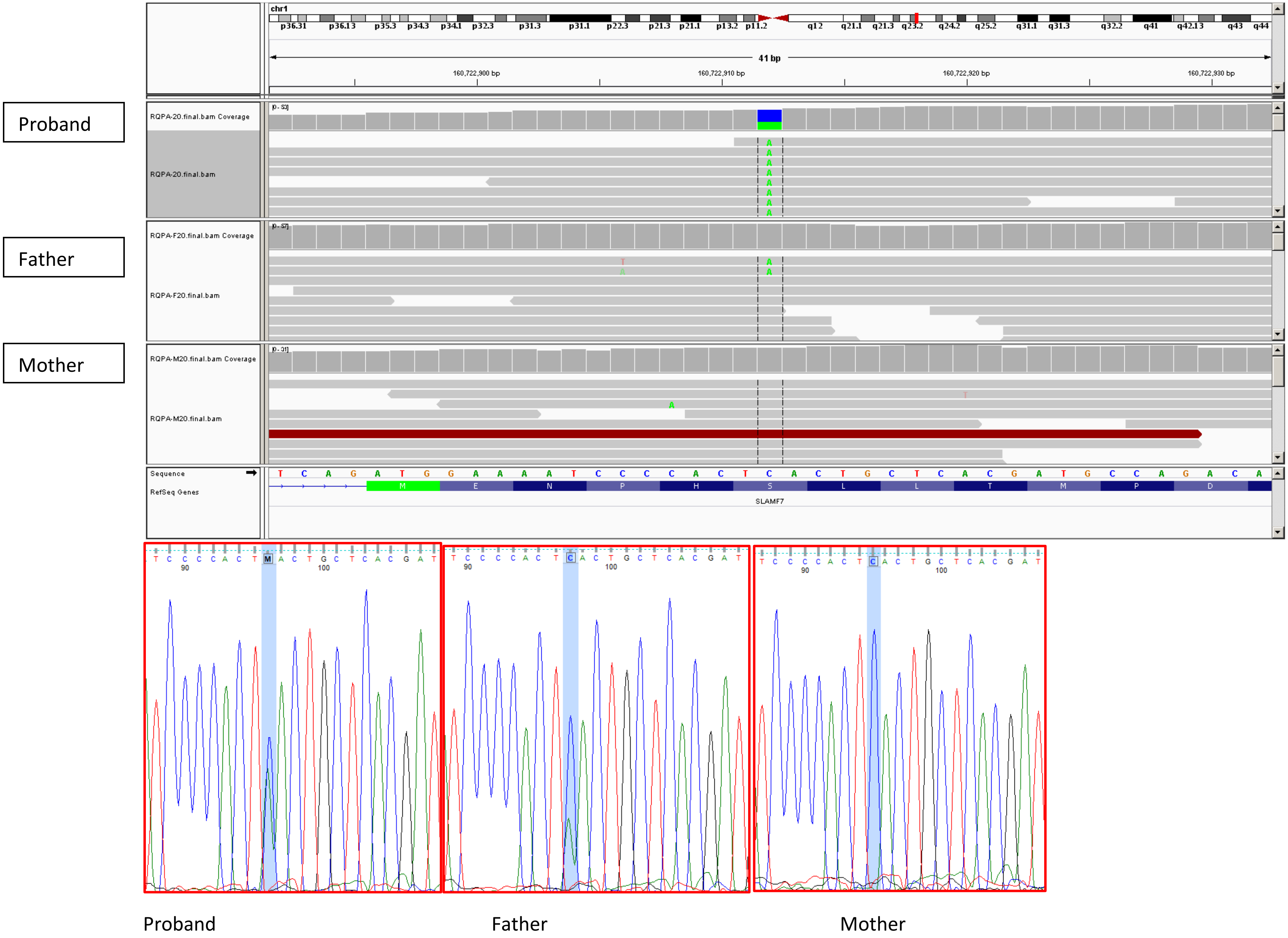

5. IAU29: *MYT1L*

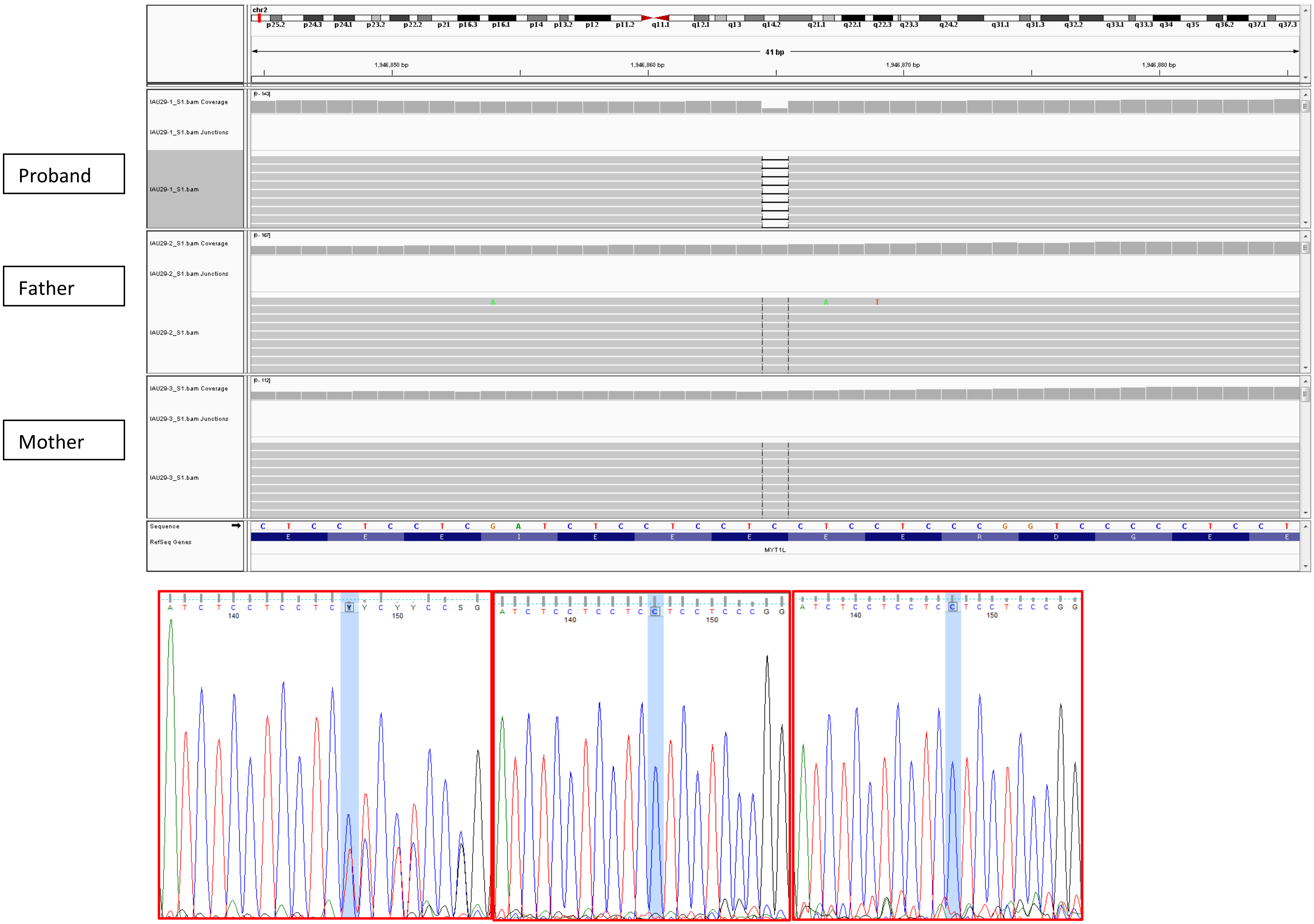


Proband Father Mother

6. SMPA53: *SCN2A*

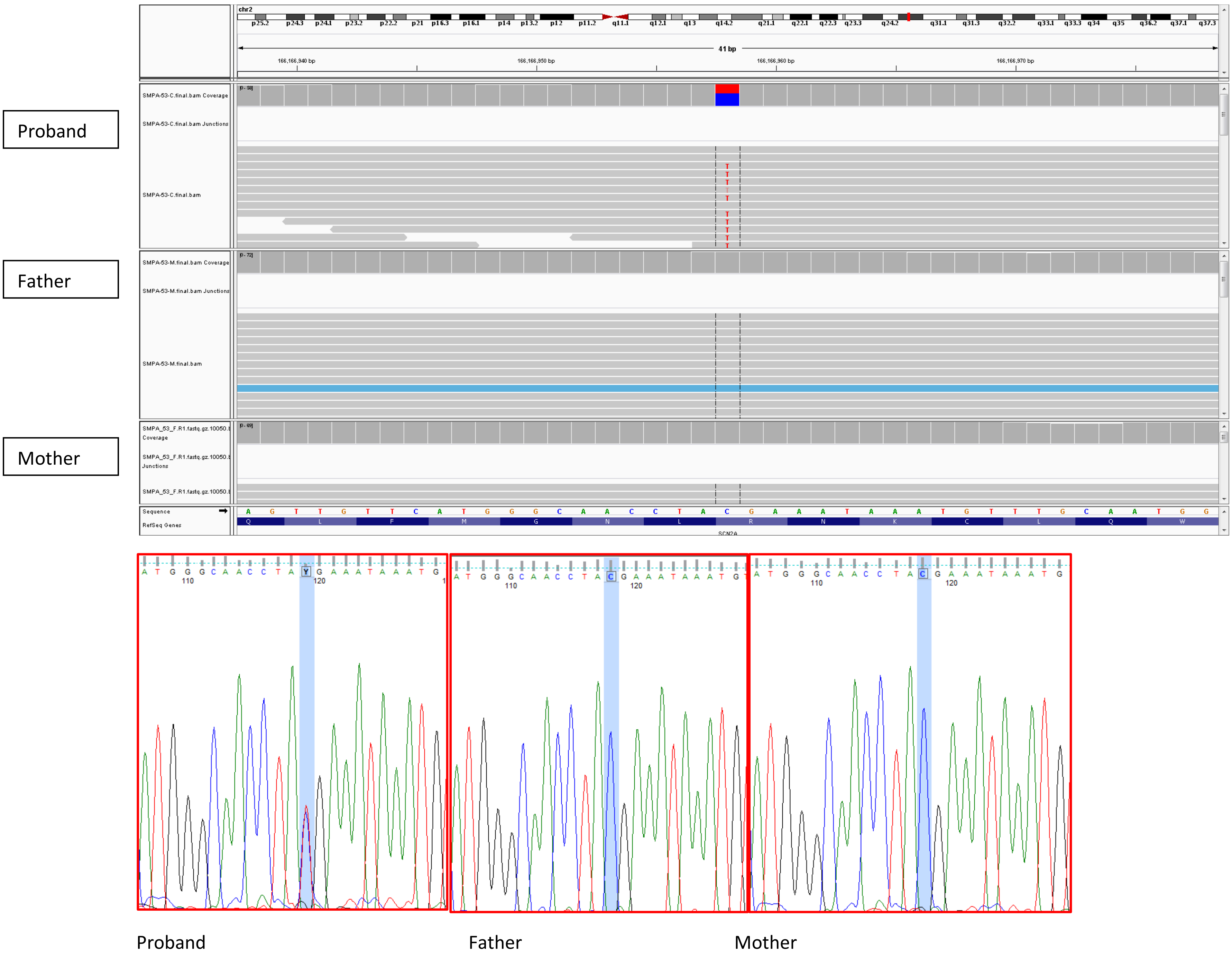

7. SMPA38: *SCN2A*

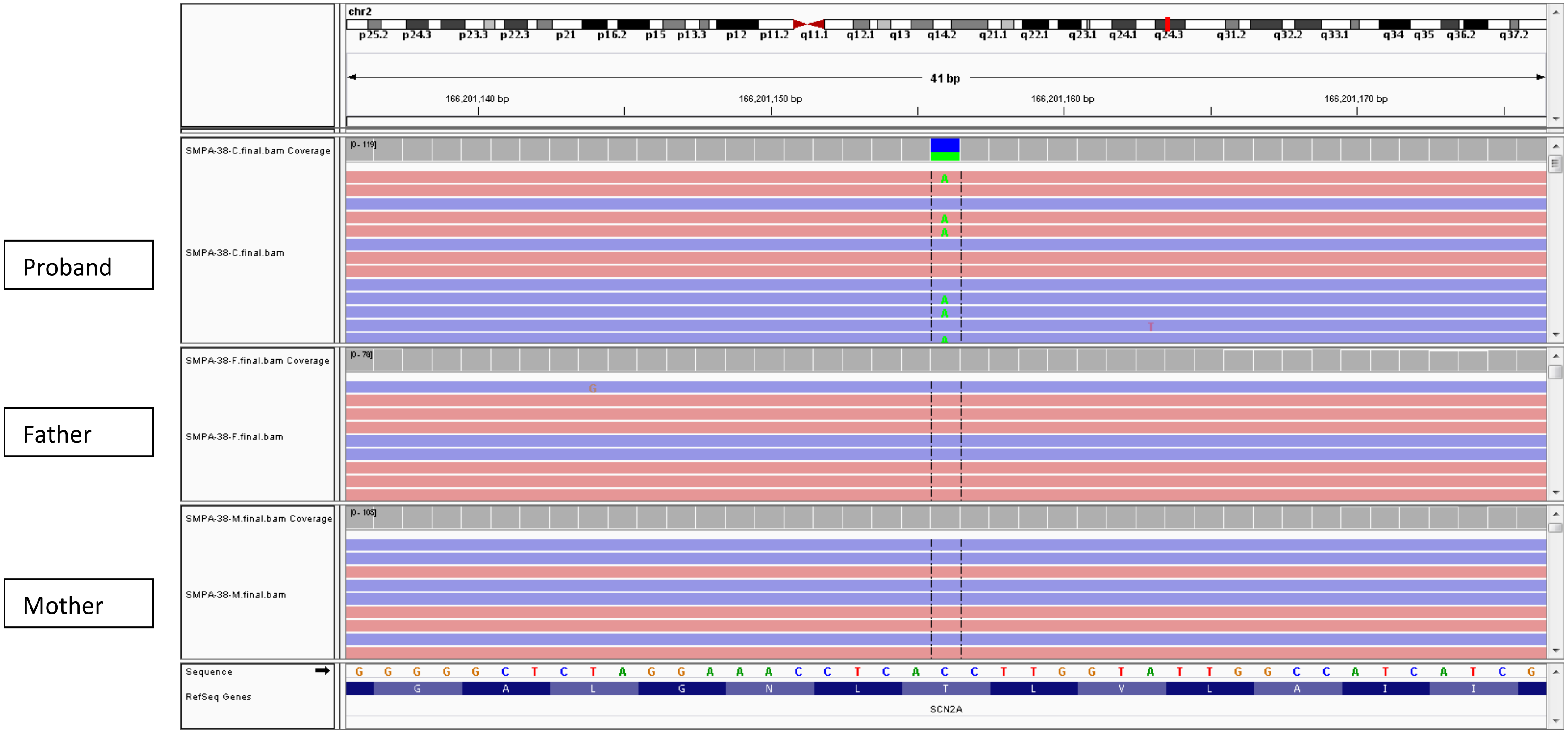


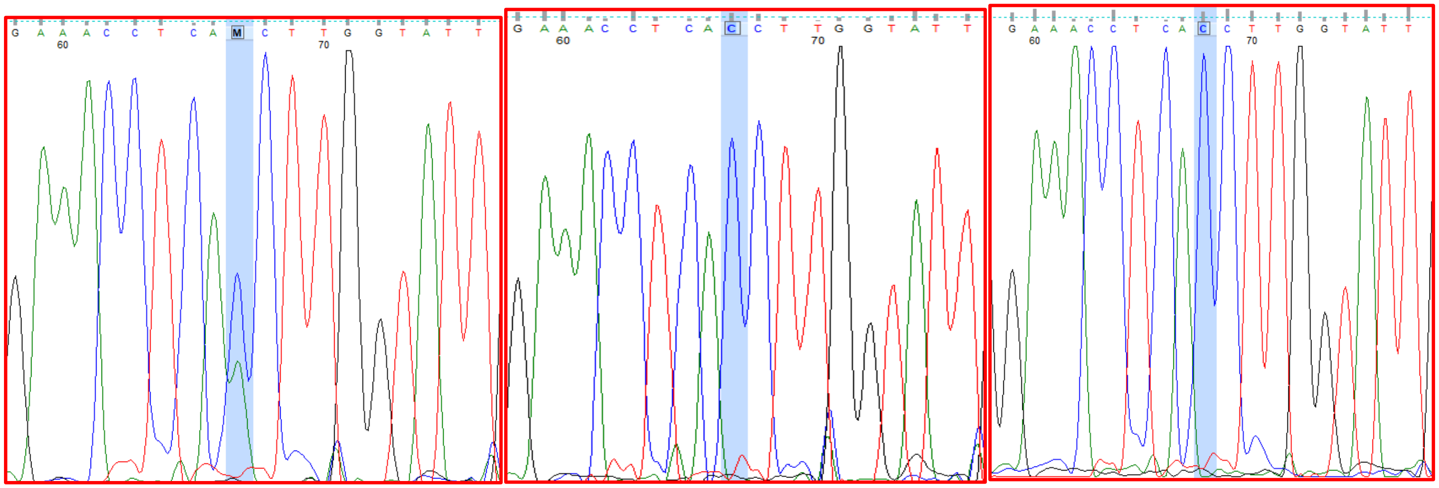

Proband Father Mother

8. IAU66: *NCL*

Proband

Father

Mother

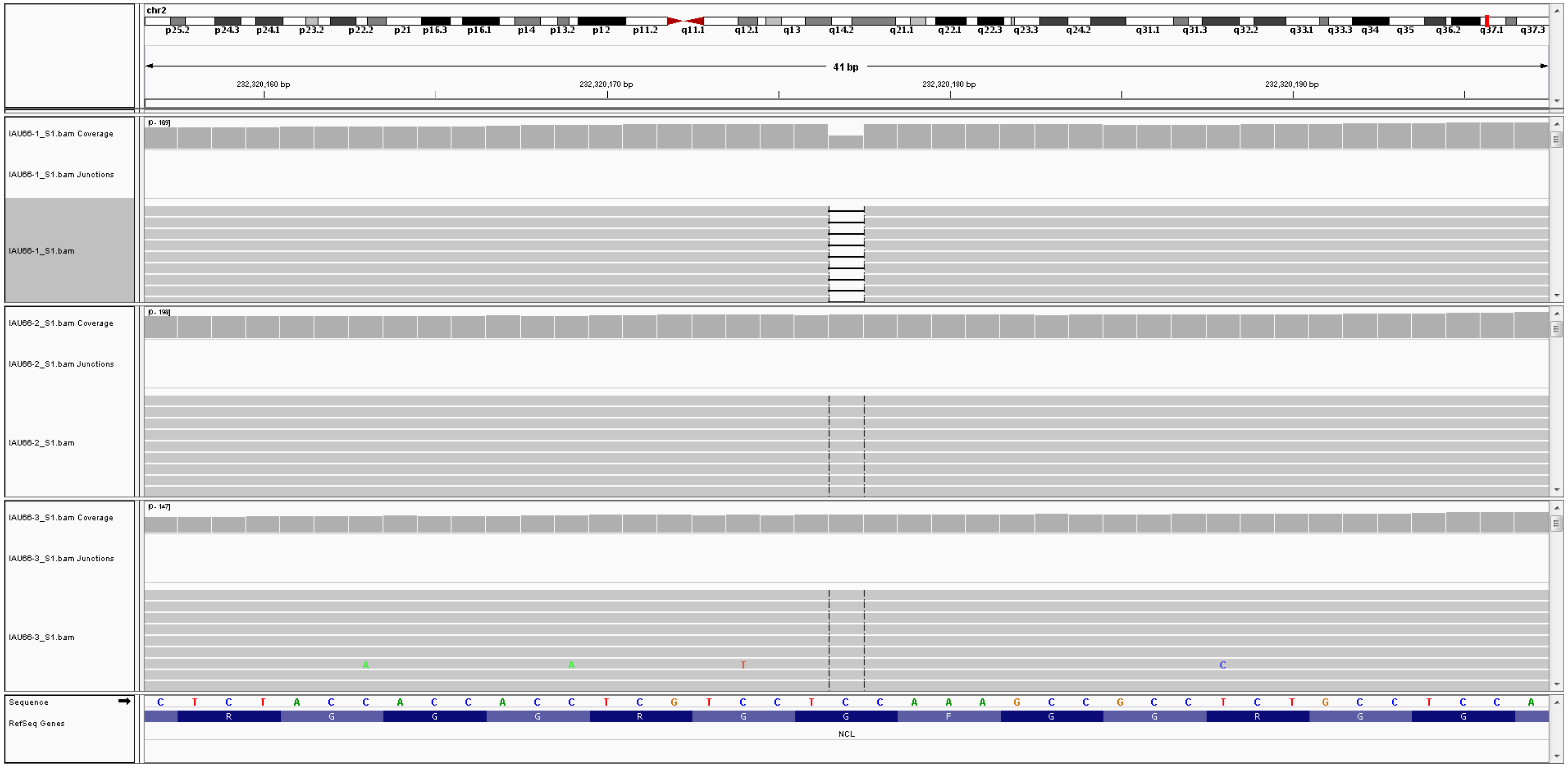

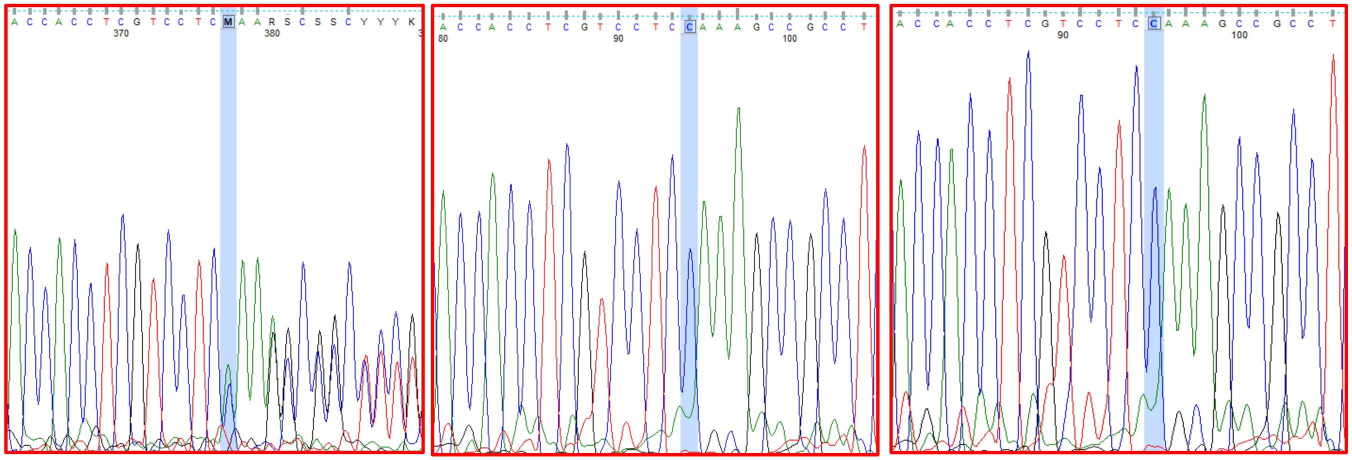

Proband Father Mother

9. SA1: SCN5A

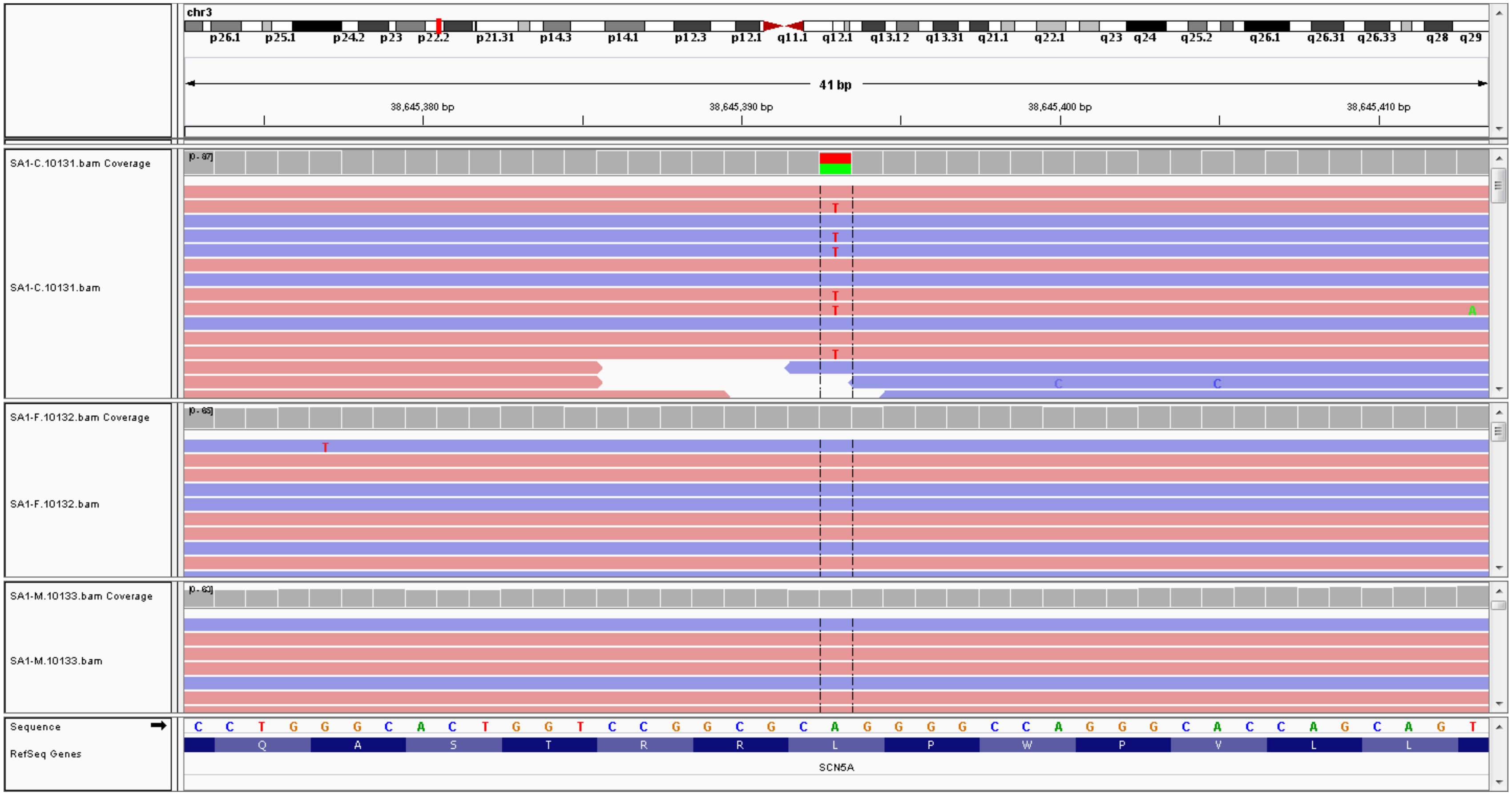


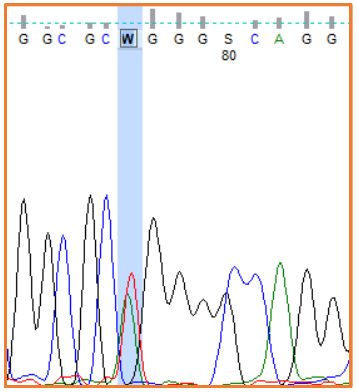

Proband

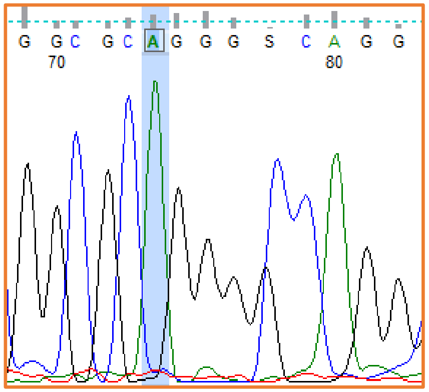

Sibling

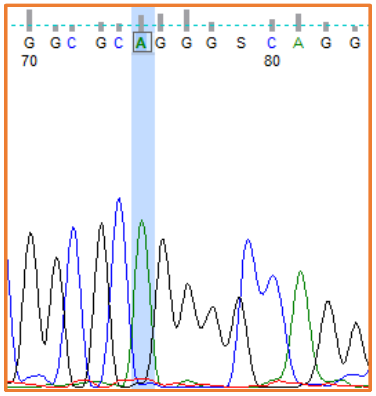

Father

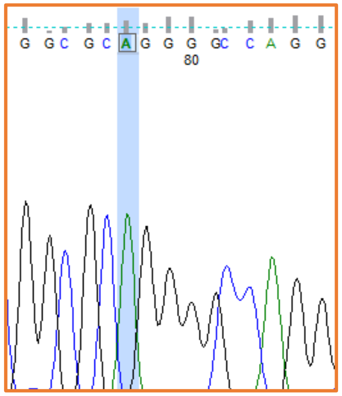

Mother

10. SMPA75: *FAM53C*

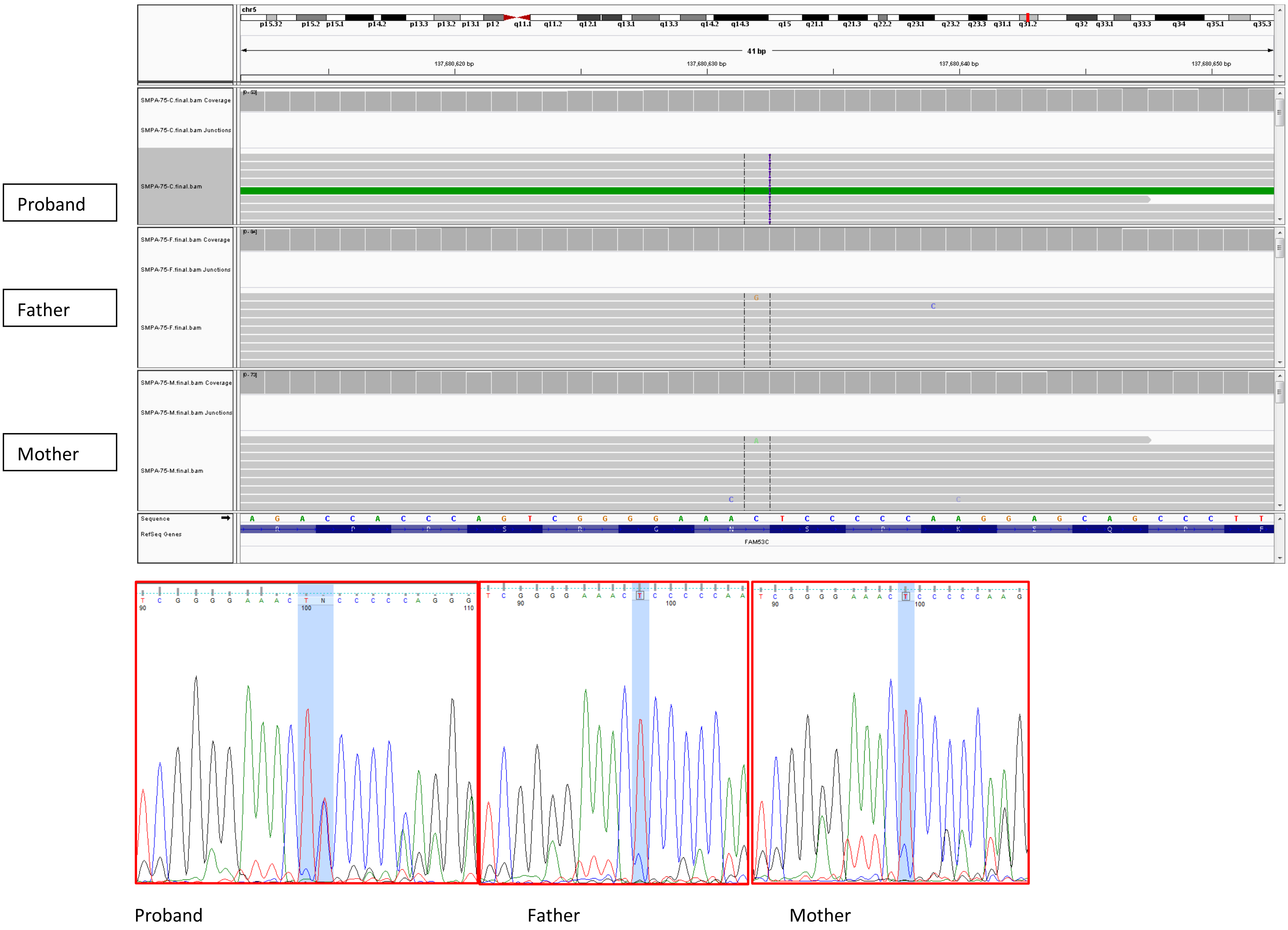

11. SMPA75: *ADGRF2*

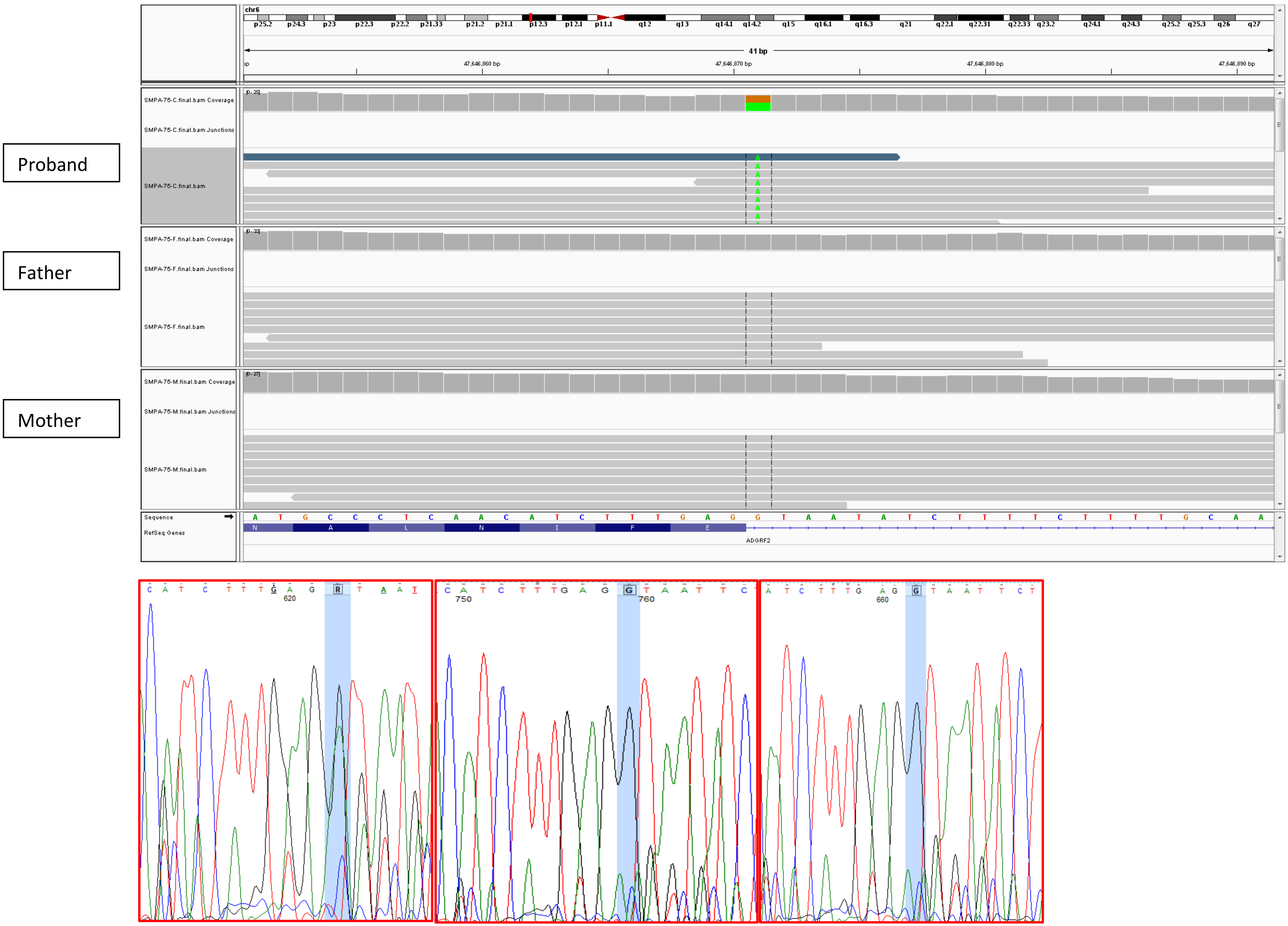


Proband Father Mother

12. Autism10: *ZNF292*

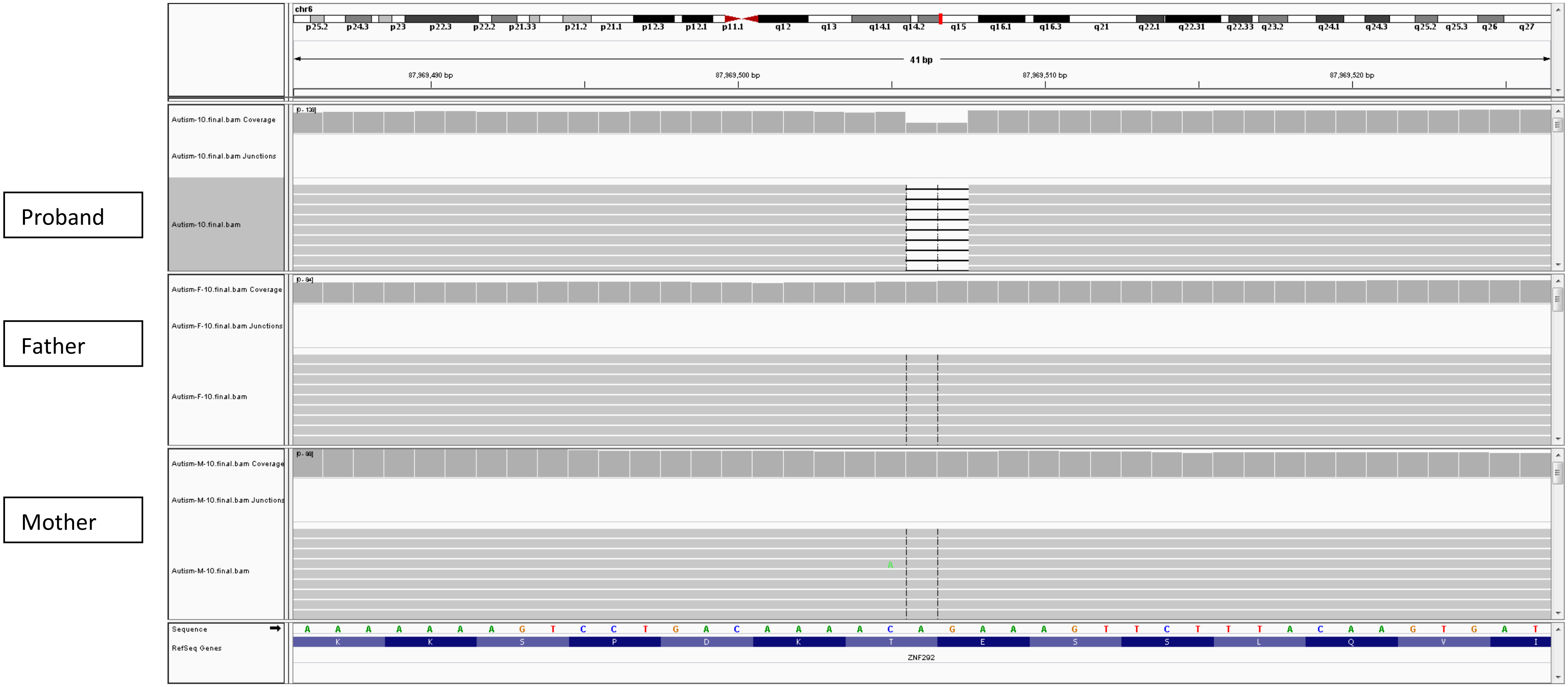


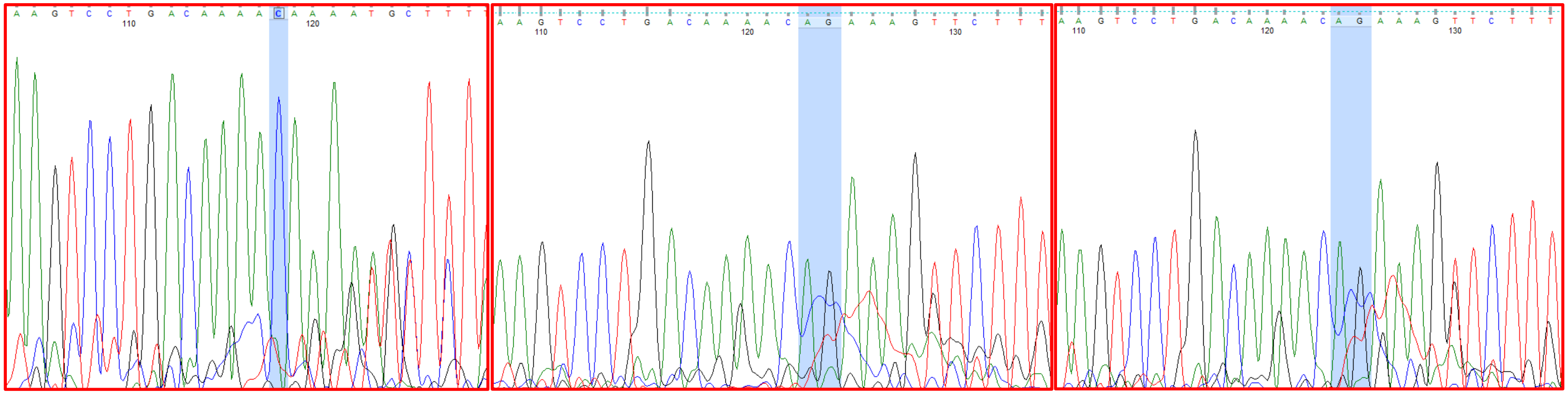

Proband Father Mother

13. IAU65: *ZNF292*

Proband

Father

Mother

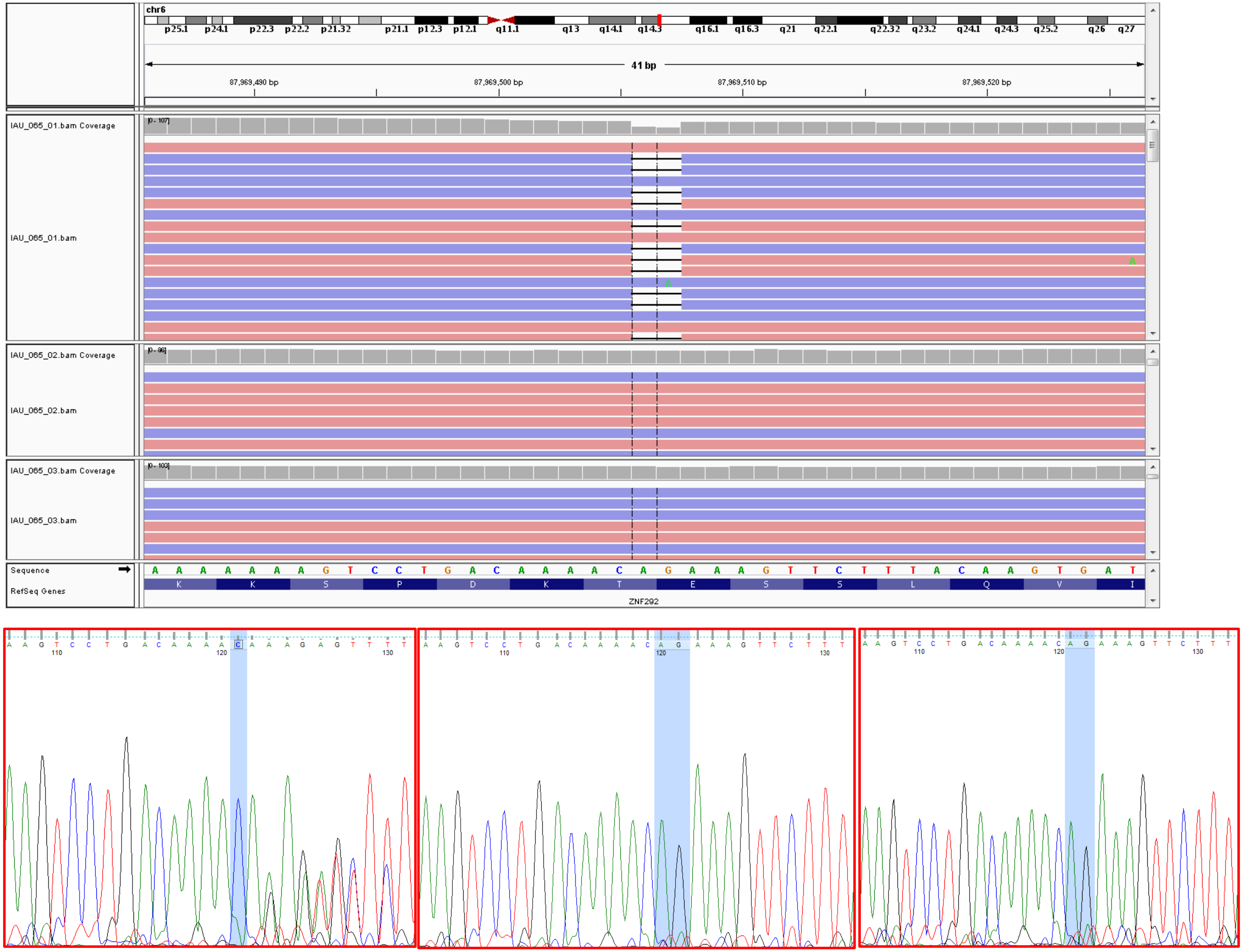


Proband Father Mother

14. RQPA20: *DGKZ*

Proband

Father

Mother

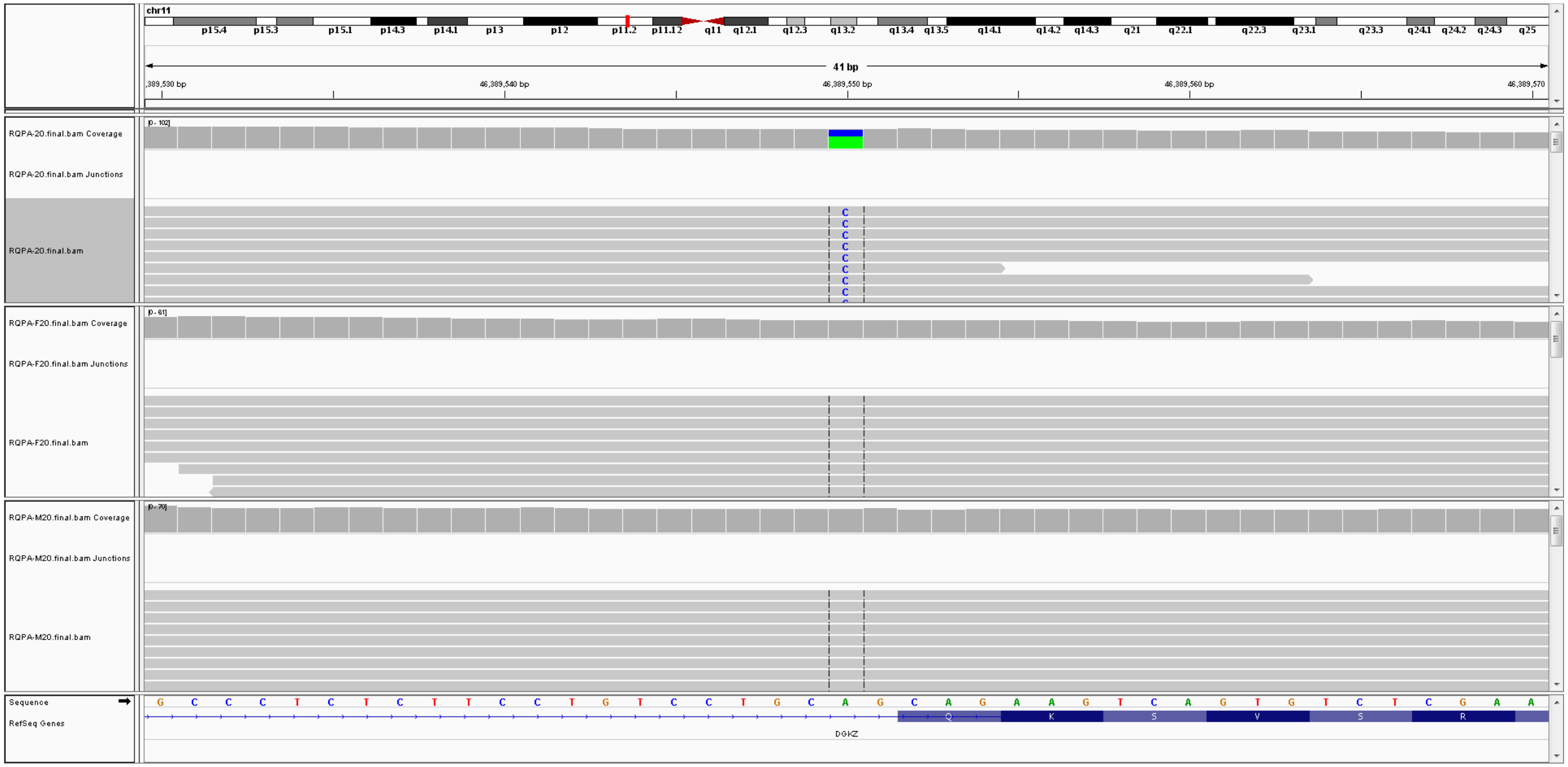


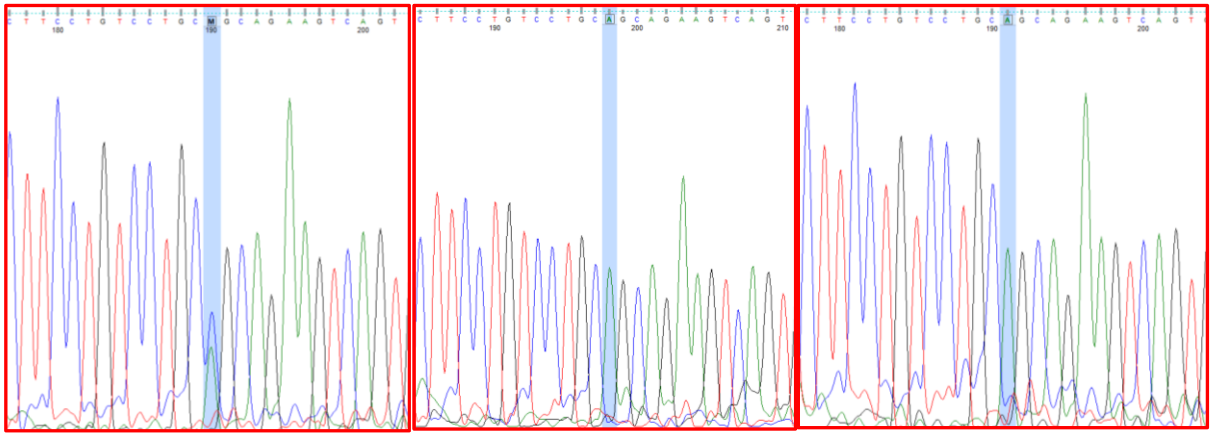

Proband Father Mother

15. Autism9: SCN8A

Proband

Father

Mother

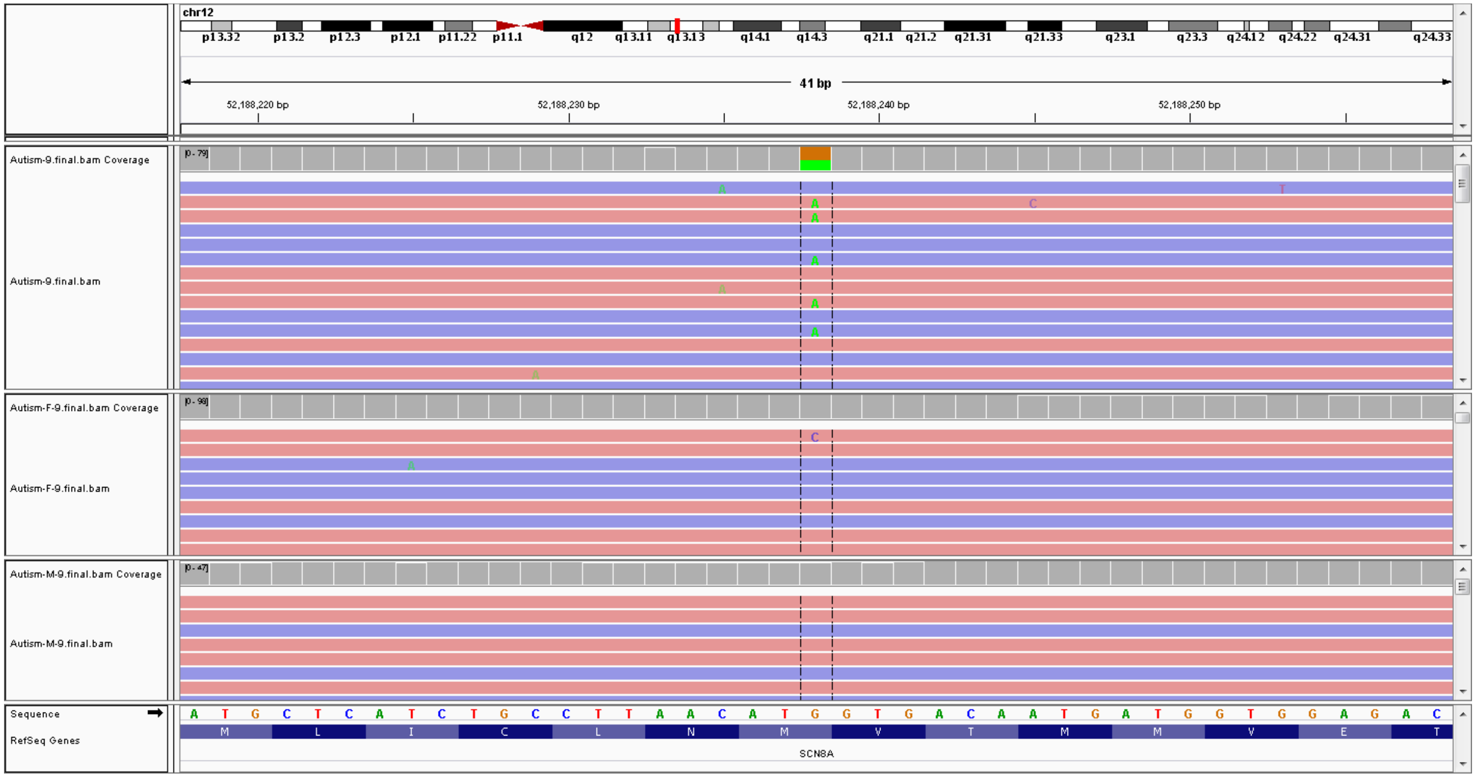


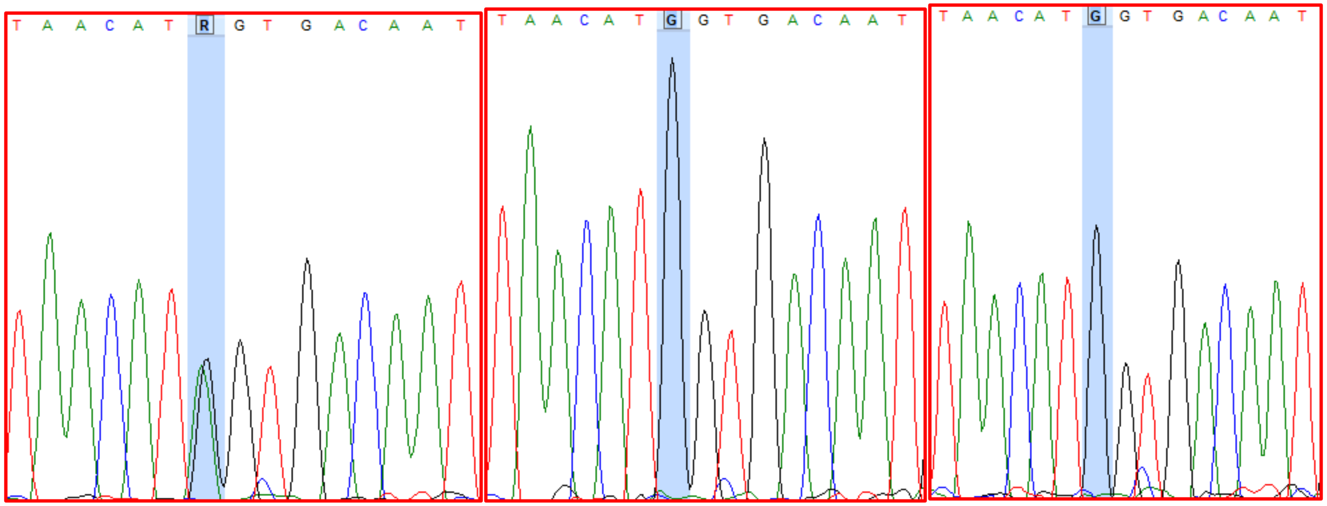


Proband

Father

Mother

16. SMPA73: *ATP2B1*

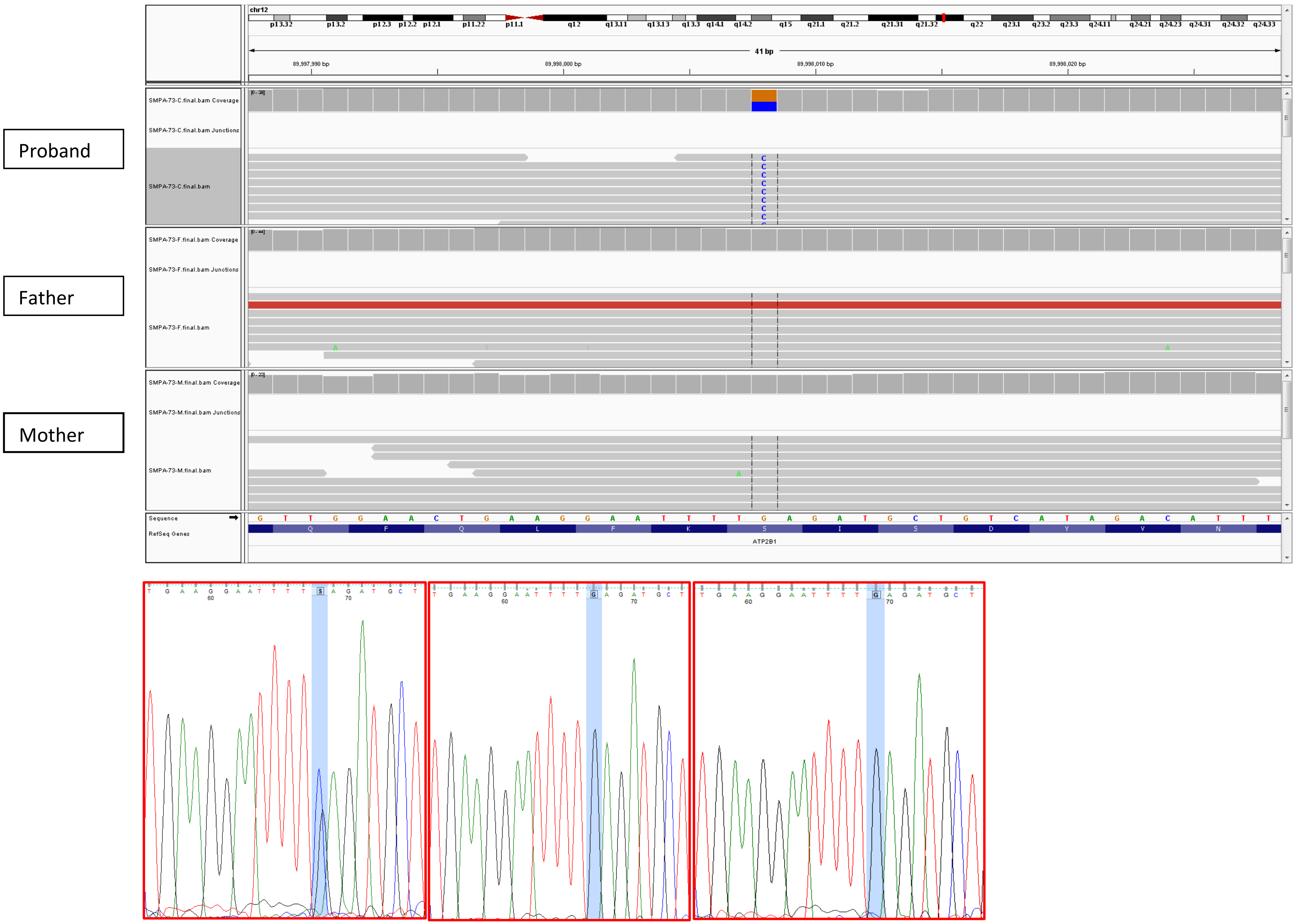


Proband Father Mother

17. RQPA20: *CBFA2T3*

Proband

Father

Mother

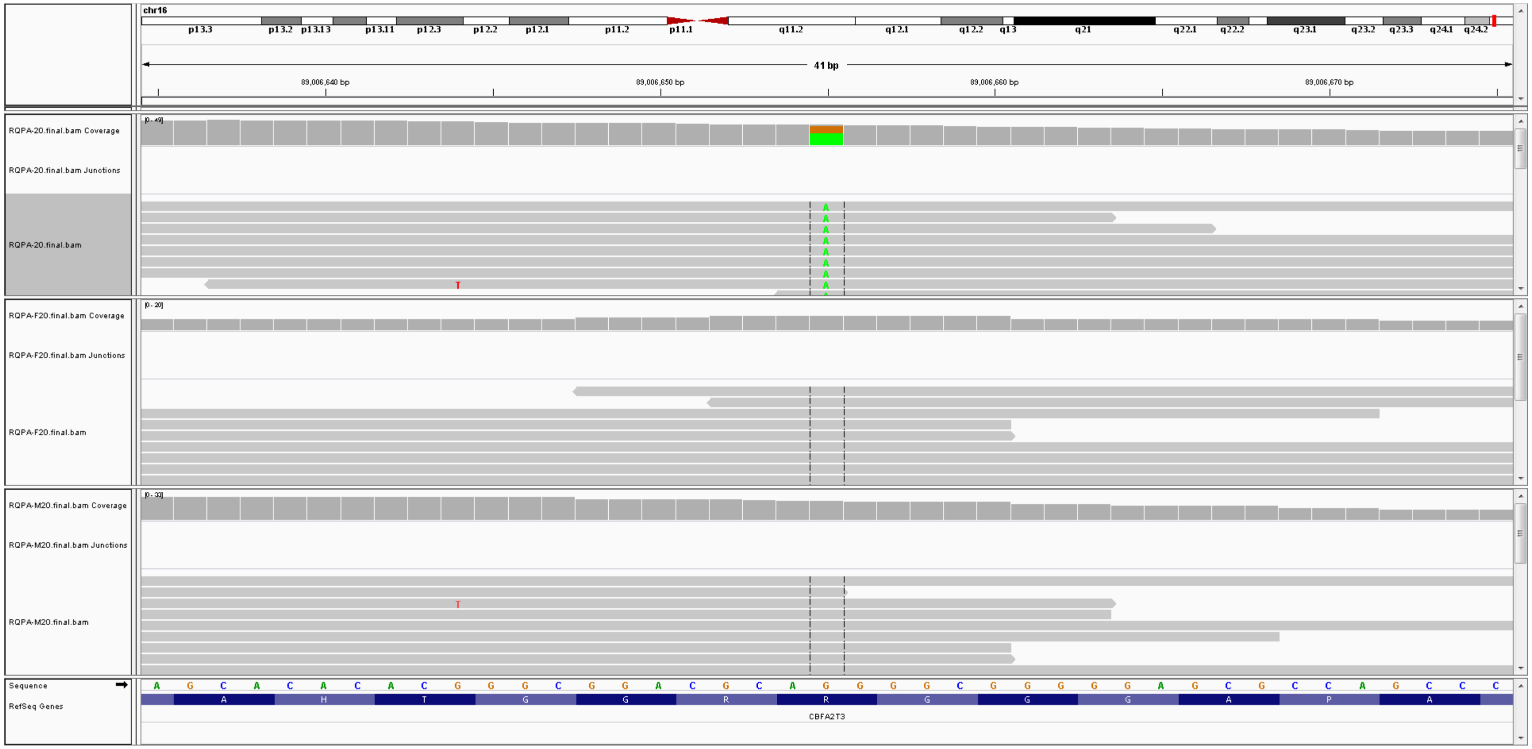


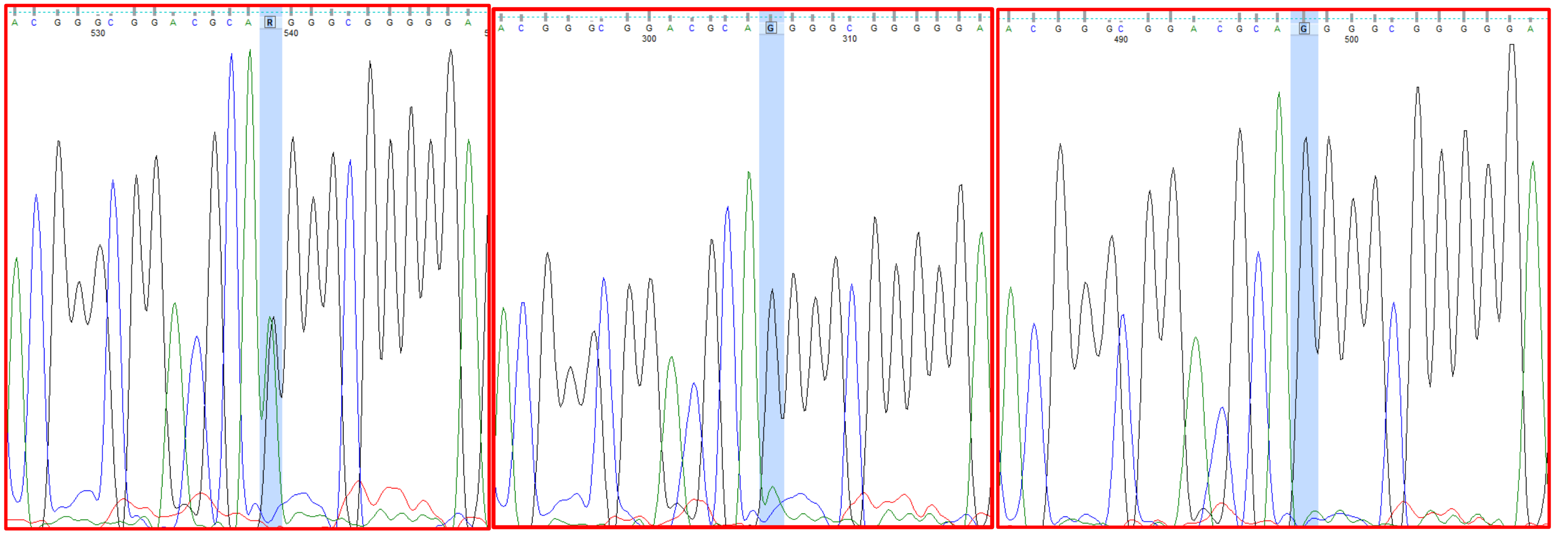

Proband Father Mother

18. SMPA75: *RETN*

Proband

Father

Mother

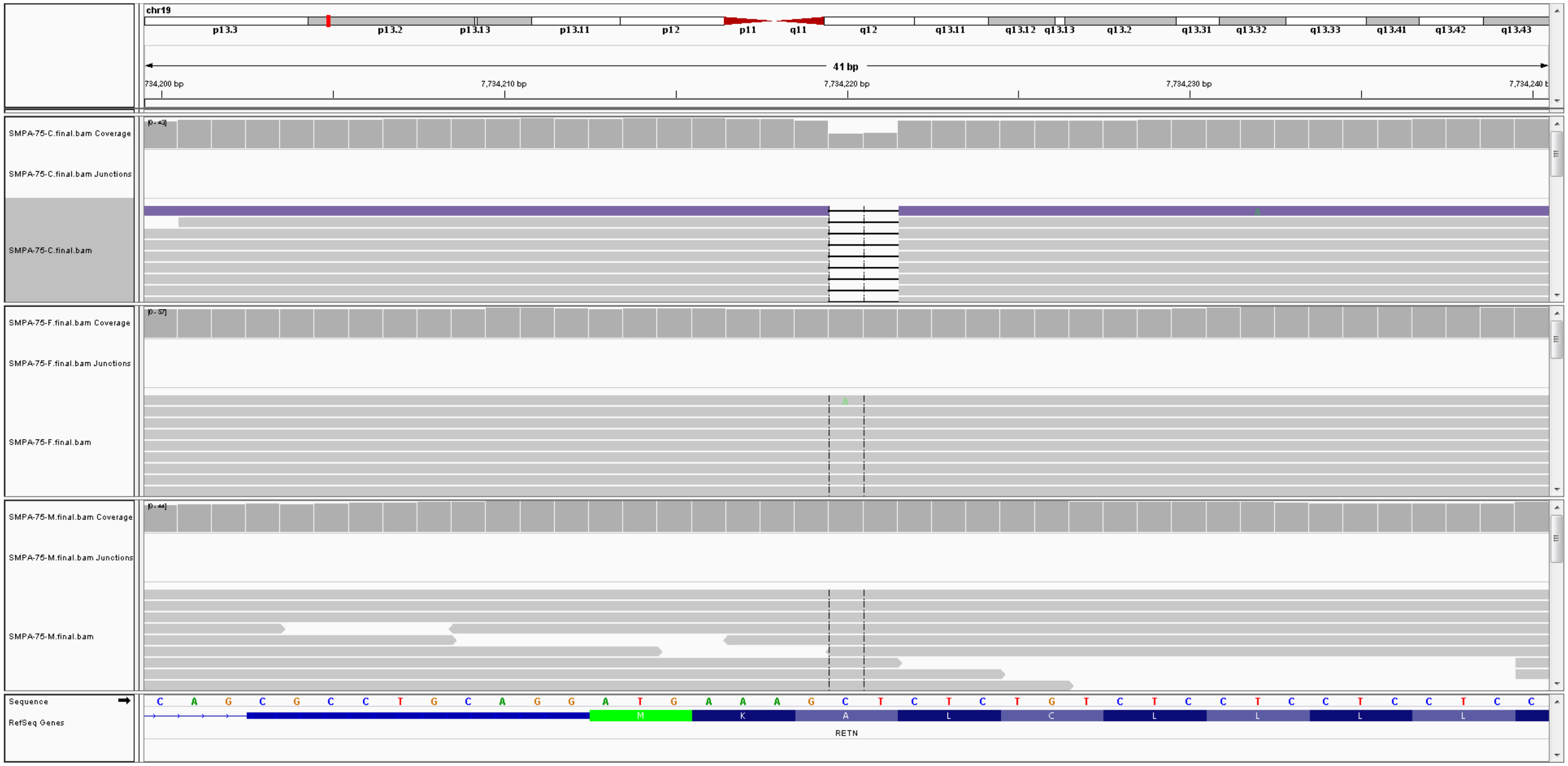

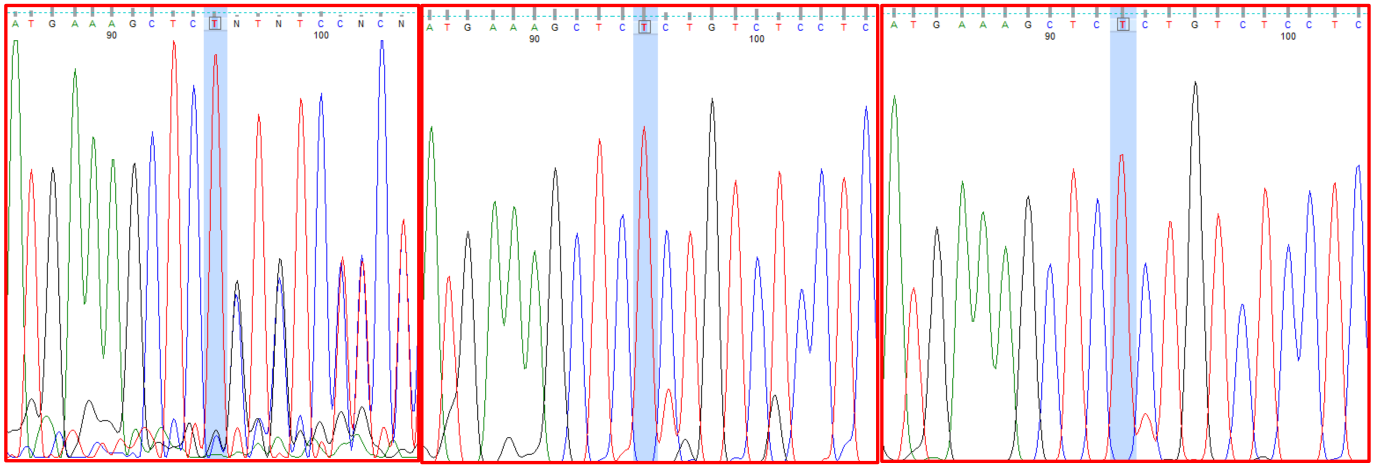

Proband

Father

Mother

19. RQPA20: TANGO2

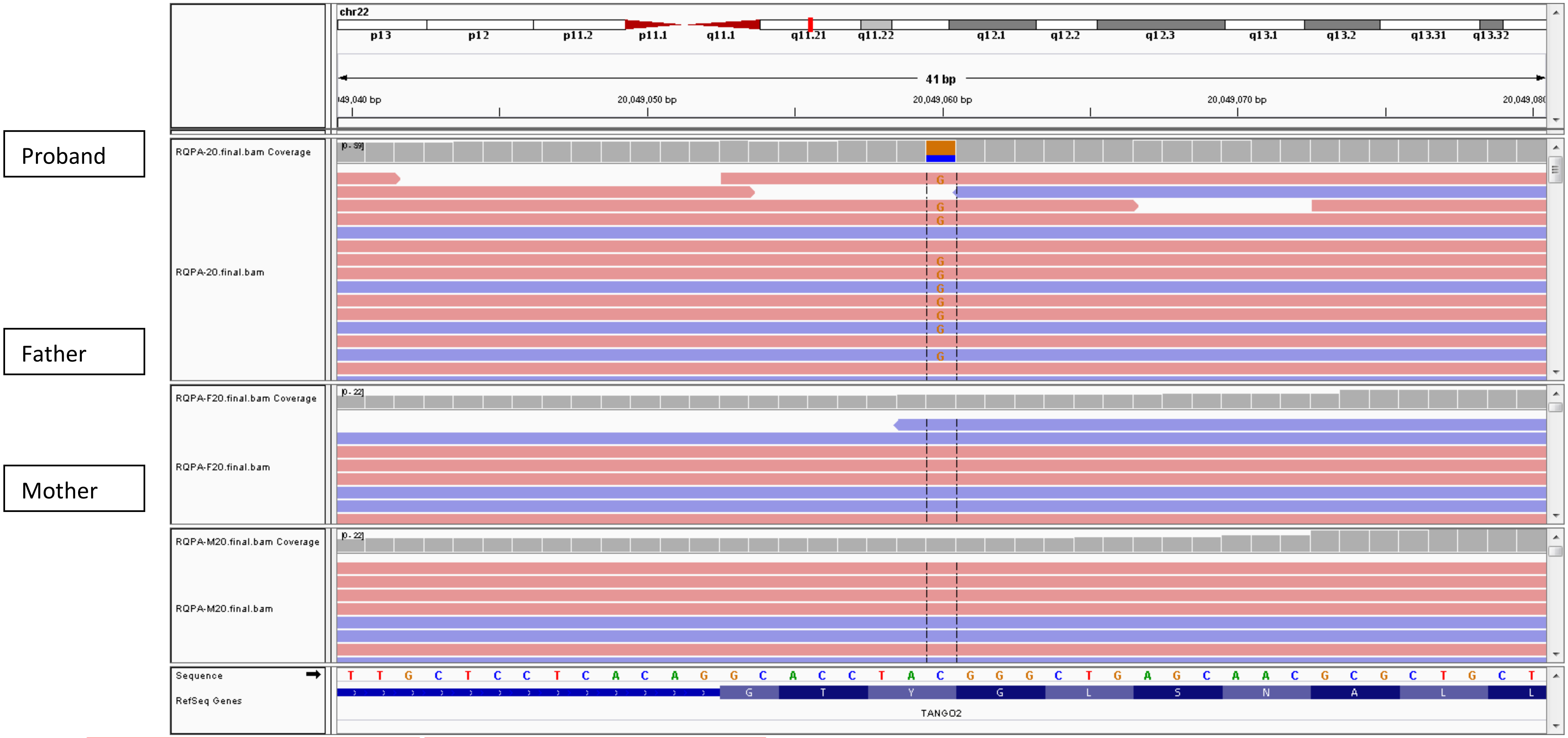


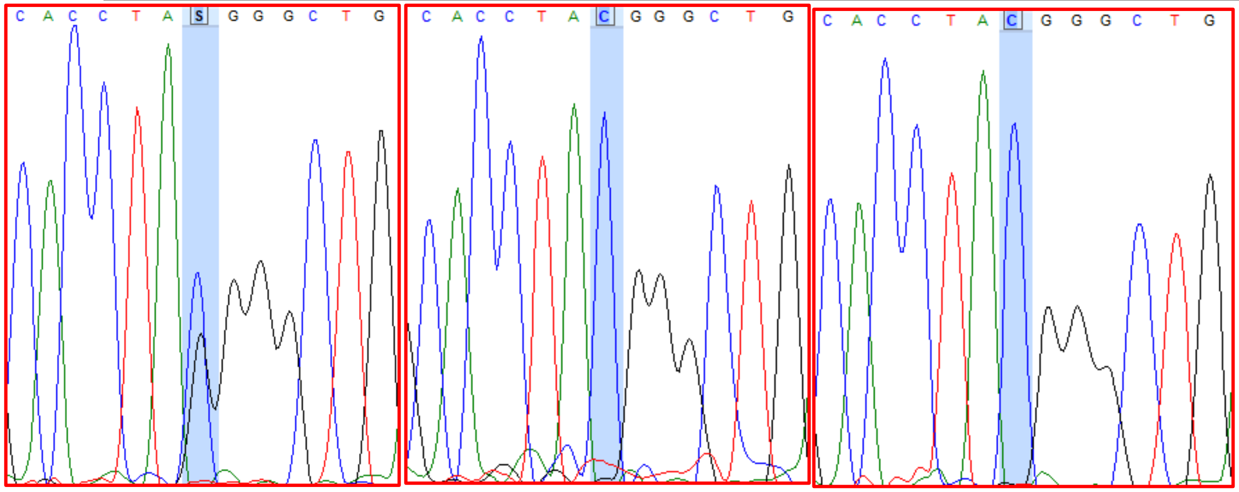


Proband Father Mother

20. RQPA20: *PPIL2*

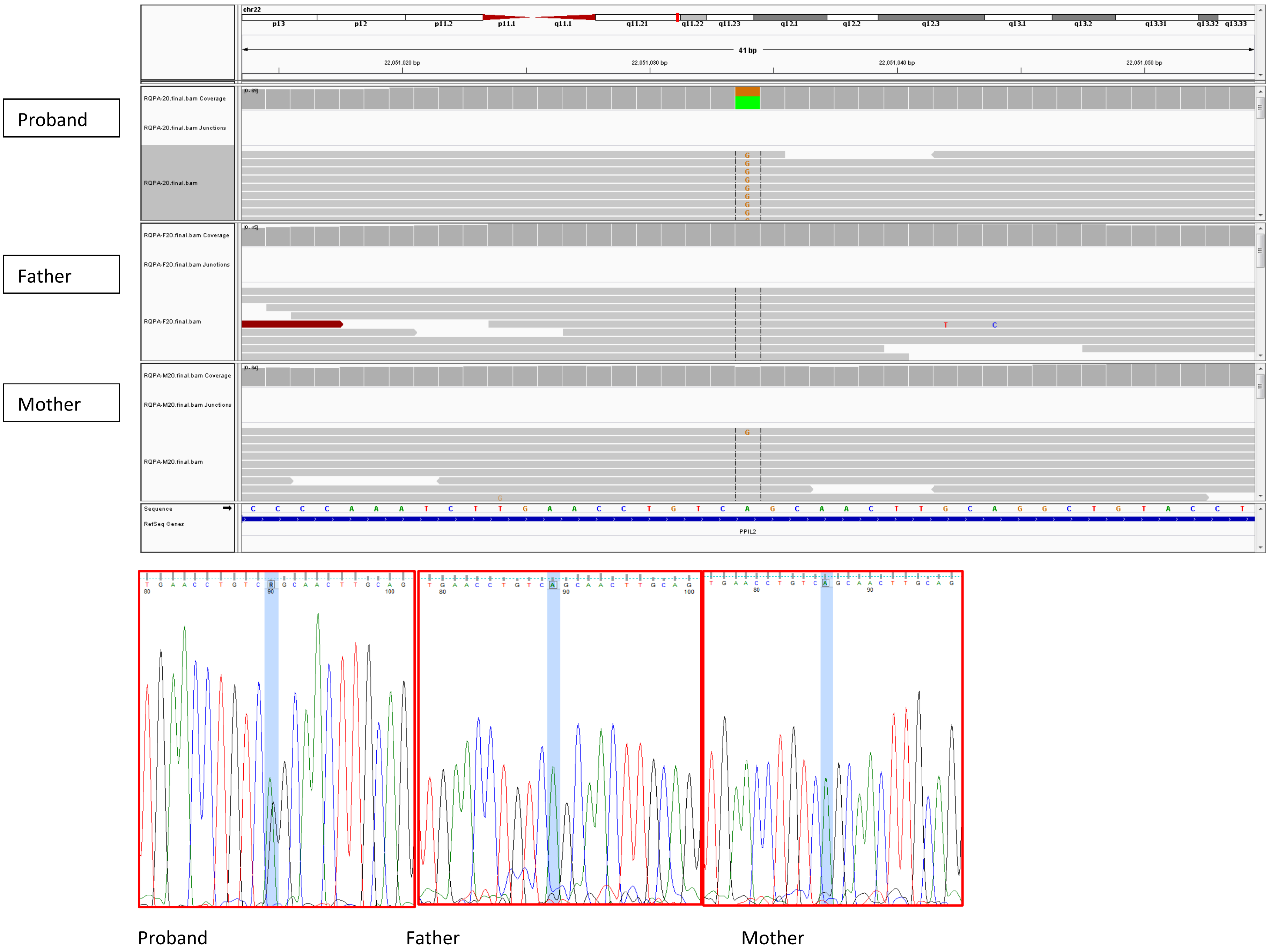

**Table 4: X-linked variants**

1. SMPA3: *MAGEB2*

Proband

Father

Mother

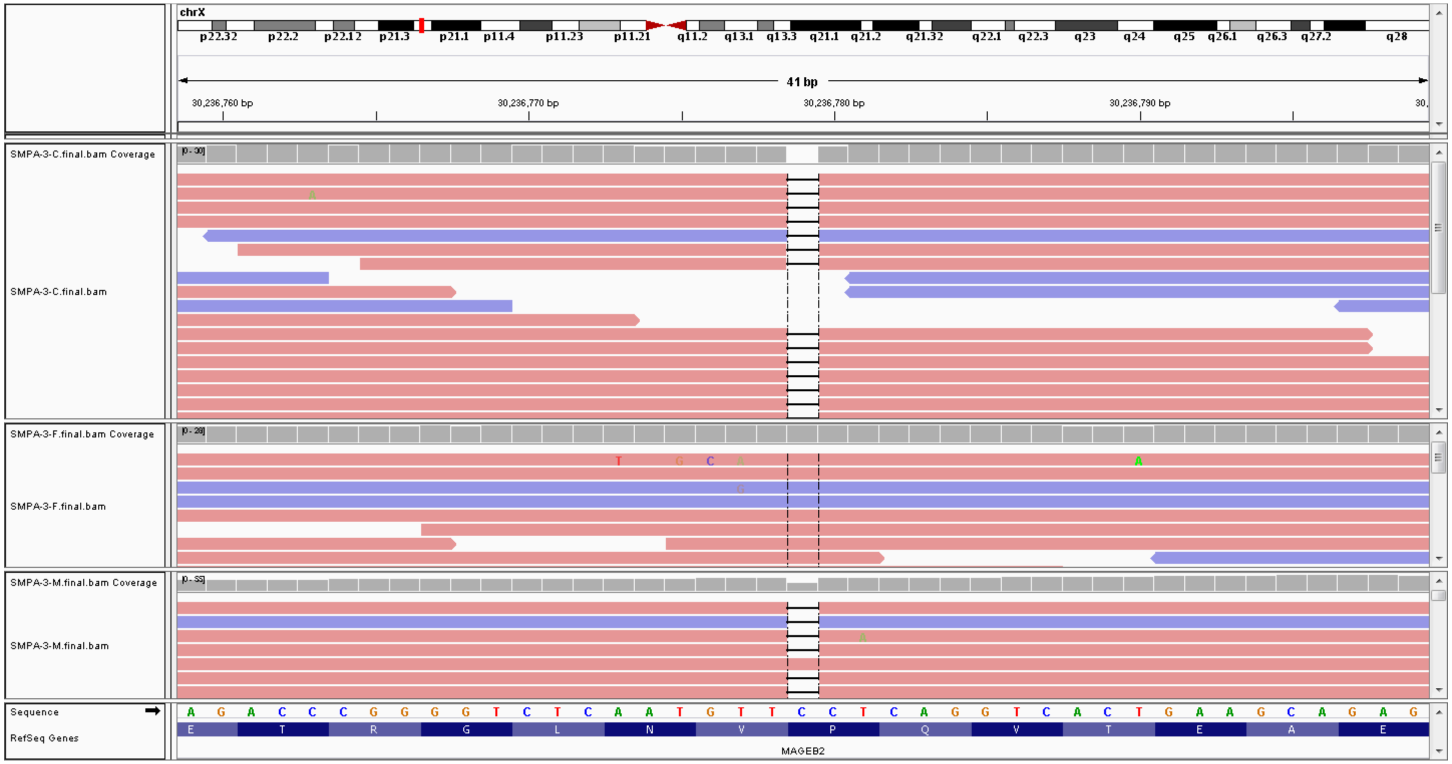


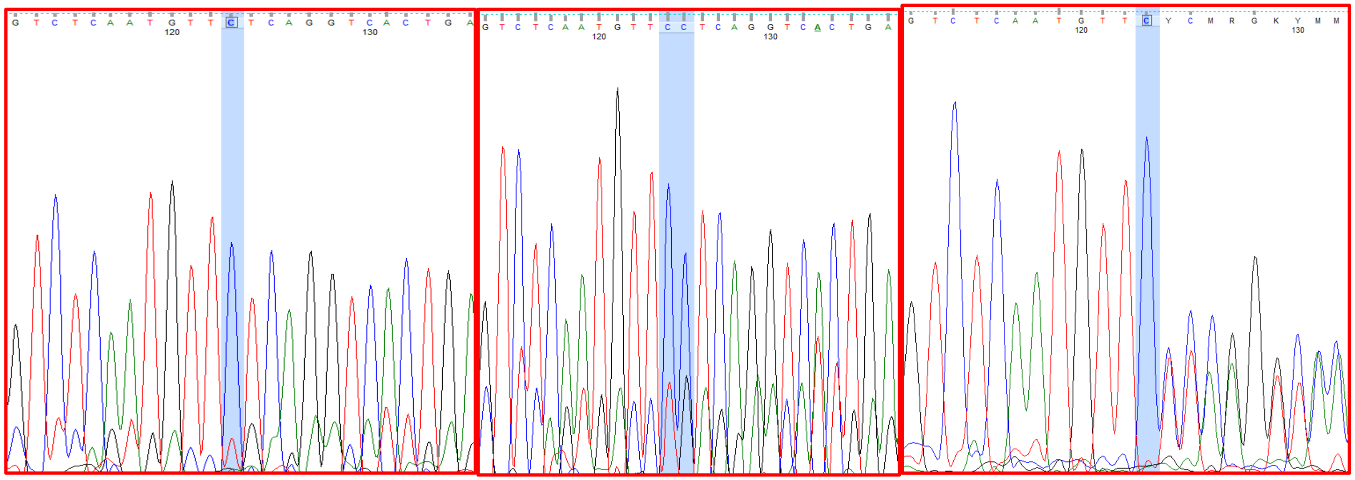

Proband Father Mother

2. IAU66: HDAC6

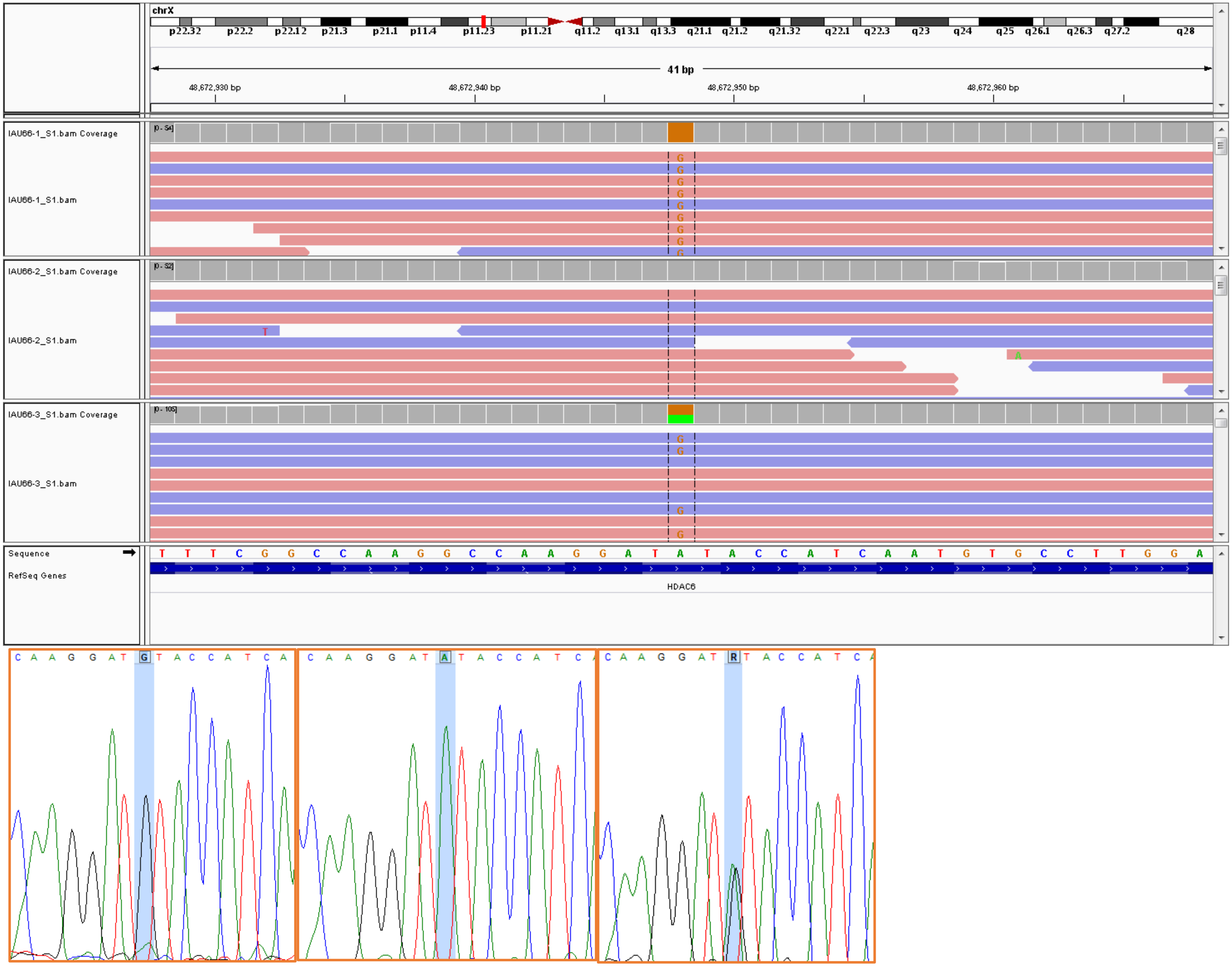


Proband

Father

Mother

3. IAU69: *RRAGB*

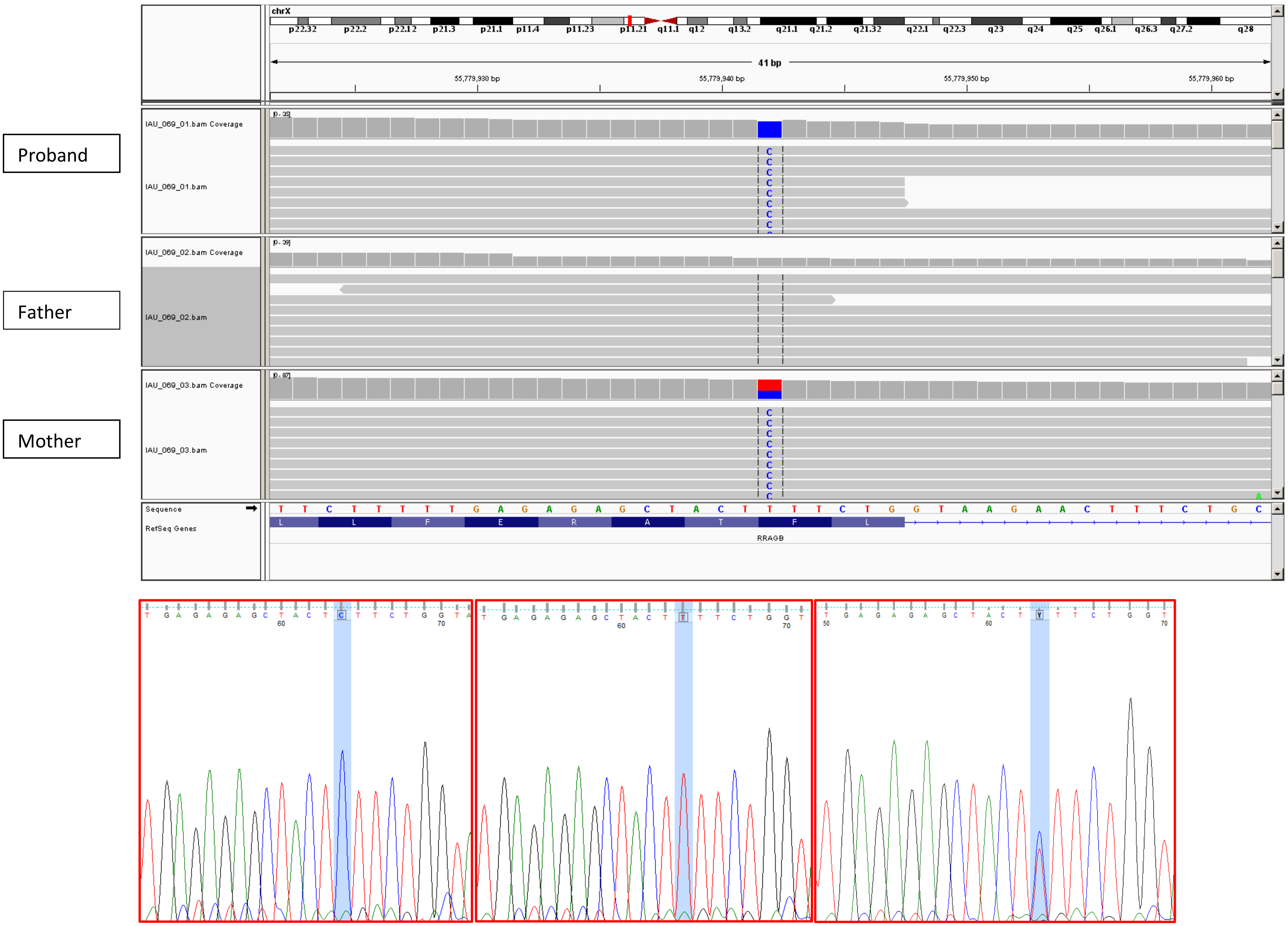


Proband Father Mother

4. IAU45: *MSN*

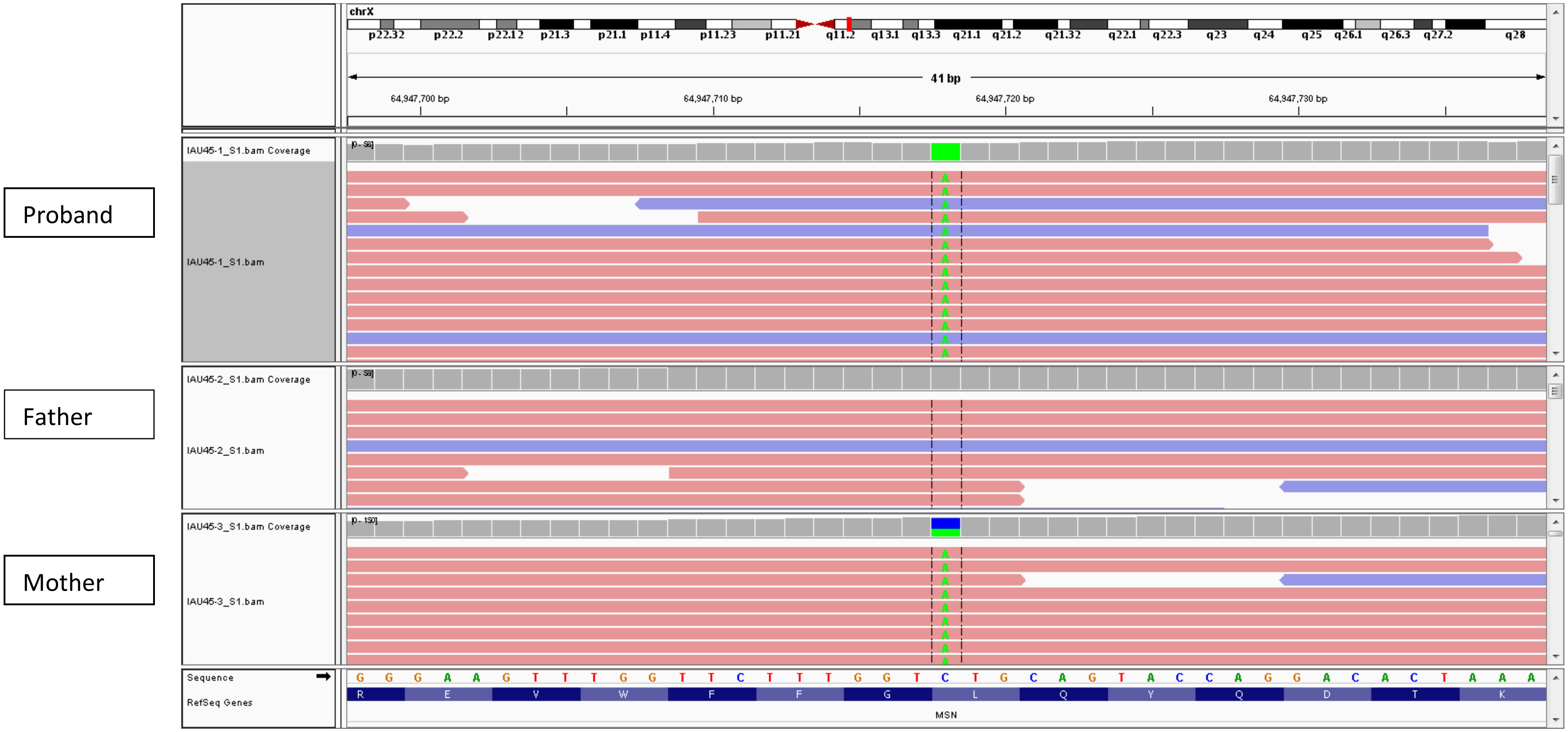


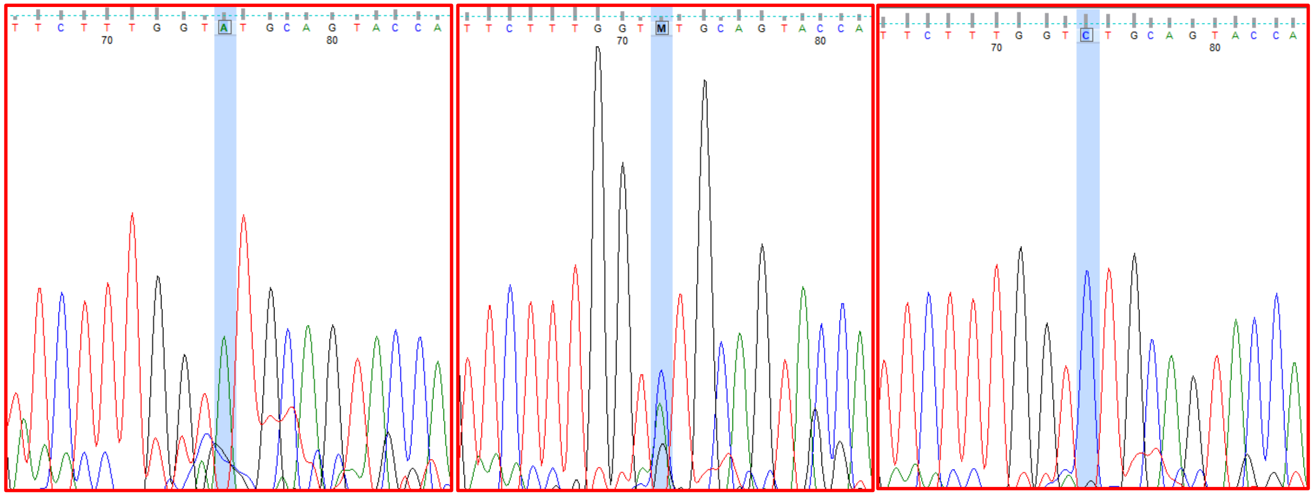

Proband Father Mother

5. SMPA15: *NRK*

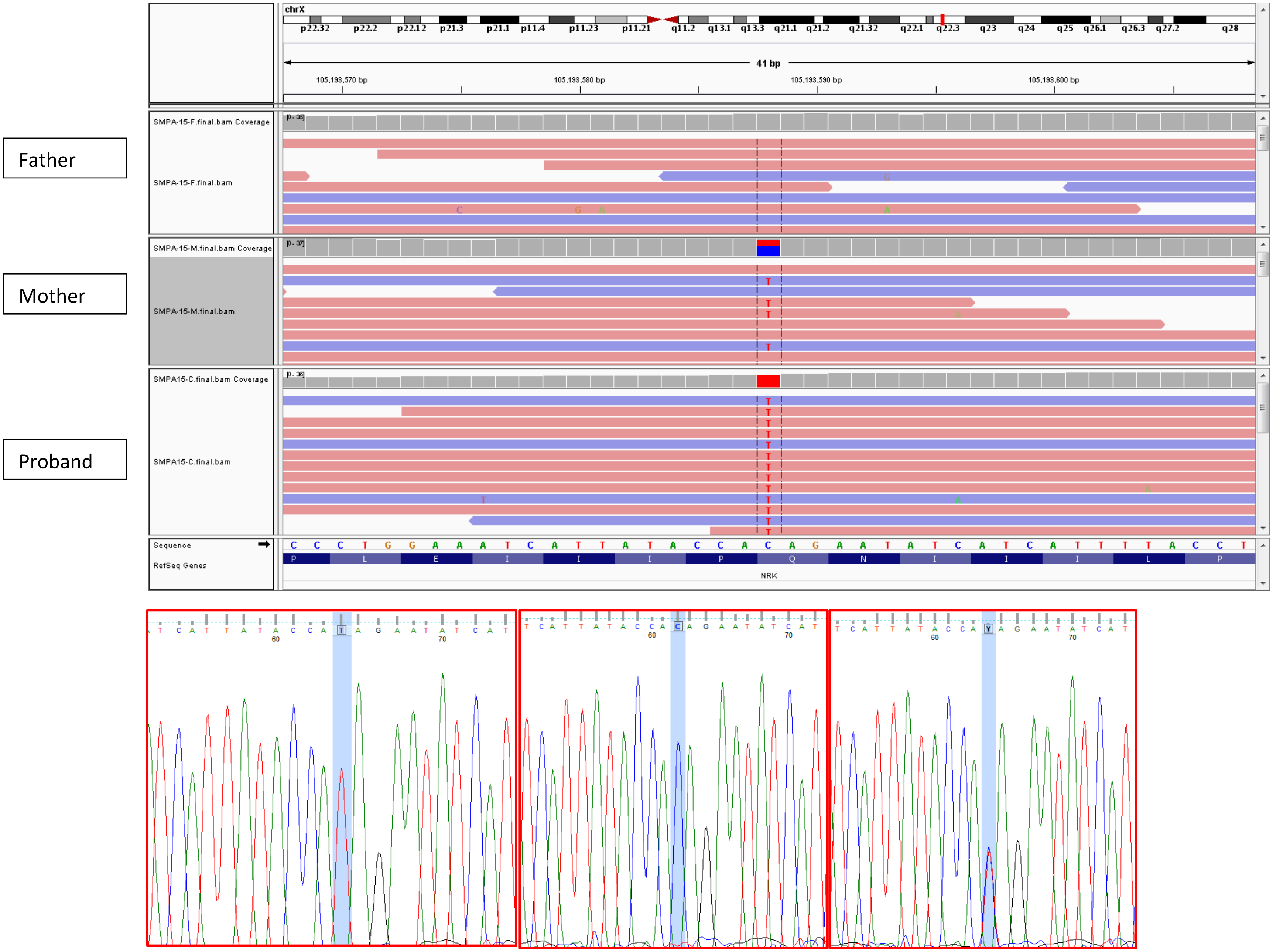


Proband Father Mother

6. IAU47: *MID2*

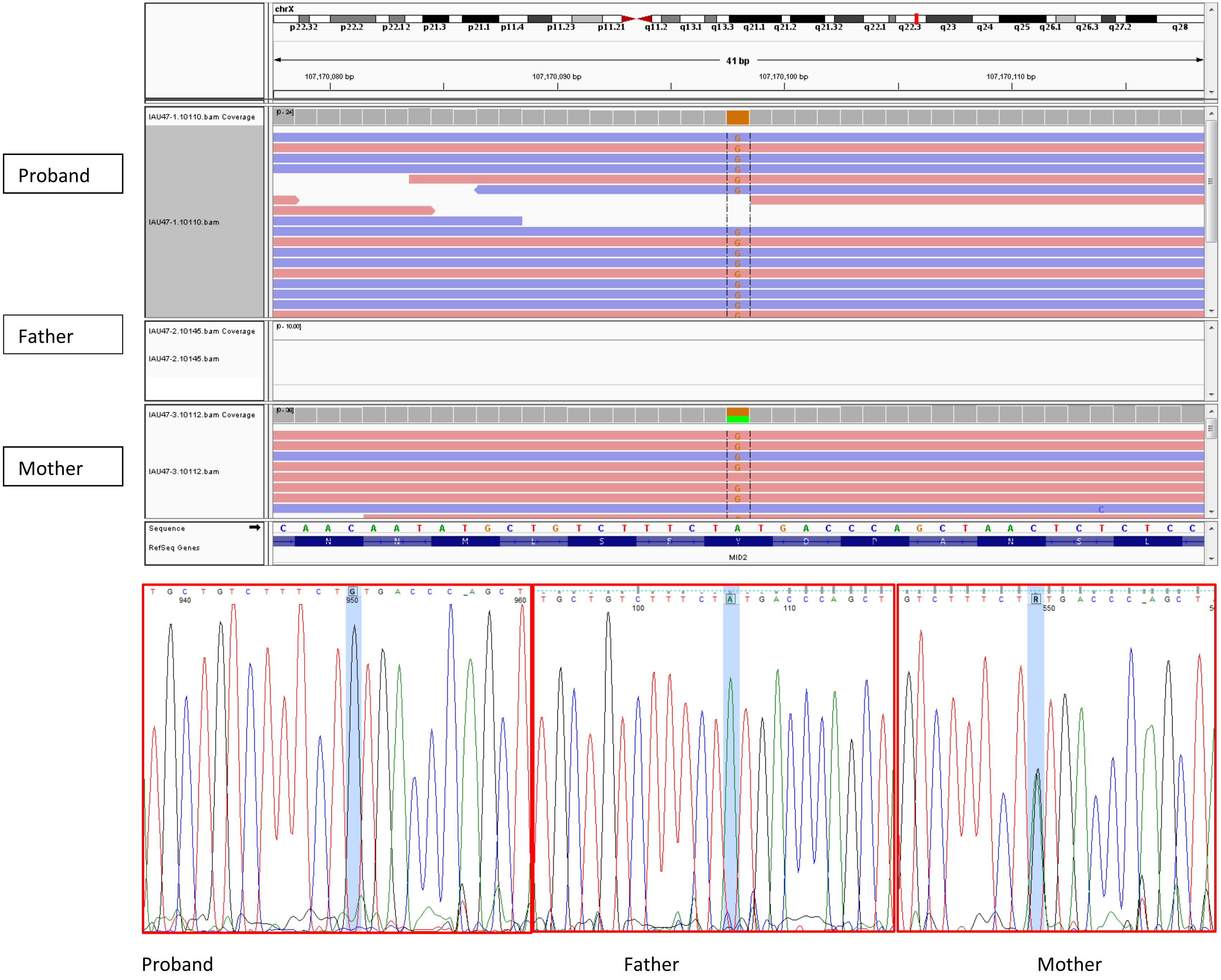
Proband
Father
Mother
chrX
41 bp
107,170,080 bp
107,170,090 bp
107,170,100 bp
107,170,110 bp
IAU47-1.10110.bam Coverage
IAU47-1.10110.bam
IAU47-2.10145.bam Coverage
IAU47-2.10145.bam
IAU47-3.10112.bam Coverage
IAU47-3.10112.bam
Sequence
RefSeq Genes
MID2
Proband
Father
Mother

7. IAU24: *AIFM1*

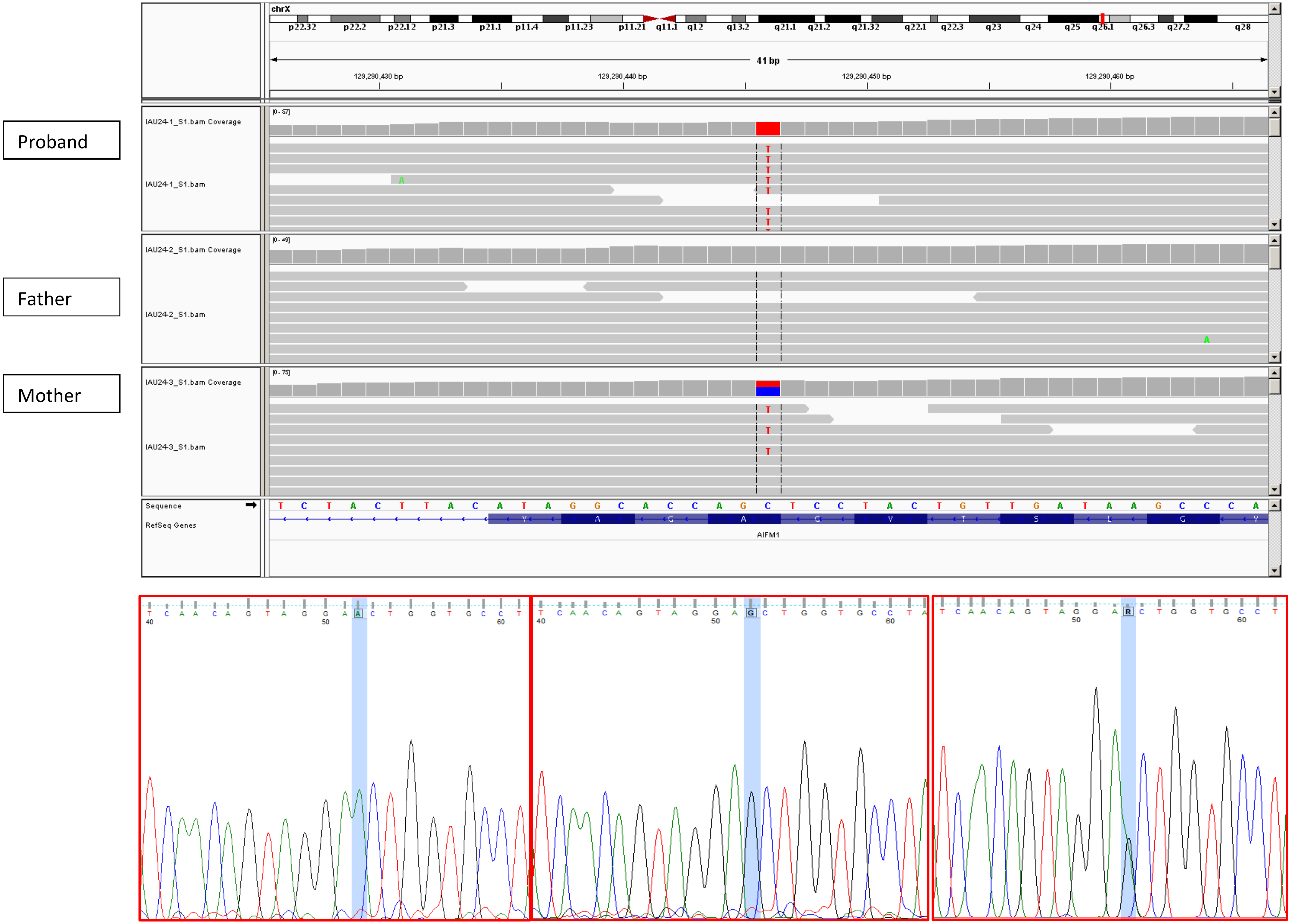


Proband Father Mother

*8.* IAU51: *MAGEC1*

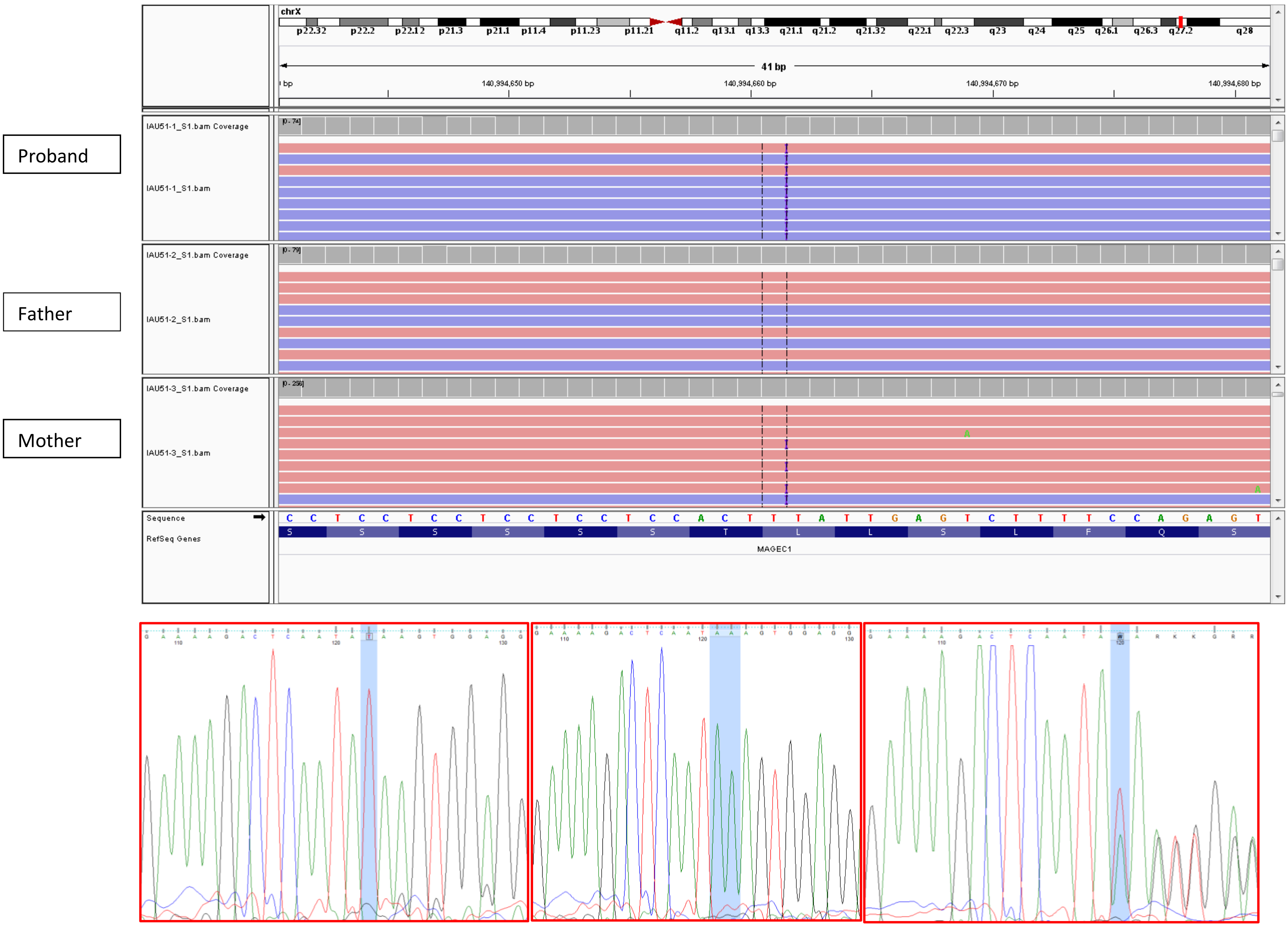


Proband Father Mother

9. IABB8: ZNF185

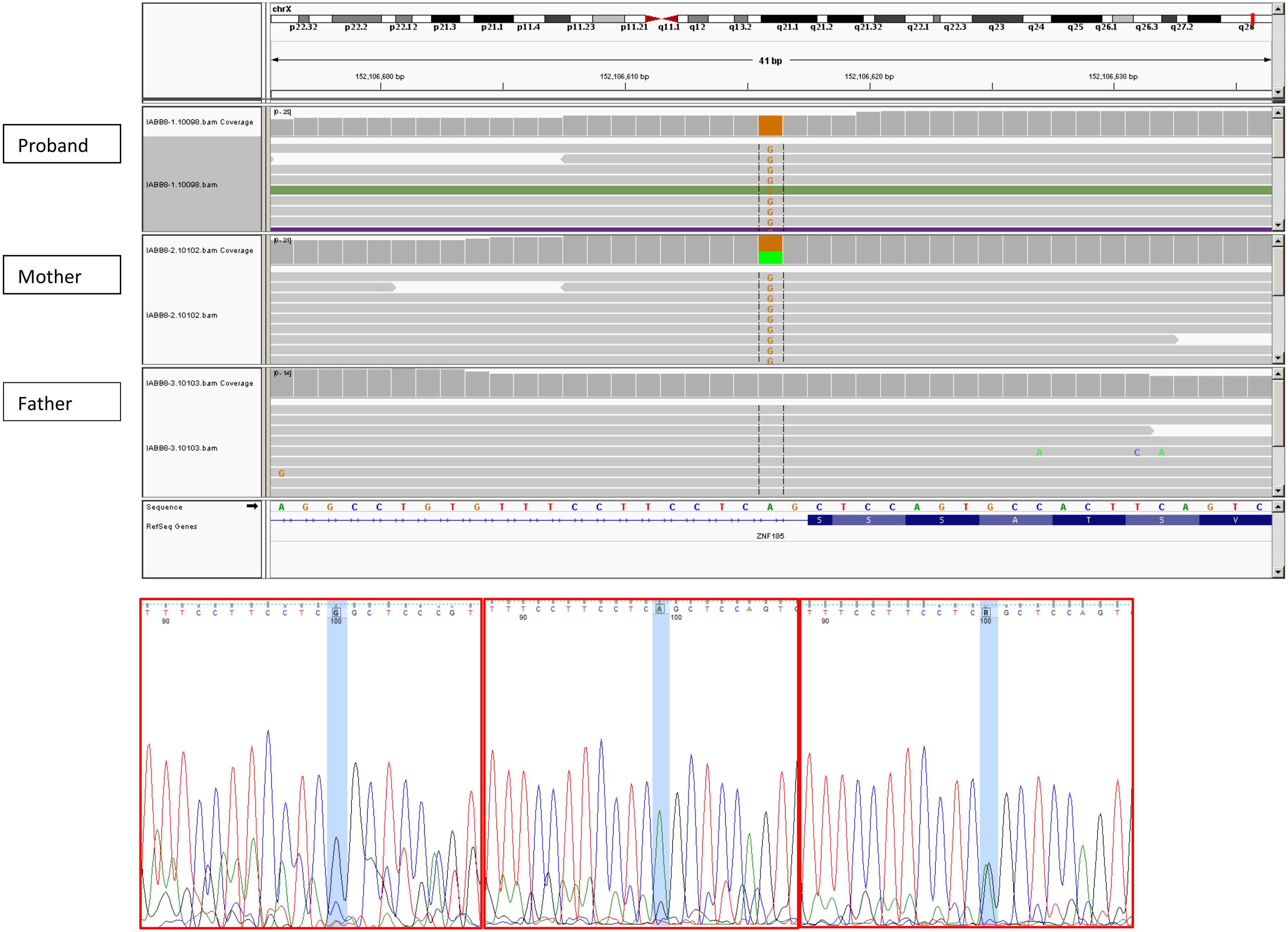


Proband Father Mother

10. SA1: *CLTRN* (*TMEM27*)- *de novo*

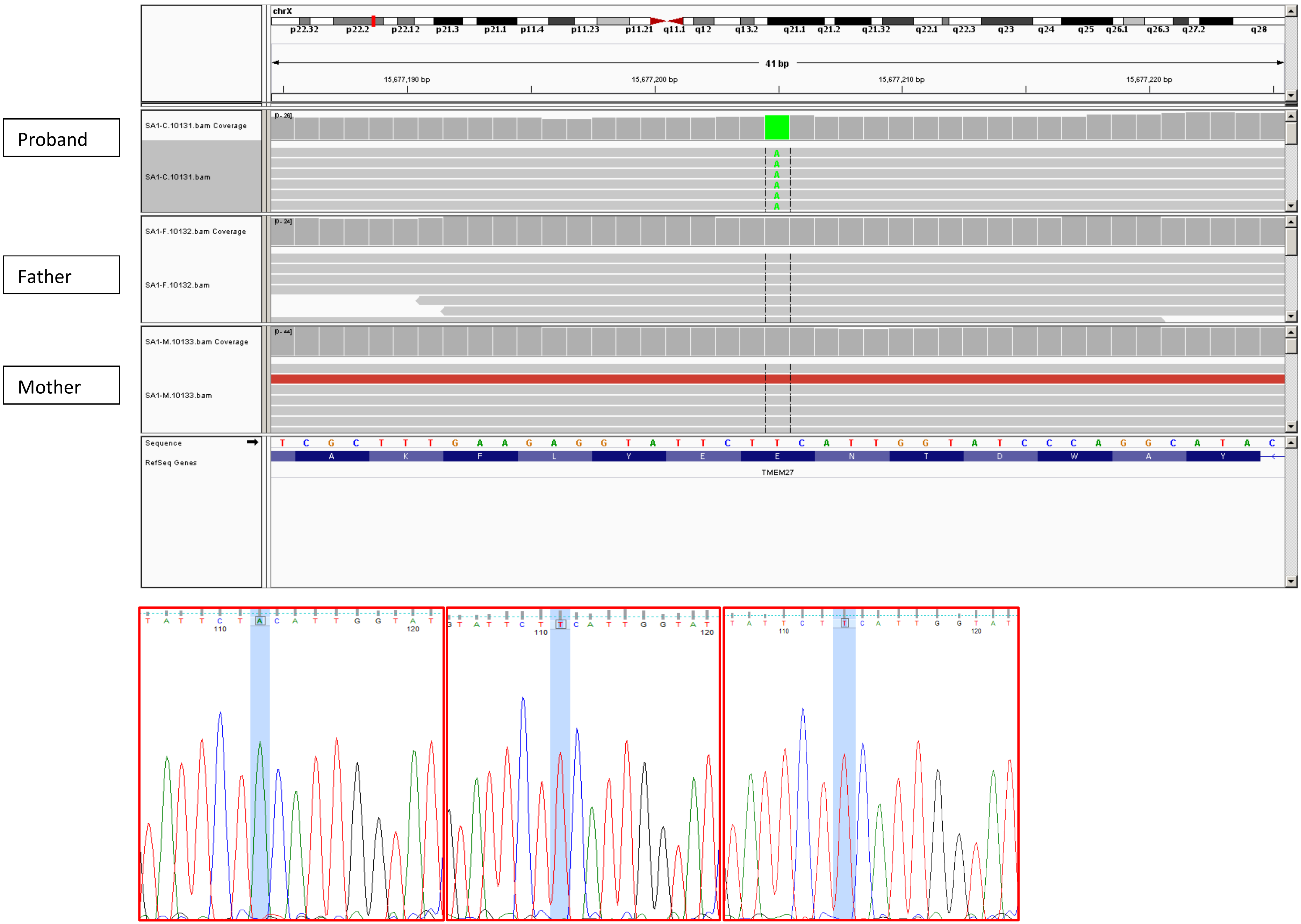


Proband Father Mother

11. SMPA71: *ARHGEF9*

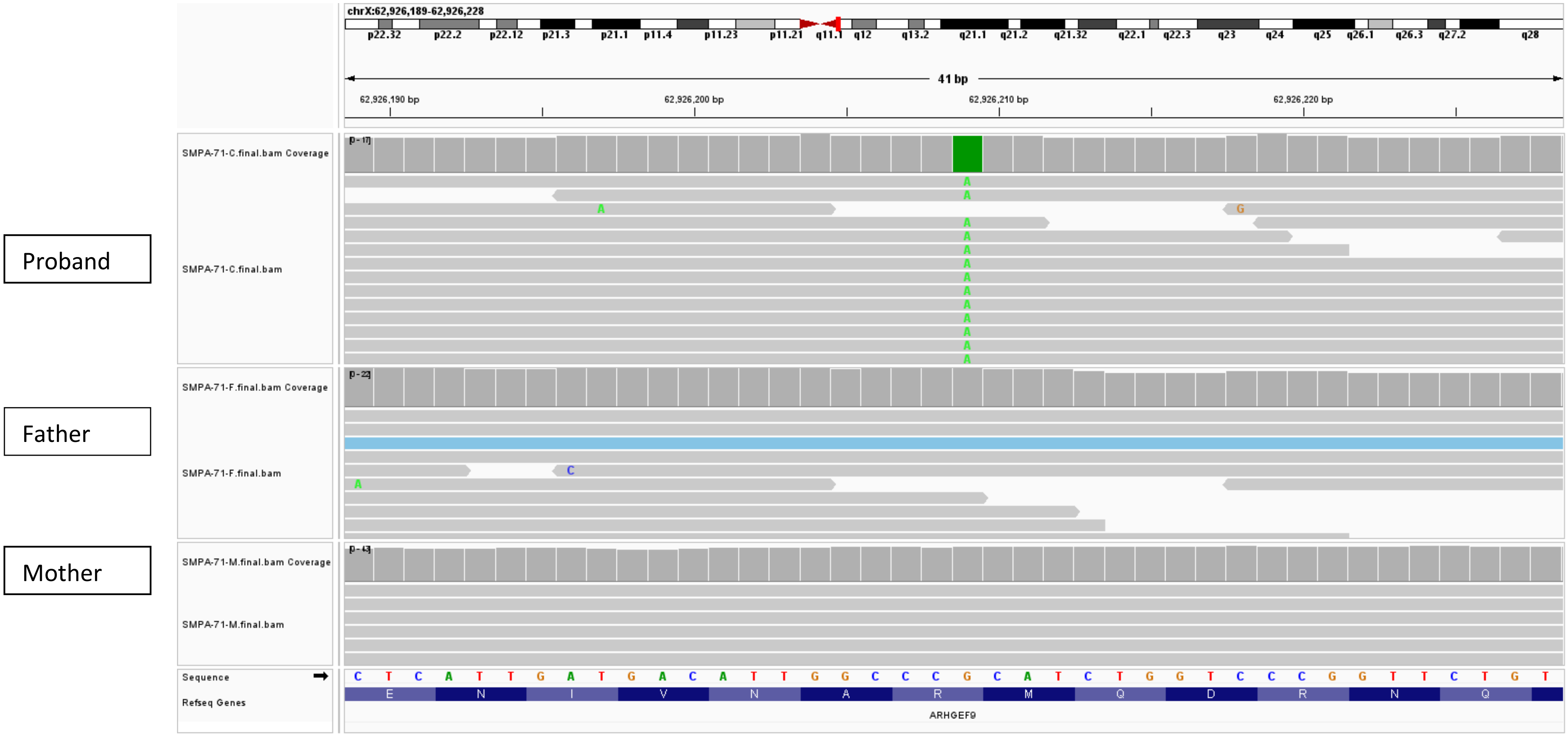


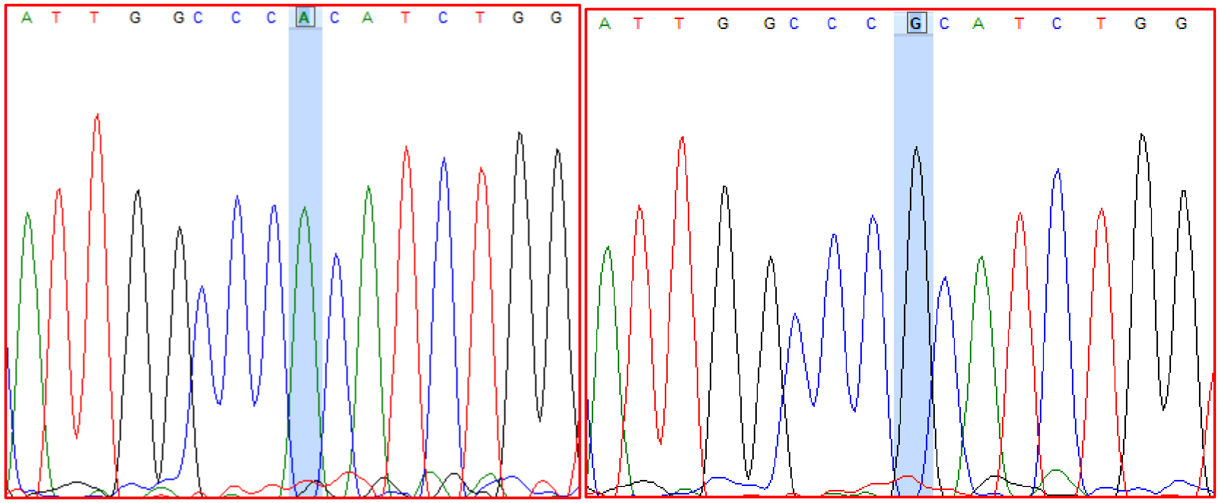


Proband Mother

12. SMPA84: *DRP2*

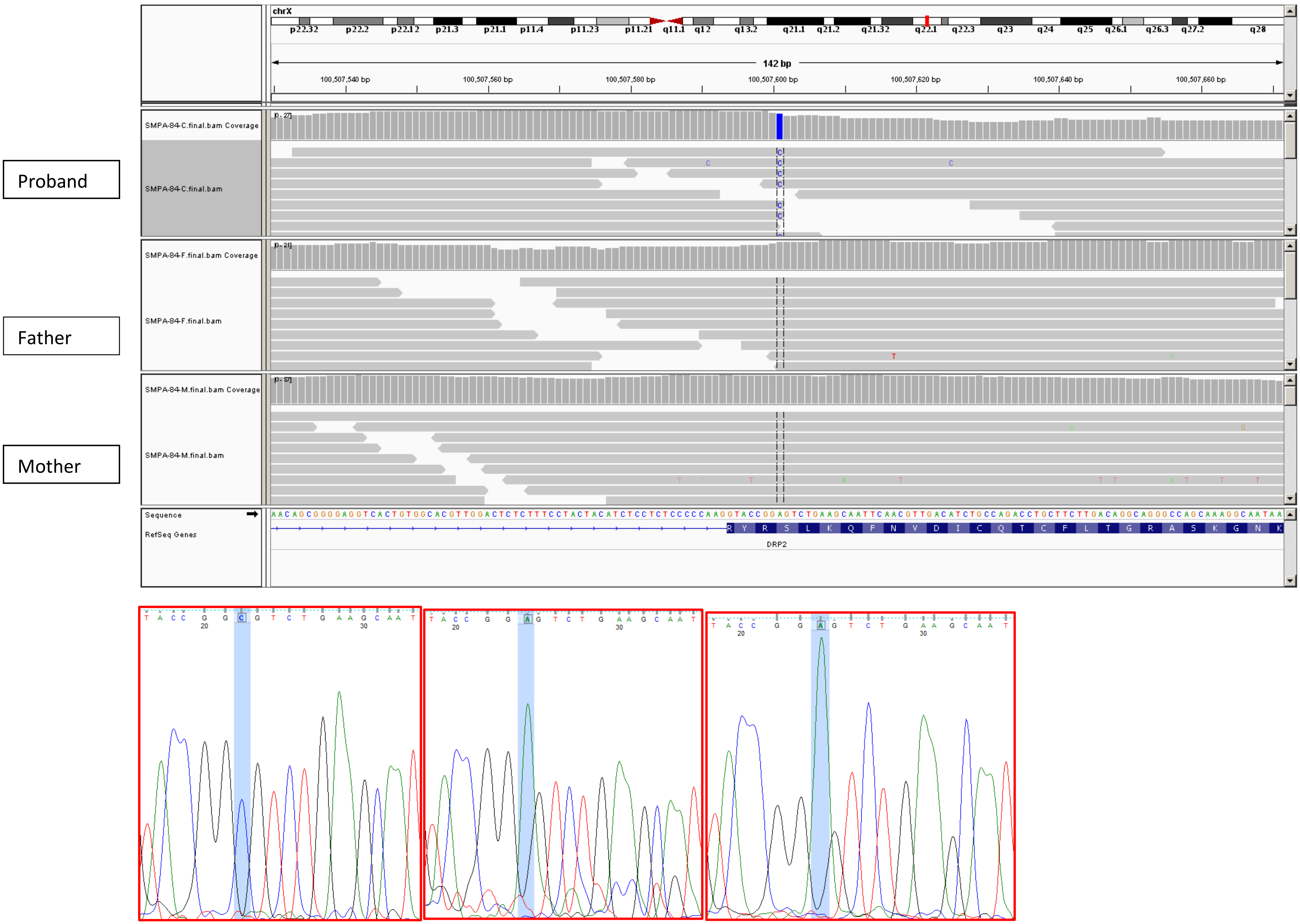


Proband Father Mother

13. IAU54: *MECP2- de novo*

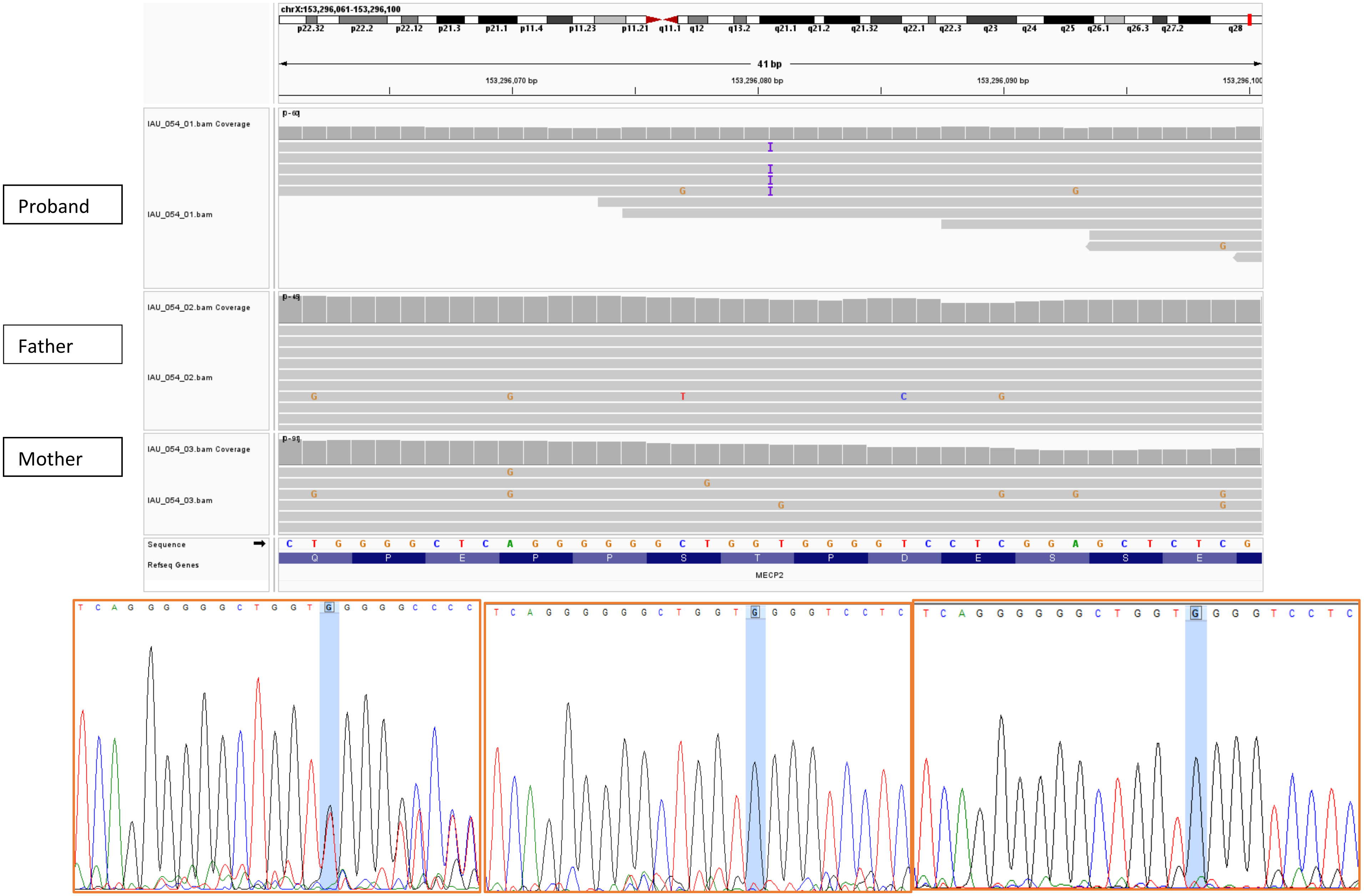


Proband Father Mother

14. SMPA78: *MECP2- de novo*

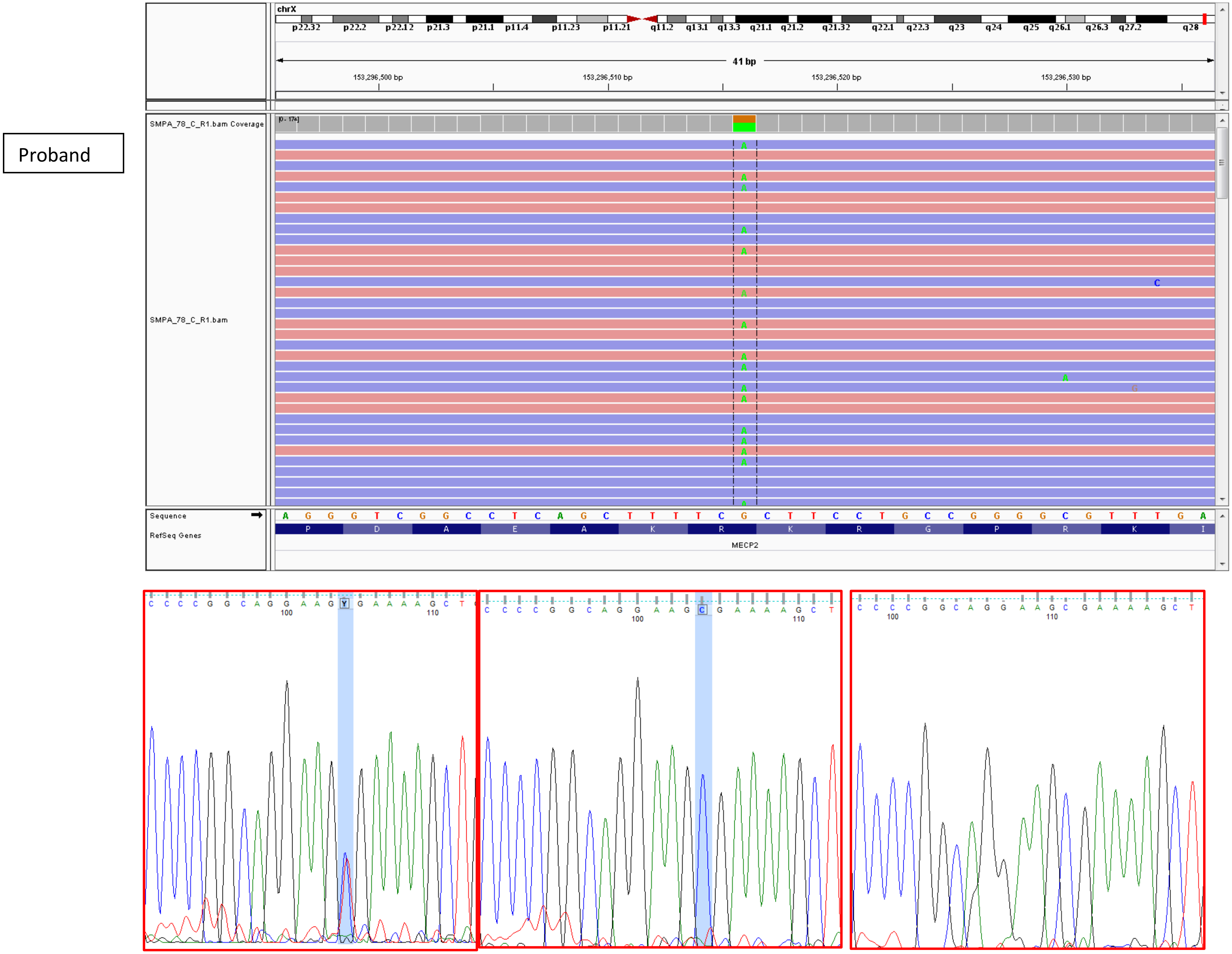


Proband Father Mother

Table 5: Biallelic loss CNVs

1. SMPA4: 14q11.2: *DHRS4/DHRS4L*

Proband

Father

Mother

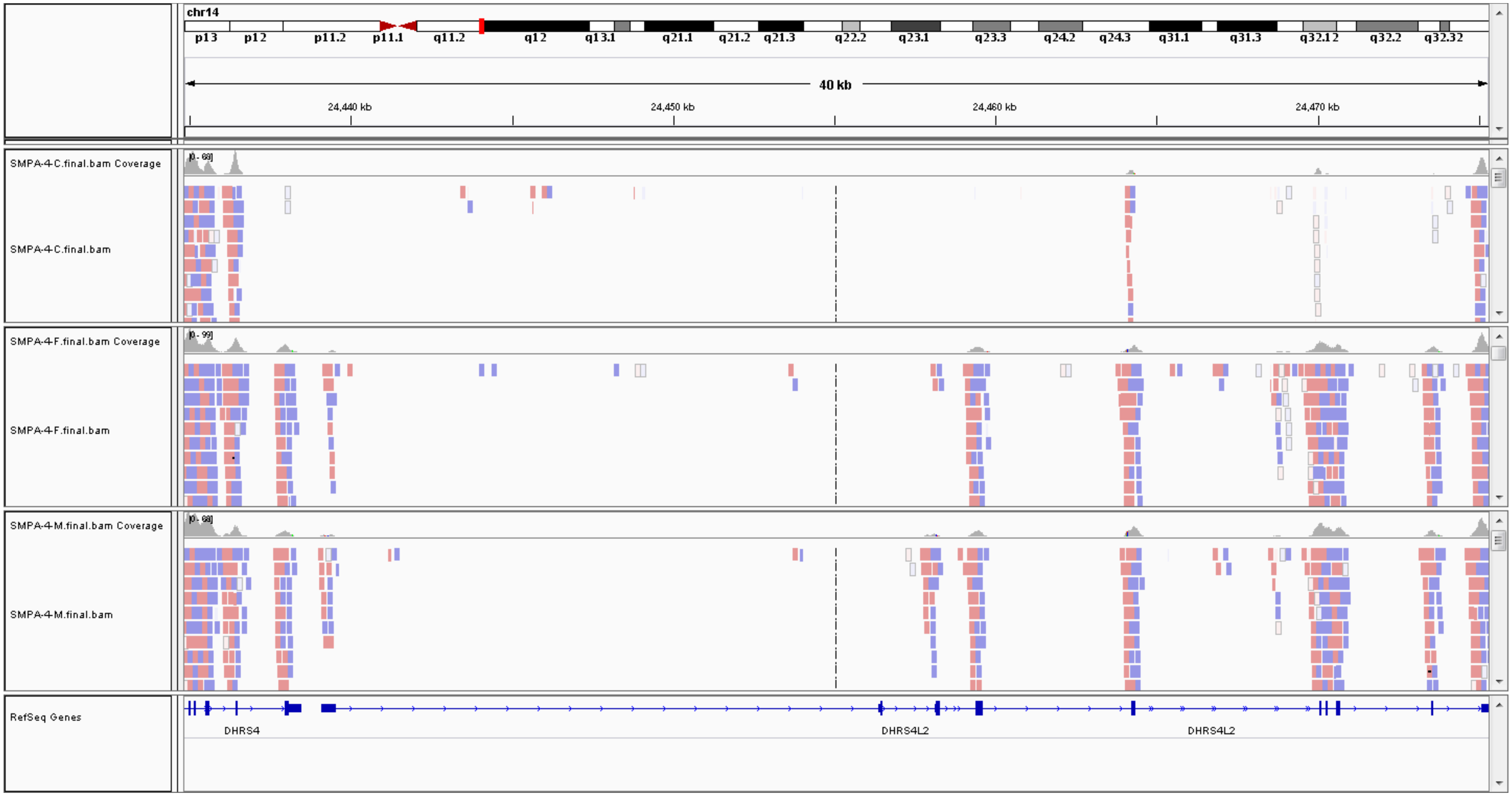


C=child/proband

F=father

M=mother

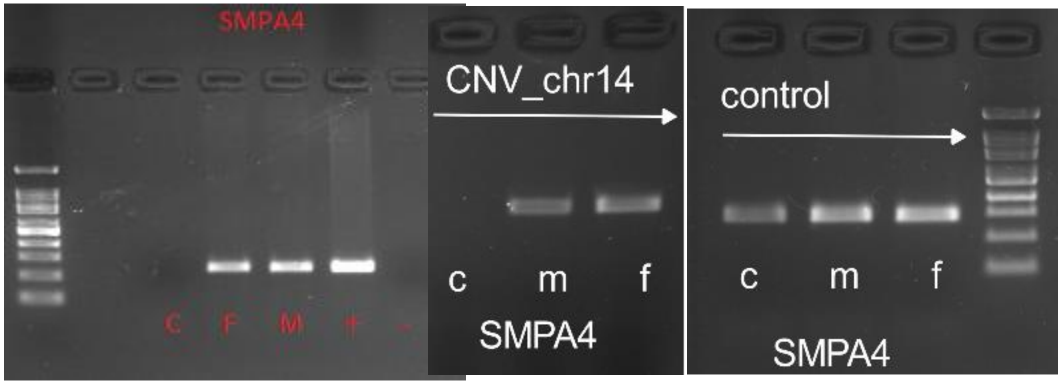

2. SMPA4: *SEMG1*

Proband

Father

Mother

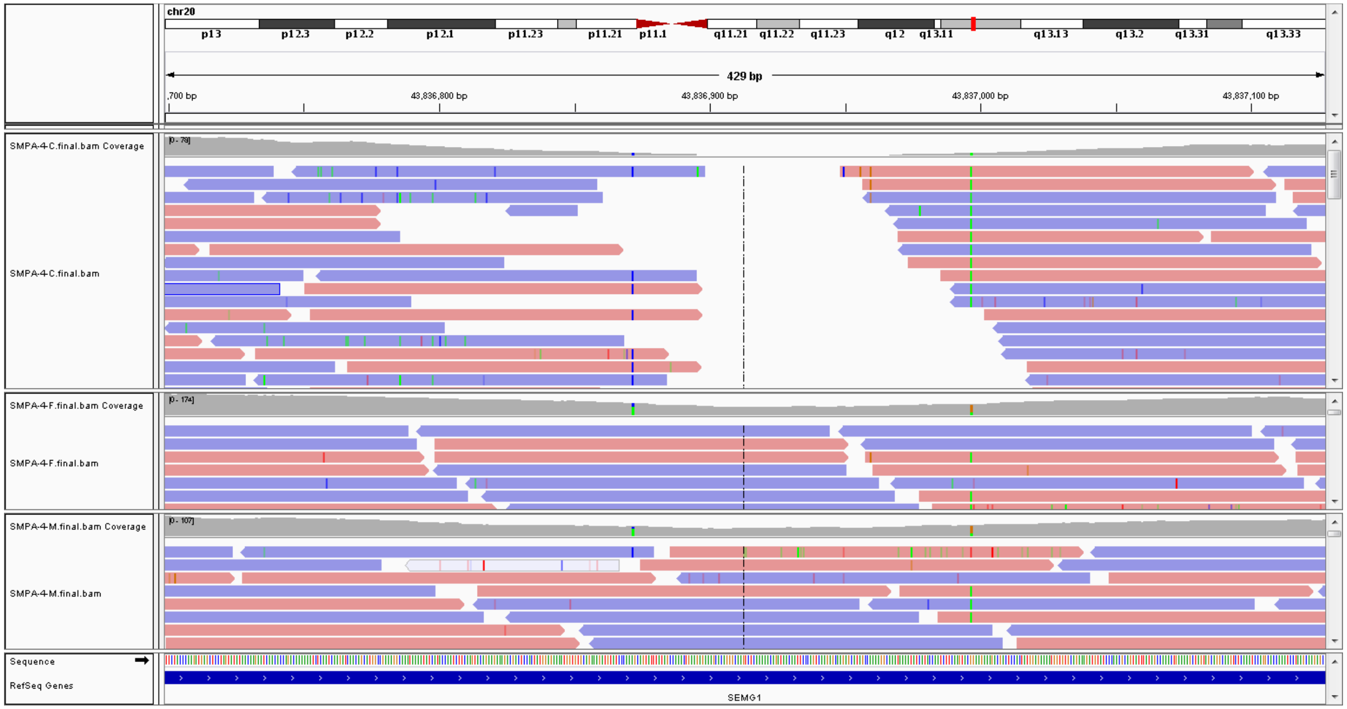


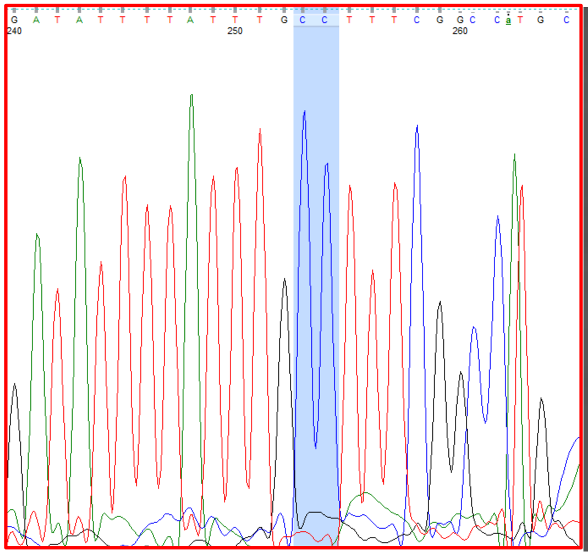

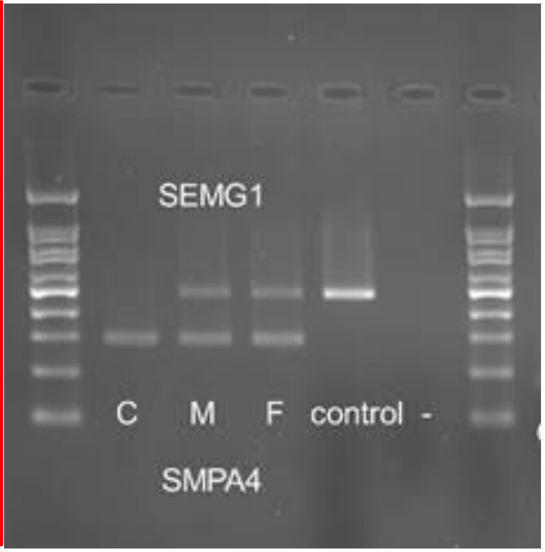


Proband (showing 180bp deletion junction)

3. SMPA8: 2q32.2: *DNAH7*

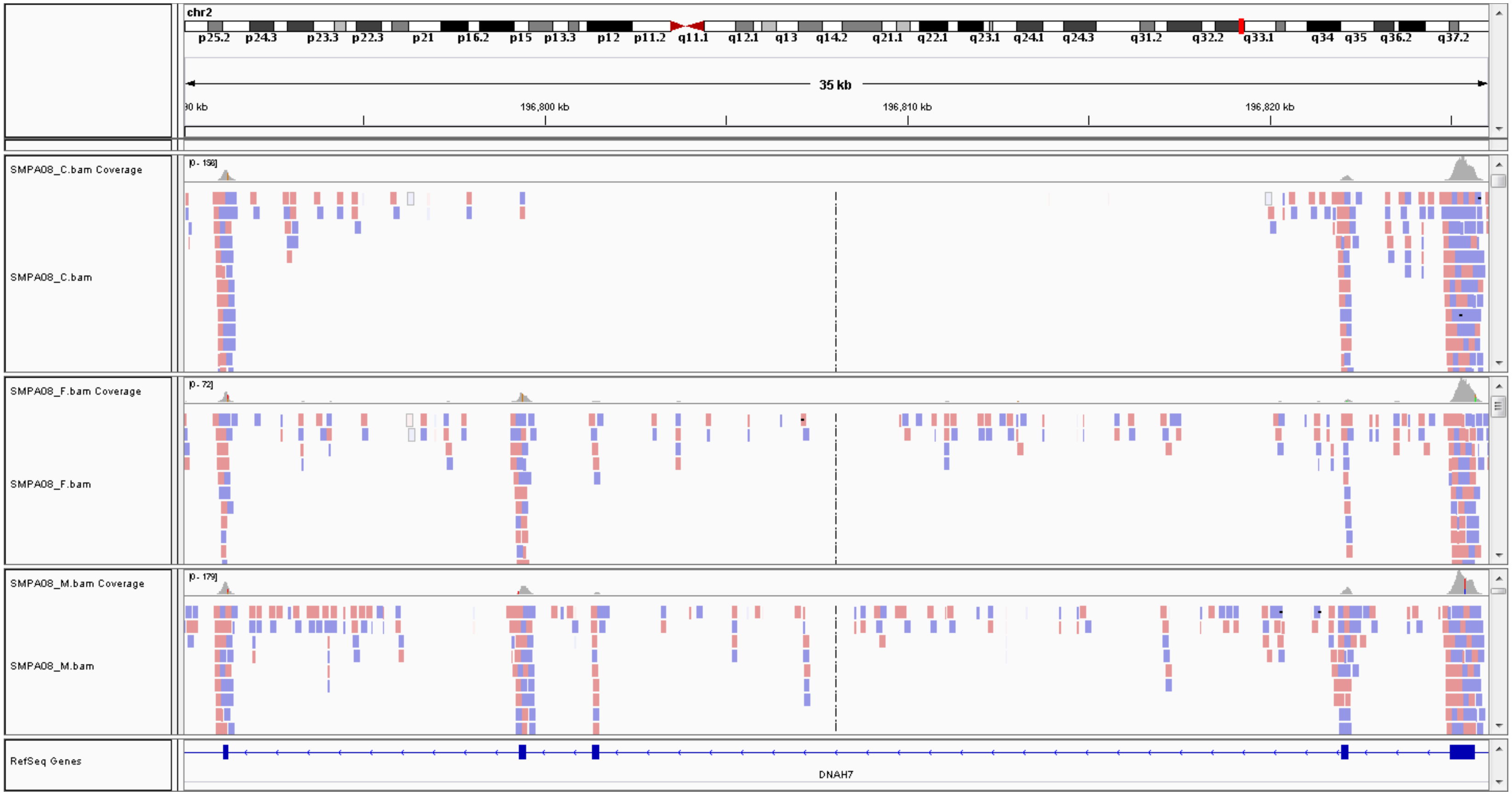


Proband

Father

Mother

C=child/proband

F=father

M=mother

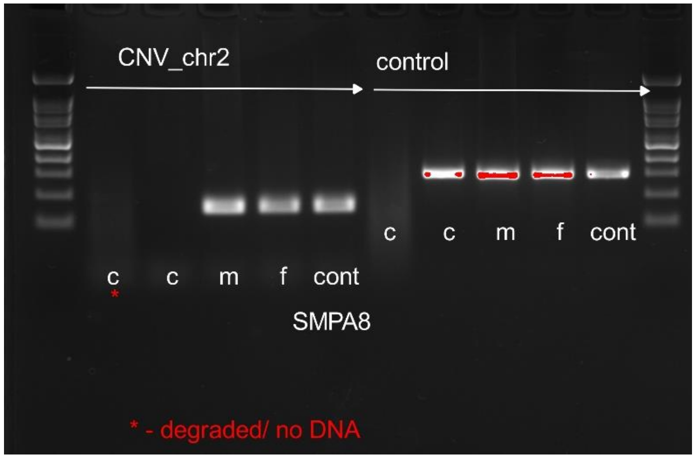

4. SMPA3: 5q31.3: *PCDHA9*, *PCDHA10*

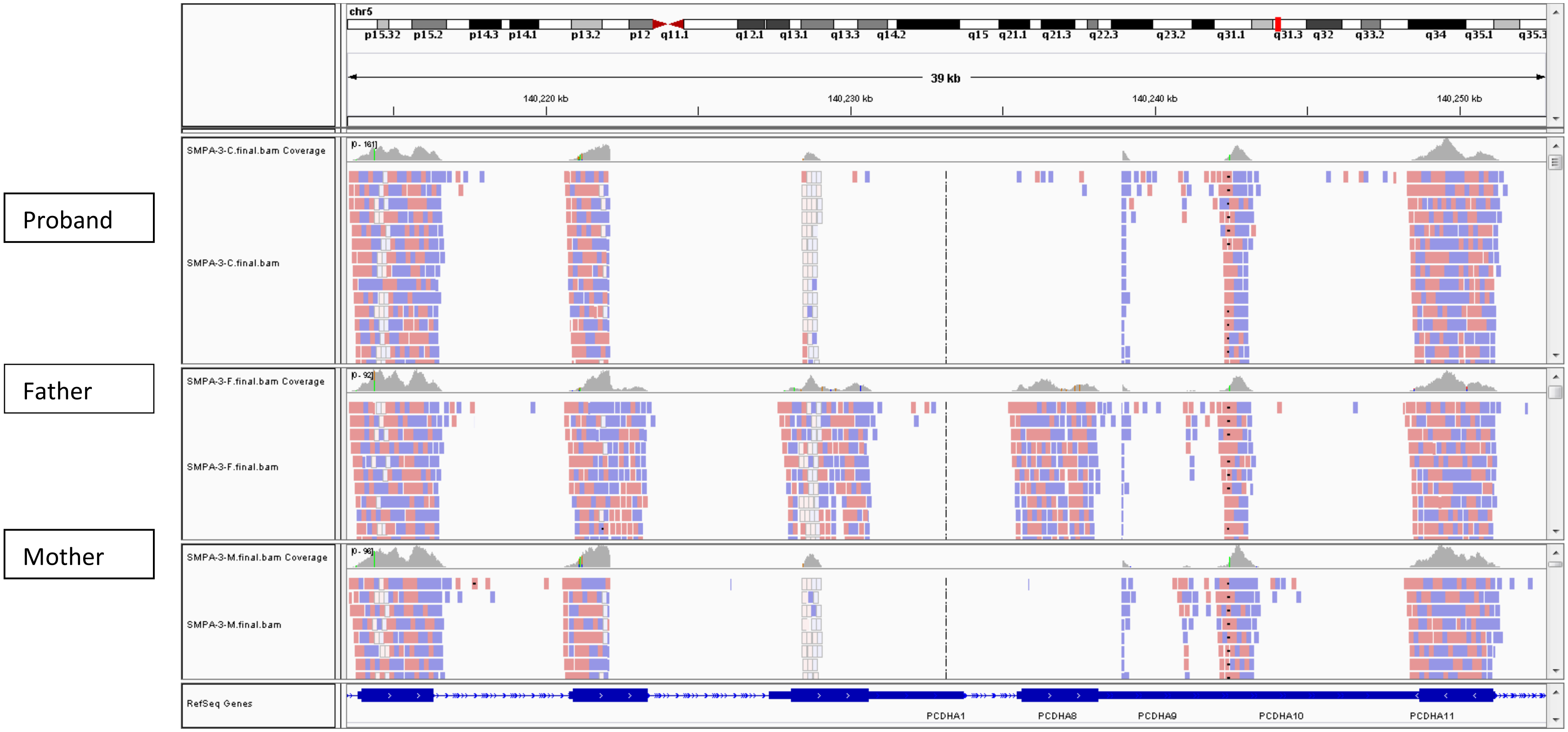


C=child/proband

F=father

M=mother

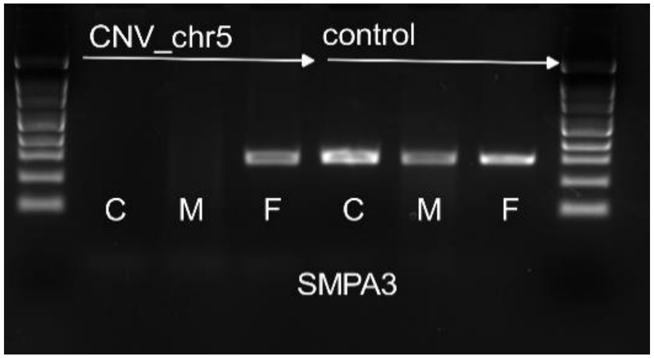

5. IAH2: 5q31.3: *PCDHA9, PCDHA10*

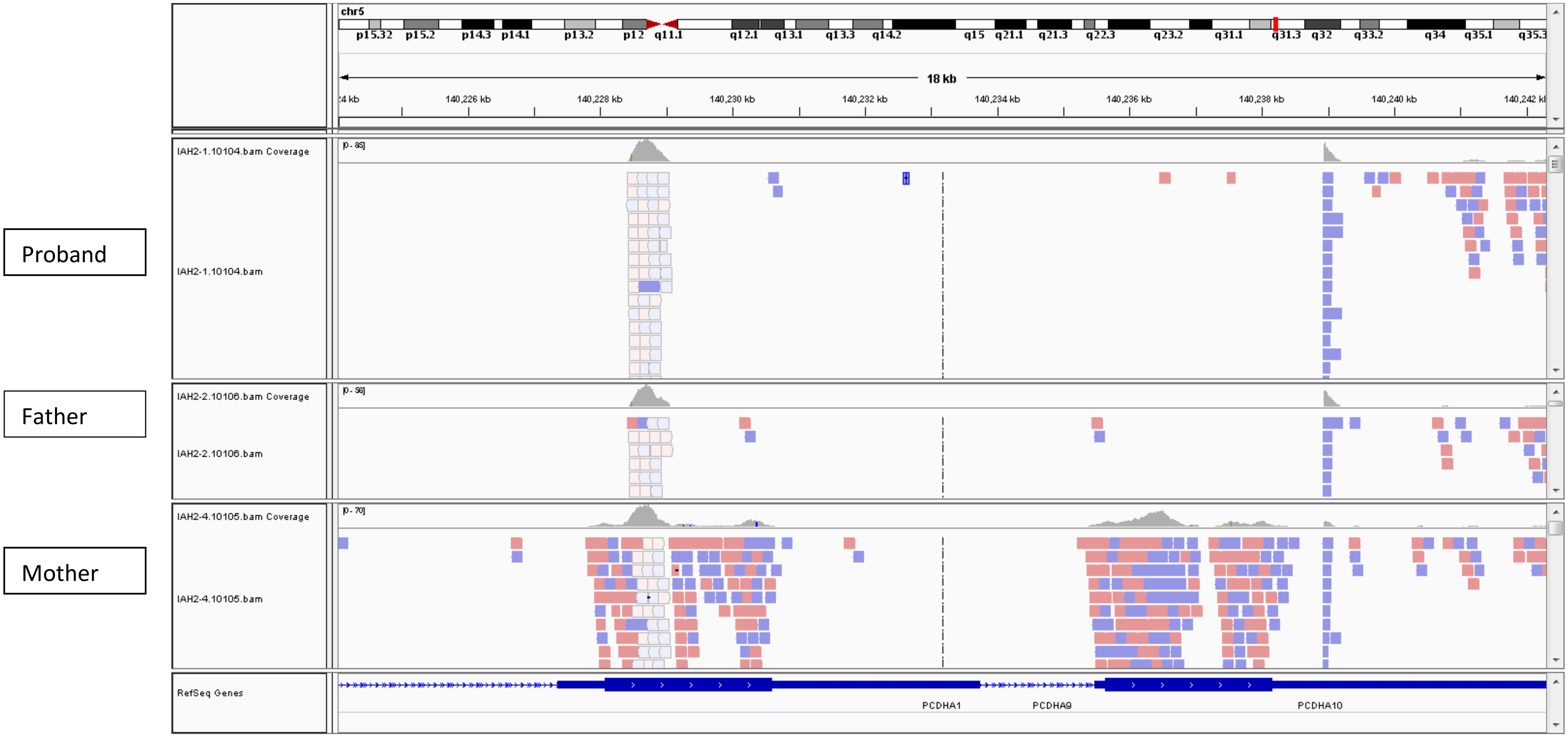


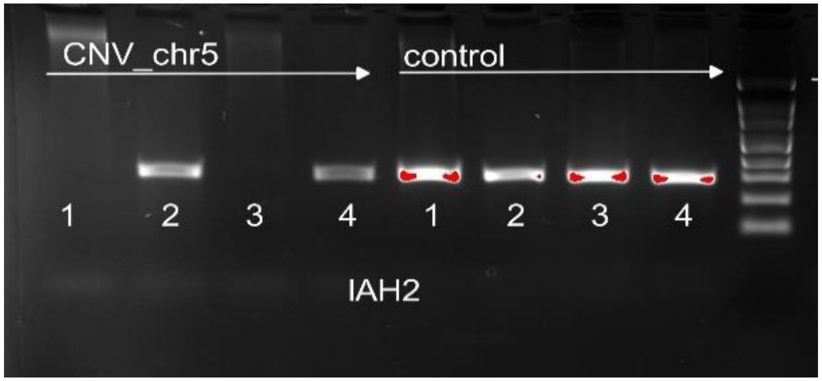


1=proband; 2=unaffected brother; 3=father; 4=mother

6. IAU79: 20p13: *SIRPB1*

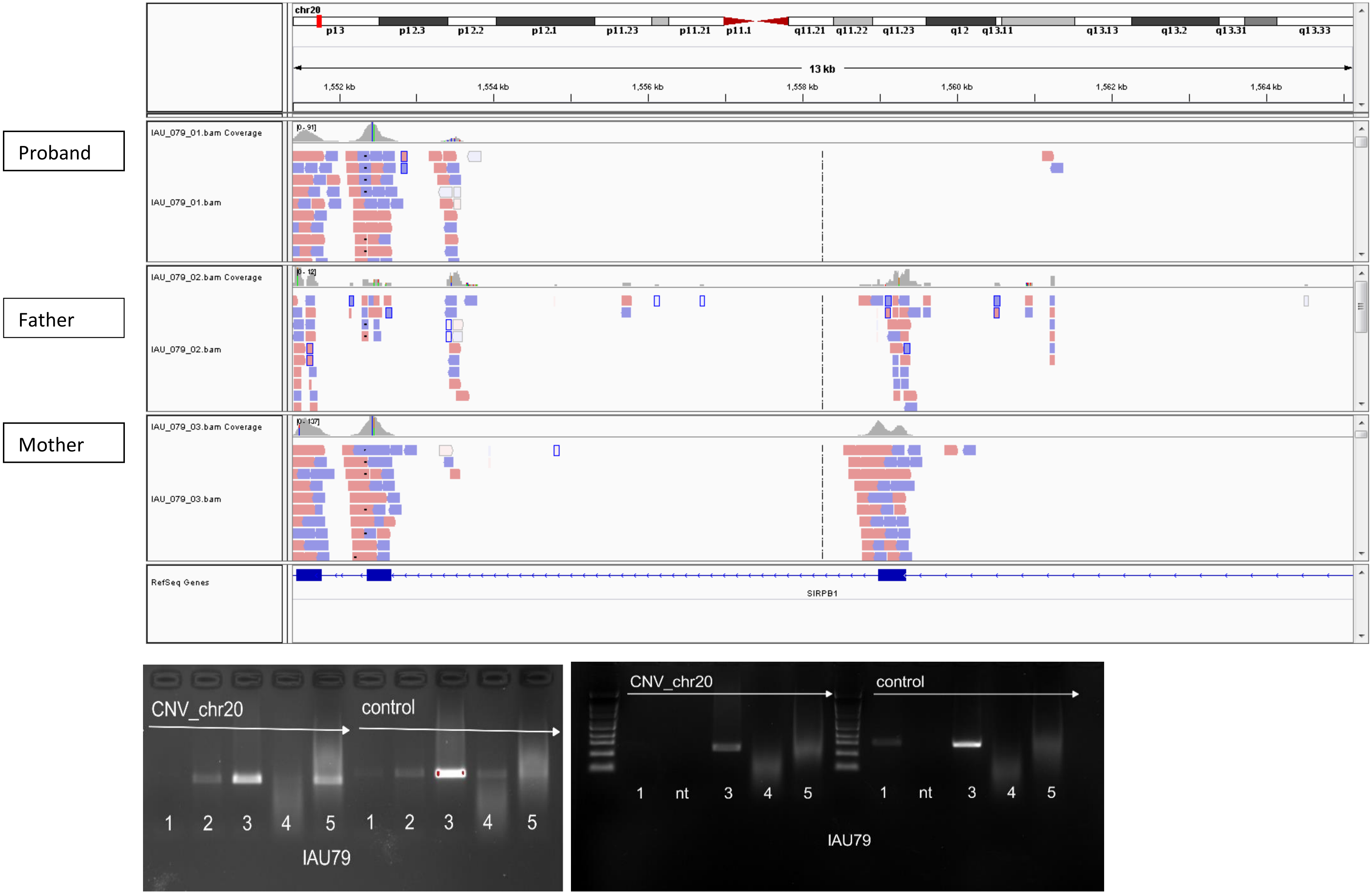

7. IABB2: 17p13.2: *SHPK* (validated by PCR and qPCR)

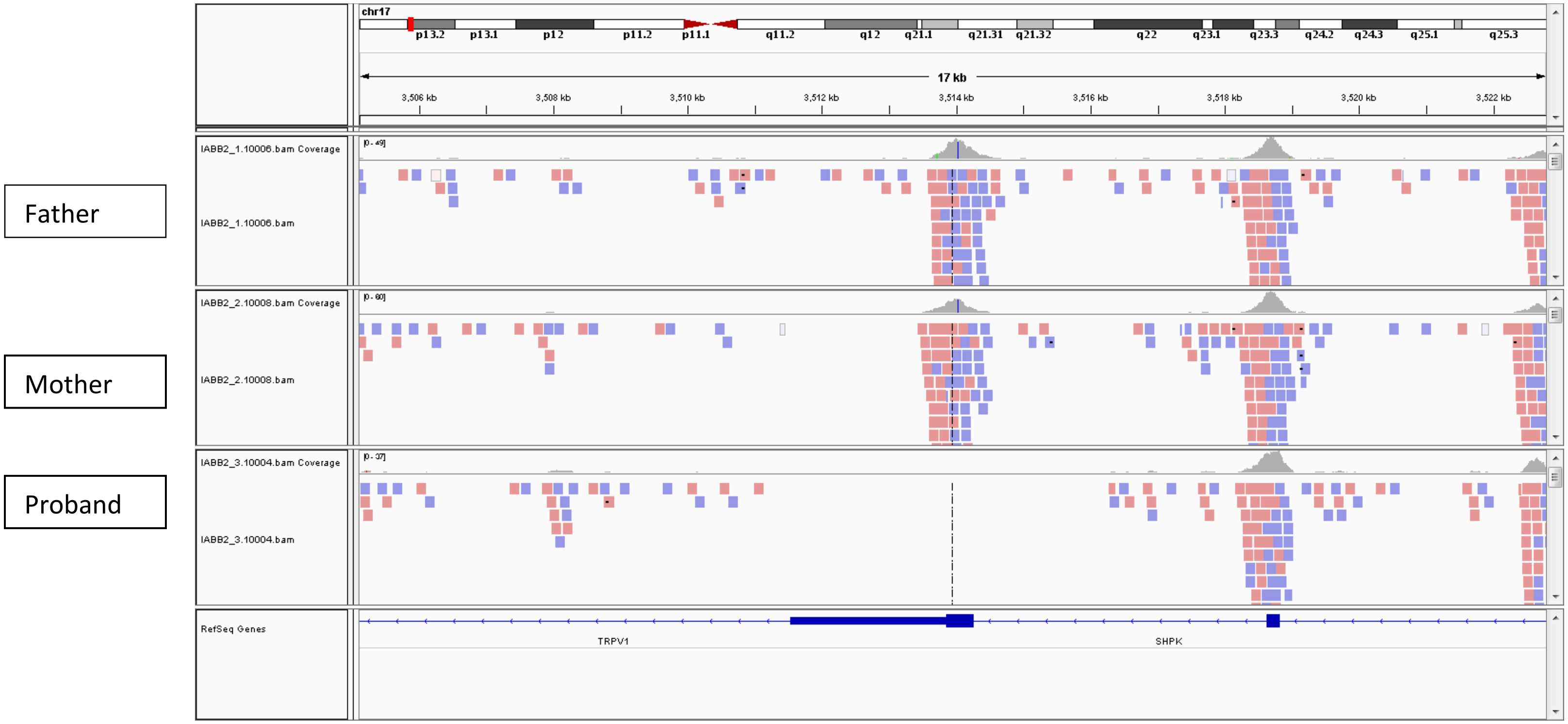


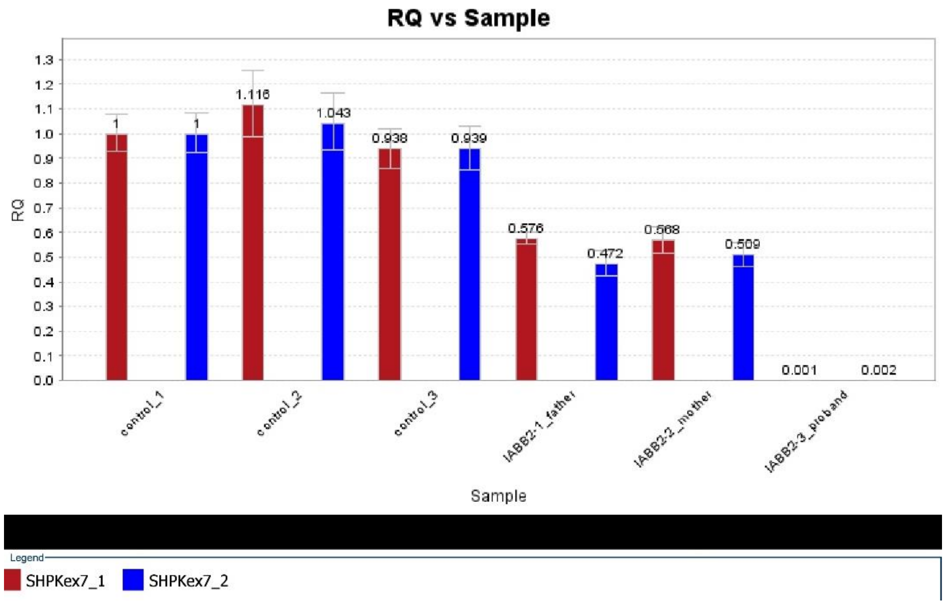

8. IABB14: *WDR73*

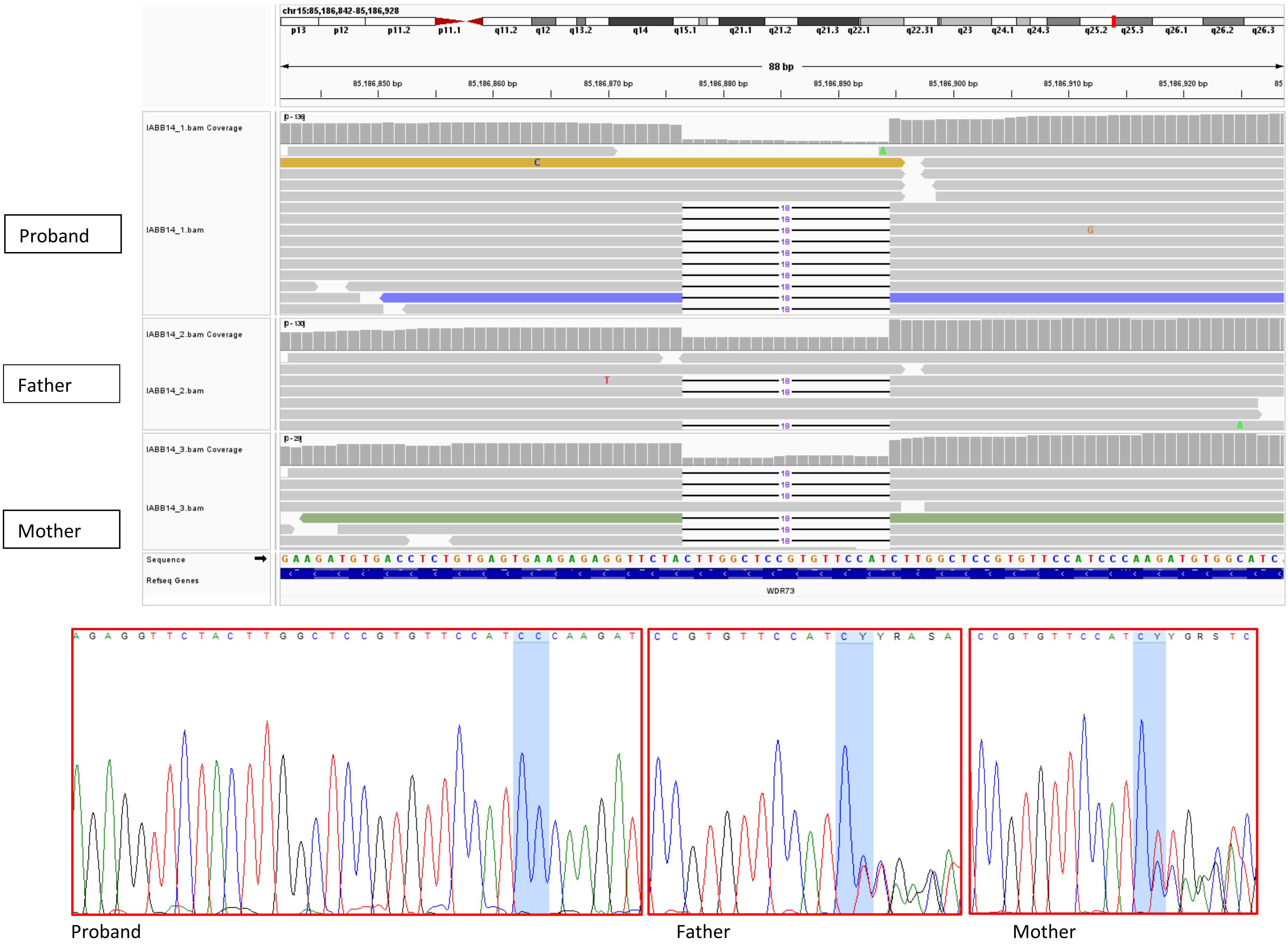

9. IABB14: 19q13.2: *CYP2A7*

Proband

Father

Mother

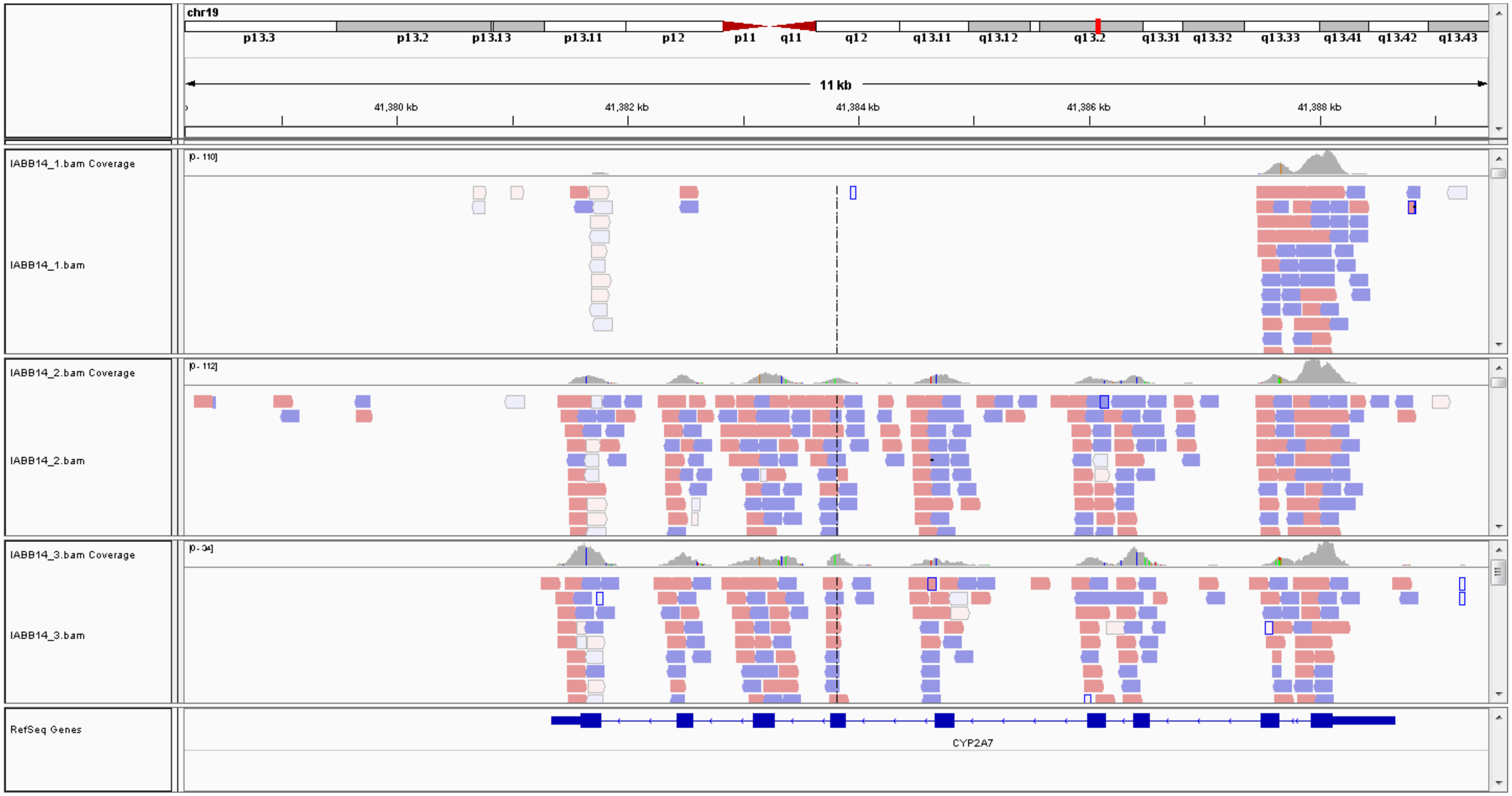


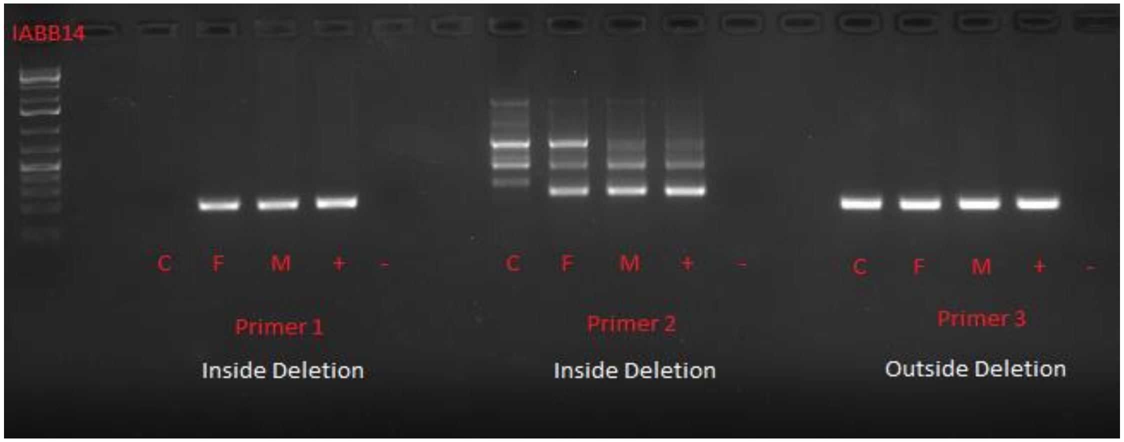


C=child/proband

F=father

M=mother

+=positive control

-=negative control

Exon 8

Intron 4

Exon 1/Intron 1

10. IABB14: 19q13.33: *KLK15* (validated by PCR and qPCR)

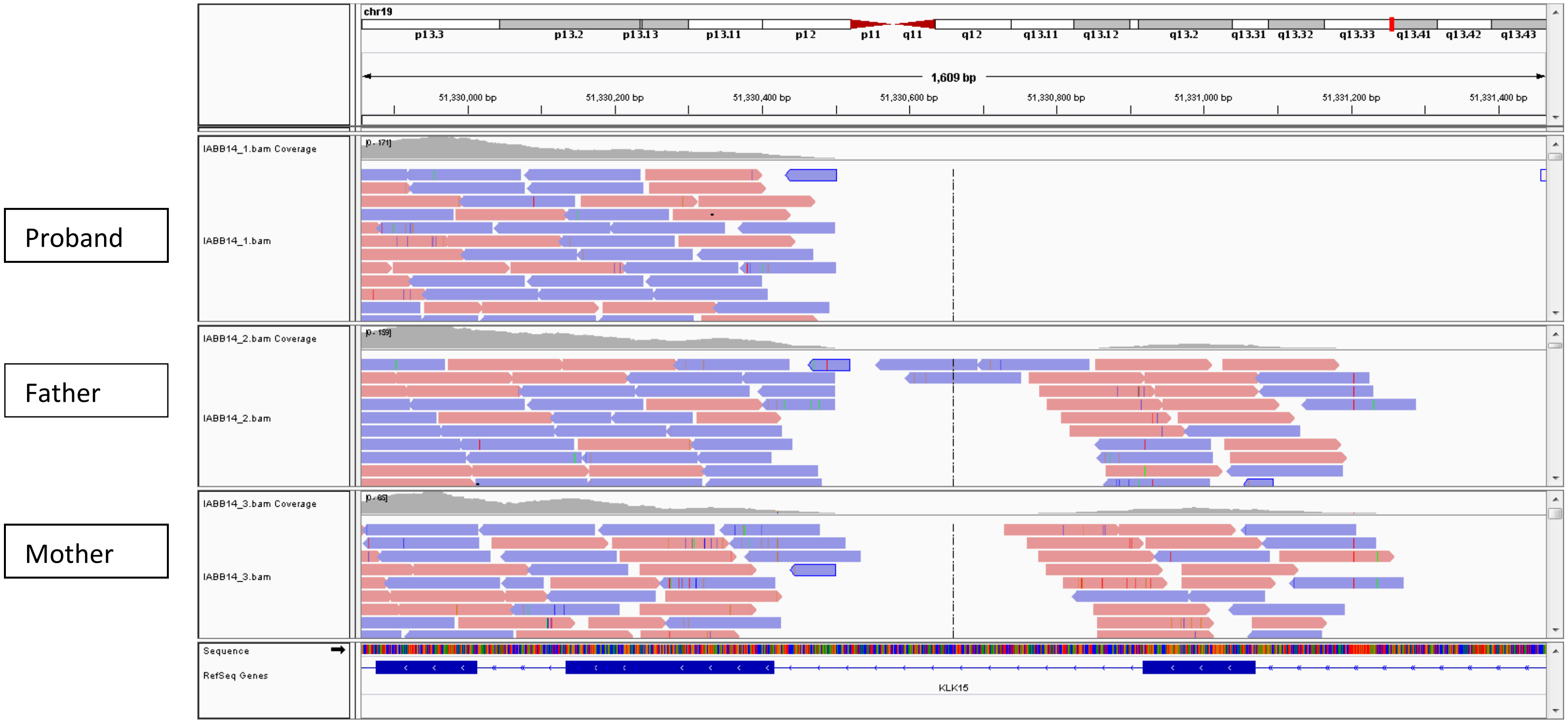


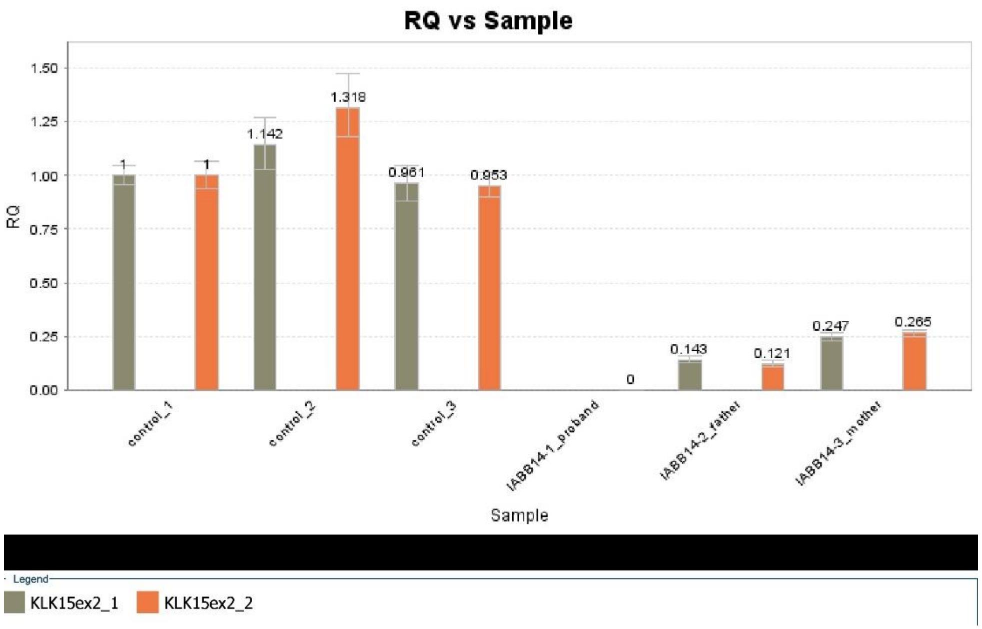

*11.* SMPA42: Xp11.23: *ZNF630/SSX6*

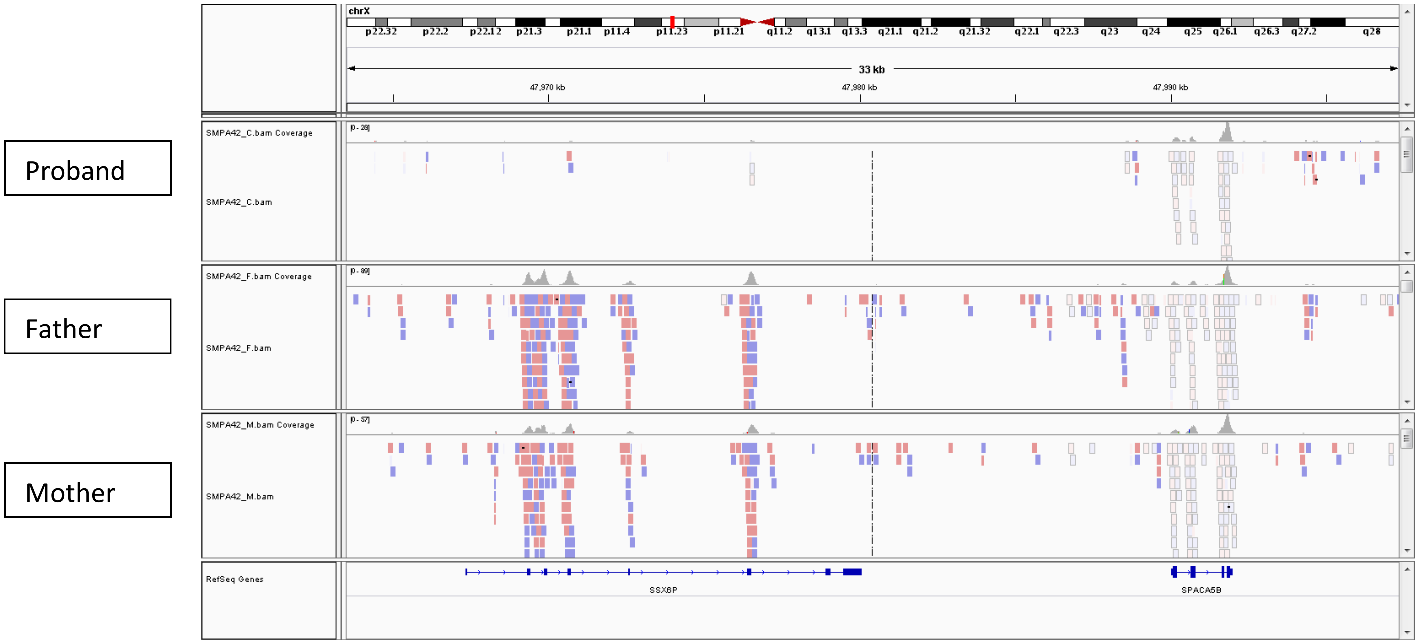


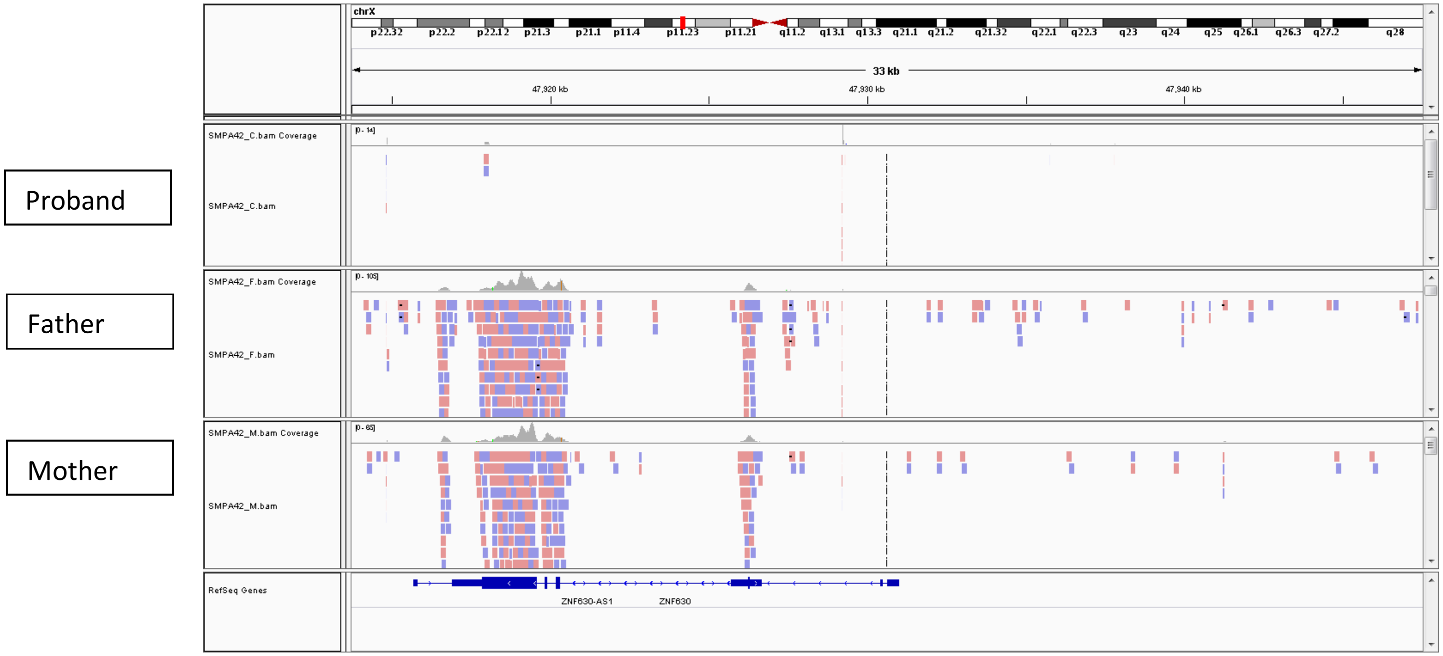

*12.* SMPA9: Xp11.23: *ZNF630/SSX6*

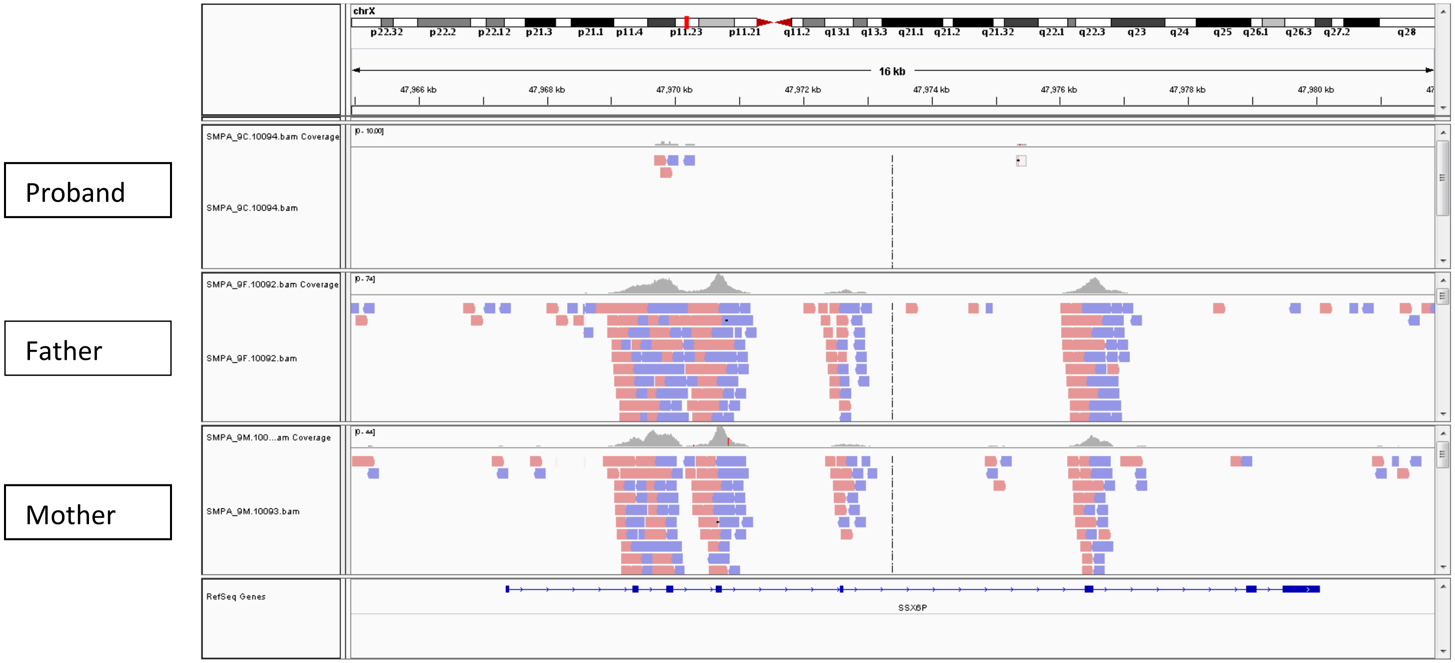


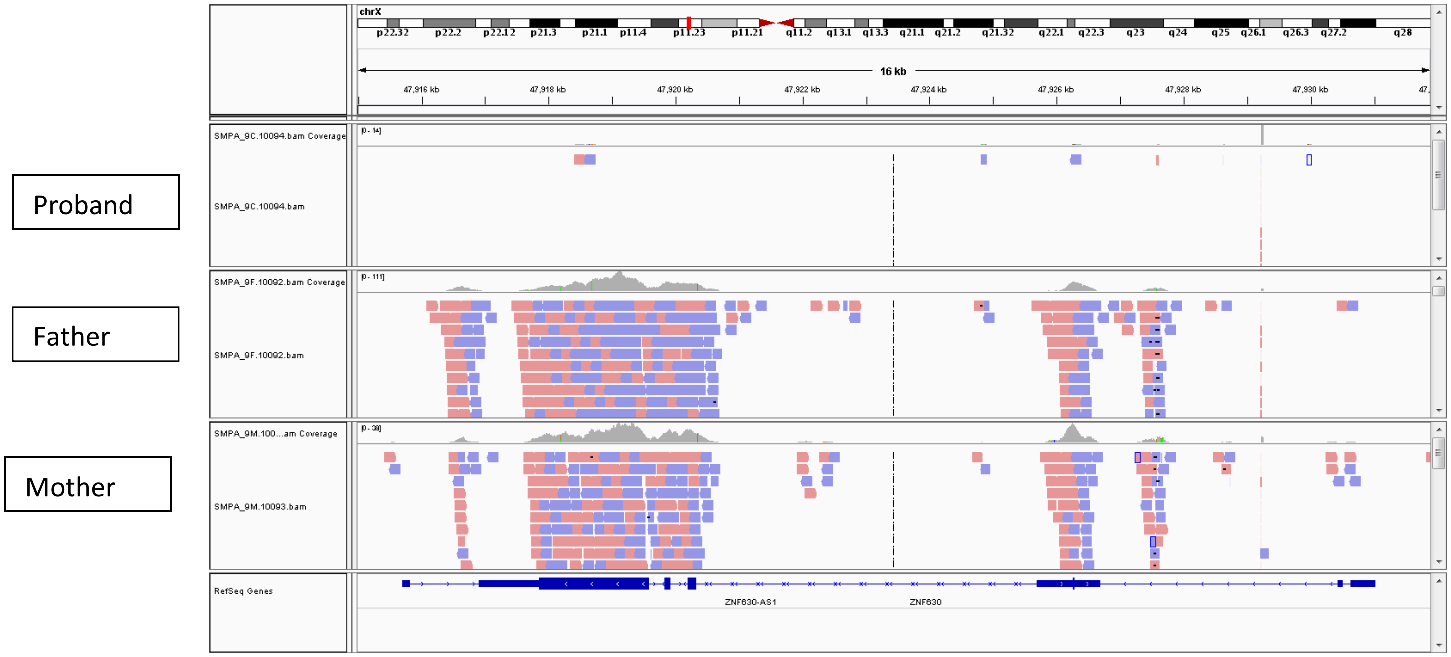

**Table 6: Hemizygous CNVs**

1. SMPA80: Chr9:370,244-13,398,587

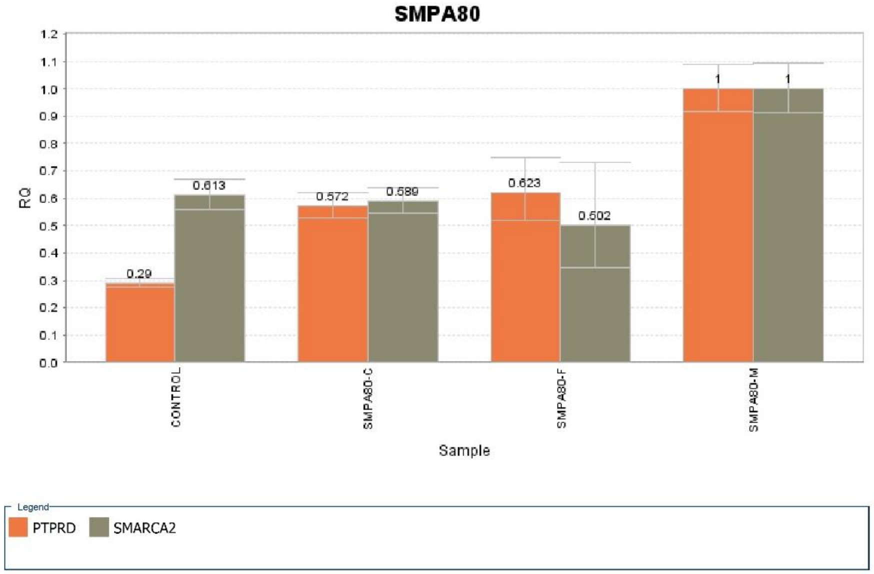


2. IAU25: Chr17:16,778,164-20,364,207

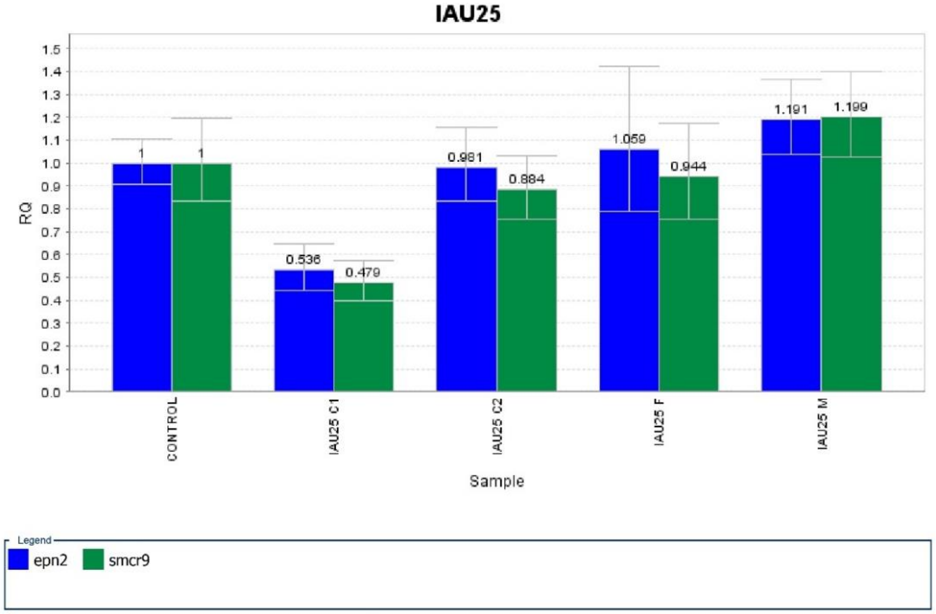

3. IAU74: Chr2:235,370,826-243,034,519

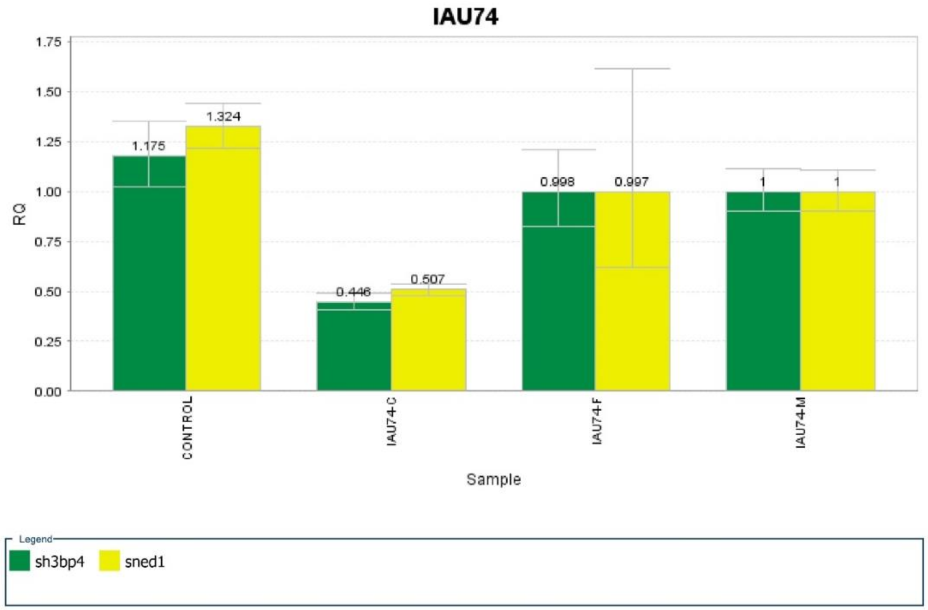


4. IAU7: Chr1:249357-699532

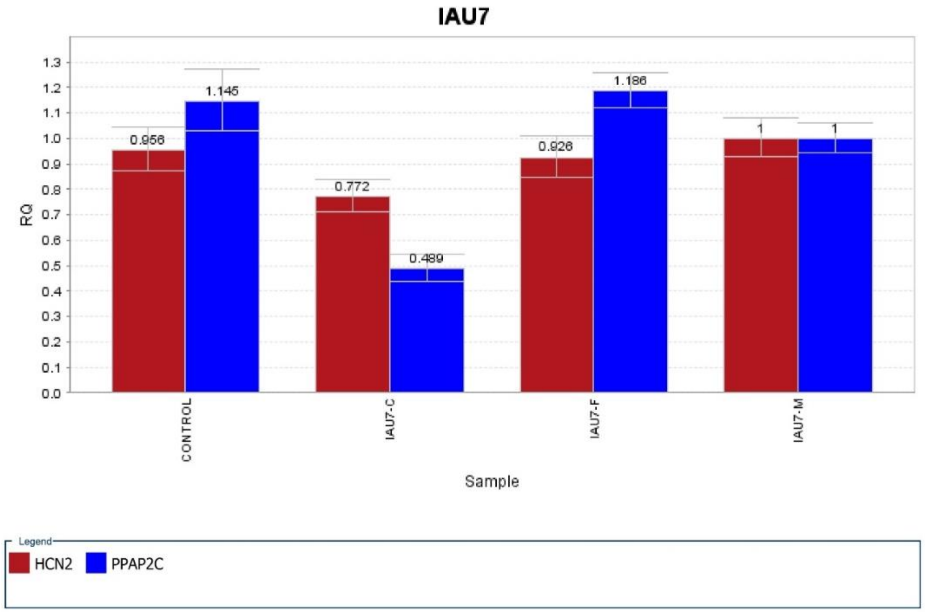

5. SMPA15: Chr15:91,290,633-91,347,451
6. SMPA15: Chr15:40,477,419-41,026,994

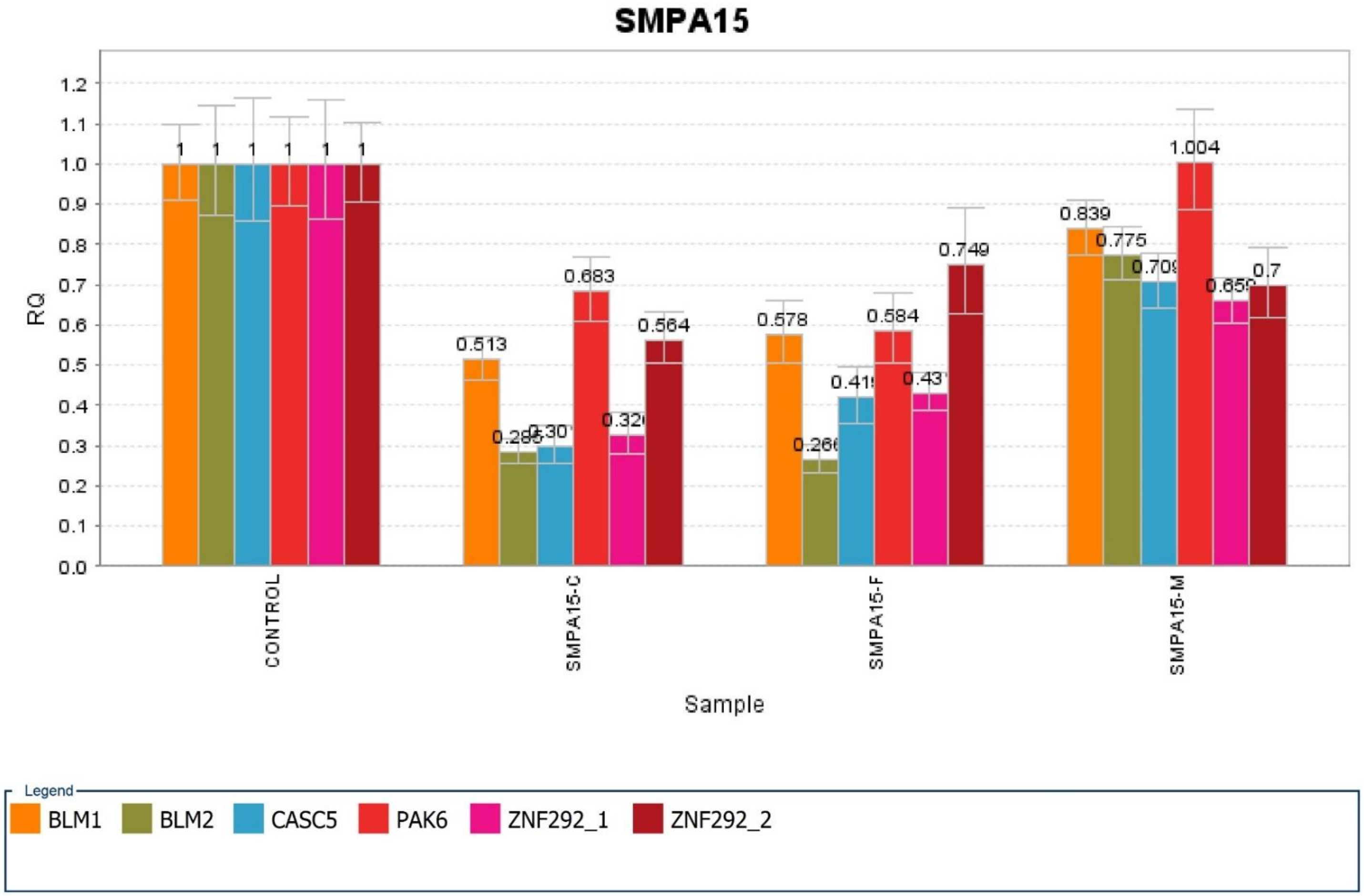
